# Limiting to English language records: A comparison of five methods on Ovid MEDLINE and Embase versus removal during screening

**DOI:** 10.64898/2026.03.18.26348470

**Authors:** Helen Fulbright, Kirsty Morrison

## Abstract

**Background:** For evidence syntheses using English language limits, several different methods and approaches are available.

**Objective:** To understand the English language (EL) limits available on Ovid MEDLINE and Embase and the application of language metadata on these databases. To compare the impact of five EL limits versus removing non-English language (NEL) records during screening.

**Methods:** Using the records included at full text screening or excluded on NEL status during screening in seven evidence syntheses, we tested five EL limits on 1,509 MEDLINE and 1,584 Embase records. ‘Includes’ removed or ‘NEL excludes’ retrieved were investigated.

**Results:** All EL limits performed identically, 99.8% of MEDLINE ‘includes’ were retrieved versus 99.7% on Embase. All five ‘includes’ incorrectly removed with EL limits had language metadata errors. Although 98.2% MEDLINE and 94.6% Embase ‘NEL excludes’ were removed with EL limits, eight MEDLINE and nine Embase records were available in English.

**Discussion:** The risk of excluding potentially eligible records due to language restrictions (whether applied during the strategies or screening) could be mitigated with forward and backward citation searching.

**Conclusion:** EL limits risk removing records with incorrect language metadata. However, EL records might also be excluded on language during screening.

## BACKGROUND

Within evidence syntheses, the exclusion of non-English language (NEL) records is referred to as ‘language bias’ [1]. Reviews that exclude records based on language could miss relevant evidence and consequently skew or prejudice the findings. This could be caused by various factors, including the types of evidence available in different languages, since positive results are more likely to be published in English language (EL) journals [2, 1]. Language bias can therefore magnify ‘publication bias’, which is the publication of research according to the direction or strength of its findings [3]. However, for some reviews, the research question or objectives might directly justify the use of language restrictions. This could be the case if only EL records are likely to be relevant, such as studies on a United Kingdom population.

Additionally, the exclusion of EL records might be considered under certain circumstances, such as constraints on time, budget, or other resources (e.g., size of the review team). These are the most common reasons for excluding NEL records, followed by a lack of resources to translate studies [4]. *The Cochrane Handbook for Systematic Reviews of Interventions* recommends awareness of the impact of language bias and how this could be minimised, though note the need to achieve a balance between thoroughness and efficiency in the use of time and funds (see chapter 4, section 4.2.2) [5].

An overview of evidence on the impact and application of language restrictions is provided in chapter 4, section 4.4.5 of the *Cochrane Handbook* [5]. One investigation of studies on conventional medicine found no direct evidence of bias [1], whereas several others found an impact on statistical significance in a minority of cases, although all conclusions remained the same [6, 7, 8]. Although some studies have observed differences in review findings for complementary and alternative medicine [9, 10], one study of reviews on acupuncture found that inclusion of Chinese resources had no significant effect on conclusions [11], suggesting variation by the intervention or topic of the review. This evidence has been used to suggest language restrictions may be a viable shortcut, especially for rapid reviews, unless eligible records are likely to be published in a NEL [7, 12].

The application of language restrictions has received less attention in the literature. Review teams that decide to limit to EL records can apply this within the search strategies, or during screening. Excluding NEL records during screening is more sensitive than within the search strategies, as this can remove records with incorrect or absent language metadata [13]. This approach gives review teams the option to translate these articles later and has also been recommended by Pieper and Puljak as it allows transparent reporting of records available in other languages [14].

The different approaches to applying language restrictions (i.e., in the search strategies or during screening) have not yet been compared. There are also several options and approaches for applying language limits in search strategies on Ovid MEDLINE and Embase, which are both widely used databases in evidence syntheses. Some information specialists use a technique to prevent the exclusion of records without language metadata [13], though both Ovid MEDLINE and Embase now provide an inbuilt limit to retrieve records with no language specified. Results can also be refined by selecting from a list of available languages or entering search terms for languages, though both databases have records in languages that are not clearly defined (e.g., ‘multilingual’ on MEDLINE). These methods on Ovid MEDLINE and Embase warrant further investigation and are also valuable to compare to the removal of NEL records during screening. Understanding the different methods and approaches available and how they compare to each other could inform decision-making by information specialists and review teams.

## OBJECTIVES

This is an exploratory study which aims to investigate the impact of five methods to limit database searches to EL records on Ovid MEDLINE and Embase, and to compare these methods to the removal of NEL records during screening. The main objectives are to:

- understand how language metadata is applied on MEDLINE and Embase via Ovid;
- explore the impact of five methods to limit to EL records on Ovid MEDLINE and Embase and how they compare to each other; and
- explore how the five methods compare with the removal of NEL records during screening.

## METHODS

A summary of the methodology is as follows, we:

1. communicated with Ovid, the National Library of Medicine (NLM), and Elsevier to ask questions on how language metadata is applied on MEDLINE and Embase;
2. examined which methods can be used to limit to EL records on MEDLINE and Embase;
3. determined how many records included at full text screening or excluded on NEL status during screening in seven evidence syntheses, would have been retrieved or removed with each method; and
4. investigated any ‘includes’ removed or ‘NEL excludes’ retrieved by each method to analyse the accuracy of the language metadata.

### Understanding how language metadata is applied on Ovid MEDLINE and Embase

We e-mailed Ovid, NLM, and Elsevier to enquire how language metadata is determined and applied to records, and whether language metadata remains the same or gets updated. The language index pages for MEDLINE and Embase were also consulted [15, 16].

Though available on the same database platform (Ovid), MEDLINE is produced by the National Library of Medicine [17], whereas Embase is produced by Elsevier [18]. This means the language metadata is applied separately, even for the content available on both databases [19, 20].

For MEDLINE via Ovid, the metadata for each record is downloaded from the file transfer protocol server of the NLM [21]. A baseline set of PubMed records are released by NLM annually with updates of new, revised, and deleted records released daily [22]. Publishers submit their citations (and metadata) to the NLM either with or without language metadata and have access to edit their citations at any time [19, 23]. In comparison, for Embase via Ovid, records are received from Elsevier and converted to a format compatible with searching on Ovid’s platform [24]. The language metadata is created manually based on information provided by the publisher or what is found in the text [20].

Ovid does not check or update the metadata from the NLM or Elsevier. Where errors are found (i.e., reported to Ovid by customers), this is communicated to the NLM or Elsevier for corrections which are then included in updates [21, 25]. This means there are no differences in language metadata between Embase on Elsevier’s platform (Embase.com) or on Ovid, or between MEDLINE on NLM’s platform (PubMed) or on Ovid, except for records yet to be updated on Ovid.

### Methods of limiting to English language records

We examined which methods could be used to apply language restrictions on Ovid MEDLINE and Embase in the following ways:

1. Ovid’s inbuilt limits for MEDLINE and Embase were explored by viewing ‘Limits’ or selecting ‘Additional Limits’ and browsing the available languages.
2. We consulted the index for ‘Language’ via the ‘Search Fields’ tab, to view the list of languages on each database and the number of records or ‘postings’ available in each language.
3. We entered search terms for languages using the language field code, including two variations of the method described by Aali and Shokraneh [13] to prevent exclusion of records without language metadata – this will subsequently be referred to as the ‘not (NEL not EL)’ method.

On MEDLINE and Embase, inbuilt language limits can be applied through a list of selectable language options under ‘Additional Limits’, though these are not exhaustive. The option to limit to records with ‘no language specified’ can be applied under ‘Limits’ or entered as a search string (see Table 1). It is also possible to enter terms for languages and apply the language field code (lg), though the option to retrieve records with ‘no language specified’ cannot be applied in this way. The language index for each database shows a range of different languages and the number of records or ‘postings’ in each language [15, 16].

**Table 1.**
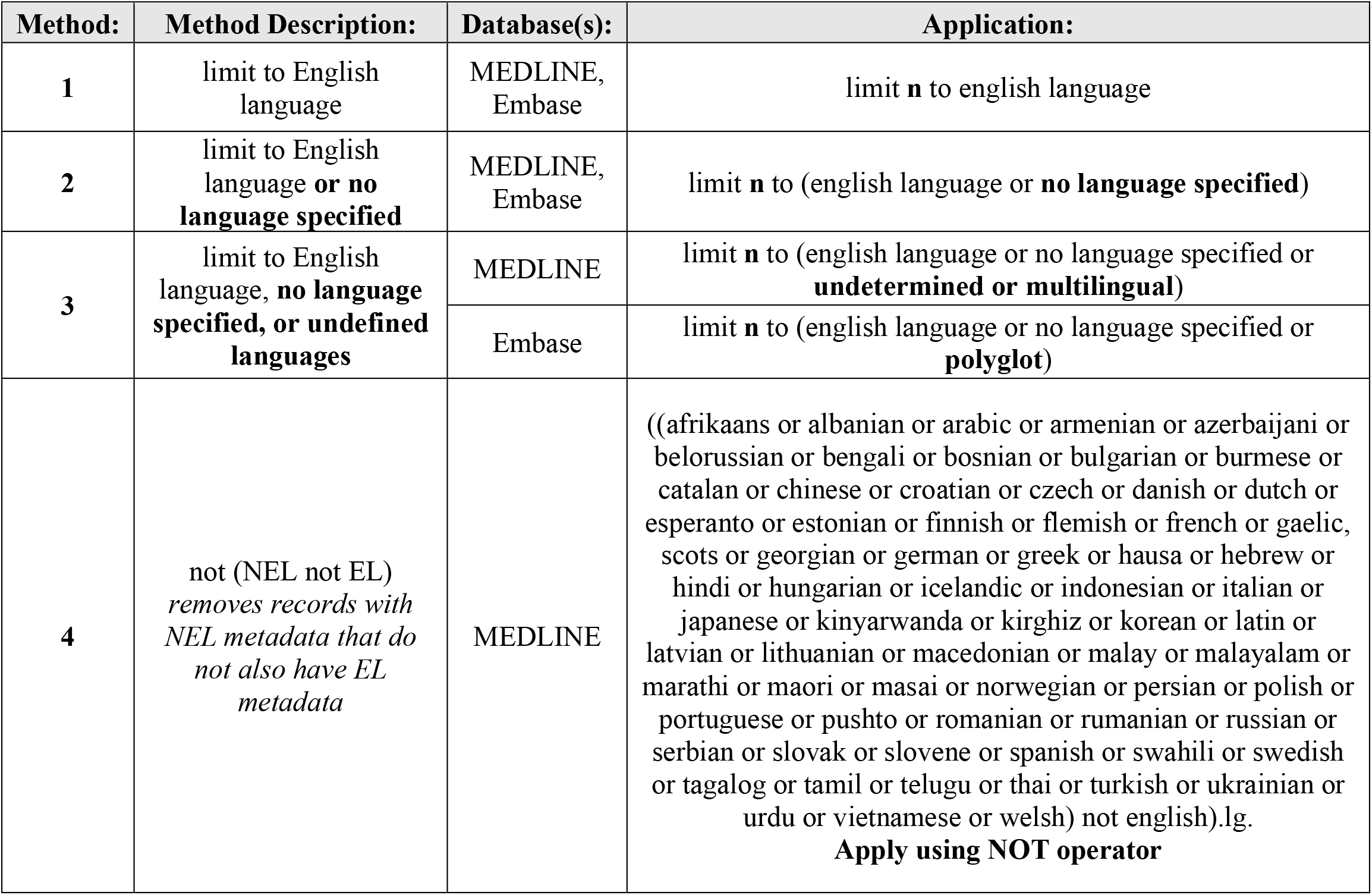
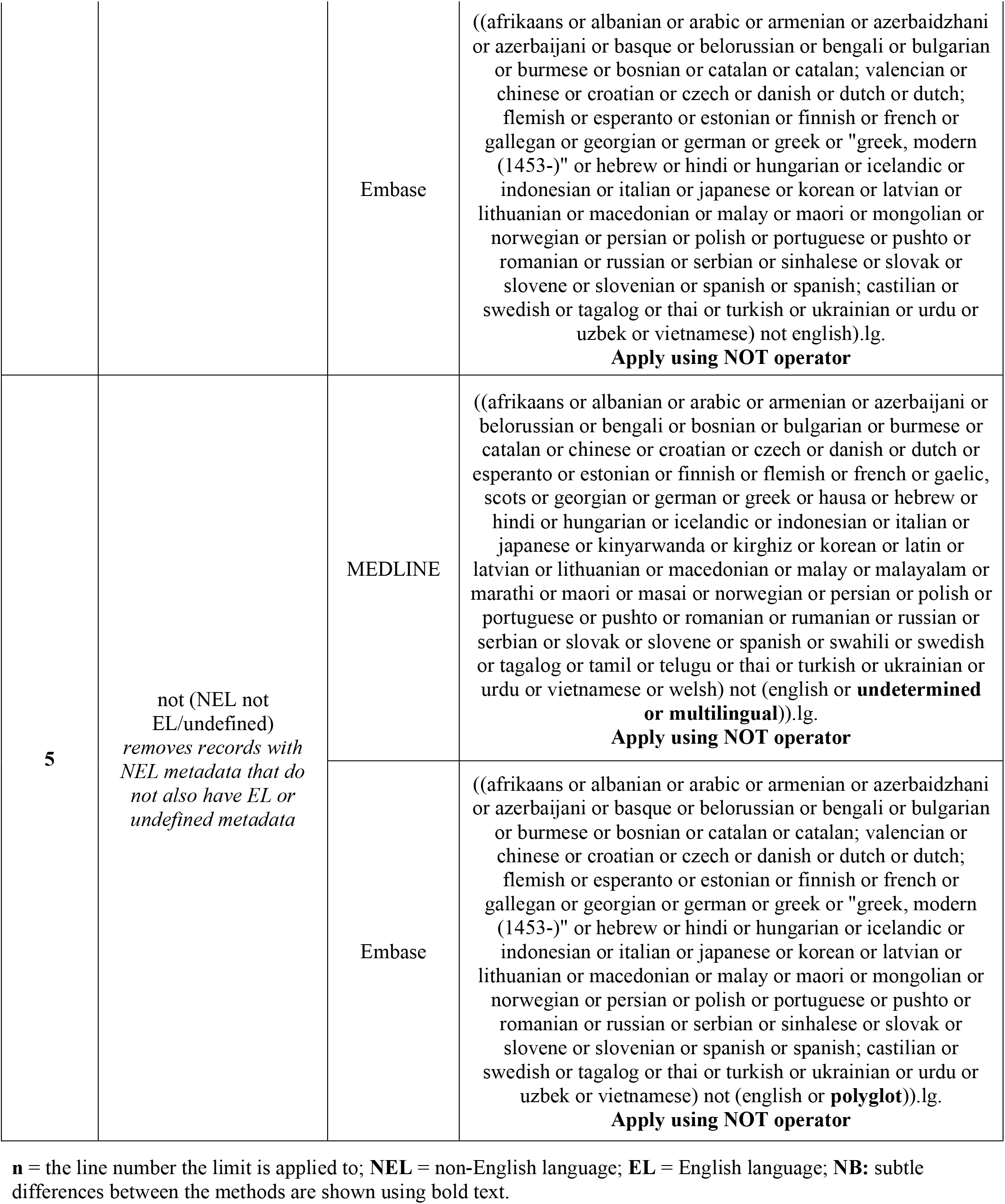
Methods of limiting to English language on Ovid MEDLINE and Embase.

The variety of languages on MEDLINE is different to those on Embase, and both databases contain postings for ‘languages’ where the language is not clearly defined. These are: ‘undetermined’, ‘multilingual’, and ‘interlingua’ on MEDLINE (which have 239,277, 1,582, and zero postings, respectively), and ‘polyglot’ on Embase (which has zero postings). Since there are various options to retrieve records with no language specified or where the language is not clearly defined, we created methods 2 and 3 to include undefined languages to see how they might compare with method 1, which limits to EL records only.

A technique developed to limit by language is the ‘not (NEL not EL)’ method, which is designed to remove records with NEL metadata that do not also have EL metadata. The method can be applied using the language field code, as shown in Table 1. Two versions of the ‘not (NEL not EL)’ method are presented for use on MEDLINE and Embase (methods 4 and 5, respectively), which were developed by browsing the selectable language options in addition to the language indexes. This was necessary as the selectable language options under ‘Additional Limits’ are not exhaustive, and there are also differences in spellings of language names. It should be noted that the languages employed in both methods 4 and 5 are not exhaustive but can be adapted as required.

In method 4, the first ‘not’ is only applied to English whereas in method 5 the first ‘not’ is applied to English or undefined languages. Method 5 was created to prevent removal of records with metadata for multiple languages, where one of the languages is an undefined language. In comparison, method 4 would remove records tagged with both a NEL and an undefined language. For both methods, the undefined languages on MEDLINE and Embase are not included alongside the terms for NELs due to their ambiguity. The language ‘interlingua’ on MEDLINE was omitted from all methods due to its zero postings. As polyglot had nine postings at the time this study was conducted, it was not omitted from methods 3 and 5 for Embase.

### Testing the performance of five English language limits

We drew on seven evidence syntheses to test how five methods of limiting to EL would have performed if they had been applied in the search strategies versus during screening [26, 27, 28, 29, 30, 31, 32]. These projects covered various healthcare topics, such as pharmaceutical interventions, mental health and neurological interventions, and public health. Datasets were obtained by appealing for evidence syntheses matching our criteria at the Centre for Reviews and Dissemination (CRD), University of York, with those used representing the number of respondents with eligible datasets. Studies were eligible if they had only restricted to EL records during screening and had used either Ovid MEDLINE, PubMed, or Ovid Embase in the searches. No geographic limits were applied in any of the search strategies or screening criteria. Dataset 4 only excluded on language where translations could not be obtained. Table 2 provides details of the datasets used.

**Table 2.** Datasets used.

| **Dataset:** | **Review:** | **Searches Last Run:** | **Date Limits:** | **Eligible Databases & Platforms Used:** | **Screening Stage 'NEL Excludes' Obtained from:** |
| --- | --- | --- | --- | --- | --- |
| **1** | Walker et al. (2024) | 2022, updated 2024 | None | Ovid MEDLINE, Ovid Embase | full text |
| **2** | Khouja et al. (2022) | 2018, updated 2021 | 2013-Current | Ovid MEDLINE, Ovid Embase | title & abstract, full text |
| **3** | Stokes et al. (2017) | 2017 | 2002-Current | Ovid MEDLINE, Ovid Embase | full text |
| 4 | Huang et al. (2023) | 2023 | None | Ovid MEDLINE | full text |
| 5 | Gallardo-Gómez et al. (2022) | 2021 | None | PubMed via NLM | title & abstract, full text |
| 6 | Gallardo-Gómez et al. (2023) | 2022 | None | PubMed via NLM | title & abstract, full text |
| 7 | Lorenc et al. (2021) | 2018 | None | Ovid MEDLINE, Ovid Embase | full text |

Seven datasets consisting of the final ‘includes’ (i.e., records which met the review’s eligibility criteria at the full text screening stage) and the ‘NEL excludes’ (i.e., records which were excluded on NEL status during screening) were created. The ‘includes’ and ‘NEL excludes’ were imported into EndNote libraries for each dataset.

For each dataset, we created four separate search strategies to find how many of the ‘includes’ and ‘NEL excludes’ were available on each database. This was achieved by searching for the title of the record as an exact phrase and combining this with additional metadata to limit to the relevant record only (where necessary). The five different methods of limiting to EL records were then applied to the search strategy (see Table 1). For methods 1, 2, and 3, we then applied the NOT operator to determine which records would be removed by each limit. For methods 4 and 5, which are applied using the NOT operator, we used the AND operator to determine which records would be removed by each limit. We calculated the percentage of ‘includes’ and ‘NEL excludes’ retrieved or removed for each methods and for each database. Results using percentages were rounded to one decimal figure.

For each dataset, the ‘includes’ and ‘NEL excludes’ that were retrieved or removed by each method for each database, were then imported into EndNote and categorised accordingly. We investigated any ‘includes’ that were removed with each method or any ‘NEL excludes’ that were retrieved, noting the language metadata and country of publication listed in the database record. Metadata were categorised as ‘correct’, ‘incorrect’, or ‘partially correct’, and were considered ‘partially correct’ if at least one of the languages listed in the metadata was confirmed to be correct by viewing the full text(s). We also made notes on whether articles appeared in journals publishing in single or multiple languages and whether English was the language of the abstract only and not the full text.

The same databases and segments on the Ovid platform were used when testing each dataset. Full search strategies are available in the appendix.

## RESULTS

### Testing the performance of five English language limits

Table 3 shows the number of ‘includes’ and ‘NEL excludes’ that were available on MEDLINE and Embase respectively out of the total number of ‘includes’ and ‘NEL excludes’ tested for each project.

**Table 3.**
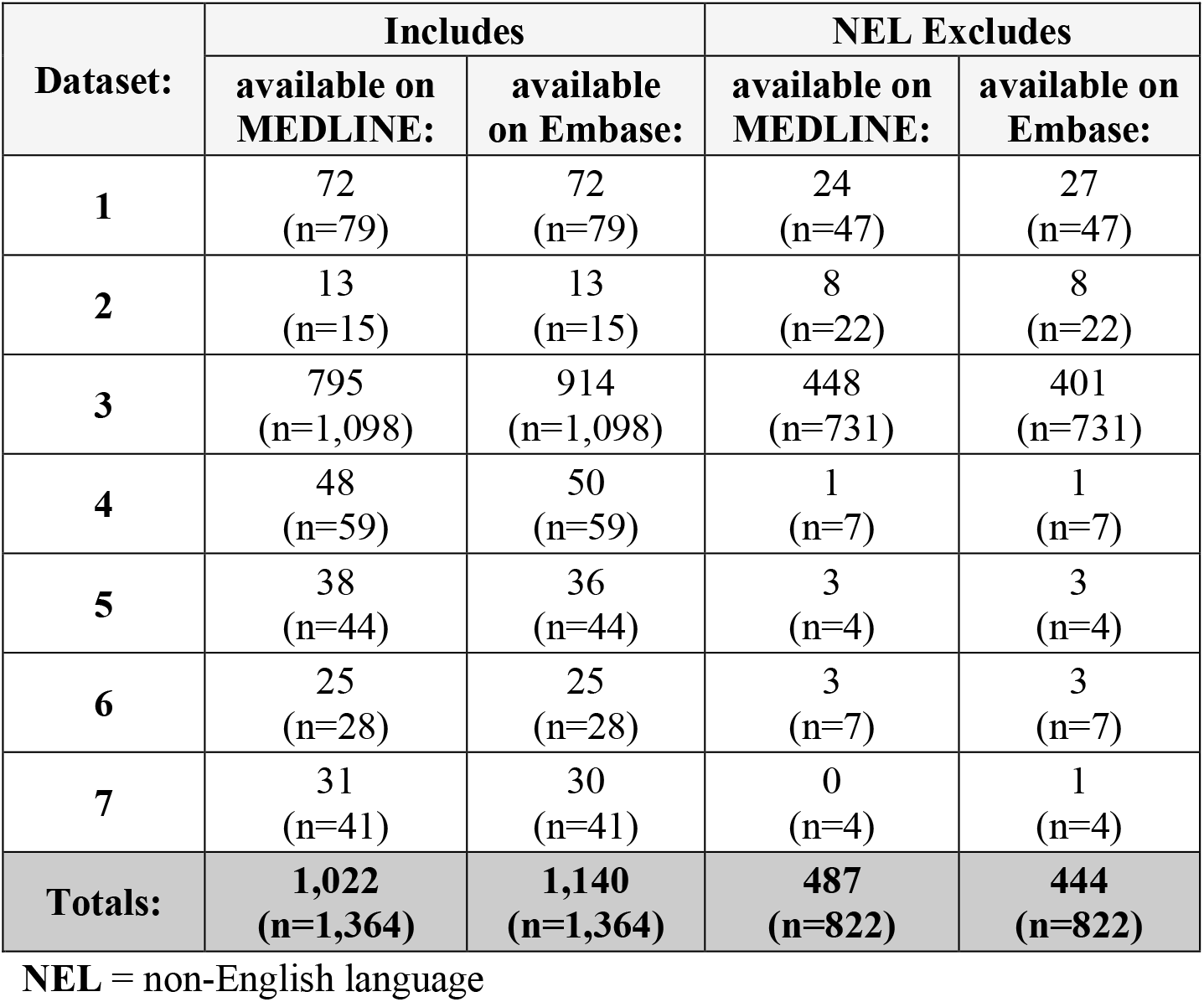
Number of ‘includes’ and ‘NEL excludes’ available on MEDLINE and Embase.

Table 4 shows the number of ‘includes’ and ‘NEL excludes’ retrieved or removed by each method on MEDLINE and Embase respectively, out of the total number available on each database.

**Table 4.** Number of ‘includes’ and ‘NEL excludes’ retrieved or excluded.

| Method: | Includes |  |  |  | NEL Excludes |  |  |  |
| --- | --- | --- | --- | --- | --- | --- | --- | --- |
|  | retrieved<br>on<br>MEDLINE | removed<br>on<br>MEDLINE | retrieved<br>on<br>Embase | removed<br>on<br>Embase | removed<br>on<br>MEDLINE | retrieved<br>on<br>MEDLINE | removed<br>on<br>Embase | retrieved<br>on<br>Embase |
|  | (N=1,022) |  | (N=1,140) |  | (N=487) |  | (N=444) |  |
| **1** | 1,020<br>(99.8%) | 2<br>(0.2%) | 1,137<br>(99.7%) | 3<br>(0.3%) | 478<br>(98.2%) | 9<br>(1.8%) | 420<br>(94.6%) | 24<br>(5.4%) |
| **2** | 1,020<br>(99.8%) | 2<br>(0.2%) | 1,137<br>(99.7%) | 3<br>(0.3%) | 478<br>(98.2%) | 9<br>(1.8%) | 420<br>(94.6%) | 24<br>(5.4%) |
| **3** | 1,020<br>(99.8%) | 2<br>(0.2%) | 1,137<br>(99.7%) | 3<br>(0.3%) | 478<br>(98.2%) | 9<br>(1.8%) | 420<br>(94.6%) | 24<br>(5.4%) |
| **4** | 1,020<br>(99.8%) | 2<br>(0.2%) | 1,137<br>(99.7%) | 3<br>(0.3%) | 478<br>(98.2%) | 9<br>(1.8%) | 420<br>(94.6%) | 24<br>(5.4%) |
| **5** | 1,020<br>(99.8%) | 2<br>(0.2%) | 1,137<br>(99.7%) | 3<br>(0.3%) | 478<br>(98.2%) | 9<br>(1.8%) | 420<br>(94.6%) | 24<br>(5.4%) |

**NEL** = non-English language; **NB:** ‘includes’ removed or ‘NEL excludes’ retrieved are highlighted in dark grey

On each database respectively, all methods performed identically, retrieving and removing the same records. For all methods, 99.8% of ‘includes’ on MEDLINE were retrieved compared with 99.7% on Embase. For all methods, 98.2% ‘NEL excludes’ on MEDLINE were removed compared with 94.6% on Embase.

For each ‘include’ removed on MEDLINE and Embase, Table 5 shows the language metadata, country of publication, and confirmed language(s). It reports on whether methods 1-5 removed the record correctly or incorrectly, the accuracy of the language metadata, and provides further notes on the journal and record. All methods removed the same records.

**Table 5.** ‘Includes’ removed:

| Database: | Language Metadata: | Country of Publication Metadata: | Confirmed Language(s): | Removed by Limits: | Language Metadata Accuracy: | Notes: |
| --- | --- | --- | --- | --- | --- | --- |
| **MEDLINE** | Spanish | Spain | Spanish, English | **Incorrectly** | **Partially correct** | Journal is bilingual, article is in multiple languages |
|  | Polish | Poland | Polish, English | **Incorrectly** | **Partially correct** | Journal is bilingual, article is in multiple languages |
| **Embase** | Polish | Poland | Polish, English | **Incorrectly** | **Partially correct** | Journal is bilingual, article is in multiple languages |
|  | Portuguese | Brazil | Portuguese, Spanish, English | **Incorrectly** | **Partially correct** | Journal is multilingual, article is in multiple languages |
|  | Spanish | Brazil | Portuguese, Spanish, English | **Incorrectly** | **Partially correct** | Journal is multilingual, article is in multiple languages |

**NB:** records incorrectly removed or with incorrect or partially correct metadata are highlighted in bold.

Only 2 of 1,022 (0.2%) ‘includes’ were removed on MEDLINE using methods 1-5, compared with 3 of 1,140 (0.3%) on Embase. These five records were all incorrectly removed by the limits (due to availability in English). The metadata for all five records was only partially correct.

For each ‘NEL exclude’ retrieved on MEDLINE and Embase, Table 6 shows the language metadata, country of publication, and confirmed language(s). It reports on whether methods 1-5 retrieved the record correctly or incorrectly, the accuracy of the language metadata, and provides further notes on the journal and record. All methods retrieved the same records.

**Table 6.** ‘NEL Excludes’ retrieved.

| **Database:** | **Language Metadata:** | **Country of Publication Metadata:** | **Confirmed Language(s):** | **Retrieved by Limits:** | **Language Metadata Accuracy:** | **Notes:** |
| --- | --- | --- | --- | --- | --- | --- |
| **MEDLINE** | English | France | English, French | Correctly | **Partially correct** | Journal is bilingual, article is in multiple languages |
|  | English French | England | English, French | Correctly | Correct | Journal is bilingual, article is in multiple languages |
|  | English | United States | English | Correctly | Correct | English language journal |
|  | English | Poland | English | Correctly | Correct | English language journal |
|  | English Norwegian | Norway | English, Norwegian | Correctly | Correct | Journal is bilingual, article is in multiple languages |
|  | English | Poland | Polish | **Incorrectly** | **Incorrect** | Polish language journal with English abstracts only |
|  | English Spanish | Colombia | English | Correctly | **Partially correct** | Journal is bilingual, article is only in English |
|  | English Turkish | Turkey | English, Turkish | Correctly | Correct | Journal is bilingual, article is in multiple languages |
|  | English | China (Republic: 1949- ) | English | Correctly | Correct | English language journal |
| **Embase** | English | Taiwan (Republic of China) | English | Correctly | Correct | English language journal |
|  | English | Croatia | Croatian | **Incorrectly** | **Incorrect** | Croatian language journal with English abstracts only |
|  | English, French | France | English, French | Correctly | Correct | Journal is bilingual, article is in multiple languages |
|  | English, French | France | French | **Incorrectly** | **Partially correct** | French language journal with English abstracts only |
|  | English, French | Canada | English, French | Correctly | Correct | Journal is bilingual, article is in multiple languages |
|  | English, Polish | Poland | Polish | **Incorrectly** | **Partially correct** | Polish language journal with English abstracts only |
|  | English, Czech | Czechia | Czech | **Incorrectly** | **Partially correct** | Bilingual journal with English abstracts only |
|  | English, Czech | Czechia | Czech | **Incorrectly** | **Partially correct** | Bilingual journal with English abstracts only |
|  | English | Bulgaria | English | Correctly | Correct | English language journal |
|  | English, Turkish | Turkey | Turkish | **Incorrectly** | **Partially correct** | Journal is bilingual, article is only in Turkish |
|  | English, Czech | Czechia | Czech | **Incorrectly** | **Partially correct** | Multilingual journal with English abstracts only |
|  | English, Turkish | Turkey | Turkish | **Incorrectly** | **Partially correct** | Journal is bilingual, article is only in Turkish |
|  | German, English | Germany | German | **Incorrectly** | **Partially correct** | Journal is bilingual, article is only in German |
|  | English | Poland | English | Correctly | Correct | English language journal |
|  | English, Polish | Poland | Polish | **Incorrectly** | **Partially correct** | Journal is bilingual, article is only in Polish |
|  | English, French | Nigeria | French | **Incorrectly** | **Partially correct** | Journal is bilingual, article is only in French |
|  | English, Norwegian | Norway | English, Norwegian | Correctly | Correct | Journal is bilingual, article is in multiple languages |
|  | English, German | Switzerland | German | **Incorrectly** | **Partially correct** | Bilingual journal article with English abstracts only |
|  | English, Portuguese | Portugal | Portuguese | **Incorrectly** | **Partially correct** | Journal is bilingual, article is only in Portuguese |
|  | English, Russian | Russian Federation | Russian | **Incorrectly** | **Partially correct** | Russian language journal with English abstracts only |
|  | English | Poland | Polish | **Incorrectly** | **Incorrect** | Polish language journal with English abstracts only |
|  | English, Spanish | Colombia | English | Correctly | **Partially correct** | Journal is bilingual, article is only in English |
|  | English, Turkish | Turkey | English, Turkish | Correctly | Correct | Journal is bilingual, article is in multiple languages |
|  | English | Netherlands | English | Correctly | Correct | English language journal |

**NB:** records incorrectly retrieved or with incorrect or partially correct metadata are highlighted in bold.

On MEDLINE, eight out of nine records were correctly retrieved by methods 1-5 due to their availability in English, with one record retrieved incorrectly due to incorrect metadata. On Embase, nine out of 24 records were correctly retrieved by methods 1-5 due to their availability in English, with 15 records retrieved incorrectly due to incorrect or partially correct metadata (where English was not listed).

## DISCUSSION

### Methods of limiting by language on Ovid MEDLINE and Embase

Any method of restricting by language on Ovid MEDLINE or Embase which aims to be more sensitive than limiting to a single language, could benefit from careful checking of the available language options and the number of postings for several reasons. Firstly, there are differences between the selectable language options under ‘Additional Limits’ and those listed in the language indexes on MEDLINE and Embase, with items in each list not represented in the other. As an example, ‘interlingua’ is a selectable language on MEDLINE which does not appear in the language index and has zero postings. This highlights that both sources should be consulted for users who want to understand all languages that are available and whether these are worth applying. Secondly, where language names have alternative spellings, the application of inbuilt limits can produce different results compared with using the language field code on both databases. For example, *selecting* the language ‘Rumanian’ on MEDLINE will also limit to records listed as ‘Romanian’, but *searching* for ‘Rumanian’ with the language field code brings back zero postings, as Romanian.lg. or rum.lg. must be used. Likewise, on Embase, *selecting* ‘Azerbaidzhani’ will also limit to records listed as ‘Azerbaijani’, but ‘Azerbaidzhani’ has zero postings if searched with the language field code, as Azerbaijani.lg. must be used.

It is also worth understanding the relevance of some of the ambiguous languages on the databases. Many of the 239,277 MEDLINE records with ‘undetermined’ listed as the language are from the OLDMEDLINE subset of PubMed and come from print indexes which date from 1946-1959, though records with this language span from 1940-2002 [21, 33, 34]. This choice of language is therefore only relevant for strategies without date limits or with date limits before 2003.

For users looking to be sensitive in their application of EL limits, it is more straightforward to limit to: ‘English, undetermined, multilingual, or no language specified’ (method 3) on MEDLINE, or ‘English or no language specified or polyglot’ (method 3) on Embase, in comparison with methods 4 and 5. This circumvents the issue of different NELs (or records with different spellings for languages) being retrieved in search results where these have not been excluded. As there are now zero postings in polyglot on Embase but were nine at the time this study was conducted, there may be no difference between methods 2 and 3 for Embase.

### The performance of five English language limits on Ovid MEDLINE and Embase

Despite the identical performance of the five EL limits for the datasets tested in this paper, the methods may not always perform identically. Data showing methods 1-5 have the potential to retrieve and remove different records from each other can be found in the supplementary material.

The finding that screening decisions were consistent with the language metadata for 1,020 of 1,022 (99.8%) ‘includes’ on MEDLINE and 1,137 of 1,140 (99.7%) ‘includes’ on Embase might lead us to believe there is minimal risk of removing EL records with the use of language limits. However, this is not a complete picture of the data as it is possible that EL records might have been incorrectly excluded on NEL status during screening and have incorrect language metadata. Moreover, the removal of any records in English that meet a review’s eligibility criteria highlights the potential risks of applying EL limits within the search strategies.

A total of five ‘includes’ were removed with methods 1-5: two on MEDLINE (0.2%) and three on Embase (0.3%), as shown in Table 5. All five records were incorrectly removed. In all cases, the language metadata was only partially correct – correctly listing a single NEL language which caused it to be removed with the use of the EL limits. All five articles (100%) were published in English and additional languages in bilingual or multilingual journals.

As the language metadata for MEDLINE and Embase originates from different sources, searching both databases might reduce the risk of removing EL records, since metadata errors might be unique to a single database. Language metadata can differ for the same article, as shown by a record listed as Scottish Gaelic on MEDLINE but Irish Gaelic on Embase [35]. However, the finding that one of the ‘includes’ in this study was incorrectly removed on both MEDLINE and Embase is interesting and shows that language metadata can be incorrect for the same article on both databases.

The ‘NEL excludes’ that were retrieved with the use of the EL limits (nine on MEDLINE and 24 on Embase) are more nuanced to analyse (see Table 6). On MEDLINE, eight out of nine ‘NEL excludes’ were correctly retrieved by methods 1-5 due to their availability in English, with the remaining one record retrieved incorrectly due to incorrect metadata. The single record that was retrieved incorrectly was in a NEL journal with English abstracts only.

On Embase, nine out of 24 ‘NEL excludes’ were correctly retrieved by methods 1-5 due to their availability in English, with 15 records retrieved incorrectly due to incorrect or partially correct metadata (where English was not listed). The 15 records retrieved incorrectly provide further evidence of incorrect language metadata. Of these 15 records: six were in bilingual/multilingual journals with the article only in a NEL (40%); five were in NEL journals with English abstracts only (33.3%); and four were in bilingual/multilingual journal with English abstracts only (26.6%). This may suggest that language metadata may be more likely to be inaccurate for bilingual/multilingual journals, or for single-language publications with abstracts in other languages, in comparison with journals publishing in a single language only. Nussbaumer-Streit et al have noted that as some studies can be published in both English and NELs, excluding NEL publications does not necessarily result in missing a study [7]. In the context of excluding NEL records within the search strategies however, Table 5 shows that where publications are available in multiple languages this is not always accurately reflected in the language metadata. It should be noted that where EL limits are used, a small number of NEL records may still have to be excluded during screening.

Using EL limits on Ovid MEDLINE and Embase may only have a small impact on the screening burden compared with removing NEL records during screening (though this will vary by topic). However, as the screening burden can affect the quality of reviews [36], it is important to ensure the volume of search results are manageable for review teams. Therefore, the use of EL limits in search strategies could be pragmatic where review teams know they will be unable to translate papers or where this is not necessary (e.g., only studies published in English are relevant). Review teams could decide whether to apply EL limits in search strategies based on the number of NEL records overall, which the information specialist could report on before searches are run.

### Removing NEL records during screening

Articles can be incorrectly excluded on NEL status during screening. Several ‘NEL excludes’ that were retrieved with the use of the EL limits were found to have been correctly retrieved due to their availability in English (eight on MEDLINE and nine on Embase).

On MEDLINE, Table 6 shows that of the eight articles correctly retrieved: four were in bilingual journals where the article was in English and other languages (50%); three were in EL journals (37.5%); and one was in a bilingual journal where the article was only in English (12.5%). On Embase, of the nine records correctly retrieved, four were in EL journals (44.4%); four were in bilingual journals where the article was in multiple languages (44.4%); and one was in a bilingual journal where the article was only available in English (11.1%). Across both databases and individually, most of these records were in bilingual/multilingual journals, though a significant proportion were in EL journals. This suggests that for reviews excluding on NEL status during screening, articles should be carefully checked for their availability in multiple languages. In some instances, there may be different digital object identifiers (DOIs) or different page numbers for records in multiple languages, as found for one record available in English [37] and French [38] which was excluded during screening. However, it is worth noting that NEL records excluded during screening might also be excluded on other criteria. Notably, none of the records found to have been incorrectly excluded on NEL status during screening would have met the inclusion criteria of the evidence syntheses they originated from. As language can be among the earliest criteria used to exclude studies during screening [39], review teams could consider moving it further down the list of eligibility criteria, so that fewer records are excluded on language alone.

Of the records published in EL journals that were excluded on NEL status during screening (three on MEDLINE, four on Embase), only two records were published in countries which speak English as a first language, according to the country of publication metadata. It is possible this factor could have contributed to their incorrect exclusion, though human error such as selecting the wrong exclusion criteria is also possible.

As noted previously, there are advantages to not applying language limits in the search strategies. This method allows transparent reporting of records available in other languages and gives research teams the option to translate articles if time permits or circumstances change [15]. A survey study by Neimann Rasmussen and Montgomery found the lack of resources to translate studies was the second most common challenge to including NEL studies [4]. However, it is possible for review teams to ask colleagues to translate articles or use free tools such as Google Translate [40] or DeepL Translate [41], so records could be excluded on language only where translation is not possible. As numerous strategies to help researchers include NEL records have been published [42, 43, 44], information specialists and review teams could factor these into their decisions on whether to exclude on NEL status. However, for reviews with limited resources interested in trials as a source of evidence, a study by Egger et al found that assessing the quality of evidence from trials may be more important than retrieving or translating them, since difficult to locate trials were found to be of lower quality which could introduce bias [45].

The risk of excluding potentially eligible records due to language restrictions (whether applied during the strategies or screening) could be mitigated with forward and backward citation searching. This could reduce the risk of missing EL or eligible records with incorrect language metadata or excluding them during screening. Review teams with constraints on time or other resources could consider including and translating NEL records from citation searches as an alternative to restricting on language altogether.

Where review teams decide to include NEL records and are using ‘priority screening’ on systematic review software such as EPPI-Reviewer (a feature where machine-learning uses screening decisions to re-order the screening list by relevance) [46], they should be aware that records without an EL title and abstract may end up being screened last [47]. Review teams using priority screening and applying ‘stopping criteria’ (a rule where screening is stopped if no new ‘includes’ have been found after a set number of records) [48], should therefore screen NEL records separately [47].

### LIMITATIONS

While this study is based on data readily available to the authors, it demonstrates the implications of limiting to EL within the search strategies in comparison to removing NEL records during screening.

1,509 MEDLINE records and 1,584 Embase records taken from healthcare evidence syntheses were tested. Although only two databases were investigated, Table 3 shows that 74.9% of ‘includes’ and 59.2% of ‘NEL excludes’ from the seven evidence syntheses were available on MEDLINE, whereas 83.6% of ‘includes’ and 54% of ‘NEL excludes’ were available on Embase. This demonstrates the value of understanding the impact of language limits on these important and widely used databases.

Searching for the ‘includes’ and ‘NEL excludes’ using the title of the record could have introduced minor errors.

For some of the evidence syntheses (three of seven), ‘NEL excludes’ were obtained from full text screening only, as opposed to from title and abstract screening and full text screening (four of seven).

For all datasets, the language status of records included or excluded by reviewers during screening was not double-checked for accuracy (except where ‘includes’ were removed or ‘NEL excludes’ were retrieved with the use of language limits). It should also be noted that not all language metadata will be accurate.

The seven evidence syntheses used covered various healthcare topics: pharmaceutical interventions, mental health and neurological interventions, and public health. As previous evidence has shown, the impact of language limits may vary for alternative or complementary medicine and where evidence on the intervention or topic is highly likely to be published in NELs.

## CONCLUSION

As this study has explored, limiting database records to English on Ovid MEDLINE and Embase can be achieved with various methods. For review teams restricting by language within the search strategies, it may be worth checking if any results have no language specified or are in undefined languages. This could reduce the risk of removing potentially eligible records (except where language metadata is incorrect). Although methods 1-5 performed identically on the datasets tested, understanding the potential impact of these methods in terms of their design and the languages they retrieve and exclude, facilitates informed decision making by information specialists and review teams.

Overall, 0.2% ‘includes’ on MEDLINE and 0.3% of ‘includes’ on Embase were incorrectly removed with methods 1-5 due to incorrect language metadata. These low numbers could suggest that it is pragmatic to apply language limits in the search strategies as there may be minimal risk of removing potentially eligible records in English. As the five ‘includes’ that were removed with methods 1-5 came from two of the seven evidence syntheses, the remaining five reviews would still have found all ‘includes’ on MEDLINE and Embase if they had applied EL limits in the search strategies. However, as the risk of removing ‘includes’ could skew or prejudice the findings, review teams should carefully consider methods of mitigating this risk (e.g., citation searching). These incorrectly removed ‘includes’ may suggest that more accurate language metadata is needed for articles in bilingual/multilingual journals.

Our investigation of the ‘NEL excludes’ retrieved found that several EL records were incorrectly excluded during screening (eight on MEDLINE, nine on Embase), though these records were not eligible for inclusion in the evidence syntheses based on other exclusion criteria. This highlights the value of detailed checking of the availability of articles in multiple languages.

Overall, our findings are informative for information specialists and review teams to understand the impact of restricting to EL within the search strategies in comparison with screening. Reviews with constraints on resources should consider the various strategies to include NEL records [42, 43, 44] and weigh up whether their inclusion could reduce bias or reduce the quality of evidence [45]. Studies that decide to limit by language should consider the different risks of removing EL records within the search strategies versus during screening. Where used, language restrictions should be justifiable and reported transparently.

Further research could explore the accuracy of language metadata on other databases, and how much the screening burden could be reduced using language limits across a wider selection of databases.

## Supporting information

Appendix - Search Strategies

## DATA AVAILABILITY STATEMENT

Data associated with this article are available in the appendix and supplementary material or can be obtained by contacting the first author.

## ACKNOWLEDGEMENTS

We would like to acknowledge NLM Support, Elsevier Customer Care, and Ovid’s Customer Success Team and their Customer Engagement Team for their time and help answering our enquiries.

## CONFLICT OF INTEREST STATEMENT

The authors have no competing interests to report.

## SOURCES OF FUNDING STATEMENT

The authors have no sources of funding to report.

## ETHICS STATEMENT

Ethical approval was not necessary for this study.

## PEER REVIEW STATEMENT

This study was submitted to a journal in August 2024 and has received two rounds of peer reviews at the time of submission to medRxiv. All data was checked and was up to date at the time of submission to medRxiv.

## Notes

### Competing Interest Statement

The authors have declared no competing interest.

