## Appendix - Search Strategies for "Limiting to English language records: A comparison of five methods on Ovid MEDLINE and Embase versus removal during screening"

### **Dataset 1:**

**Includes: MEDLINE® ALL** via Ovid http://ovidsp.ovid.com/

Date range searched: <1946 to January 24, 2024>

Date searched: 25 January 2024

1	"Addition of 3-day aprepitant to ondansetron and dexamethasone for prophylaxis of chemotherapy-induced nausea and vomiting among patients with diffuse large b cell lymphoma receiving 5-day cisplatin-based chemotherapy".ti. (1)

2	"Randomized double-blind crossover ondansetron-dexamethasone versus ondansetron-placebo study for the treatment of chemotherapy-induced nausea and vomiting in pediatric patients with malignancies".ti. (1)

3	"The antiemetic efficacy and safety of granisetron compared with metoclopramide plus dexamethasone in patients receiving fractionated chemotherapy over 5 days. The granisetron study group".ti. (1)

4	"Aprepitant as an add-on therapy in children receiving highly emetogenic chemotherapy: A randomized, double-blind, placebo-controlled trial".ti. (1)

5	"A double-blind, crossover, randomized dose-comparison trial of granisetron for the prevention of acute and delayed nausea and emesis in children receiving moderately emetogenic carboplatin-based chemotherapy".ti. (1)

6	"An increased loading dose of ondansetron: A north european, double-blind randomised study in children, comparing 5 mg/m2 with 10 mg/m2".ti. (1)

7	"Cardiac effects of granisetron in a prospective crossover randomized dose comparison trial".ti. (1)

8	"Palonosetron is a better choice compared with ondansetron for the prevention of chemotherapy-induced nausea and vomiting (cinv) in a resource-limited pediatric oncology center: Results from a randomized control trial".ti. (1)

9	"The control of acute cisplatin-induced emesis--a comparative study of granisetron and a combination regimen of high-dose metoclopramide and dexamethasone. Granisetron study group".ti. (1)

10	"A randomized trial of individualized versus standard of care antiemetic therapy for breast cancer patients at high risk for chemotherapy-induced nausea and vomiting".ti. (1)

11	"Randomised comparison of ondansetron and metoclopramide plus dexamethasone for chemotherapy induced emesis".ti. (1)

12	"Palonosetron for prevention of acute and delayed nausea and vomiting in non-small-cell lung carcinoma patients".ti. (1)

13	"A randomised, double-blind, parallel-group study to compare the efficacy and safety of ondansetron (gr38032f) plus dexamethasone with metoclopramide plus dexamethasone in the prophylaxis of nausea and emesis induced by carboplatin chemotherapy".ti. (1)

14	"Evaluating aprepitant single-dose plus granisetron and dexamethasone in children receiving highly emetogenic chemotherapy for the prevention of chemotherapy-induced nausea and vomiting: A triple-blinded randomized clinical trial".ti. (1)

15	"Granisetron, tropisetron, and ondansetron in the prevention of acute emesis induced by a combination of cisplatin-adriamycin and by high-dose ifosfamide delivered in multiple-day continuous infusions".ti. (1)

16	"Double-blind comparative trial of oral ondansetron versus oral granisetron versus iv ondansetron in the prevention of nausea and vomiting associated with highly emetogenic preparative regimens prior to stem cell transplantation".ti. (1)

17	"Ondansetron versus granisetron in the prevention of chemotherapy-induced nausea and vomiting. Results of a prospective randomized trial".ti. (1)

18	"Comparing different antiemetic regimens for chemotherapy induced nausea and vomiting".ti. (0)

19	"Aprepitant in adolescent patients for prevention of chemotherapy-induced nausea and vomiting: A randomized, double-blind, placebo-controlled study of efficacy and tolerability".ti. (1)

20	"Palonosetron improves prevention of chemotherapy-induced nausea and vomiting following moderately emetogenic chemotherapy: Results of a double-blind randomized phase iii trial comparing single doses of palonosetron with ondansetron".ti. (1)

21	"Single-dose oral granisetron has equivalent antiemetic efficacy to intravenous ondansetron for highly emetogenic cisplatin-based chemotherapy".ti. (1)

22	"Single-dose fosaprepitant for the prevention of chemotherapy-induced nausea and vomiting associated with cisplatin therapy: Randomized, double-blind study protocol--ease".ti. (1)

23	"A single-blind comparison of intravenous ondansetron, a selective serotonin antagonist, with intravenous metoclopramide in the prevention of nausea and vomiting associated with high-dose cisplatin chemotherapy".ti. (1)

24	"Efficacy and tolerability of aprepitant for the prevention of chemotherapy-induced nausea and emesis over multiple cycles of moderately emetogenic chemotherapy".ti. (1)

25	"The oral neurokinin-1 antagonist aprepitant for the prevention of chemotherapy-induced nausea and vomiting: A multinational, randomized, double-blind, placebo-controlled trial in patients receiving high-dose cisplatin--the aprepitant protocol 052 study group".ti. (1)

26	"The effect of palonosetron hydrochloride in the prevention of chemotherapy-induced moderate and severe nausea and vomiting".ti. (1)

27	"Randomized, double-blind, placebo-controlled study of aprepitant versus two dosages of olanzapine with ondansetron plus dexamethasone for prevention of chemotherapy-induced nausea and vomiting in patients receiving high-emetogenic chemotherapy".ti. (1)

28	"A randomized, open-label non-inferiority study to compare palonosetron and ondansetron for prevention of acute chemotherapy-induced vomiting in children with cancer receiving moderate or high emetogenic chemotherapy".ti. (1)

29	"Single-dose oral granisetron versus multidose intravenous ondansetron for moderately emetogenic cyclophosphamide-based chemotherapy in pediatric outpatients with acute lymphoblastic lukemia".ti. (0)

30	"A comparison of ondansetron with metoclopramide in the prophylaxis of chemotherapy-induced nausea and vomiting: A randomized, double-blind study. International emesis study group".ti. (1)

31	"Aprepitant for the prevention of chemotherapy-induced nausea and vomiting in paediatric subjects: An analysis by age group".ti. (1)

32	"Aprepitant for the prevention of chemotherapy-induced nausea and vomiting in children: A randomised, double-blind, phase 3 trial".ti. (1)

33	"Clinical evaluation of two antiemetic combinations palonosetron dexamethasone versus ondansetron dexamethasone in chemotherapy of head and neck cancer".ti. (1)

34	"Efficacy and safety of aprepitant for the prevention of chemotherapy-induced nausea and vomiting during the first cycle of moderately emetogenic chemotherapy in korean patients with a broad range of tumor types".ti. not published erratum/ (1)

35	"A randomised dose-comparison trial of granisetron in preventing emesis in children with leukaemia receiving emetogenic chemotherapy".ti. (1)

36	"Effectiveness of oral ondansetron in the management of nausea and vomiting induced by moderately emetogenic chemotherapy".ti. (0)

37	"Comparison of the efficacy and side-effects of ondansetron and metoclopramide-diphenhydramine administered to control nausea and vomiting in children treated with antineoplastic chemotherapy: A prospective randomized study".ti. (1)

38	"Palonosetron compared with ondansetron in pediatric cancer patients: Multicycle analysis of a randomized phase iii study".ti. (1)

39	"Palonosetron versus ondansetron for prevention of chemotherapy-induced nausea and vomiting in paediatric patients with cancer receiving moderately or highly emetogenic chemotherapy: A randomised, phase 3, double-blind, double-dummy, non-inferiority study".ti. (1)

40	"Ondansetron compared with dexamethasone and metoclopramide as antiemetics in the chemotherapy of breast cancer with cyclophosphamide, methotrexate, and fluorouracil".ti. (1)

41	"Does a single dose of palonosetron have any role in preventing acute chemotherapy-induced nausea and vomiting in pediatric osteosarcoma patients without dexamethasone? A randomized clinical trial".ti. (1)

42	"Contribution to the treatment of nausea and emesis induced by chemotherapy in children and adolescents with osteosarcoma".ti. (1)

43	"A prospective triple-blind randomized trial on safety and efficacy of abitant in the prevention of chemotherapy-induced nausea and vomiting".ti. (0)

44	"An open randomised cross-over study on granisetron versus ondansetron in the prevention of acute emesis induced by moderate dose cisplatin-containing regimens".ti. (1)

45	"Daily palonosetron is superior to ondansetron in the prevention of delayed chemotherapy-induced nausea and vomiting in patients with acute myelogenous leukemia".ti. (1)

46	"Comparison of acute and delayed antiemetic effect of adding aprepitant to odansitron and dexamethasone regimen in patients with hodgkin lymphoma receiving highly emetogenic chemotherapy".ti. (0)

47	"Efficacy and safety of olanzapine in children receiving highly emetogenic chemotherapy: A randomized, double-blind placebo-controlled phase 3 trial".ti. (1)

48	"Triplet aprepitant/dexamethasone/ondansetron versus doublet dexamethasone/ondansetron for prevention of moderately emetogenic chemotherapy: A placebo-controlled, double-blind, randomized clinical trial of efficacy".ti. (0)

49	"Efficacy and safety of adding olanzapine to the standard preventive regimen for chemotherapy-induced nausea and vomiting in children: A randomized double-blind controlled trial".ti. (1)

50	"Olanzapine for prevention of vomiting in children and adolescents receiving highly emetogenic chemotherapy: Investigator-initiated, randomized, open-label trial".ti. (1)

51	"Comparative clinical trial of granisetron and ondansetron in the prophylaxis of cisplatin-induced emesis. The granisetron study group".ti. (1)

52	"A double-blind, randomised, crossover comparison of granisetron and ondansetron in 5-day fractionated chemotherapy: Assessment of efficacy, safety and patient preference. The granisetron study group".ti. (1)

53	"A comparative study of intravenous granisetron versus intravenous and oral ondansetron in the prevention of nausea and vomiting associated with moderately emetogenic chemotherapy".ti. (1)

54	"Randomized open-label phase ii trial of 5-day aprepitant plus ondansetron compared to ondansetron alone in the prevention of chemotherapy-induced nausea-vomiting (cinv) in glioma patients receiving adjuvant temozolomide".ti. not letter/ (1)

55	"Comparison of antiemetic efficacy and safety of palonosetron vs ondansetron in the prevention of chemotherapy-induced nausea and vomiting in children".ti. (1)

56	"Addition of the neurokinin 1 receptor antagonist aprepitant to standard antiemetic therapy improves control of chemotherapy-induced nausea and vomiting. Results from a randomized, double-blind, placebo-controlled trial in latin america".ti. (1)

57	"Intravenous fosaprepitant for the prevention of chemotherapy induced vomiting in children, a randomized, phase 3 trial".ti. (0)

58	"Aprepitant for the prevention of chemotherapy-induced nausea and vomiting associated with a broad range of moderately emetogenic chemotherapies and tumor types: A randomized, double-blind study".ti. (1)

59	"Ondansetron compared with granisetron in the prophylaxis of cisplatin-induced acute emesis: A multicentre double-blind, randomised, parallel-group study".ti. (1)

60	"Single daily dosing versus divided dosing intravenous ondansetron to prevent chemotherapy-induced nausea and vomiting among children: A comparative randomized double-blind controlled trial".ti. (1)

61	"Randomized double-blind comparison of single high-dose ondansetron and multiple standard-dose ondansetron in chemotherapy-naive pediatric oncology patients".ti. (1)

62	"Comparison of an aprepitant regimen with a multiple-day ondansetron regimen, both with dexamethasone, for antiemetic efficacy in high-dose cisplatin treatment".ti. (1)

63	"Palonosetron hydrochloride is an effective and safe option to prevent chemotherapy-induced nausea and vomiting in children".ti. (1)

64	"Addition of aprepitant improves acute emesis control in children and adolescents receiving induction chemotherapy for acute myeloid leukaemia: A randomised, open-label trial".ti. (1)

65	"Ondansetron versus granisetron in the prevention of chemotherapy induced nausea and vomiting in children with acute lymphoblastic leukemia".ti. (1)

66	"Ondansetron compared with metoclopramide in the control of emesis and quality of life during repeated chemotherapy for breast cancer".ti. (1)

67	"A comparison of oral ondansetron and intravenous granisetron for the prevention of nausea and emesis associated with cisplatin-based chemotherapy".ti. (1)

68	"Ondansetron and tropisetron in the control of nausea and vomiting in children receiving combined cancer chemotherapy".ti. (1)

69	"Prevention of nausea and vomiting associated with stem cell transplant: Results of a prospective, randomized trial of aprepitant used with highly emetogenic preparative regimens".ti. (1)

70	"Palonosetron is nonsuperior to ondansetron in acute phase but provides superior antiemetic control in delayed phase for pediatric patients administered highly emetogenic chemotherapy".ti. (1)

71	"A randomized, double-blind, placebo-controlled study evaluating the efficacy of combination olanzapine, ondansetron and dexamethasone for prevention of chemotherapy-induced nausea and vomiting in patients receiving doxorubicin plus cyclophosphamide".ti. (1)

72	"Effects of granisetron in children undergoing high-dose chemotherapy: A multi-institutional, cross-over study".ti. (1)

73	"The efficacy and safety of the addition of olanzapine to ondansetron and dexamethasone for prevention of chemotherapy-induced nausea and vomiting in patients receiving highly emetogenic chemotherapy".ti. (1)

74	"Efficacy and tolerability of aprepitant for the prevention of chemotherapy-induced nausea and vomiting in patients with breast cancer after moderately emetogenic chemotherapy".ti. (1)

75	"Single-dose fosaprepitant for the prevention of chemotherapy-induced nausea and vomiting associated with moderately emetogenic chemotherapy: Results of a randomized, double-blind phase iii trial".ti. (1)

76	"A comparison of oral ondansetron syrup or intravenous ondansetron loading dose regimens given in combination with dexamethasone for the prevention of nausea and emesis in pediatric and adolescent patients receiving moderately/highly emetogenic chemotherapy".ti. (1)

77	"A randomized study of olanzapine-containing versus standard antiemetic regimens for the prevention of chemotherapy-induced nausea and vomiting in chinese breast cancer patients".ti. (1)

78	"A randomized study of aprepitant, ondansetron and dexamethasone for chemotherapy-induced nausea and vomiting in chinese breast cancer patients receiving moderately emetogenic chemotherapy".ti. (1)

79	"The efficacy and safety of palonosetron compared with granisetron in preventing highly emetogenic chemotherapy-induced vomiting in the chinese cancer patients: A phase ii, multicenter, randomized, double-blind, parallel, comparative clinical trial".ti. (1)

80	or/1-79 (72)

81	80 (72)

82	limit 81 to english language (72)

83	81 not 82 (0)

84	limit 81 to (english language or no language specified) (72)

85	81 not 84 (0)

86	limit 81 to (english language or no language specified or multilingual or undetermined) (72)

87	81 not 86 (0)

88	((afrikaans or albanian or arabic or armenian or azerbaijani or belorussian or bengali or bosnian or bulgarian or burmese or catalan or chinese or croatian or czech or danish or dutch or esperanto or estonian or finnish or flemish or french or gaelic, scots or georgian or german or greek or hausa or hebrew or hindi or hungarian or icelandic or indonesian or italian or japanese or kinyarwanda or kirghiz or korean or latin or latvian or lithuanian or macedonian or malay or malayalam or marathi or maori or masai or norwegian or persian or polish or portuguese or pushto or romanian or rumanian or russian or serbian or slovak or slovene or spanish or swahili or swedish or tagalog or tamil or telugu or thai or turkish or ukrainian or urdu or vietnamese or welsh) not english).lg. (4602638)

89	81 not 88 (72)

90	81 and 88 (0)

91	((afrikaans or albanian or arabic or armenian or azerbaijani or belorussian or bengali or bosnian or bulgarian or burmese or catalan or chinese or croatian or czech or danish or dutch or esperanto or estonian or finnish or flemish or french or gaelic, scots or georgian or german or greek or hausa or hebrew or hindi or hungarian or icelandic or indonesian or italian or japanese or kinyarwanda or kirghiz or korean or latin or latvian or lithuanian or macedonian or malay or malayalam or marathi or maori or masai or norwegian or persian or polish or portuguese or pushto or romanian or rumanian or russian or serbian or slovak or slovene or spanish or swahili or swedish or tagalog or tamil or telugu or thai or turkish or ukrainian or urdu or vietnamese or welsh) not (english or undetermined or multilingual)).lg. (4602596)

92	81 not 91 (72)

93	81 and 91 (0)

**Includes: Embase** via Ovid http://ovidsp.ovid.com/

Date range searched: <1974 to 2024 January 24>

Date searched: 25 January 2024"

1	"Addition of 3-day aprepitant to ondansetron and dexamethasone for prophylaxis of chemotherapy-induced nausea and vomiting among patients with diffuse large b cell lymphoma receiving 5-day cisplatin-based chemotherapy".ti. (1)

2	"Randomized double-blind crossover ondansetron-dexamethasone versus ondansetron-placebo study for the treatment of chemotherapy-induced nausea and vomiting in pediatric patients with malignancies".ti. (1)

3	"The antiemetic efficacy and safety of granisetron compared with metoclopramide plus dexamethasone in patients receiving fractionated chemotherapy over 5 days. The granisetron study group".ti. (0)

4	"Aprepitant as an add-on therapy in children receiving highly emetogenic chemotherapy: A randomized, double-blind, placebo-controlled trial".ti. not conference abstract.pt. (1)

5	"A double-blind, crossover, randomized dose-comparison trial of granisetron for the prevention of acute and delayed nausea and emesis in children receiving moderately emetogenic carboplatin-based chemotherapy".ti. (1)

6	"An increased loading dose of ondansetron: A north european, double-blind randomised study in children, comparing 5 mg/m2 with 10 mg/m2".ti. (0)

7	"Cardiac effects of granisetron in a prospective crossover randomized dose comparison trial".ti. (1)

8	"Palonosetron is a better choice compared with ondansetron for the prevention of chemotherapy-induced nausea and vomiting (cinv) in a resource-limited pediatric oncology center: Results from a randomized control trial".ti. (1)

9	"The control of acute cisplatin-induced emesis--a comparative study of granisetron and a combination regimen of high-dose metoclopramide and dexamethasone. Granisetron study group".ti. (0)

10	"A randomized trial of individualized versus standard of care antiemetic therapy for breast cancer patients at high risk for chemotherapy-induced nausea and vomiting".ti. (1)

11	"Randomised comparison of ondansetron and metoclopramide plus dexamethasone for chemotherapy induced emesis".ti. (1)

12	"Palonosetron for prevention of acute and delayed nausea and vomiting in non-small-cell lung carcinoma patients".ti. (1)

13	"A randomised, double-blind, parallel-group study to compare the efficacy and safety of ondansetron (gr38032f) plus dexamethasone with metoclopramide plus dexamethasone in the prophylaxis of nausea and emesis induced by carboplatin chemotherapy".ti. (1)

14	"Evaluating aprepitant single-dose plus granisetron and dexamethasone in children receiving highly emetogenic chemotherapy for the prevention of chemotherapy-induced nausea and vomiting: A triple-blinded randomized clinical trial".ti. (1)

15	"Granisetron, tropisetron, and ondansetron in the prevention of acute emesis induced by a combination of cisplatin-adriamycin and by high-dose ifosfamide delivered in multiple-day continuous infusions".ti. (1)

16	"Double-blind comparative trial of oral ondansetron versus oral granisetron versus iv ondansetron in the prevention of nausea and vomiting associated with highly emetogenic preparative regimens prior to stem cell transplantation".ti. (1)

17	"Ondansetron versus granisetron in the prevention of chemotherapy-induced nausea and vomiting. Results of a prospective randomized trial".ti. (1)

18	"Comparing different antiemetic regimens for chemotherapy induced nausea and vomiting".ti. (1)

19	"Aprepitant in adolescent patients for prevention of chemotherapy-induced nausea and vomiting: A randomized, double-blind, placebo-controlled study of efficacy and tolerability".ti. (1)

20	"Palonosetron improves prevention of chemotherapy-induced nausea and vomiting following moderately emetogenic chemotherapy: Results of a double-blind randomized phase iii trial comparing single doses of palonosetron with ondansetron".ti. (1)

21	"Single-dose oral granisetron has equivalent antiemetic efficacy to intravenous ondansetron for highly emetogenic cisplatin-based chemotherapy".ti. (1)

22	"Single-dose fosaprepitant for the prevention of chemotherapy-induced nausea and vomiting associated with cisplatin therapy: Randomized, double-blind study protocol--ease".ti. (1)

23	"A single-blind comparison of intravenous ondansetron, a selective serotonin antagonist, with intravenous metoclopramide in the prevention of nausea and vomiting associated with high-dose cisplatin chemotherapy".ti. (1)

24	"Efficacy and tolerability of aprepitant for the prevention of chemotherapy-induced nausea and emesis over multiple cycles of moderately emetogenic chemotherapy".ti. not erratum/ (1)

25	"The oral neurokinin-1 antagonist aprepitant for the prevention of chemotherapy-induced nausea and vomiting: A multinational, randomized, double-blind, placebo-controlled trial in patients receiving high-dose cisplatin--the aprepitant protocol 052 study group".ti. (2)

26	"The effect of palonosetron hydrochloride in the prevention of chemotherapy-induced moderate and severe nausea and vomiting".ti. (1)

27	"Randomized, double-blind, placebo-controlled study of aprepitant versus two dosages of olanzapine with ondansetron plus dexamethasone for prevention of chemotherapy-induced nausea and vomiting in patients receiving high-emetogenic chemotherapy".ti. (1)

28	"A randomized, open-label non-inferiority study to compare palonosetron and ondansetron for prevention of acute chemotherapy-induced vomiting in children with cancer receiving moderate or high emetogenic chemotherapy".ti. (1)

29	"Single-dose oral granisetron versus multidose intravenous ondansetron for moderately emetogenic cyclophosphamide-based chemotherapy in pediatric outpatients with acute lymphoblastic lukemia".ti. (1)

30	"A comparison of ondansetron with metoclopramide in the prophylaxis of chemotherapy-induced nausea and vomiting: A randomized, double-blind study. International emesis study group".ti. (0)

31	"Aprepitant for the prevention of chemotherapy-induced nausea and vomiting in paediatric subjects: An analysis by age group".ti. (1)

32	"Aprepitant for the prevention of chemotherapy-induced nausea and vomiting in children: A randomised, double-blind, phase 3 trial".ti. not erratum/ (1)

33	"Clinical evaluation of two antiemetic combinations palonosetron dexamethasone versus ondansetron dexamethasone in chemotherapy of head and neck cancer".ti. (1)

34	"Efficacy and safety of aprepitant for the prevention of chemotherapy-induced nausea and vomiting during the first cycle of moderately emetogenic chemotherapy in korean patients with a broad range of tumor types".ti. not erratum/ (1)

35	"A randomised dose-comparison trial of granisetron in preventing emesis in children with leukaemia receiving emetogenic chemotherapy".ti. (1)

36	"Effectiveness of oral ondansetron in the management of nausea and vomiting induced by moderately emetogenic chemotherapy".ti. (1)

37	"Comparison of the efficacy and side-effects of ondansetron and metoclopramide-diphenhydramine administered to control nausea and vomiting in children treated with antineoplastic chemotherapy: A prospective randomized study".ti. (1)

38	"Palonosetron compared with ondansetron in pediatric cancer patients: Multicycle analysis of a randomized phase iii study".ti. (1)

39	"Palonosetron versus ondansetron for prevention of chemotherapy-induced nausea and vomiting in paediatric patients with cancer receiving moderately or highly emetogenic chemotherapy: A randomised, phase 3, double-blind, double-dummy, non-inferiority study".ti. (1)

40	"Ondansetron compared with dexamethasone and metoclopramide as antiemetics in the chemotherapy of breast cancer with cyclophosphamide, methotrexate, and fluorouracil".ti. (1)

41	"Does a single dose of palonosetron have any role in preventing acute chemotherapy-induced nausea and vomiting in pediatric osteosarcoma patients without dexamethasone? A randomized clinical trial".ti. (1)

42	"Contribution to the treatment of nausea and emesis induced by chemotherapy in children and adolescents with osteosarcoma".ti. (1)

43	"A prospective triple-blind randomized trial on safety and efficacy of abitant in the prevention of chemotherapy-induced nausea and vomiting".ti. (1)

44	"An open randomised cross-over study on granisetron versus ondansetron in the prevention of acute emesis induced by moderate dose cisplatin-containing regimens".ti. (1)

45	"Daily palonosetron is superior to ondansetron in the prevention of delayed chemotherapy-induced nausea and vomiting in patients with acute myelogenous leukemia".ti. (1)

46	"Comparison of acute and delayed antiemetic effect of adding aprepitant to odansitron and dexamethasone regimen in patients with hodgkin lymphoma receiving highly emetogenic chemotherapy".ti. (1)

47	"Efficacy and safety of olanzapine in children receiving highly emetogenic chemotherapy: A randomized, double-blind placebo-controlled phase 3 trial".ti. (1)

48	"Triplet aprepitant/dexamethasone/ondansetron versus doublet dexamethasone/ondansetron for prevention of moderately emetogenic chemotherapy: A placebo-controlled, double-blind, randomized clinical trial of efficacy".ti. (1)

49	"Efficacy and safety of adding olanzapine to the standard preventive regimen for chemotherapy-induced nausea and vomiting in children: A randomized double-blind controlled trial".ti. (1)

50	"Olanzapine for prevention of vomiting in children and adolescents receiving highly emetogenic chemotherapy: Investigator-initiated, randomized, open-label trial".ti. (0)

51	"Comparative clinical trial of granisetron and ondansetron in the prophylaxis of cisplatin-induced emesis. The granisetron study group".ti. (0)

52	"A double-blind, randomised, crossover comparison of granisetron and ondansetron in 5-day fractionated chemotherapy: Assessment of efficacy, safety and patient preference. The granisetron study group".ti. (0)

53	"A comparative study of intravenous granisetron versus intravenous and oral ondansetron in the prevention of nausea and vomiting associated with moderately emetogenic chemotherapy".ti. (1)

54	"Randomized open-label phase ii trial of 5-day aprepitant plus ondansetron compared to ondansetron alone in the prevention of chemotherapy-induced nausea-vomiting (cinv) in glioma patients receiving adjuvant temozolomide".ti. not letter/ (1)

55	"Comparison of antiemetic efficacy and safety of palonosetron vs ondansetron in the prevention of chemotherapy-induced nausea and vomiting in children".ti. (1)

56	"Addition of the neurokinin 1 receptor antagonist aprepitant to standard antiemetic therapy improves control of chemotherapy-induced nausea and vomiting. Results from a randomized, double-blind, placebo-controlled trial in latin america".ti. (1)

57	"Intravenous fosaprepitant for the prevention of chemotherapy induced vomiting in children, a randomized, phase 3 trial".ti. (1)

58	"Aprepitant for the prevention of chemotherapy-induced nausea and vomiting associated with a broad range of moderately emetogenic chemotherapies and tumor types: A randomized, double-blind study".ti. not conference abstract.pt. (1)

59	"Ondansetron compared with granisetron in the prophylaxis of cisplatin-induced acute emesis: A multicentre double-blind, randomised, parallel-group study".ti. (1)

60	"Single daily dosing versus divided dosing intravenous ondansetron to prevent chemotherapy-induced nausea and vomiting among children: A comparative randomized double-blind controlled trial".ti. (1)

61	"Randomized double-blind comparison of single high-dose ondansetron and multiple standard-dose ondansetron in chemotherapy-naive pediatric oncology patients".ti. (1)

62	"Comparison of an aprepitant regimen with a multiple-day ondansetron regimen, both with dexamethasone, for antiemetic efficacy in high-dose cisplatin treatment".ti. (1)

63	"Palonosetron hydrochloride is an effective and safe option to prevent chemotherapy-induced nausea and vomiting in children".ti. (1)

64	"Addition of aprepitant improves acute emesis control in children and adolescents receiving induction chemotherapy for acute myeloid leukaemia: A randomised, open-label trial".ti. (2)

65	"Ondansetron versus granisetron in the prevention of chemotherapy induced nausea and vomiting in children with acute lymphoblastic leukemia".ti. (1)

66	"Ondansetron compared with metoclopramide in the control of emesis and quality of life during repeated chemotherapy for breast cancer".ti. (1)

67	"A comparison of oral ondansetron and intravenous granisetron for the prevention of nausea and emesis associated with cisplatin-based chemotherapy".ti. (1)

68	"Ondansetron and tropisetron in the control of nausea and vomiting in children receiving combined cancer chemotherapy".ti. (1)

69	"Prevention of nausea and vomiting associated with stem cell transplant: Results of a prospective, randomized trial of aprepitant used with highly emetogenic preparative regimens".ti. (1)

70	"Palonosetron is nonsuperior to ondansetron in acute phase but provides superior antiemetic control in delayed phase for pediatric patients administered highly emetogenic chemotherapy".ti. (1)

71	"A randomized, double-blind, placebo-controlled study evaluating the efficacy of combination olanzapine, ondansetron and dexamethasone for prevention of chemotherapy-induced nausea and vomiting in patients receiving doxorubicin plus cyclophosphamide".ti. not conference abstract.pt. not medline.af. (1)

72	"Effects of granisetron in children undergoing high-dose chemotherapy: A multi-institutional, cross-over study".ti. (1)

73	"The efficacy and safety of the addition of olanzapine to ondansetron and dexamethasone for prevention of chemotherapy-induced nausea and vomiting in patients receiving highly emetogenic chemotherapy".ti. (1)

74	"Efficacy and tolerability of aprepitant for the prevention of chemotherapy-induced nausea and vomiting in patients with breast cancer after moderately emetogenic chemotherapy".ti. not erratum/ not medline.af. (1)

75	"Single-dose fosaprepitant for the prevention of chemotherapy-induced nausea and vomiting associated with moderately emetogenic chemotherapy: Results of a randomized, double-blind phase iii trial".ti. (1)

76	"A comparison of oral ondansetron syrup or intravenous ondansetron loading dose regimens given in combination with dexamethasone for the prevention of nausea and emesis in pediatric and adolescent patients receiving moderately/highly emetogenic chemotherapy".ti. (1)

77	"A randomized study of olanzapine-containing versus standard antiemetic regimens for the prevention of chemotherapy-induced nausea and vomiting in chinese breast cancer patients".ti. (1)

78	"A randomized study of aprepitant, ondansetron and dexamethasone for chemotherapy-induced nausea and vomiting in chinese breast cancer patients receiving moderately emetogenic chemotherapy".ti. (1)

79	"The efficacy and safety of palonosetron compared with granisetron in preventing highly emetogenic chemotherapy-induced vomiting in the chinese cancer patients: A phase ii, multicenter, randomized, double-blind, parallel, comparative clinical trial".ti. (1)

80	or/1-79 (74)

81	remove duplicates from 80 (72)

82	81 (72)

83	limit 82 to english language (72)

84	82 not 83 (0)

85	limit 82 to (english language or no language specified) (72)

86	82 not 85 (0)

87	limit 82 to (english language or no language specified or polyglot) (72)

88	82 not 87 (0)

89	((afrikaans or albanian or arabic or armenian or azerbaidzhani or azerbaijani or basque or belorussian or bengali or bulgarian or burmese or bosnian or catalan or catalan; valencian or chinese or croatian or czech or danish or dutch or dutch; flemish or esperanto or estonian or finnish or french or gallegan or georgian or german or greek or "greek, modern (1453-)" or hebrew or hindi or hungarian or icelandic or indonesian or italian or japanese or korean or latvian or lithuanian or macedonian or malay or maori or mongolian or norwegian or persian or polish or portuguese or pushto or romanian or russian or serbian or sinhalese or slovak or slovene or slovenian or spanish or spanish; castilian or swedish or tagalog or thai or turkish or ukrainian or urdu or uzbek or vietnamese) not english).lg. (4690154)

90	82 not 89 (72)

91	82 and 89 (0)

92	((afrikaans or albanian or arabic or armenian or azerbaidzhani or azerbaijani or basque or belorussian or bengali or bulgarian or burmese or bosnian or catalan or catalan; valencian or chinese or croatian or czech or danish or dutch or dutch; flemish or esperanto or estonian or finnish or french or gallegan or georgian or german or greek or "greek, modern (1453-)" or hebrew or hindi or hungarian or icelandic or indonesian or italian or japanese or korean or latvian or lithuanian or macedonian or malay or maori or mongolian or norwegian or persian or polish or portuguese or pushto or romanian or russian or serbian or sinhalese or slovak or slovene or slovenian or spanish or spanish; castilian or swedish or tagalog or thai or turkish or ukrainian or urdu or uzbek or vietnamese) not (english or polyglot)).lg. (4690154)

93	82 not 92 (72)

94	82 and 92 (0)

**Excludes: MEDLINE® ALL** via Ovid http://ovidsp.ovid.com/

Date range searched: <1946 to January 24, 2024>

Date searched: 25 January 2024

1	"Role of metoclopramide in the prevention of chemotherapy-induced nausea and vomiting".ti. (0)

2	"Treatment of chemotherapy-induced nausea and vomiting with 5-hydroxytryptamine type 3 receptor antagonists".ti. (1)

3	"Olanzapine: Extension of preventive options for nausea and vomiting after highly emetogenic chemotherapy".ti. (0)

4	"Comparison of intravenous granisetron with metoclopramide plus dexamethasone in the prevention of nausea and vomiting associated with emetogenic cytotoxic chemotherapy".ti. (1)

5	"A comparison of the acute antiemetic effect of tropisetron with ondansetron in patients receiving cisplatin".ti. (0)

6	"A prospective randomized comparison of ondansetron and metoclopramide in the prophylaxis of emesis induced by cisplatinum based combination chemotherapy".ti. (0)

7	"Prevention of cytostatic-induced vomiting with high-dose metoclopramide".ti. (1)

8	"Trial of premedication with lorazepam for the prevention of emetic effects of cisplatin".ti. (1)

9	"Tropisetron plus dexamethasone to control nausea and vomiting in patients with emetogenic chemotherapy".ti. (0)

10	"Antiemetic efficacy of granisetron in the treatment of pediatric cancer--(1). Clinical evaluation of granisetron at a dose of 40 micrograms/kg".ti. (1)

11	"Examination on efficacy and safety of concurrent use of ondansetron hydrochloride and steroid in lung cancer patients on cisplatin".ti. (1)

12	"Effect of ondansetron in prevention of nausea and vomiting induced by cancer chemotherapy".ti. (0)

13	"Granisetron (per os) compared with ondansetron (per os) in the prevention of nausea and vomiting induced by mildly emetogenic chemotherapies. Groupe de recherches en cancerologie du nord".ti. (1)

14	"Antiemetic efficacy of granisetron compared with high-dose metoclopramide plus dexamethasone in patients with primary lung cancer receiving chemotherapy: A randomized crossover trial".ti. (0)

15	"Comparison of antiemetic efficacy of granisetron as premedication versus combination therapy of granisetron".ti. (1)

16	"Evaluatin of sn-307 (ondansetron), given intravenously for the treatment of nausea and vomiting caused by anticancer drugs including cisplatin - a placebo-controlled, double-blind comparative study".ti. (0)

17	"A randomized comparison of antiemetic effect of ondansetron versus mdl (metoclopramide/dexamethasone/lorazepam) in patients receiving cisplatin-based combination chemotherapy".ti. (0)

18	"Anti-emetic treatment with metoclopramide and other drugs during cddp therapy".ti. (1)

19	"Clinical phase iii study of tropisetron capsule in the treatment of nausea and vomiting induced by anti-cancer drug; a placebo-controlled, multicenter, double-blind comparative study".ti. (1)

20	"Ondansetron for prophylaxis of nausea and vomiting after bone marrow transplantation".ti. (0)

21	"Comparison of tropisetron with ondansetron in the prevention of cisplatin-induced nausea and vomiting".ti. (0)

22	"Clinical study of ondansetron versus tropisetron in the prevention of nausea and vomiting caused by fractionated radiotherapy to the upper abdomen".ti. (0)

23	"Antiemetic effect of low dose olanzapine in solid tumor chemotherapy".ti. (0)

24	"Oral granisetron solution as prophylaxis for chemotherapy-induced emesis in children: Double-blind study of 2 doses".ti. (1)

25	"Comparative trial of oral granisetron and intravenous ondansetron in patients receiving chemotherapy for breast cancer".ti. (1)

26	"Curative efficacy of ondansetron against nausea and emesis induced by anticancer drugs: A study versus metoclopramide".ti. (0)

27	"Effects of an anti-emetic tropisetron capsule on qol of patients with delayed nausea and vomiting induced by cancer chemotherapy. Group for investigation of qol questionnaire for anti-emetics used in cancer chemotherapy. Joint research group for tropisetron double-blind comparative study".ti. (1)

28	"Comparative clinical evaluation of lorazepam (author's transl)".ti. (1)

29	"A prospective randomized comparative clinical trial comparing the efficacy between ondansetron and metoclopramide for prevention of nausea and vomiting in patients undergoing fractionated radiotherapy to the abdominal region".ti. (0)

30	"Efficacy and safety of palonosetron versus tropisetron in the prevention of highly emetogenic chemotherapy-induced acute and delayed vomiting in chinese cancer patients".ti. (1)

31	"Evaluation of the anti-emetic effectiveness of two drug formulations of ondansetron in combined chemotherapy for children with malignant tumors".ti. (1)

32	"Randomized control study of high-dose metoclopramide in the prevention of cddp-induced emesis".ti. (1)

33	"Efficacy of granisetron in the prevention of emesis induced by conditioning chemotherapy for allogeneic bone marrow transplantation".ti. (1)

34	"Efficacy of palonosetron hydrochloride injection in preventing gastrointestinal reactions caused by high-dose chemotherapy in osteosarcoma patients".ti. (0)

35	"Effect of concurrent use of ondansetron hydrochloride and dexamethasone against nausea and vomiting in lung cancer patients receiving cisplatin".ti. (1)

36	"Effect of palonosetron in preventing chemotherapy-induced vomiting".ti. (0)

37	"Randomized controlled study of high-dose metoclopramide and dexamethasone in the prevention of cddp-induced emesis".ti. (1)

38	"Efficacy and safety of multiple-dose 5-ht3 receptor antagonists in preventing multi-day-based and highly emetogenic chemotherapy-induced nausea and vomiting".ti. (0)

39	"The clinical effect of tropisetron in the prevention of nausea and vomiting induced by anti-cancer drugs".ti. (1)

40	"A comparison of ondansetron plus dexamethasone with metoclopramide plus dexamethasone for the prevention of emesis induced by non-cisplatin containing chemotherapy regimens".ti. (0)

41	"The effect of ondansetron in prevention of vomiting induced by combination chemotherapy with cisplation".ti. (0)

42	"Preventive efficacies of china-made tropisetron hydrochloride and navoban on chemotherapy-induced nausea and vomiting: A randomized controlled clinical trial".ti. (1)

43	"Comparing the preventive effect of granisetron versus ondansetron (made in china) on chemotherapy-induced nausea and vomiting: Report on a prospective multiple center randomized controlled trial".ti. (0)

44	"The role of ondansetron (qilu) in the prevention of non- cisplatin-induced vomiting--a randomised clinical trial".ti. (0)

45	"Anti-emetic effect of ondansetron in cisplatin induced nausea and vomiting--a randomized clinical trial".ti. (1)

46	"A randomized trial of tropisetron in the prophylaxis of nausea and vomiting induced by chemotherapy".ti. (1)

47	"Clinical study on granisetron in the prophylaxis of nausea and vomiting iduced by chemotherapeutic agents of gastric cancer".ti. (0)

48	or/1-47 (24)

49	48 (24)

50	limit 49 to english language (1)

51	49 not 50 (23)

52	limit 49 to (english language or no language specified) (1)

53	49 not 52 (23)

54	limit 49 to (english language or no language specified or multilingual or undetermined) (1)

55	49 not 54 (23)

56	((afrikaans or albanian or arabic or armenian or azerbaijani or belorussian or bengali or bosnian or bulgarian or burmese or catalan or chinese or croatian or czech or danish or dutch or esperanto or estonian or finnish or flemish or french or gaelic, scots or georgian or german or greek or hausa or hebrew or hindi or hungarian or icelandic or indonesian or italian or japanese or kinyarwanda or kirghiz or korean or latin or latvian or lithuanian or macedonian or malay or malayalam or marathi or maori or masai or norwegian or persian or polish or portuguese or pushto or romanian or rumanian or russian or serbian or slovak or slovene or spanish or swahili or swedish or tagalog or tamil or telugu or thai or turkish or ukrainian or urdu or vietnamese or welsh) not english).lg. (4602638)

57	49 not 56 (1)

58	49 and 56 (23)

59	((afrikaans or albanian or arabic or armenian or azerbaijani or belorussian or bengali or bosnian or bulgarian or burmese or catalan or chinese or croatian or czech or danish or dutch or esperanto or estonian or finnish or flemish or french or gaelic, scots or georgian or german or greek or hausa or hebrew or hindi or hungarian or icelandic or indonesian or italian or japanese or kinyarwanda or kirghiz or korean or latin or latvian or lithuanian or macedonian or malay or malayalam or marathi or maori or masai or norwegian or persian or polish or portuguese or pushto or romanian or rumanian or russian or serbian or slovak or slovene or spanish or swahili or swedish or tagalog or tamil or telugu or thai or turkish or ukrainian or urdu or vietnamese or welsh) not (english or undetermined or multilingual)).lg. (4602596)

60	49 not 59 (1)

61	49 and 59 (23)

**Excludes: Embase** via Ovid http://ovidsp.ovid.com/

Date range searched: <1974 to 2024 January 24>

Date searched: 25 January 2024

1	"Role of metoclopramide in the prevention of chemotherapy-induced nausea and vomiting".ti. (0)

2	"Treatment of chemotherapy-induced nausea and vomiting with 5-hydroxytryptamine type 3 receptor antagonists".ti. (1)

3	"Olanzapine: Extension of preventive options for nausea and vomiting after highly emetogenic chemotherapy".ti. (1)

4	"Comparison of intravenous granisetron with metoclopramide plus dexamethasone in the prevention of nausea and vomiting associated with emetogenic cytotoxic chemotherapy".ti. (1)

5	"A comparison of the acute antiemetic effect of tropisetron with ondansetron in patients receiving cisplatin".ti. (0)

6	"A prospective randomized comparison of ondansetron and metoclopramide in the prophylaxis of emesis induced by cisplatinum based combination chemotherapy".ti. (0)

7	"Prevention of cytostatic-induced vomiting with high-dose metoclopramide".ti. (0)

8	"Trial of premedication with lorazepam for the prevention of emetic effects of cisplatin".ti. (0)

9	"Tropisetron plus dexamethasone to control nausea and vomiting in patients with emetogenic chemotherapy".ti. (0)

10	"Antiemetic efficacy of granisetron in the treatment of pediatric cancer--(1). Clinical evaluation of granisetron at a dose of 40 micrograms/kg".ti. (0)

11	"Examination on efficacy and safety of concurrent use of ondansetron hydrochloride and steroid in lung cancer patients on cisplatin".ti. (1)

12	"Effect of ondansetron in prevention of nausea and vomiting induced by cancer chemotherapy".ti. (0)

13	"Granisetron (per os) compared with ondansetron (per os) in the prevention of nausea and vomiting induced by mildly emetogenic chemotherapies. Groupe de recherches en cancerologie du nord".ti. (0)

14	"Antiemetic efficacy of granisetron compared with high-dose metoclopramide plus dexamethasone in patients with primary lung cancer receiving chemotherapy: A randomized crossover trial".ti. (1)

15	"Comparison of antiemetic efficacy of granisetron as premedication versus combination therapy of granisetron".ti. (1)

16	"Evaluatin of sn-307 (ondansetron), given intravenously for the treatment of nausea and vomiting caused by anticancer drugs including cisplatin - a placebo-controlled, double-blind comparative study".ti. (1)

17	"A randomized comparison of antiemetic effect of ondansetron versus mdl (metoclopramide/dexamethasone/lorazepam) in patients receiving cisplatin-based combination chemotherapy".ti. (0)

18	"Anti-emetic treatment with metoclopramide and other drugs during cddp therapy".ti. (1)

19	"Clinical phase iii study of tropisetron capsule in the treatment of nausea and vomiting induced by anti-cancer drug; a placebo-controlled, multicenter, double-blind comparative study".ti. (1)

20	"Ondansetron for prophylaxis of nausea and vomiting after bone marrow transplantation".ti. (1)

21	"Comparison of tropisetron with ondansetron in the prevention of cisplatin-induced nausea and vomiting".ti. (0)

22	"Clinical study of ondansetron versus tropisetron in the prevention of nausea and vomiting caused by fractionated radiotherapy to the upper abdomen".ti. (1)

23	"Antiemetic effect of low dose olanzapine in solid tumor chemotherapy".ti. (1)

24	"Oral granisetron solution as prophylaxis for chemotherapy-induced emesis in children: Double-blind study of 2 doses".ti. (0)

25	"Comparative trial of oral granisetron and intravenous ondansetron in patients receiving chemotherapy for breast cancer".ti. (1)

26	"Curative efficacy of ondansetron against nausea and emesis induced by anticancer drugs: A study versus metoclopramide".ti. (1)

27	"Effects of an anti-emetic tropisetron capsule on qol of patients with delayed nausea and vomiting induced by cancer chemotherapy. Group for investigation of qol questionnaire for anti-emetics used in cancer chemotherapy. Joint research group for tropisetron double-blind comparative study".ti. (0)

28	"Comparative clinical evaluation of lorazepam (author's transl)".ti. (0)

29	"A prospective randomized comparative clinical trial comparing the efficacy between ondansetron and metoclopramide for prevention of nausea and vomiting in patients undergoing fractionated radiotherapy to the abdominal region".ti. (0)

30	"Efficacy and safety of palonosetron versus tropisetron in the prevention of highly emetogenic chemotherapy-induced acute and delayed vomiting in chinese cancer patients".ti. (1)

31	"Evaluation of the anti-emetic effectiveness of two drug formulations of ondansetron in combined chemotherapy for children with malignant tumors".ti. (1)

32	"Randomized control study of high-dose metoclopramide in the prevention of cddp-induced emesis".ti. (1)

33	"Efficacy of granisetron in the prevention of emesis induced by conditioning chemotherapy for allogeneic bone marrow transplantation".ti. (1)

34	"Efficacy of palonosetron hydrochloride injection in preventing gastrointestinal reactions caused by high-dose chemotherapy in osteosarcoma patients".ti. (1)

35	"Effect of concurrent use of ondansetron hydrochloride and dexamethasone against nausea and vomiting in lung cancer patients receiving cisplatin".ti. (1)

36	"Effect of palonosetron in preventing chemotherapy-induced vomiting".ti. (1)

37	"Randomized controlled study of high-dose metoclopramide and dexamethasone in the prevention of cddp-induced emesis".ti. (1)

38	"Efficacy and safety of multiple-dose 5-ht3 receptor antagonists in preventing multi-day-based and highly emetogenic chemotherapy-induced nausea and vomiting".ti. (1)

39	"The clinical effect of tropisetron in the prevention of nausea and vomiting induced by anti-cancer drugs".ti. (1)

40	"A comparison of ondansetron plus dexamethasone with metoclopramide plus dexamethasone for the prevention of emesis induced by non-cisplatin containing chemotherapy regimens".ti. (0)

41	"The effect of ondansetron in prevention of vomiting induced by combination chemotherapy with cisplation".ti. (0)

42	"Preventive efficacies of china-made tropisetron hydrochloride and navoban on chemotherapy-induced nausea and vomiting: A randomized controlled clinical trial".ti. (1)

43	"Comparing the preventive effect of granisetron versus ondansetron (made in china) on chemotherapy-induced nausea and vomiting: Report on a prospective multiple center randomized controlled trial".ti. (0)

44	"The role of ondansetron (qilu) in the prevention of non- cisplatin-induced vomiting--a randomised clinical trial".ti. (0)

45	"Anti-emetic effect of ondansetron in cisplatin induced nausea and vomiting--a randomized clinical trial".ti. (1)

46	"A randomized trial of tropisetron in the prophylaxis of nausea and vomiting induced by chemotherapy".ti. (1)

47	"Clinical study on granisetron in the prophylaxis of nausea and vomiting iduced by chemotherapeutic agents of gastric cancer".ti. (0)

48	or/1-47 (27)

49	48 (27)

50	limit 49 to english language (1)

51	49 not 50 (25)

52	limit 49 to (english language or no language specified) (2)

53	49 not 52 (25)

54	limit 49 to (english language or no language specified or polyglot) (2)

55	49 not 54 (25)

56	((afrikaans or albanian or arabic or armenian or azerbaidzhani or azerbaijani or basque or belorussian or bengali or bulgarian or burmese or bosnian or catalan or catalan; valencian or chinese or croatian or czech or danish or dutch or dutch; flemish or esperanto or estonian or finnish or french or gallegan or georgian or german or greek or "greek, modern (1453-)" or hebrew or hindi or hungarian or icelandic or indonesian or italian or japanese or korean or latvian or lithuanian or macedonian or malay or maori or mongolian or norwegian or persian or polish or portuguese or pushto or romanian or russian or serbian or sinhalese or slovak or slovene or slovenian or spanish or spanish; castilian or swedish or tagalog or thai or turkish or ukrainian or urdu or uzbek or vietnamese) not english).lg. (4690154)

57	49 not 56 (2)

58	49 and 56 (25)

59	((afrikaans or albanian or arabic or armenian or azerbaidzhani or azerbaijani or basque or belorussian or bengali or bulgarian or burmese or bosnian or catalan or catalan; valencian or chinese or croatian or czech or danish or dutch or dutch; flemish or esperanto or estonian or finnish or french or gallegan or georgian or german or greek or "greek, modern (1453-)" or hebrew or hindi or hungarian or icelandic or indonesian or italian or japanese or korean or latvian or lithuanian or macedonian or malay or maori or mongolian or norwegian or persian or polish or portuguese or pushto or romanian or russian or serbian or sinhalese or slovak or slovene or slovenian or spanish or spanish; castilian or swedish or tagalog or thai or turkish or ukrainian or urdu or uzbek or vietnamese) not (english or polyglot)).lg. (4690154)

60	49 not 59 (2)

61	49 and 59 (25)

### **Dataset 2:**

**Includes: MEDLINE® ALL** via Ovid http://ovidsp.ovid.com/

Date range searched: <1946 to January 24, 2024>

Date searched: 25 January 2024

1	"Energy drinks in the gulf cooperation council states: A review".ti. (1)

2	"Energy drinks and their adverse health effects: A systematic review of the current evidence".ti. (1)

3	"The negative impact of sugar-sweetened beverages on children's health: An update of the literature".ti. (1)

4	"Energy drink consumption among adolescents and young adults: Health effects and implications for practice".ti. (0)

5	"Extensive literature search as preparatory work for the safety assessment for caffeine".ti. (0)

6	"Behavioural correlates of energy drink consumption among adolescents: A review of the literature".ti. (1)

7	"Review of published cases of adverse cardiovascular events after ingestion of energy drinks".ti. (1)

8	"Energy drinks and myocardial ischemia: A review of case reports".ti. (1)

9	"Energy drinks and their adverse health effects: A systematic review and meta-analysis".ti. (1)

10	"A review of energy drinks and mental health, with a focus on stress, anxiety, and depression".ti. (1)

11	"Alcohol mixed with energy drinks and risk of injury: A systematic review".ti. (1)

12	("Alcohol mixed with energy drink (amed)" and "A critical review").ti. (1)

13	"Caffeine intake and its sources: A review of national representative studies".ti. (1)

14	"Consumption of energy drinks by children and young people: A rapid review examining evidence of physical effects and consumer attitudes".ti. (1)

15	"Association between energy drink consumption and substance use in adolescence: A systematic review of prospective cohort studies".ti. (1)

16	or/1-15 (13)

17	16 (13)

18	limit 17 to english language (13)

19	17 not 18 (0)

20	limit 17 to (english language or no language specified) (13)

21	17 not 20 (0)

22	limit 17 to (english language or no language specified or multilingual or undetermined) (13)

23	17 not 22 (0)

24	((afrikaans or albanian or arabic or armenian or azerbaijani or belorussian or bengali or bosnian or bulgarian or burmese or catalan or chinese or croatian or czech or danish or dutch or esperanto or estonian or finnish or flemish or french or gaelic, scots or georgian or german or greek or hausa or hebrew or hindi or hungarian or icelandic or indonesian or italian or japanese or kinyarwanda or kirghiz or korean or latin or latvian or lithuanian or macedonian or malay or malayalam or marathi or maori or masai or norwegian or persian or polish or portuguese or pushto or romanian or rumanian or russian or serbian or slovak or slovene or spanish or swahili or swedish or tagalog or tamil or telugu or thai or turkish or ukrainian or urdu or vietnamese or welsh) not english).lg. (4602638)

25	17 not 24 (13)

26	17 and 24 (0)

27	((afrikaans or albanian or arabic or armenian or azerbaijani or belorussian or bengali or bosnian or bulgarian or burmese or catalan or chinese or croatian or czech or danish or dutch or esperanto or estonian or finnish or flemish or french or gaelic, scots or georgian or german or greek or hausa or hebrew or hindi or hungarian or icelandic or indonesian or italian or japanese or kinyarwanda or kirghiz or korean or latin or latvian or lithuanian or macedonian or malay or malayalam or marathi or maori or masai or norwegian or persian or polish or portuguese or pushto or romanian or rumanian or russian or serbian or slovak or slovene or spanish or swahili or swedish or tagalog or tamil or telugu or thai or turkish or ukrainian or urdu or vietnamese or welsh) not (english or undetermined or multilingual)).lg. (4602596)

28	17 not 27 (13)

29	17 and 27 (0)

**Includes: Embase** via Ovid http://ovidsp.ovid.com/

Date range searched: <1974 to 2024 January 24>

Date searched: 25 January 2024

1	"Energy drinks in the gulf cooperation council states: A review".ti. (1)

2	"Energy drinks and their adverse health effects: A systematic review of the current evidence".ti. (1)

3	"The negative impact of sugar-sweetened beverages on children's health: An update of the literature".ti. (1)

4	"Energy drink consumption among adolescents and young adults: Health effects and implications for practice".ti. (0)

5	"Extensive literature search as preparatory work for the safety assessment for caffeine".ti. (0)

6	"Behavioural correlates of energy drink consumption among adolescents: A review of the literature".ti. (1)

7	"Review of published cases of adverse cardiovascular events after ingestion of energy drinks".ti. (1)

8	"Energy drinks and myocardial ischemia: A review of case reports".ti. (1)

9	"Energy drinks and their adverse health effects: A systematic review and meta-analysis".ti. (1)

10	"A review of energy drinks and mental health, with a focus on stress, anxiety, and depression".ti. (1)

11	"Alcohol mixed with energy drinks and risk of injury: A systematic review".ti. (1)

12	("Alcohol mixed with energy drink (amed)" and "A critical review").ti. (1)

13	"Caffeine intake and its sources: A review of national representative studies".ti. (1)

14	"Consumption of energy drinks by children and young people: A rapid review examining evidence of physical effects and consumer attitudes".ti. (1)

15	"Association between energy drink consumption and substance use in adolescence: A systematic review of prospective cohort studies".ti. (1)

16	or/1-15 (13)

17	16 (13)

18	limit 17 to english language (13)

19	17 not 18 (0)

20	limit 17 to (english language or no language specified) (13)

21	17 not 20 (0)

22	limit 17 to (english language or no language specified or polyglot) (13)

23	17 not 22 (0)

24	((afrikaans or albanian or arabic or armenian or azerbaidzhani or azerbaijani or basque or belorussian or bengali or bulgarian or burmese or bosnian or catalan or catalan; valencian or chinese or croatian or czech or danish or dutch or dutch; flemish or esperanto or estonian or finnish or french or gallegan or georgian or german or greek or "greek, modern (1453-)" or hebrew or hindi or hungarian or icelandic or indonesian or italian or japanese or korean or latvian or lithuanian or macedonian or malay or maori or mongolian or norwegian or persian or polish or portuguese or pushto or romanian or russian or serbian or sinhalese or slovak or slovene or slovenian or spanish or spanish; castilian or swedish or tagalog or thai or turkish or ukrainian or urdu or uzbek or vietnamese) not english).lg. (4690154)

25	17 not 24 (13)

26	17 and 24 (0)

27	((afrikaans or albanian or arabic or armenian or azerbaidzhani or azerbaijani or basque or belorussian or bengali or bulgarian or burmese or bosnian or catalan or catalan; valencian or chinese or croatian or czech or danish or dutch or dutch; flemish or esperanto or estonian or finnish or french or gallegan or georgian or german or greek or "greek, modern (1453-)" or hebrew or hindi or hungarian or icelandic or indonesian or italian or japanese or korean or latvian or lithuanian or macedonian or malay or maori or mongolian or norwegian or persian or polish or portuguese or pushto or romanian or russian or serbian or sinhalese or slovak or slovene or slovenian or spanish or spanish; castilian or swedish or tagalog or thai or turkish or ukrainian or urdu or uzbek or vietnamese) not (english or polyglot)).lg. (4690154)

28	17 not 27 (13)

29	17 and 27 (0)

**Excludes: MEDLINE® ALL** via Ovid http://ovidsp.ovid.com/

Date range searched: <1946 to January 24, 2024>

Date searched: 25 January 2024

1	"Toxicological evaluation of energy drinks on health effects".ti. (0)

2	"[caffeine in nutrition. Article 1. Consumption with food and regulation]".ti. (1)

3	"Revisión bibliográfica sobre el efecto de la cafeína en el rendimiento y en la salud de los deportistas ; bibliographic review effect of cafeine on the performance and health of athletes".ti. (0)

4	"[pharmaceutical cognitive doping in students: A chimeric way to get-a-head?]".ti. (2)

5	"Intoxicación por teobromina en pequeños animales, una revisión a un problema frecuente".ti. (0)

6	"Utilidad de las recomendaciones generales en la enfermedad por reflujo gastroesofágico: Una revisión sistemática de la literatura".ti. (0)

7	"Energidrycker - en hälsorisk? En litteraturbaserad studie med folkhälsovetenskapligt perspektiv ; energy drinks – a health risk? A literature-based study with public health perspective".ti. (0)

8	"Consumption of energy drinks with alcohol, hazardous mixture".ti. (1)

9	"Barn og unge må beskyttes mot energidrikker".ti. (0)

10	"Zwischenevaluation einer lebensstilintervention bei jugendlichen mit neet-status".ti. (0)

11	"Energy drinks in emergency medicine—give more than wings".ti. (0)

12	"Optimization and application of flowing atmospheric-pressure afterglow mass spectrometry for direct surface sampling and quantitative analysis ; optimierung und anwendung der flowing atmospheric-pressure afterglow quelle für die massenspektrometrie zur direkten oberflächenbeprobung und quantitativen analyse".ti. (0)

13	"Efecto del secado solar intermitente en la composición química del café ; intermittent solar drying effect on the chemical composition of coffee".ti. (0)

14	"Caffeinated energy drinks: Neurological and cardiovascular effects".ti. (0)

15	"Consumo de café e risco de diabetes tipo 2".ti. (0)

16	"[abuse of energy drinks: Does it pose a risk?]".ti. (1)

17	("Energy and sports drinks in children and adolescents" not "Re:").ti. (1)

18	"[health hazards of energy drinks--the situation in israel and the world]".ti. (1)

19	"Review of the composition of energy drinks and health effects perceived by young consumers ; revisión de la composición de las bebidas energizantes y efectos en la salud percibidos por jóvenes consumidores".ti. (0)

20	"Energy drinks are a real danger to the health of children, adolescents, young people, and the adult population. Part 2. Risks associated with consumption of alcohol-containing energy drinks. World health organization guidelines. Legislative regulation in different countries".ti. (0)

21	"[food, diet and dietetic in treatment of urinary tract dysfunctions. A review]".ti. (1)

22	"能量飲料對運動表現影響之研究 ; effects of energy drink on exercise performances".ti. (0)

23	or/1-22 (8)

24	remove duplicates from 23 (8)

25	24 (8)

26	limit 25 to english language (2)

27	25 not 26 (6)

28	limit 25 to (english language or no language specified) (2)

29	25 not 28 (6)

30	limit 25 to (english language or no language specified or multilingual or undetermined) (2)

31	25 not 30 (6)

32	((afrikaans or albanian or arabic or armenian or azerbaijani or belorussian or bengali or bosnian or bulgarian or burmese or catalan or chinese or croatian or czech or danish or dutch or esperanto or estonian or finnish or flemish or french or gaelic, scots or georgian or german or greek or hausa or hebrew or hindi or hungarian or icelandic or indonesian or italian or japanese or kinyarwanda or kirghiz or korean or latin or latvian or lithuanian or macedonian or malay or malayalam or marathi or maori or masai or norwegian or persian or polish or portuguese or pushto or romanian or rumanian or russian or serbian or slovak or slovene or spanish or swahili or swedish or tagalog or tamil or telugu or thai or turkish or ukrainian or urdu or vietnamese or welsh) not english).lg. (4602638)

33	25 not 32 (2)

34	25 and 32 (6)

35	((afrikaans or albanian or arabic or armenian or azerbaijani or belorussian or bengali or bosnian or bulgarian or burmese or catalan or chinese or croatian or czech or danish or dutch or esperanto or estonian or finnish or flemish or french or gaelic, scots or georgian or german or greek or hausa or hebrew or hindi or hungarian or icelandic or indonesian or italian or japanese or kinyarwanda or kirghiz or korean or latin or latvian or lithuanian or macedonian or malay or malayalam or marathi or maori or masai or norwegian or persian or polish or portuguese or pushto or romanian or rumanian or russian or serbian or slovak or slovene or spanish or swahili or swedish or tagalog or tamil or telugu or thai or turkish or ukrainian or urdu or vietnamese or welsh) not (english or undetermined or multilingual)).lg. (4602596)

36	25 not 35 (2)

37	25 and 35 (6)

**Excludes: Embase** via Ovid http://ovidsp.ovid.com/

Date range searched: <1974 to 2024 January 24>

Date searched: 25 January 2024

1	"Toxicological evaluation of energy drinks on health effects".ti. (0)

2	"[caffeine in nutrition. Article 1. Consumption with food and regulation]".ti. (1)

3	"Revisión bibliográfica sobre el efecto de la cafeína en el rendimiento y en la salud de los deportistas ; bibliographic review effect of cafeine on the performance and health of athletes".ti. (0)

4	"[pharmaceutical cognitive doping in students: A chimeric way to get-a-head?]".ti. and "Dopage cognitif chez les etudiants".ot. (1)

5	"Intoxicación por teobromina en pequeños animales, una revisión a un problema frecuente".ti. (0)

6	"Utilidad de las recomendaciones generales en la enfermedad por reflujo gastroesofágico: Una revisión sistemática de la literatura".ti. (0)

7	"Energidrycker - en hälsorisk? En litteraturbaserad studie med folkhälsovetenskapligt perspektiv ; energy drinks – a health risk? A literature-based study with public health perspective".ti. (0)

8	"Consumption of energy drinks with alcohol, hazardous mixture".ti. (1)

9	"Barn og unge må beskyttes mot energidrikker".ti. (0)

10	"Zwischenevaluation einer lebensstilintervention bei jugendlichen mit neet-status".ti. (0)

11	"Energy drinks in emergency medicine—give more than wings".ti. (0)

12	"Optimization and application of flowing atmospheric-pressure afterglow mass spectrometry for direct surface sampling and quantitative analysis ; optimierung und anwendung der flowing atmospheric-pressure afterglow quelle für die massenspektrometrie zur direkten oberflächenbeprobung und quantitativen analyse".ti. (0)

13	"Efecto del secado solar intermitente en la composición química del café ; intermittent solar drying effect on the chemical composition of coffee".ti. (0)

14	"Caffeinated energy drinks: Neurological and cardiovascular effects".ti. (1)

15	"Consumo de café e risco de diabetes tipo 2".ti. (0)

16	"[abuse of energy drinks: Does it pose a risk?]".ti. (1)

17	"Energy and sports drinks in children and adolescents".ti. and "Les boissons pour sportifs et les boissons".ot. (1)

18	"[health hazards of energy drinks--the situation in israel and the world]".ti. (1)

19	"Review of the composition of energy drinks and health effects perceived by young consumers ; revisión de la composición de las bebidas energizantes y efectos en la salud percibidos por jóvenes consumidores".ti. (0)

20	"Energy drinks are a real danger to the health of children, adolescents, young people, and the adult population. Part 2. Risks associated with consumption of alcohol-containing energy drinks. World health organization guidelines. Legislative regulation in different countries".ti. (0)

21	"[food, diet and dietetic in treatment of urinary tract dysfunctions. A review]".ti. (1)

22	"能量飲料對運動表現影響之研究 ; effects of energy drink on exercise performances".ti. (0)

23	or/1-22 (8)

24	23 (8)

25	limit 24 to english language (3)

26	24 not 25 (5)

27	limit 24 to (english language or no language specified) (3)

28	24 not 27 (5)

29	limit 24 to (english language or no language specified or polyglot) (3)

30	24 not 29 (5)

31	((afrikaans or albanian or arabic or armenian or azerbaidzhani or azerbaijani or basque or belorussian or bengali or bulgarian or burmese or bosnian or catalan or catalan; valencian or chinese or croatian or czech or danish or dutch or dutch; flemish or esperanto or estonian or finnish or french or gallegan or georgian or german or greek or "greek, modern (1453-)" or hebrew or hindi or hungarian or icelandic or indonesian or italian or japanese or korean or latvian or lithuanian or macedonian or malay or maori or mongolian or norwegian or persian or polish or portuguese or pushto or romanian or russian or serbian or sinhalese or slovak or slovene or slovenian or spanish or spanish; castilian or swedish or tagalog or thai or turkish or ukrainian or urdu or uzbek or vietnamese) not english).lg. (4690154)

32	24 not 31 (3)

33	24 and 31 (5)

34	((afrikaans or albanian or arabic or armenian or azerbaidzhani or azerbaijani or basque or belorussian or bengali or bulgarian or burmese or bosnian or catalan or catalan; valencian or chinese or croatian or czech or danish or dutch or dutch; flemish or esperanto or estonian or finnish or french or gallegan or georgian or german or greek or "greek, modern (1453-)" or hebrew or hindi or hungarian or icelandic or indonesian or italian or japanese or korean or latvian or lithuanian or macedonian or malay or maori or mongolian or norwegian or persian or polish or portuguese or pushto or romanian or russian or serbian or sinhalese or slovak or slovene or slovenian or spanish or spanish; castilian or swedish or tagalog or thai or turkish or ukrainian or urdu or uzbek or vietnamese) not (english or polyglot)).lg. (4690154)

35	24 not 34 (3)

36	24 and 34 (5)

### **Dataset 3:**

**Includes: MEDLINE® ALL** via Ovid http://ovidsp.ovid.com/

Date range searched: <1946 to January 30, 2024>

Date searched: 31st January 2024

1	"Exposure and preventive behaviours toward ticks and lyme disease in canada: Results from a first national survey".ti. and "2017".yr. (1)

2	"Lyme and tick-borne infections patient survey".ti. and "2012".yr. (0)

3	"An internet survey of lyme patients experience of access to care".ti. and "2013".yr. (0)

4	"A patient survey of a community: Lyme patients experiences of health care".ti. and "2013".yr. (0)

5	"Lyme disease on the nhs: Patient survey results 2016".ti. and "2016".yr. (0)

6	"Lyme disease, data and statistics".ti. and "2016".yr. (0)

7	"Patient narratives and an umwelten-based account of the more-than-human".ti. and "2016".yr. (0)

8	"Stress, despair, and quality of life: The lived experience of families with children below 26 years of age diagnosed with lyme disease".ti. and "2016".yr. (0)

9	"Zoonoses report united kingdom 2003".ti. and "2004".yr. (0)

10	"Zoonoses summary report: Uk 2014".ti. and "2015".yr. (0)

11	"National lyme disease surveillance in canada 2013: Web report".ti. and "2015".yr. (0)

12	"National lyme disease surveillance in canada 2009-2012".ti. and "2015".yr. (0)

13	"Surveillance of lyme disease in canada".ti. and "2016".yr. (0)

14	"Epidemiology and risks of tick-borne lyme borreliosis".ti. and "2009".yr. (0)

15	"Lyme neuroborreliosis in ireland 2012 and 2013".ti. and "2013".yr. (0)

16	"Foreign travel associated illness, england, wales and northern ireland: 2007 report".ti. and "2007".yr. (0)

17	"Pediatric lyme disease prevention".ti. and "2015".yr. (0)

18	"Tick-borne diseases: Assessing the knowledge, attitudes, and behaviors of college students".ti. and "2015".yr. (0)

19	"Fight lyme now survey".ti. and "2017".yr. (0)

20	"Common animal associated infections quarterly report (england and wales) - third quarter 2014".ti. and "2014".yr. (0)

21	"Lyme disease in ireland survey results 2016".ti. and "2016".yr. (0)

22	"Nhs testing and investigation of lyme borreliosis – patient experiences survey".ti. and "2014".yr. (0)

23	"German patient survey 2015 results".ti. and "2015".yr. (0)

24	"Waterborne zoonoses".ti. and "2004".yr. (0)

25	"How to design work related information for low-literate employees?".ti. and "2014".yr. (0)

26	"Multi-center validation of a clinical decision rule to distinguish lyme from aseptic meningitis in children in lyme disease endemic areas".ti. and "2011".yr. (0)

27	"Lyme disease as a cause of aseptic meningitis in children in a lyme-endemic region...2007 society for academic emergency medicine annual meeting".ti. and "2007".yr. (0)

28	"Clinical predictors of lyme disease among pediatric patients with acute monoarticular arthritis...2007 society for academic emergency medicine annual meeting".ti. and "2007".yr. (0)

29	"Acute polyradiculoneuritis syndrome: Clinical observations and differential diagnosis".ti. and "2012".yr. (1)

30	"Course of borrelia burgdorferi dna shedding in urine after treatment".ti. and "2007".yr. (1)

31	"The many faces of solitary and multiple erythema migrans".ti. and "2013".yr. (1)

32	"Clinical spectrum of skin manifestations of lyme borreliosis in 204 children in austria".ti. and "2015".yr. (1)

33	"Detection of spirochaetal dna simultaneously in skin biopsies, peripheral blood and urine from patients with erythema migrans".ti. and "2004".yr. (1)

34	"Granuloma annulare displaying pseudorosettes in borelia infection".ti. and "2008".yr. (1)

35	"Laboratory diagnostic methods in suspected disseminated lyme disease: A comparison of different techniques".ti. and "2011".yr. (0)

36	"Peripheral neuropathy in acrodermatitis chronica atrophicans - effect of treatment".ti. and "2002".yr. (1)

37	"Chronic symptoms are common in patients with neuroborreliosis -- a questionnaire follow-up study".ti. and "2002".yr. (1)

38	"European neuroborreliosis: Quality of life 30 months after treatment".ti. and "2011".yr. (1)

39	"Risk factors for a non-favorable outcome after treated european neuroborreliosis".ti. and "2013".yr. (1)

40	"Pcr in lyme neuroborreliosis: A prospective study".ti. and "2005".yr. (1)

41	"Multiple sclerosis in vest-agder county, norway".ti. and "2011".yr. (1)

42	"Cxcl13 chemokine in pediatric and adult neuroborreliosis".ti. and "2011".yr. (1)

43	"Use of electrocochleography for assessing endolymphatic hydrops in patients with lyme disease and meniere's disease".ti. and "2002".yr. (1)

44	"Brain magnetic resonance imaging does not contribute to the diagnosis of chronic neuroborreliosis".ti. and "2007".yr. (1)

45	"The analysis of the course of early localized lyme disease among patients of the infectious diseases clinic in szczecin".ti. and "2013".yr. (0)

46	"Borrelia burgdorferi sensu lato in brazil: Occurrence confirmed by immunohistochemistry and focus floating microscopy".ti. and "2010".yr. (1)

47	"Lyme neuroborreliosis".ti. and "2011".yr. and Acta Veterinaria.jn. (0)

48	"Serological markers of borrelia burgdorferi infection among forestry workers in west pomerania during a five-year period".ti. and "2004".yr. (0)

49	"Lyme borreliosis--the utility of improved real-time pcr assay in the detection of borrelia burgdorferi infections".ti. and "2015".yr. (1)

50	"Non-specific arthralgia as the main manifestation of borrelia burgdorferi sensu lato infection".ti. and "2008".yr. (0)

51	"Rational design of antigens to improve the serodiagnosis of tick-borne borreliosis in central regions of russia".ti. and "2014".yr. (1)

52	"Lyme disease in poland - a serious problem?".ti. and "2016".yr. (1)

53	"Recent lesson from a clinical and seroepidemiological survey: Low positive predictive value of borrelia burgdorferi antibody testing in a high risk population".ti. and "2012".yr. (1)

54	"Atovaquone plus cholestyramine in patients coinfected with babesia microti and borrelia burgdorferi refractory to other treatment".ti. and "2006".yr. (1)

55	"Mr imaging assessment of brain and cervical cord damage in patients with neuroborreliosis".ti. and "2006".yr. (1)

56	"Mri features of lyme arthritis in children".ti. and "2005".yr. (1)

57	"Immunological aspects of lyme borreliosis co-infection".ti. and "2011".yr. (0)

58	"Longitudinal evaluation of an educational intervention for preventing tick bites in an area with endemic lyme disease in baltimore county, maryland".ti. and "2003".yr. (1)

59	"Lyme disease incidence in wisconsin: A comparison of state-reported rates and rates from a population-based cohort".ti. and "2002".yr. (1)

60	"Effects of climate on variability in lyme disease incidence in the northeastern united states".ti. and "2003".yr. (1)

61	"Medical risks of wilderness hiking".ti. and "2003".yr. (1)

62	"Long-term follow-up of patients with culture-confirmed lyme disease".ti. and "2003".yr. (1)

63	"Systemic symptoms without erythema migrans as the presenting picture of early lyme disease".ti. and "2003".yr. (1)

64	"Lyme borreliosis and facial paralysis--a prospective analysis of risk factors and outcome".ti. and "2002".yr. (1)

65	"Higher prevalence of antibodies to borrelia burgdorferi in psychiatric patients than in healthy subjects".ti. and "2002".yr. (1)

66	"Borrelia-associated primary cutaneous malt lymphoma in a nonendemic region".ti. and "2003".yr. (1)

67	"Fourteen-year seroepidemiological study of zoonoses in a greek village".ti. and "2002".yr. (1)

68	"Lyme disease testing by multiplexed bead analysis using vlse1 and pepc10 antigens compared with 2-tiered testing".ti. and "2010".yr. (0)

69	"Focus floating microscopy: Gold standard for cutaneous borreliosis?".ti. and "2007".yr. (1)

70	"Cutaneous lymphoid hyperplasia and other lymphoid infiltrates of the breast nipple: A retrospective clinicopathologic study of fifty-six patients".ti. and "2005".yr. (1)

71	"Acrodermatitis chronica atrophicans with pseudolymphomatous infiltrates".ti. and "2013".yr. (1)

72	"Association between lyme disease and weather types in new york state".ti. and "2013".yr. (0)

73	"An evaluation of a lyme disease prevention program in a working population".ti. and "2006".yr. (1)

74	"Emergence of a persistent symptomatology in a cohort of patients reporting previous diagnosis and treatment for lyme disease".ti. and "2015".yr. (0)

75	"Subjective symptoms after treatment of early lyme disease".ti. and "2010".yr. (1)

76	"Etiology of pericarditis in a prospective cohort of 1162 cases".ti. and "2015".yr. (1)

77	"Psychiatric comorbidity and other psychological factors in patients with chronic lyme disease ".ti. and "2009".yr. (1)

78	"Long-term assessment of fatigue in patients with culture-confirmed lyme disease".ti. and "2015".yr. (1)

79	"No value for routine serologic screening for borrelia burgdorferi in patients with uveitis in the netherlands".ti. and "2016".yr. (1)

80	"Cardiac tropism of borrelia burgdorferi: An autopsy study of sudden cardiac death associated with lyme carditis".ti. and "2016".yr. (1)

81	"Peridomestic lyme disease prevention: Results of a population-based case-control study".ti. and "2009".yr. (1)

82	"Risk perceptions regarding ticks and lyme disease: A national survey".ti. and "2004".yr. (1)

83	"Disparities in reportable communicable disease incidence by census tract-level poverty, new york city, 2006-2013".ti. and "2015".yr. (1)

84	"Hyperplasia of hair follicles and other adnexal structures in cutaneous lymphoproliferative disorders: A study of 53 cases, including so-called pseudolymphomatous folliculitis and overt lymphomas".ti. and "2008".yr. (1)

85	"Spatial patterns of lyme disease risk in california based on disease incidence data and modeling of vector-tick exposure".ti. and "2006".yr. (1)

86	"Reinfection and relapse in early lyme disease".ti. and "2006".yr. (1)

87	"Lyme disease risk influences human settlement in the wildland-urban interface: Evidence from a longitudinal analysis of counties in the northeastern united states".ti. and "2014".yr. (1)

88	"Spatial and temporal emergence pattern of lyme disease in virginia".ti. and "2014".yr. (1)

89	"Meteorological influences on the seasonality of lyme disease in the united states".ti. and "2014".yr. (1)

90	"Survey for tick-borne zoonoses in the state of espirito santo, southeastern brazil".ti. and "2010".yr. (1)

91	"Human behavioral and ecological risk factors for lyme disease infection on block island, rhode island".ti. and "2013".yr. (0)

92	"Linking acarological risk and lyme disease incidence in the usa".ti. and "2011".yr. (0)

93	"Lyme disease and convergence insufficiency: Is it a near fit?".ti. and "2006".yr. (1)

94	"Epidemiological, clinical and paraclinical aspects of lyme disease in bihor county between 2007-2011".ti. and "2011".yr. (0)

95	"Particular aspects of lyme disease in bihor county".ti. and "2014".yr. (0)

96	"Borrelia burgdorferi infection among forestry workers - assessed with an immunoenzymatic method (elisa), pcr and correlated with the clinical state of the patients".ti. and "2003".yr. (1)

97	"Clinical characteristics and treatment outcome of early lyme disease in patients with microbiologically confirmed erythema migrans".ti. and "2002".yr. (1)

98	"Duration of antibiotic therapy for early lyme disease. A randomized, double-blind, placebo-controlled trial".ti. and "2003".yr. (1)

99	"Risk of tick-borne bacterial diseases in humans in the florence area, tuscany".ti. and "2003".yr. (1)

100	"Epidemiological and clinical differences among rickettsia slovaca rickettsiosis and other tick-borne diseases in spain".ti. and "2003".yr. (1)

101	"Bartonella henselae and borrelia burgdorferi infections of the central nervous system".ti. and "2003".yr. (1)

102	"Lyme borreliosis in south-eastern poland: Relationships with environmental factors and medical attention standards".ti. and "2011".yr. (1)

103	"Occurrence of cases of borreliosis certified as an occupational disease in the province of wielkopolska (poland)".ti. and "2009".yr. (1)

104	"Seroepidemiological study of lyme borreliosis among forestry workers in southern poland".ti. and "2009".yr. (1)

105	"Correlation of tests for detection of borrelia burgdorferi sensu lato infection in patients with diagnosed borreliosis".ti. and "2006".yr. (1)

106	"Serological survey in persons occupationally exposed to tick-borne pathogens in cases of co-infections with borrelia burgdorferi, anaplasma phagocytophilum, bartonella spp. And babesia microti".ti. and "2012".yr. (1)

107	"Risk of tick-borne bacterial diseases among workers of roztocze national park (south-eastern poland)".ti. and "2005".yr. (1)

108	"Study on lyme borreliosis focus in the lublin region (eastern poland)".ti. and "2008".yr. (1)

109	"Risk of tick-borne diseases in various categories of employment among forestry workers in eastern poland".ti. and "2012".yr. (1)

110	"Incidence and prevalence of infection with anaplasma phagocytophilum. Prospective study in healthy individuals exposed to ticks".ti. and "2004".yr. (1)

111	"Human anaplasmosis in north-eastern poland: Seroprevalence in humans and prevalence in ixodes ricinus ticks".ti. and "2004".yr. (1)

112	"Detecting dnas of anaplasma phagocytophilum and babesia in the blood of patients suspected of lyme disease".ti. and "2004".yr. (1)

113	"Occurrence of igg antibodies to anaplasma phagocytophilum in humans suspected of lyme borreliosis in eastern slovakia".ti. and "2009".yr. (1)

114	"Molecular and serological diagnosis of borrelia burgdorferi infection among patients with diagnosed erythema migrans".ti. and "2007".yr. (1)

115	"Epidemiology of lyme disease among workers of forest inspectorates in poland".ti. and "2013".yr. (1)

116	"Exposure to ticks and seroprevalence of borrelia burgdorferi among a healthy young population living in the area of southern podlasie, poland".ti. and "2014".yr. (1)

117	"Occupational exposure to zoonotic agents among agricultural workers in lombardy region, northern italy".ti. and "2013".yr. (1)

118	"Association between incidence of lyme disease and spring-early summer season temperature changes in hungary--1998-2010".ti. and "2013".yr. (1)

119	"Sero-epidemiological study of lyme disease among high-risk population groups in eastern slovakia".ti. and "2015".yr. (1)

120	"Risk compensation and vaccination: Can getting vaccinated cause people to engage in risky behaviors?".ti. and "2007".yr. (1)

121	"Risk perceptions and their relation to risk behavior".ti. and "2004".yr. (1)

122	"Prophylaxis with single-dose doxycycline for the prevention of lyme disease after an ixodes scapularis tick bite".ti. and "2002".yr. (0)

123	"Potential misdiagnoses of bell's palsy in the emergency department".ti. and "2014".yr. (1)

124	"Appropriateness of lyme disease serologic testing".ti. and "2004".yr. (1)

125	"Brief communication: Hematogenous dissemination in early lyme disease.[summary for patients in ann intern med. 2005 may 3;142(9):I48; pmid: 15867401]".ti. and "2005".yr. (0)

126	"Effect of iv antibiotic therapy on antibody profile in patients with post-treatment lyme encephalopathy".ti. and "2010".yr. (0)

127	"Anti-flagellin antibodies in patients with pandas-like illnesses".ti. and "2010".yr. (0)

128	"Prevalence of chlamydophila psittaci and borrelia burgdorferi infections in a series of 108 primary cutaneous lymphomas".ti. and "2011".yr. (0)

129	"Human exposure to anaplasma phagocytophilum in portugal".ti. and "2006".yr. (1)

130	"Joint involvement in russian patients with lyme borreliosis presenting with acrodermatitis chronica atrophicans".ti. and "2003".yr. (0)

131	"Is there an association between spondyloarthritis and antibodies towards borrelia, ehrlichia and chlamydia species?".ti. and "2015".yr. (0)

132	"Antibodies to borrelia burgdorferi in sera and synovial fluid in patients with lyme arthritis".ti. and "2006".yr. (0)

133	"Juvenile lyme arthritis".ti. and "2003".yr. (0)

134	"Serologic validation of four western blots for diagnosis of borrelia burgdorferi infection".ti. and "2009".yr. (0)

135	"Simultaneous use of serum igg and igm for risk scoring of suspected early lyme borreliosis: Graphical and bivariate analyses".ti. and "2010".yr. (1)

136	"Lyme borreliosis in sweden--diagnostic performance of five commercial borrelia serology kits using sera from well-defined patient groups".ti. and "2004".yr. (1)

137	"Seroprevalence of antibodies to borrelia burgdorferi sensu lato in healthy adults from western norway: Risk factors and methodological aspects".ti. and "2014".yr. (1)

138	"Improved serodiagnosis of early lyme borreliosis: Immunoblot with local borrelia afzelii strain".ti. and "2003".yr. (1)

139	"C6-peptide serology as diagnostic tool in neuroborreliosis".ti. and "2008".yr. (1)

140	"Memory and executive functions in adolescents with posttreatment lyme disease".ti. and "2008".yr. (1)

141	"Possible role of borrelia burgdorferi sensu lato infection in lichen sclerosus".ti. and "2008".yr. (1)

142	"How accurate is a clinical diagnosis of erythema chronicum migrans? Prospective study comparing the diagnostic accuracy of general practitioners and dermatologists in an area where lyme borreliosis is endemic".ti. and "2004".yr. (1)

143	"Lyme disease - united states, 2000".ti. and "2002".yr. and MMWR - Morbidity & Mortality Weekly Report.jn. (1)

144	"Clinical presentation of childhood neuroborreliosis; neurological examination may be normal".ti. and "2010".yr. (1)

145	"Seroprevalence of borrelia igg antibodies among young swedish children in relation to reported tick bites, symptoms and previous treatment for lyme borreliosis: A population-based survey".ti. and "2010".yr. (1)

146	"Lyme meningitis, the major cause of childhood meningitis in an endemic area: A population based study".ti. and "2012".yr. (1)

147	"Integrating spatial epidemiology into a decision model for evaluation of facial palsy in children".ti. and "2011".yr. (1)

148	"Epidemiological situation of lyme borreliosis in belgium, 2003 to 2012".ti. and "2015".yr. (1)

149	"Serodiagnosis of borreliosis: Indirect immunofluorescence assay, enzyme-linked immunosorbent assay and immunoblotting".ti. and "2011".yr. (1)

150	"Hyperosmia in lyme disease".ti. and "2014".yr. (1)

151	"Role of psychiatric comorbidity in chronic lyme disease".ti. and "2008".yr. (1)

152	"Burden and viability of borrelia burgdorferi in skin and joints of patients with erythema migrans or lyme arthritis".ti. and "2011".yr. (1)

153	"A82: Lyme arthritis: An emerging clinical problem in nova scotia, canada".ti. and "2014".yr. (0)

154	"Brief report: Long-term assessment of fibromyalgia in patients with culture-confirmed lyme disease".ti. and "2015".yr. (0)

155	"The utility of borrelia burgdorferi specific pcr of synovial fluid as a diagnostic tool for lyme arthritis".ti. and "2002".yr. (0)

156	"Prolonged synovitis in pediatric lyme disease: A study on clinical and laboratory predictors".ti. and "2002".yr. (0)

157	"A comparison of the presentation, synovial fluid analysis, and treatment course in children and adults with lyme arthritis".ti. and "2010".yr. (0)

158	"Prevalence and clinical presentation of lyme arthritis in a large cohort of patients with recent-onset arthritis".ti. and "2013".yr. (0)

159	"Sleep disturbances, fatigue and memory complaints attributed to chronic lyme disease".ti. and "2003".yr. (0)

160	"Maladaptive schemas in chronic lyme disease".ti. and "2004".yr. (0)

161	"Co-morbid mental disorders in chronic lyme disease: The first 50 patients".ti. and "2003".yr. (0)

162	"A prospective, longitudinal cohort study evaluating psychosocial risk and protective factors for post lyme disease syndrome".ti. and "2010".yr. (0)

163	"A lyme disease education program changes knowledge, attitudes and behaviors in elementary school children living in an endemic area".ti. and "2004".yr. (0)

164	("Diminished health-related quality-of-life improves over time in lyme disease" and "12 year followup from the nantucket lyme disease cohort study").ti. and "2002".yr. (0)

165	"Components of the health belief model are determinants of lyme disease preventive behavior among school-aged children in an endemic area of massachusetts".ti. and "2006".yr. (0)

166	"Interest of systematic lyme serology in context of recent onset arthritis".ti. and "2014".yr. (0)

167	"Lyme arthritis: A comparison of presentation, synovial fluid analysis, and treatment course in children and adults".ti. and "2013".yr. (1)

168	"Cost-effectiveness analysis of the lyme disease vaccine".ti. and "2002".yr. (1)

169	"Investigation of anaplasmosis in yiyuan county, shandong province, china".ti. and "2011".yr. (1)

170	"Seropositivity for anti-borrelia igg antibody is independently associated with carotid atherosclerosis".ti. and "2006".yr. (1)

171	"Gender differences in childhood lyme neuroborreliosis".ti. and "2015".yr. (1)

172	"Evaluation of a new euroimmun anti-borrelia select elisa".ti. and "2013".yr. (0)

173	"Lyme disease in the differential diagnosis of psychiatric patients".ti. and "2003".yr. (0)

174	"Seroepidemiological investigation of lyme disease and human granulocytic anaplasmosis among people living in forest areas of eight provinces in china".ti. and "2013".yr. (1)

175	"Evaluation of six recombinant proteins for serological diagnosis of lyme borreliosis in china".ti. and "2016".yr. (1)

176	"A study of the technique of western blot for diagnosis of lyme disease caused by borrelia afzelii in china".ti. and "2013".yr. (1)

177	"Absence of borrelia burgdorferi in the myocardium of subjects with normal left ventricular systolic function: A study using pcr and electron microscopy".ti. and "2016".yr. (1)

178	"Ethic of consensibility, subaltern ethicality: The clinical application of embryonic stem cells in india".ti. and "2013".yr. (0)

179	"Tick-borne infections and pediatric bipolar disorder".ti. and "2015".yr. (0)

180	"Personal and family history of immune-related conditions increase the risk of plasma cell disorders: A population-based study".ti. and "2011".yr. (1)

181	"Borrelia infection and risk of non-hodgkin lymphoma".ti. and "2008".yr. (1)

182	"P05.22. Health beliefs and experiences of patients with chronic lyme disease: A qualitative study...International research congress on integrative medicine and health 2012 portland, oregon, usa. 15-18 may 2012".ti. and "2012".yr. (0)

183	"Exploring the association between morgellons disease and lyme disease: Identification of borrelia burgdorferi in morgellons disease patients".ti. and "2015".yr. (1)

184	"Experiences of patients identifying with chronic lyme disease in the healthcare system: A qualitative study".ti. and "2014".yr. (1)

185	"Diagnostic challenges of early lyme disease: Lessons from a community case series".ti. and "2009".yr. (1)

186	"Probable late lyme disease: A variant manifestation of untreated borrelia burgdorferi infection".ti. and "2012".yr. (1)

187	"Clinical characteristics and cerebrospinal fluid parameters in patients with peripheral facial palsy caused by lyme neuroborreliosis compared with facial palsy of unknown origin (bell's palsy)".ti. and "2011".yr. (1)

188	"Utilization of serology for the diagnosis of suspected lyme borreliosis in denmark: Survey of patients seen in general practice".ti. and "2010".yr. (1)

189	"Lyme arthritis in southern norway--an endemic area for lyme borreliosis".ti. and "2014".yr. (1)

190	"Public health impact of strain specific immunity to borrelia burgdorferi".ti. and "2015".yr. (1)

191	"The diagnostic accuracy of serological tests for lyme borreliosis in europe: A systematic review and meta-analysis".ti. and "2016".yr. (1)

192	"Doxycycline-mediated effects on persistent symptoms and systemic cytokine responses post-neuroborreliosis: A randomized, prospective, cross-over study".ti. and "2012".yr. (1)

193	"Mapping of hormones and cortisol responses in patients after lyme neuroborreliosis".ti. and "2010".yr. (1)

194	"Efficacy and safety of pharmacological treatments for lyme neuroborreliosis in children: A systematic review".ti. and "2015".yr. (0)

195	"Neurocognitive functions and brain atrophy after proven neuroborreliosis: A case-control study".ti. and "2015".yr. (1)

196	"The nebop score - a clinical prediction test for evaluation of children with lyme neuroborreliosis in europe".ti. and "2015".yr. (1)

197	"Multi-criteria decision analysis as an innovative approach to managing zoonoses: Results from a study on lyme disease in canada".ti. and "2013".yr. (1)

198	"Factors associated with preventive behaviors regarding lyme disease in canada and switzerland: A comparative study".ti. and "2015".yr. (1)

199	"Acceptability of tick control interventions to prevent lyme disease in switzerland and canada: A mixed-method study".ti. and "2016".yr. (1)

200	"From lyme disease emergence to endemicity: A cross sectional comparative study of risk perceptions in different populations".ti. and "2014".yr. (1)

201	"Education on tick bite and lyme borreliosis prevention, aimed at schoolchildren in the netherlands: Comparing the effects of an online educational video game versus a leaflet or no intervention".ti. and "2016".yr. (1)

202	"Study on public perceptions and protective behaviors regarding lyme disease among the general public in the netherlands: Implications for prevention programs".ti. and "2013".yr. (1)

203	"Comparing the effect of a leaflet and a movie in preventing tick bites and lyme disease in the netherlands".ti. and "2016".yr. (1)

204	"Determinants and protective behaviours regarding tick bites among school children in the netherlands: A cross-sectional study".ti. and "2013".yr. (1)

205	"Subjective health complaints are not associated with tick bites or antibodies to borrelia burgdorferi sensu lato in blood donors in western norway: A cross-sectional study".ti. and "2015".yr. (1)

206	"Decrease in tick bite consultations and stabilization of early lyme borreliosis in the netherlands in 2014 after 15 years of continuous increase".ti. and "2016".yr. (1)

207	"Are the current notification criteria for lyme borreliosis in norway suitable? Results of an evaluation of lyme borreliosis surveillance in norway, 1995-2013".ti. and "2016".yr. (1)

208	"Experience of lyme disease and preferences for precautions: A cross-sectional survey of uk patients".ti. and "2013".yr. (1)

209	"Exploratory spatial analysis of lyme disease in texas -what can we learn from the reported cases?".ti. and "2015".yr. (1)

210	"Early lyme disease with spirochetemia - diagnosed by dna sequencing".ti. and "2010".yr. (1)

211	"Long-term prognosis of patients treated for erythema migrans in france".ti. and "2002".yr. (1)

212	"Borreliosis - a cause of obstructive sleep apnea syndrome".ti. and "2012".yr. (0)

213	"Contribution to laboratory diagnostics of neuroborreliosis".ti. and "2003".yr. (1)

214	"Reported incidence of lyme disease in slovakia and antibodies to b. Burgdorferi antigens detected in healthy population".ti. and "2005".yr. (1)

215	"Our experience with examination of antibodies against antigens of borrelia burgdorferi in patients with suspected lyme disease".ti. and "2010".yr. (1)

216	"Antibodies to borrelia burgdorferi in erythema migrans patients".ti. and "2007".yr. (1)

217	"Prevalence of antibodies to borrelia burgdorferi sensu stricto in humans from a cuban village".ti. and "2012".yr. (1)

218	"Zoonoses in humans from small rural properties in jataizinho, parana, brazil".ti. and "2013".yr. (1)

219	"Borrelia burgdorferi sensu lato in humans in a rural area of parana state, brazil".ti. and "2015".yr. (1)

220	"Seroprevalence of borrelia burgdorferi in occupationally exposed persons in the belgrade area, serbia".ti. and "2015".yr. (1)

221	"Laboratory diagnosis of lyme borreliosis in scottish patients: A novel approach".ti. and "2014".yr. (1)

222	"Mycosis fungoides: Is it a borrelia burgdorferi-associated disease?".ti. and "2006".yr. (1)

223	"Morphoea: A manifestation of infection with borrelia species?".ti. and "2007".yr. (1)

224	"Lyme neuroborreliosis: A treatable cause of acute ocular motor disturbances in children".ti. and "2015".yr. (1)

225	"Intermediate uveitis in children and young adults: Differences in clinical course, associations and visual outcome".ti. and "2014".yr. (1)

226	"Clinico-biological aspects in human neuroborreliosis".ti. and "2013".yr. (0)

227	"Awareness that dogs can be carriers for ticks that transmit lyme disease".ti. and "2016".yr. (0)

228	"Surveillance for lyme disease in canada, 2009 to 2012".ti. and "2015".yr. (1)

229	"Descriptive epidemiology of lyme disease in ontario: 1999-2004".ti. and "2006".yr. (1)

230	"Lyme disease: Knowledge, beliefs, and practices of physicians in a low-endemic area".ti. and "2012".yr. (1)

231	"Lyme disease: Knowledge and practices of family practitioners in southern quebec".ti. and "2015".yr. (1)

232	"Challenges with lyme disease diagnostics".ti. and "2012".yr. (0)

233	"Evaluation of a new health canada-approved lyme immunoblot assay for confirmation of positive eia screen results".ti. and "2011".yr. (0)

234	"Immune complexes in early lyme disease".ti. and "2007".yr. (1)

235	"A systematic review and meta-analysis for the adverse effects, immunogenicity and efficacy of lyme disease vaccines: Guiding novel vaccine development".ti. and "2017".yr. (1)

236	"Seroprevalence study of tick-borne encephalitis, borrelia burgdorferi, dengue and toscana virus in turin province".ti. and "2007".yr. (1)

237	"An enhanced elispot assay for sensitive detection of antigen-specific t cell responses to borrelia burgdorferi".ti. and "2013".yr. (1)

238	"Can elispot be applied to a clinical setting as a diagnostic utility for neuroborreliosis?".ti. and "2012".yr. (1)

239	"Study of awareness of tick-borne diseases among children and young people in the czech republic".ti. and "2003".yr. (1)

240	"Long term and repeated electron microscopy and pcr detection of borrelia burgdorferi sensu lato after an antibiotic treatment".ti. and "2004".yr. (1)

241	"Direct detection of borrelia burgdorferi spirochetes in patients with early disseminated lyme borreliosis".ti. and "2009".yr. (1)

242	"Pilot cross-sectional study of three zoonoses (lyme disease, tularaemia, leptospirosis) among healthy blood donors in eastern slovakia".ti. and "2015".yr. (1)

243	"Tick-borne encephalitis: Course and complications - our observations from 2009 to 2012".ti. and "2014".yr. (0)

244	"Standard criteria of western blot for the diagnosis of lyme disease caused by borrelia garinii in china".ti. and "2005".yr. (0)

245	"Diagnosis of lyme borreliosis by polymerase chain reaction".ti. and "2002".yr. (1)

246	"Ability of the borreliacidal antibody test to confirm lyme disease in clinical practice".ti. and "2002".yr. (1)

247	"Detection of borrelia burgdorferi by species-specific and broad-range pcr of synovial fluid and synovial tissue of lyme arthritis patients before and after antibiotic treatment".ti. and "2003".yr. (1)

248	"Disease-specific diagnosis of coinfecting tickborne zoonoses: Babesiosis, human granulocytic ehrlichiosis, and lyme disease".ti. and "2002".yr. (1)

249	"Prospective study of coinfection in patients with erythema migrans".ti. and "2003".yr. (1)

250	"Asymptomatic infection with borrelia burgdorferi".ti. and "2003".yr. (1)

251	"Parental knowledge of and attitudes toward lymerix (recombinant osp--a lyme vaccine)".ti. and "2002".yr. (1)

252	"An open-label, nonrandomized, single-center, prospective extension, clinical trial of booster dose schedules to assess the safety profile and immunogenicity of recombinant outer-surface protein a (ospa) lyme disease vaccine".ti. and "2003".yr. (1)

253	"Serodiagnosis of lyme borreliosis infection using surface plasmon resonance".ti. and "2008".yr. (1)

254	"Long-term effects of immunization with recombinant lipoprotein outer surface protein a on serologic test for lyme disease".ti. and "2004".yr. (1)

255	"Comparison of western immunoblotting and the c6 lyme antibody test for laboratory detection of lyme disease".ti. and "2004".yr. (1)

256	"Ccl19 as a chemokine risk factor for post-treatment lyme disease syndrome: A prospective clinical cohort study".ti. and "2016".yr. (0)

257	"Misdiagnosis of late-onset lyme arthritis by inappropriate use of borrelia burgdorferi immunoblot testing with synovial fluid".ti. and "2012".yr. (1)

258	"Rapid, simple, quantitative, and highly sensitive antibody detection for lyme disease".ti. and "2010".yr. (1)

259	"Lack of serum antibodies against borrelia burgdorferi in children with autism".ti. and "2013".yr. (1)

260	"Humoral immune responses in patients with lyme neuroborreliosis".ti. and "2010".yr. (1)

261	"Study of a cohort of 1,886 persons to determine changes in antibody reactivity to borrelia burgdorferi 3 months after a tick bite".ti. and "2015".yr. (1)

262	"Simple objective detection of human lyme disease infection using immuno-pcr and a single recombinant hybrid antigen".ti. and "2014".yr. (1)

263	"Significantly improved accuracy of diagnosis of early lyme disease by peptide enzyme-linked immunosorbent assay based on the borreliacidal antibody epitope of borrelia burgdorferi ospc".ti. and "2008".yr. (1)

264	"Evaluation of the recombinant vlse-based liaison chemiluminescence immunoassay for detection of borrelia burgdorferi and diagnosis of lyme disease".ti. and "2008".yr. (1)

265	"A decrease in the immunoglobulin g antibody response against the vlse protein of borrelia burgdorferi sensu lato correlates with the resolution of clinical signs in antibiotic-treated patients with early lyme disease".ti. and "2006".yr. (1)

266	"Multiplex immunoassay for lyme disease using vlse1-igg and pepc10-igm antibodies: Improving test performance through bioinformatics".ti. and "2011".yr. (1)

267	"Evaluation of selected borrelia burgdorferi lp54 plasmid-encoded gene products expressed during mammalian infection as antigens to improve serodiagnostic testing for early lyme disease".ti. and "2015".yr. (1)

268	"Impact of clinical variables on borrelia burgdorferi-specific antibody seropositivity in acute-phase sera from patients in north america with culture-confirmed early lyme disease".ti. and "2008".yr. (1)

269	"A novel multivalent ospa vaccine against lyme borreliosis is safe and immunogenic in an adult population previously infected with borrelia burgdorferi sensu lato".ti. and "2014".yr. (1)

270	"Evaluating frequency, diagnostic quality, and cost of lyme borreliosis testing in germany: A retrospective model analysis".ti. and "2012".yr. (1)

271	"The impact of immunosuppression on erythema migrans. A retrospective study of clinical presentation, response to treatment and production of borrelia antibodies in 33 patients".ti. and "2006".yr. (1)

272	"Frequency and clinical features of intraocular inflammation in tokyo".ti. and "2009".yr. (1)

273	"Uveitis in children: Single center experience [p064]".ti. and "2011".yr. (0)

274	"Automated cerebrospinal fluid cell count--new reference ranges and evaluation of its clinical use in central nervous system infections".ti. and "2014".yr. (1)

275	"Liaison (r) borrelia assay: A recombinant antigen based, automated chemiluminescence immunoassay for the diagnosis of lyme disease".ti. and "2006".yr. (0)

276	"Clinical comparison of two commercially available lyme screening elisa assays".ti. and "2007".yr. (0)

277	"Development and validation of a novel ifn-gamma elispot assay for sensitive and specific detection of antigen-specific t cell response to borrelia burgdorferi".ti. and "2013".yr. (0)

278	"Validation and correlation of vidas iggii and igmii lyme antibodies vs vidas lyme igg/igm in the laboratory evaluation of lyme disease: Clinical implications".ti. and "2015".yr. (0)

279	"Method comparison of two elisa platforms for the detection of b.Burgdorferi (lyme) antibodies; c6 specific peptide vs. Whole cell sonicate".ti. and "2013".yr. (0)

280	"Bead based multiplex immunoassays for the laboratory neuroborreliosis".ti. and "2012".yr. (0)

281	"Detection of borrelia burgdorferi in formalin fixed paraffin embedded skin biopsies".ti. and "2012".yr. (0)

282	"Enzyme-linked immunospot assay in the laboratory diagnostics of lyme borreliosis".ti. and "2015".yr. (0)

283	"2-tiered antibody testing for early and late lyme disease using only an immunoglobulin g blot with the addition of a vlse band as the second-tier test".ti. and "2010".yr. (1)

284	"Two-tiered antibody testing for lyme disease with use of 2 enzyme immunoassays, a whole-cell sonicate enzyme immunoassay followed by a vlse c6 peptide enzyme immunoassay".ti. and "2011".yr. (1)

285	"Performance of united states serologic assays in the diagnosis of lyme borreliosis acquired in europe".ti. and "2013".yr. (1)

286	"A comparison of lyme disease serologic test results from 4 laboratories in patients with persistent symptoms after antibiotic treatment".ti. and "2014".yr. (1)

287	"Lyme disease testing by large commercial laboratories in the united states".ti. and "2014".yr. (1)

288	"Lyme disease and human granulocytic anaplasmosis coinfection: Impact of case definition on coinfection rates and illness severity".ti. and "2013".yr. (1)

289	"Higher mrna levels of chemokines and cytokines associated with macrophage activation in erythema migrans skin lesions in patients from the united states than in patients from austria with lyme borreliosis".ti. and "2008".yr. (1)

290	"Antibiotic treatment duration and long-term outcomes of patients with early lyme disease from a lyme disease-hyperendemic area".ti. and "2010".yr. (1)

291	"Poor positive predictive value of lyme disease serologic testing in an area of low disease incidence".ti. and "2015".yr. (1)

292	"Evaluation of the c6 lyme enzyme immunoassay for the diagnosis of lyme disease in children and adolescents".ti. and "2016".yr. (1)

293	"Borrelial lymphocytoma in adult patients".ti. and "2016".yr. (1)

294	"Course and outcome of early lyme borreliosis in patients with hematological malignancies".ti. and "2015".yr. (1)

295	"Xenodiagnosis to detect borrelia burgdorferi infection: A first-in-human study".ti. and "2014".yr. (1)

296	"Development of a metabolic biosignature for detection of early lyme disease".ti. and "2015".yr. (1)

297	"Blood cultures for patients with extracutaneous manifestations of lyme disease in the united states".ti. and "2009".yr. (1)

298	"Suspected early lyme neuroborreliosis in patients with erythema migrans".ti. and "2013".yr. (1)

299	("Course and outcome of early european lyme neuroborreliosis" and "Clinical and laboratory findings").ti. and "2016".yr. (1)

300	"Lyme disease diagnosed by alternative methods: A phenotype similar to that of chronic fatigue syndrome".ti. and "2015".yr. (1)

301	"Prospective study of serologic tests for lyme disease".ti. and "2008".yr. (1)

302	"Comparison of findings for patients with borrelia garinii and borrelia afzelii isolated from cerebrospinal fluid".ti. and "2006".yr. (1)

303	"Treatment of erythema migrans with doxycycline for 10 days versus 15 days".ti. and "2012".yr. (1)

304	"Incidence and patterns of extended-course antibiotic therapy in patients evaluated for lyme disease".ti. and "2015".yr. (1)

305	"Long-term assessment of post-treatment symptoms in patients with culture-confirmed early lyme disease".ti. and "2015".yr. (1)

306	"Long-term follow-up of patients with lyme disease: Longitudinal analysis of clinical and quality-of-life measures".ti. and "2016".yr. (1)

307	"Extensive limb swelling after immunization: Reports to the vaccine adverse event reporting system".ti. and "2003".yr. (1)

308	"Effect of borrelia burgdorferi genotype on the sensitivity of c6 and 2-tier testing in north american patients with culture-confirmed lyme disease".ti. and "2008".yr. (1)

309	"Long-term assessment of health-related quality of life in patients with culture-confirmed early lyme disease".ti. and "2015".yr. and Wormser.ax. (1)

310	"Evaluation of a new multiparametric microspot array for serodiagnosis of lyme borreliosis".ti. and "2015".yr. (1)

311	"Lyme disease in the u.K.: Clinical and laboratory features and response to treatment".ti. and "2010".yr. (1)

312	"Comparison of pcr methods and culture for the detection of borrelia spp. In patients with erythema migrans".ti. and "2008".yr. (1)

313	"Seroprevalence of tick-borne infections in forestry rangers from northeastern italy".ti. and "2004".yr. (1)

314	"Ticking the right boxes: Classification of patients suspected of lyme borreliosis at an academic referral center in the netherlands".ti. and "2015".yr. (1)

315	"Anaplasma phagocytophilum infection in patients with early lyme borreliosis, erythema migrans, in north-eastern poland".ti. and "2009".yr. (1)

316	"Neuroborreliosis--an epidemiological, clinical and healthcare cost study from an endemic area in the south-east of sweden".ti. and "2010".yr. (1)

317	"Analysis of borrelia burgdorferi igg antibodies with a combination of igg elisa and vlse c6 peptide elisa".ti. and "2005".yr. (1)

318	"Peripheral facial palsy in patients with tick-borne encephalitis".ti. and "2012".yr. (1)

319	"Testing patients with non-specific symptoms for antibodies against borrelia burgdorferi sensu lato does not provide useful clinical information about their aetiology".ti. and "2015".yr. (1)

320	"Lyme borreliosis caused by diverse genospecies of borrelia burgdorferi sensu lato in northeastern china".ti. and "2014".yr. (1)

321	"Seroprevalence of seven pathogens transmitted by the ixodes ricinus tick in forestry workers in france".ti. and "2016".yr. (1)

322	"Comparison of isolation rate of borrelia burgdorferi sensu lato in two different culture media, mkp and bsk-h".ti. and "2014".yr. (1)

323	"High frequency of false positive igm immunoblots for borrelia burgdorferi in clinical practice".ti. and "2012".yr. (1)

324	"Comparison of five different immunoassays for the detection of borrelia burgdorferi igm and igg antibodies".ti. and "2006".yr. (1)

325	"High sensitivity and specificity of the c6-peptide elisa on cerebrospinal fluid in lyme neuroborreliosis patients".ti. and "2011".yr. (1)

326	"Performance of four commercial immunoblots for serological confirmation of lyme borreliosis in rhone-alpes (france)".ti. and "2009".yr. (0)

327	"Lyme borreliosis: Descriptive study of 151 patients followed up in a university hospital, lyon (france)".ti. and "2009".yr. (0)

328	"Value of the lymphocyte transformation test to determine the acuity of neuroborreliosis".ti. and "2012".yr. (0)

329	"Evaluation of two automated tests on the vidas instrument to detect anti-lyme disease antibodies igg and igm in human serum, plasma and csf".ti. and "2010".yr. (0)

330	"Molecular identification of borrelia spirochetes isolated from skin biopsies of patients characterised with an unusual dermal manifestation (prurigo pigmentosa) in taiwan".ti. and "2012".yr. (0)

331	"Preliminary study on immunological reactivity in people occupationally exposed to tick-transmitted pathogens".ti. and "2009".yr. (0)

332	"Methodology in reporting diagnostic accuracy for borrelia specific igg and igm antibodies. Tests often differ but performance of the test antigens may be quite similar anyway".ti. and "2012".yr. (0)

333	"Direct detection of early lyme borreliosis from whole blood".ti. and "2011".yr. (0)

334	"German investigation of lyme borreliosis: Evaluation of therapeutic and diagnostic cost (gilead)".ti. and "2011".yr. (0)

335	"Detection of borrelia burgdorferi sensu lato in urine specimens from patients with early and late lyme borreliosis".ti. and "2012".yr. (0)

336	"Preferential reactivity towards borrelia afzelii in the sera of patients with lyme borreliosis from the north-east of poland".ti. and "2010".yr. (0)

337	"Combining c6 elisa and igm immunoblot for the detection of antibodies in early lyme infection".ti. and "2010".yr. (0)

338	"Lyme borreliosis and tick-borne encephalitis in latvia: A comparative study of cases reported in 2007-2010".ti. and "2012".yr. (0)

339	"Course and outcome of erythema migrans in patients with underlying rheumatological disease".ti. and "2011".yr. (0)

340	"Neuroborreliosis: Detection of intrathecally produced antibodies against borrelia in cerebrospinal fluid using the line blot immunoassay anti-borrelia euroline-rn-at".ti. and "2011".yr. (0)

341	"Correlation between indirect immunofluorescence and western blot, serological techniques used for the diagnosis of lyme disease".ti. and "2012".yr. (0)

342	"Spectrum of clinical forms and seasonal distribution of lyme borreliosis in children".ti. and "2010".yr. (0)

343	"Co-infection with borrelia burgdorferi and tick-borne encephalitis virus in humans, ticks and tick cells - analysis of clinical cases, literature and experimental possibilities".ti. and "2012".yr. (0)

344	"Performance evaluation of the new vidas lyme igm and igg assays compared to the previous vidas lyme total antibodies assay on fresh prospective sera".ti. and "2012".yr. (0)

345	"Dimeric ospcadv - advanced recombinant ospc antigens for the sensitive and specific detection of class igm antibodies in early stages of borrelia infections".ti. and "2011".yr. (0)

346	"Evaluation of different immunoassays on liaison".ti. and "2009".yr. (0)

347	"Infectious diseases among eufor military personnel".ti. and "2011".yr. (0)

348	"Childhood neuroborreliosis does not lead to long-term cognitive disturbances".ti. and "2011".yr. (0)

349	"Patients after proven neuroborreliosis - how severe is the persisting neuropsychological damage?".ti. and "2009".yr. (0)

350	"Neuroborreliosis in bulgaria - clinical manifestation and serological findings".ti. and "2011".yr. (0)

351	"Clinical manifestations of lyme borreliosis in bulgaria - comparison with tick studies".ti. and "2010".yr. (0)

352	"Evaluation of the new vidas lyme igm and vidas lyme igg kits as screening test for the serological diagnosis of lyme borreliosis".ti. and "2012".yr. (0)

353	"Evaluation of two line immunoassays for the detection of borrelia antibodies".ti. and "2009".yr. (0)

354	"Attention and learning in children with lyme disease and children with attention deficit hyperactivity disorder".ti. and "2013".yr. (0)

355	"Personality function in chronic lyme disease".ti. and "2008".yr. (0)

356	"Spect brain imaging in chronic lyme disease".ti. and "2012".yr. (1)

357	"Lyme arthritis in 20 children residing in a non-endemic area".ti. and "2005".yr. (1)

358	"Prolonged synovitis in pediatric lyme arthritis cannot be predicted by clinical or laboratory parameters".ti. and "2009".yr. (1)

359	"Effect of latitude on the rate of change in incidence of lyme disease in the united states".ti. and "2013".yr. (1)

360	"Treatment for the neurological complications of lyme disease".ti. and "2008".yr. (0)

361	"Antibiotics for the neurological complications of lyme disease".ti. and "2016".yr. (1)

362	"A comparison of analysis units for associating lyme disease with forest-edge habitat".ti. and "2006".yr. (0)

363	"Serologically detected new tick-borne zoonoses in eastern croatia".ti. and "2003".yr. (1)

364	"Systematic analysis of wide-field fundus autofluorescence (faf) imaging in posterior uveitis".ti. and "2014".yr. (1)

365	"Season is an unreliable predictor of lyme neuroborreliosis".ti. and "2015".yr. (1)

366	"Lyme disease trends: Delaware, 2000 - 2004".ti. and "2007".yr. (1)

367	"Comparison of a two- or three-week regimen and a review of treatment of erythema migrans with phenoxymethylpenicillin".ti. and "2006".yr. (1)

368	"Mini erythema migrans--a sign of early lyme borreliosis".ti. and "2006".yr. (1)

369	"Gabapentin for the symptomatic treatment of chronic neuropathic pain in patients with late-stage lyme borreliosis: A pilot study".ti. and "2005".yr. (1)

370	"Bull's-eye and nontarget skin lesions of lyme disease: An internet survey of identification of erythema migrans".ti. and "2012".yr. (1)

371	"Utility of a commercial immunoblot kit (bag-borrelia blot) in the diagnosis of the preliminary stages of lyme disease".ti. and "2003".yr. (1)

372	"A dutch nationwide evaluation of serological assays for detection of borrelia antibodies in clinically well-defined patients".ti. and "2015".yr. (1)

373	"Percent positive rate of lyme real-time polymerase chain reaction in blood, cerebrospinal fluid, synovial fluid, and tissue".ti. and "2008".yr. (1)

374	"Evaluation of commercial screening tests and blot assays for the diagnosis of lyme borreliosis".ti. and "2012".yr. (1)

375	"Presence of anti-borrelia burgdorferi antibodies and borrelia burgdorferi sensu lato dna in samples of subjects in an area of the northern italy in the period 2002-2008".ti. and "2011".yr. (1)

376	"Antigen biochips verify and extend the scope of antibody detection in lyme borreliosis".ti. and "2007".yr. (1)

377	"A lightcycler taqman assay for detection of borrelia burgdorferi sensu lato in clinical samples".ti. and "2007".yr. (1)

378	"Comparison of five diagnostic modalities for direct detection of borrelia burgdorferi in patients with early lyme disease".ti. and "2012".yr. (1)

379	"Pregnant women with erythema migrans and isolation of borreliae from blood: Course and outcome after treatment with ceftriaxone".ti. and "2011".yr. (1)

380	"Cross-reactivity between lyme and syphilis screening assays: Lyme disease does not cause false-positive syphilis screens".ti. and "2016".yr. (1)

381	"The c6 lyme antibody test has low sensitivity for antibody detection in cerebrospinal fluid".ti. and "2009".yr. (1)

382	"Comparison of males versus females with culture-confirmed early lyme disease at presentation and at 11-20 years after diagnosis".ti. and "2016".yr. (1)

383	"Single-tier testing with the c6 peptide elisa kit compared with two-tier testing for lyme disease".ti. and "2013".yr. (1)

384	"Tick-borne diseases: An evaluation of a lyme disease prevention education program for eight, nine, and ten year olds".ti. and "2009".yr. (0)

385	"Understanding lyme disease: Illness experience, prevention, and the health belief model".ti. and "2008".yr. (0)

386	"Quaternary lyme disease: Symptomatology, epidemiology and anthropology of an emerging disease".ti. and "2015".yr. (0)

387	"Opportunities and barriers in the control and prevention of lyme disease: Implications for practice".ti. and "2015".yr. (0)

388	"A comparison of memory and executive functioning in children with lyme disease and children with attention-deficit/hyperactivity disorder".ti. and "2014".yr. (0)

389	"A national survey of risk perceptions and practices to prevent tick-borne lyme disease and mosquito-borne viral encephalitis".ti. and "2003".yr. (0)

390	"Spatial analysis of human lyme disease risk in an endemic county".ti. and "2015".yr. (0)

391	"Memory and executive functions in adolescents with lyme disease".ti. and "2007".yr. (0)

392	"Personality traits, perceived stress, and coping styles in patients with chronic lyme disease and fibromyalgia".ti. and "2009".yr. (0)

393	"Coping, depression, and quality of life of individuals with chronic lyme disease".ti. and "2015".yr. (0)

394	"Audiologic manifestations of patients with post-treatment lyme disease syndrome".ti. and "2003".yr. (1)

395	"Disease risk in a dynamic environment: The spread of tick-borne pathogens in minnesota, usa".ti. and "2015".yr. (1)

396	"An examination of the demographic and environmental variables correlated with lyme disease emergence in virginia".ti. and "2015".yr. (1)

397	"Effects of landscape fragmentation and climate on lyme disease incidence in the northeastern united states".ti. and "2013".yr. (1)

398	"Increasing residential proximity of lyme borreliosis cases to high-risk habitats: A retrospective study in central bohemia, the czech republic, 1987-2010".ti. and "2015".yr. (1)

399	"Mapping lyme disease incidence for diagnostic and preventive decisions, maryland".ti. and "2002".yr. (1)

400	"Infections with candidatus neoehrlichia mikurensis and cytokine responses in 2 persons bitten by ticks, sweden".ti. and "2015".yr. (1)

401	"Lyme disease, virginia, usa, 2000-2011".ti. and "2014".yr. (1)

402	"Hypersensitivity to ticks and lyme disease risk".ti. and "2005".yr. (1)

403	"Enhancing lyme disease surveillance by using administrative claims data, tennessee, usa".ti. and "2015".yr. (1)

404	"Effect of surveillance method on reported characteristics of lyme disease, connecticut, 1996-2007".ti. and "2012".yr. (1)

405	"No geographic correlation between lyme disease and death due to 4 neurodegenerative disorders, united states, 2001-2010".ti. and "2015".yr. (1)

406	"Epidemiology of lyme disease, nova scotia, canada, 2002-2013".ti. and "2015".yr. (1)

407	"Borrelia miyamotoi sensu lato seroreactivity and seroprevalence in the northeastern united states".ti. and "2014".yr. (1)

408	"Geographic distribution and expansion of human lyme disease, united states".ti. and "2015".yr. (1)

409	"Bartonella spp. Bacteremia and rheumatic symptoms in patients from lyme disease-endemic region".ti. and "2012".yr. and Maggi.ax. (1)

410	"Incidence of clinician-diagnosed lyme disease, united states, 2005-2010".ti. and "2015".yr. (1)

411	"Lyme disease in hispanics, united states, 2000-2013".ti. and "2016".yr. (1)

412	"Human babesiosis, maine, usa, 1995-2011".ti. and "2014".yr. (1)

413	"Effectiveness of personal protective measures to prevent lyme disease".ti. and "2008".yr. (1)

414	"Antibodies against borrelia burgdorferi sensu lato among adults, germany, 2008-2011".ti. and "2015".yr. (1)

415	"Economic impact of lyme disease".ti. and "2006".yr. (1)

416	"Spatio-temporal patterns in county-level incidence and reporting of lyme disease in the northeastern united states, 1990-2000".ti. and "2007".yr. (0)

417	"Temporal and spatial distribution of tick-borne disease cases among humans and canines in illinois (2000-2009)".ti. and "2014".yr. (1)

418	"Validating negative binomial lyme disease regression model with bootstrap resampling".ti. and "2016".yr. (0)

419	"Variability in results from negative binomial models for lyme disease measured at different spatial scales".ti. and "2015".yr. (1)

420	"Lyme borreliosis--epidemiological analysis of incidence in the northern region of slovakia".ti. and "2012".yr. (1)

421	"Improvement of diagnostic approach to lyme neuroborreliosis in children by using recombinant antigens in detection of intrathecally produced igm/igg".ti. and "2016".yr. (1)

422	"Synoptic spatial classification and lyme disease in new york state (1991-2006)".ti. and "2012".yr. (0)

423	"Lyme borreliosis in southern united kingdom and a case for a new syndrome, chronic arthropod-borne neuropathy".ti. and "2015".yr. (1)

424	"Lyme disease in france: A primary care-based prospective study".ti. and "2005".yr. (1)

425	"Neuroborreliosis in the south west of england".ti. and "2008".yr. (1)

426	"Tick-borne encephalitis and lyme borreliosis in belarus republic, 1998-2007".ti. and "2008".yr. (0)

427	"Epidemiology of tick-borne encephalitis and lyme disease in the republic of belarus, 1998-2007".ti. and "2009".yr. (0)

428	"Epidemiological situation for tick-borne encephalitis and lyme disease in the republic of karelia, russia, in 2002-2011".ti. and "2012".yr. (0)

429	"Incidence of tick-borne encephalitis and lyme disease in leninggrad oblast".ti. and "2012".yr. (0)

430	"Serological evidence for human granulocytic ehrlichiosis in western austria".ti. and "2003".yr. (1)

431	"The phenomenon of chronic lyme ; an observational study".ti. and "2012".yr. (1)

432	"Clinical data and cerebrospinal fluid findings in lyme meningitis versus aseptic meningitis".ti. and "2003".yr. (1)

433	"Improving national surveillance of lyme neuroborreliosis in denmark through electronic reporting of specific antibody index testing from 2010 to 2012".ti. and "2015".yr. (1)

434	"Lyme borreliosis in the netherlands: Strong increase in gp consultations and hospital admissions in past 10 years".ti. and "2006".yr. (1)

435	"Laboratory diagnosis of lyme borreliosis at the portuguese national institute of health (1990-2004)".ti. and "2006".yr. (1)

436	"Surveillance of lyme borreliosis in germany, 2002 and 2003".ti. and "2005".yr. and Maggi.ax. (0)

437	"Disseminated and chronic lyme borreliosis in norway, 1995 - 2004".ti. and "2005".yr. and borreliosis.kw. (1)

438	"Lyme borreliosis: Europe-wide coordinated surveillance and action needed?".ti. and "2006".yr. (1)

439	"Incidence and hospitalisation rates of lyme borreliosis, france, 2004 to 2012".ti. and "2014".yr. (1)

440	"Lv and rv function in patients with lyme disease diagnosed with endomyocardial biopsy-a long term follow-up".ti. and "2012".yr. (0)

441	"Is improvement of the left ventricular systolic function in patients with new-onset dilated cardiomyopathy related to the presence of virus or inflammation in endomycardial biopsy?".ti. and "2012".yr. (0)

442	"Has serological testing of antibodies to cardiotropic agents any role in the diagnosis of inflammatory cardiomyopathy?".ti. and "2010".yr. (0)

443	"Large differences between test strategies for the detection of anti-borrelia antibodies are revealed by comparing eight elisas and five immunoblots".ti. and "2011".yr. (1)

444	"Increased incidence of lyme borreliosis in southern sweden following mild winters and during warm, humid summers".ti. and "2006".yr. (1)

445	"Borrelia burgdorferi sensu lato serology in the netherlands: Guidelines versus daily practice".ti. and "2014".yr. (1)

446	"Risk of acquiring tick-borne infections in forestry workers from lazio, italy".ti. and "2010".yr. (1)

447	"Management of paediatric lyme disease in non-endemic and endemic areas: Data from the registry of the italian society for pediatric infectious diseases".ti. and "2013".yr. (1)

448	"Pre-treatment and post-treatment assessment of the c(6) test in patients with persistent symptoms and a history of lyme borreliosis".ti. and "2004".yr. (1)

449	"Laboratory diagnosis of lyme neuroborreliosis: A comparison of three csf anti-borrelia antibody assays".ti. and "2014".yr. (1)

450	"Seroreactivity for spotted fever rickettsiae and co-infections with other tick-borne agents among habitants in central and southern sweden".ti. and "2013".yr. (1)

451	"Borrelia burgdorferi vlse antigen for the serological diagnosis of lyme borreliosis".ti. and "2008".yr. (1)

452	"Co-infections with borrelia species, anaplasma phagocytophilum and babesia spp. In patients with tick-borne encephalitis".ti. and "2014".yr. (1)

453	"Risk of borrelia burgdorferi infection in western switzerland following a tick bite".ti. and "2004".yr. (1)

454	"Duration of antibiotic treatment in disseminated lyme borreliosis: A double-blind, randomized, placebo-controlled, multicenter clinical study".ti. and "2007".yr. (1)

455	"Comparison of an automated borrelia indirect chemiluminescent immunoassay (clia) with a vlse/c6 elisa and immunoblot".ti. and "2007".yr. (1)

456	"Lyme meningoradiculitis: Prospective evaluation of biological diagnosis methods".ti. and "2007".yr. (1)

457	"Differential diagnoses of suspected lyme borreliosis or post-lyme-disease syndrome".ti. and "2007".yr. (1)

458	"Sensitivity and specificity of a commercial c6 peptide enzyme immuno assay in diagnosis of acute lyme neuroborreliosis".ti. and "2007".yr. (1)

459	"Evaluation of two commercially available rapid diagnostic tests for lyme borreliosis".ti. and "2015".yr. (1)

460	"C6 peptide elisa test in the serodiagnosis of lyme borreliosis in sweden".ti. and "2007".yr. (1)

461	"Prevalence and incidence of antibodies to borrelia burgdorferi and to tick-borne encephalitis virus in agricultural and forestry workers from tuscany, italy".ti. and "2005".yr. (1)

462	"Incidence and management of presumption of lyme borreliosis in belgium: Recent data from the sentinel network of general practitioners".ti. and "2012".yr. (1)

463	"The improvement of left ventricular systolic function induced by antibiotic therapy in patients with inflammatory cardiomyopathy caused by borrelia burgdorferi infection".ti. and "2010".yr. (0)

464	"Hypothalamic-pituitary insufficiency following infectious diseases of the central nervous system".ti. and "2008".yr. (1)

465	"North atlantic weather oscillation and human infectious diseases in the czech republic, 1951-2003".ti. and "2005".yr. (1)

466	"Screening of the prevalence of antibodies to borrelia burgdorferi in madrid province, spain".ti. and "2004".yr. (1)

467	"Detection of borrelia burgdorferi sensu lato in endomyocardial biopsy specimens in individuals with recent-onset dilated cardiomyopathy".ti. and "2012".yr. (1)

468	"Comprehensive assessment of endomyocardial biopsy specimens in patienswith recent-onset dilated cardiomyopathy".ti. and "2011".yr. (0)

469	"Borrelia burgdorferi infection in iranian patients with multiple sclerosis".ti. and "2009".yr. (0)

470	"Clinical effects of fluconazole in patients with neuroborreliosis".ti. and "2004".yr. (1)

471	"Parkinson's syndrome in chronic neuroborreliosis".ti. and "2012".yr. (0)

472	"Oral doxycycline for lyme neuroborreliosis with symptoms of encephalitis, myelitis, vasculitis or intracranial hypertension".ti. and "2014".yr. (1)

473	"A study on the association between infectious burden and alzheimer's disease".ti. and "2015".yr. (1)

474	"Borreliosis. Cause of obstructive sleep apnoea syndrome".ti. and "2008".yr. (0)

475	"The prevalence of neuropsychiatric symptoms in lyme borreliosis patients".ti. and "2015".yr. (0)

476	"Efficacy and safety of pharmacological treatments for acute lyme neuroborreliosis - a systematic review".ti. and "2015".yr. (1)

477	"The diagnostic spectrum in patients with suspected chronic lyme neuroborreliosis--the experience from one year of a university hospital's lyme neuroborreliosis outpatients clinic".ti. and "2011".yr. (1)

478	"European neuroborreliosis: Neuropsychological findings 30 months post-treatment".ti. and "2012".yr. (1)

479	"Clinical and immunological characteristics of multiple sclerosis patients with b.Burgdorferi antibodies".ti. and "2008".yr. (0)

480	"Clinical and serological features of borrelial infection in multiple sclerosis patients".ti. and "2010".yr. (0)

481	"Antibodies against ebv, cmv, vzv, hsv-1, hsv-2, hhv-6b and borrelia burgdorferi and viral nucleic acids in serum and csf samples of patients with multiple sclerosis".ti. and "2012".yr. (0)

482	"Borreliosis and associated autoimmunity".ti. and "2016".yr. (0)

483	"Oral doxycycline vs. Iv centriaxone for european lume neuro-borreliosis. A double-blind, randomized controlled clinical trial".ti. and "2008".yr. (0)

484	"Remaining complaints 1 year after treatment for acute lyme neuroborreliosis; frequency, pattern and risk factors".ti. and "2010".yr. (1)

485	"Clinical usefulness of intrathecal antibody testing in acute lyme neuroborreliosis".ti. and "2007".yr. (1)

486	"Borrelia burgdorferi: Risk factor in alzheimer's disease".ti. and "2011".yr. (0)

487	"Borrelia burgdorferi: Risk factor in alzheimer's disease p1021.[abstract]".ti. and "2012".yr. (0)

488	"Chronic polyneuropathy and lyme disease".ti. and "2006".yr. (1)

489	"Antibiotic therapy in patients with the combination of definite multiple sclerosis and chronic lyme disease".ti. and "2012".yr. (0)

490	"Peripheral facial palsy associated with viral infections in children".ti. and "2011".yr. (0)

491	"Outcome of peripheral facial palsy in children - a catamnestic study".ti. and "2013".yr. (1)

492	"Canalicular magnetic stimulation lacks specificity to differentiate idiopathic facial palsy from borreliosis in children".ti. and "2008".yr. (1)

493	"Facial palsy: Etiology, outcome and management in children".ti. and "2011".yr. (1)

494	"Uncommon synovial cysts in children".ti. and "2006".yr. (1)

495	"Pediatric tick-borne infections of the central nervous system in an endemic region of sweden: A prospective evaluation of clinical manifestations".ti. and "2012".yr. (1)

496	"Tick bites, erythema migrans and lyme borreliosis: Incidence and management by the gp in belgium".ti. and "2004".yr. (0)

497	"The burden of lyme borreliosis expressed in disability-adjusted life years".ti. and "2015".yr. (1)

498	"Cognitive functioning in lyme borreliosis".ti. and "2004".yr. (0)

499	"Mental disorders in patients with lyme disease".ti. and "2003".yr. (0)

500	"Clinical and demographic characteristics of psychiatric patients seropositive for borrelia burgdorferi".ti. and "2006".yr. (1)

501	("Two great masqueraders" and "in a psychiatry clinic-case series of neurosyphilis and neuroborreliosis").ti. and "2012".yr. (0)

502	"Analysis of weather effects on variability in lyme disease incidence in the northeastern united states".ti. and "2002".yr. (1)

503	"Evaluation of clinical specimens for rickettsia, bartonella, borrelia, coxiella, anaplasma, franciscella and diplorickettsia positivity using serological and molecular biology methods".ti. and "2012".yr. (1)

504	"Detection of borrelia burgdorferi sensu lato dna by pcr in serum of patients with clinical symptoms of lyme borreliosis".ti. and "2008".yr. (1)

505	"Cerebrospinal-fluid profile in neuroborreliosis and its diagnostic significance".ti. and "2006".yr. (1)

506	"Short-term stability of borrelia garinii in cerebrospinal fluid".ti. and "2016".yr. (1)

507	"Dna persistence after treatment of lyme borreliosis".ti. and "2014".yr. (1)

508	"Lyme borreliosis--analysis of the trends in slovakia, 1999-2008".ti. and "2011".yr. (1)

509	"Lipid peroxidation in the pathogenesis of neuroborreliosis".ti. and "2016".yr. (1)

510	"Sex differences in the clinical and serologic presentation of early lyme disease: Results from a retrospective review".ti. and "2010".yr. (1)

511	"A determination of the spatial concordance between lyme disease incidence and habitat probability of its primary vector ixodes scapularis (black-legged tick)".ti. and "2014".yr. (1)

512	"Diluting the dilution effect: A spatial lyme model provides evidence for the importance of habitat fragmentation with regard to the risk of infection".ti. and "2009".yr. (1)

513	"Molecular survey of five tick-borne pathogens (ehrlichia chaffeensis, ehrlichia ewingii, anaplasma phagocytophilum, borrelia burgdorferi sensu lato and babesia microti) in egyptian farmers".ti. and "2011".yr. (0)

514	"Recurrent early pregnancy loss, intrauterine death-causes, diagnosis and therapeutical approach".ti. and "2012".yr. (0)

515	"Making sense of unfamiliar risks in the countryside: The case of lyme disease".ti. and "2011".yr. (1)

516	"Whose lyme is it anyway? Subject positions and the construction of responsibility for managing the health risks from lyme disease".ti. and "2012".yr. (1)

517	"A controlled trial of a novel primary prevention program for lyme disease and other tick-borne illnesses".ti. and "2007".yr. (1)

518	"Developing requirements for a mobile app to support citizens in dealing with ticks and tick bites via end-user profiling".ti. and "2015".yr. (1)

519	"Healthcare access and burden of care for patients with lyme disease: A large united states survey".ti. and "2011".yr. (1)

520	"Increasing burden of lyme carditis in us children's hospitals".ti. and "2015".yr. (0)

521	"Suspected inflammatory cardiomyopathy. Prevalence of borrelia burgdorferi in endomyocardial biopsies with positive serological evidence".ti. and "2015".yr. (1)

522	"Recent-onset dilated cardiomyopathy associated with borrelia burgdorferi infection".ti. and "2015".yr. (1)

523	"Detection of spirochaetal microorganisms by focus floating microscopy in necrobiosis lipoidica in patients from central europe".ti. and "2008".yr. (1)

524	"Lyme borreliosis: Differences between the patients who are tested and those with the infection".ti. and "2008".yr. (0)

525	"Fabric impregnation using acaricides: Effective and safe method for the prevention of tick-infestation and tick-borne diseases".ti. and "2008".yr. (0)

526	"Etiology of tick-borne febrile illnesses in adult residents of north-eastern poland: Report from a prospective clinical study".ti. and "2006".yr. (1)

527	"Disappearance of specific immune response after successful therapy of chronic lyme borreliosis".ti. and "2004".yr. (1)

528	"Serological description of estonian patients with lyme disease, a comparison with control sera from endemic and non-endemic areas".ti. and "2004".yr. (1)

529	"Comparison of different borrelia burgdorferi sensu lato strains for detection of immune response in patients with erythema migrans".ti. and "2008".yr. (1)

530	"Significant improvement of the recombinant borrelia igg immunoblot for serodiagnosis of early neuroborreliosis".ti. and "2004".yr. (1)

531	"Immune responses to borrelial vlse ir6 peptide variants".ti. and "2007".yr. (1)

532	"Antibody responses to borrelia ir(6) peptide variants and the c6 peptide in swedish patients with erythema migrans".ti. and "2009".yr. (1)

533	"Serological evidence for tick-borne encephalitis, borreliosis, and human granulocytic anaplasmosis in mongolia".ti. and "2006".yr. (1)

534	"Lyme borreliosis reinfection: Might it be explained by a gender difference in immune response?".ti. and "2006".yr. (1)

535	"Could co-infection with anaplasma play a role in borrelia-associated primary cutaneous marginal zone b-cell lymphomas?".ti. and "2016".yr. (1)

536	"Factors affecting duration of chronic fatigue syndrome in pediatric patients".ti. and "2012".yr. (1)

537	"Validate or falsify: Lessons learned from a microscopy method claimed to be useful for detecting borrelia and babesia organisms in human blood".ti. and "2016".yr. (1)

538	"Solitary and multiple erythema migrans in children: Comparison of demographic, clinical and laboratory findings".ti. and "2003".yr. (1)

539	"Borrelia antibodies in children evaluated for lyme neuroborreliosis".ti. and "2008".yr. (1)

540	"Oral antibiotic treatment and long-term outcomes of lyme facial nerve palsy".ti. and "2011".yr. (1)

541	"Comparison of erythema migrans caused by borrelia afzelii and borrelia garinii".ti. and "2004".yr. (1)

542	"Isolation of borrelia burgdorferi sensu lato from blood of adult patients with borrelial lymphocytoma, lyme neuroborreliosis, lyme arthritis and acrodermatitis chronica atrophicans".ti. and "2011".yr. (1)

543	"Comparative analysis of the infectivity rate of both borrelia burgdorferi and anaplasma phagocytophilum in humans and dogs in a new jersey community".ti. and "2014".yr. (1)

544	"Evidence for strain-specific immunity in patients treated for early lyme disease".ti. and "2014".yr. (1)

545	"Lyme borreliosis: A european perspective on diagnosis and clinical management".ti. and "2009".yr. (1)

546	"The positive predictive value of borrelia burgdorferi serology in the light of symptoms of patients sent to an outpatient service for tick-borne diseases".ti. and "2010".yr. (1)

547	"Commercial test kits for detection of lyme borreliosis: A meta-analysis of test accuracy".ti. and "2016".yr. (1)

548	"Risk factors in lyme borreliosis: A german case-control study".ti. and "2002".yr. (1)

549	"Seroprevalence of infection with borrelia burgdorferi s. L. In two adjacent regions of eastern france and southwestern germany".ti. and "2002".yr. (1)

550	"Foci of tick-borne diseases in southwest germany".ti. and "2002".yr. (1)

551	"Seroprevalence of borrelia infection in occupational tick-exposed people in bavaria (germany)".ti. and "2002".yr. (1)

552	"Seroprevalence of antibodies against borrelia burgdorferi s. L. And tick-borne encephalitis virus in the bergische land region of northrhine westfalia".ti. and "2002".yr. (1)

553	"Acute facial palsy in children--a 2-year follow-up study with focus on lyme neuroborreliosis".ti. and "2003".yr. (1)

554	"Project management of lyme disease through monitoring of seroprevalence of anti-borrelia antibodies in macedonian patients".ti. and "2014".yr. (0)

555	"Retrospective study of lyme borreliosis positive serologies in 2003".ti. and "2004".yr. (0)

556	"Evaluation of plateliatm lyme igm and plateliatm lyme igg assays for lyme borreliosis diagnosis".ti. and "2007".yr. (0)

557	"Comparison between enzygnost((r)) lyme link vlse/igg and liaison((r)) borrelia igg for the laboratory diagnosis of lyme disease".ti. and "2007".yr. (0)

558	"Evaluation of five commercial screening assays and two commercial immunoblots for the serological diagnosis of lyme borreliosis".ti. and "2007".yr. (0)

559	"Trends in blacklegged tick submissions and lyme disease serology at ontario public health laboratories".ti. and "2009".yr. (0)

560	"Neuroborreliosis and tick-borne encephalitis - same vector, but different clinical course".ti. and "2007".yr. (0)

561	"Usefulness of c6 peptide antibody assay in early localised lyme borreliosis".ti. and "2007".yr. (0)

562	"The level of morbidity of lyme borreliosis after antibiotic preventive treatment".ti. and "2007".yr. (0)

563	"Seroprevalence of lyme disease and associated risk factors in rural population of beijing".ti. and "2015".yr. (1)

564	"Diagnostic use of the lymphocyte transformation test-memory lymphocyte immunostimulation assay in confirming active lyme borreliosis in clinically and serologically ambiguous cases".ti. and "2014".yr. (1)

565	"Test of 259 serums from patients with arthritis or neurological symptoms confirmed existence of lyme disease in hainan province, china".ti. and "2015".yr. (1)

566	"Examination of specific dna by pcr in patients with different forms of lyme borreliosis".ti. and "2008".yr. (1)

567	"Antibody reactivity to borrelia burgdorferi sensu stricto antigens in patients from the brazilian amazon region with skin diseases not related to lyme disease".ti. and "2010".yr. (1)

568	"Presence of borrelia burgdorferi sensu lato in patients with morphea from the amazonic region in brazil".ti. and "2011".yr. (1)

569	"Relating land cover and spatial distribution of nephropathia epidemica and lyme borreliosis in belgium".ti. and "2013".yr. (1)

570	"The paradox of the binomial ixodes ricinus activity and the observed unimodal lyme borreliosis season in hungary".ti. and "2014".yr. (1)

571	"A seventeen-year epidemiological surveillance study of borrelia burgdorferi infections in two provinces of northern spain".ti. and "2014".yr. (1)

572	"Gis and remote sensing use in the exploration of lyme disease epidemiology".ti. and "2015".yr. (1)

573	"Towards landscape design guidelines for reducing lyme disease risk".ti. and "2006".yr. (1)

574	"Relevance of chronic lyme disease to family medicine as a complex multidimensional chronic disease construct: A systematic review".ti. and "2014".yr. (1)

575	"Emerging incidence of lyme borreliosis, babesiosis, bartonellosis, and granulocytic ehrlichiosis in australia".ti. and "2011".yr. (1)

576	"Clinical determinants of lyme borreliosis, babesiosis, bartonellosis, anaplasmosis, and ehrlichiosis in an australian cohort".ti. and "2015".yr. (1)

577	"Benefit of intravenous antibiotic therapy in patients referred for treatment of neurologic lyme disease".ti. and "2011".yr. (1)

578	"Determinants of the geographic distribution of puumala virus and lyme borreliosis infections in belgium".ti. and "2007".yr. (1)

579	"Enzyme-linked immunosorbent assay, immunofluorescent assay, and recombinant immunoblotting in the serodiagnosis of early lyme borreliosis".ti. and "2003".yr. (1)

580	"Multicentric study of seroprevalence of borrelia burgdorferi and anaplasma phagocytophila in high-risk groups in regions of central and southern italy".ti. and "2004".yr. (1)

581	"Prevalence of igg antibodies against borrelia burgdorferi s.L. And ehrlichia phagocytophila in sera of patients presenting symptoms of lyme disease in a central region of italy".ti. and "2002".yr. (1)

582	"Detection of different borrelia burgdorferi genospecies in serum of people with different occupational risks: Short report".ti. and "2009".yr. (1)

583	"Diagnostic evaluation of human babesiosis in montenegro".ti. and "2012".yr. (0)

584	"Development of a foundation for a case definition of post-treatment lyme disease syndrome".ti. and "2013".yr. (1)

585	"Ultrasonographic evaluation of knee joints in patients with lyme disease".ti. and "2012".yr. (1)

586	"Serological diagnosis of lyme disease in valencia (spain)".ti. and "2010".yr. (0)

587	"Low risk of developing borrelia burgdorferi infection in the south-east of sweden after being bitten by a borrelia burgdorferi-infected tick".ti. and "2011".yr. (1)

588	"Epidemiological, clinical and zoonotic evidences for the existence of lyme disease in central of mexico".ti. and "2010".yr. (0)

589	"Maternal lyme borreliosis and pregnancy outcome".ti. and "2010".yr. (1)

590	"Clinical aspects of neuroborreliosis in bydgoszcz, poland".ti. and "2010".yr. (0)

591	"Antibodies to decorin-binding protein b (dbpb) in the diagnosis of lyme neuroborreliosis in children".ti. and "2014".yr. (1)

592	"Study of lyme borreliosis in ukraine".ti. and "2008".yr. (0)

593	"Comparison of immunofluorescence assay (ifa) and enzyme immuno assay (eia) for borrelial antibody detection in patients with signs and symptoms suggesting neuroborreliosis".ti. and "2006".yr. (0)

594	"The influence of meteorological conditions of the preceding winter on the incidences of tick-borne encephalitis and lyme borreliosis in the czech republic".ti. and "2008".yr. (0)

595	"Diversity of borrelia burgdorferi sensu lato in natural foci of novosibirsk region".ti. and "2008".yr. (0)

596	"A new recombinant igg and igm line immuno blot improves serodiagnosis of lyme borreliosis".ti. and "2004".yr. (0)

597	"Improvement of lyme borreliosis serodiagnosis by a new recombinant igg and igm line immunoblot".ti. and "2006".yr. (0)

598	"Comparison of a new line assay using purified and recombinant antigens with a european lysate blot for serodiagnosis of lyme borreliosis".ti. and "2006".yr. (0)

599	"Serological survey of bartonella spp., borrelia burgdorferi, brucella spp., coxiella burnetii, francisella tularensis, leptospira spp., echinococcus, hanta-,tick-borne encephalitis (tbe)-and xenotropic murine leukemia virus-related virus (xmrv) infection in employees of two forestry enterprises in north rhine-westphalia (nrw), germany, 2011-2013".ti. and "2015".yr. (0)

600	"Lydi-sentinel - a surveillance network on the incidence of lyme disease in bavaria".ti. and "2013".yr. (0)

601	"Serologic evidence of borrelia burgdorferi and anaplasma phagocytophilum infection among patients hospitalized with tick-borne encephalitis (tbe)".ti. and "2006".yr. (0)

602	"Lyme borreliosis: Prevalence of antibodies against non-proteinic (lipid) antigens".ti. and "2008".yr. (0)

603	"Quality of lyme disease serology. Insights and trends from the german proficiency testing program 2000-2005".ti. and "2006".yr. (0)

604	"Clinical assessment of patients with suspected lyme borreliosis".ti. and "2008".yr. (0)

605	"Evaluation of a new lyme borreliosis immunoblot including ebv and syphilis antigens as differential diagnosis tools".ti. and "2009".yr. (0)

606	"Pcr-detection and molecular-genetic analysis of tick-transmitted pathogens in patients of novosibirsk region, russia".ti. and "2008".yr. (0)

607	"The usefulness of 'in vivo' antigens in the diagnosis of human lyme borreliosis".ti. and "2008".yr. (0)

608	"Laboratory diagnosis of early lyme borreliosis - comparison of elisa, western blot (ecoline), and pcr results".ti. and "2006".yr. (0)

609	"Improved culture conditions for the growth and detection of borrelia from human serum".ti. and "2013".yr. (1)

610	"Detection of borreliae in archived sera from patients with clinically suspect lyme disease".ti. and "2014".yr. (1)

611	"A novel isothermal assay of borrelia burgdorferi by recombinase polymerase amplification with lateral flow detection".ti. and "2016".yr. (1)

612	"Psychiatric symptoms in the course of lyme borreliosis and tick borne encephalitis".ti. and "2004".yr. (0)

613	"Knowledge on lyme disease among foresters".ti. and "2011".yr. (1)

614	"Neurological complications of vaccination with outer surface protein a (ospa)".ti. and "2011".yr. (1)

615	"Urinary bladder detrusor dysfunction symptoms in lyme disease".ti. and "2013".yr. (1)

616	"The association of lyme disease with loss of sexual libido and the role of urinary bladder detrusor dysfunction".ti. and "2014".yr. (1)

617	"Absence of borrelia burgdorferi antibodies in the sera of venezuelan patients with localized scleroderma (morphea)".ti. and "2006".yr. (1)

618	"The clinical spectrum of lyme neuroborreliosis".ti. and "2010".yr. (1)

619	"Interstitial granulomatous dermatitis with histiocytic pseudorosettes: A new histopathologic pattern in cutaneous borreliosis. Detection of borrelia burgdorferi dna sequences by a highly sensitive pcr-elisa".ti. and "2003".yr. (1)

620	"Lyme disease knowledge, beliefs, and practices of new hampshire primary care physicians".ti. and "2002".yr. (1)

621	("Lyme arthritis in children presenting with joint effusions".ti. and "2011".yr.) not comment.pt. (1)

622	"Seroprevalence of antibodies that react with anaplasma phagocytophila, the agent of human granulocytic ehrlichiosis, in different populations in westchester county, new york".ti. and "2002".yr. (1)

623	"Importance of sample preparation for molecular diagnosis of lyme borreliosis from urine".ti. and "2002".yr. (1)

624	"Two-year evaluation of borrelia burgdorferi culture and supplemental tests for definitive diagnosis of lyme disease".ti. and "2005".yr. (1)

625	"Recombinant assay for serodiagnosis of lyme disease regardless of ospa vaccination status".ti. and "2002".yr. (1)

626	"Recombinant bbk32 protein in serodiagnosis of early and late lyme borreliosis".ti. and "2002".yr. (1)

627	"Comparison of the borrelia dotblot g, mardx, and vidas enzyme immunoassays for detecting immunoglobulin g antibodies to borrelia burgdorferi in human serum".ti. and "2002".yr. (1)

628	"Assessment of new culture method for detection of borrelia species from serum of lyme disease patients".ti. and "2014".yr. (1)

629	"Quantitative detection of borrelia burgdorferi in 2-millimeter skin samples of erythema migrans lesions: Correlation of results with clinical and laboratory findings".ti. and "2002".yr. (1)

630	"Evaluation of the c6 peptide enzyme-linked immunosorbent assay for individuals vaccinated with the recombinant ospa vaccine".ti. and "2002".yr. (1)

631	"Significant improvement of the recombinant borrelia-specific immunoglobulin g immunoblot test by addition of vlse and a dbpa homologue derived from borrelia garinii for diagnosis of early neuroborreliosis".ti. and "2003".yr. (1)

632	"Varicella zoster and borrelia burgdorferi are the main agents associated with facial paresis, especially in children".ti. and "2003".yr. (1)

633	"Patterns of lyme disease diagnosis and treatment by family physicians in a southeastern state".ti. and "2002".yr. (1)

634	"Role of experience and context in learning to diagnose lyme disease".ti. and "2002".yr. (1)

635	"Specific cutaneous infiltrates of b-cell chronic lymphocytic leukemia (b-cll) at sites typical for borrelia burgdorferi infection".ti. and "2002".yr. (1)

636	"Serodiagnosis of lyme disease by kinetic enzyme-linked immunosorbent assay using recombinant vlse1 or peptide antigens of borrelia burgdorferi compared with 2-tiered testing using whole-cell lysates".ti. and "2003".yr. (1)

637	"Persistence of the antibody response to the vlse sixth invariant region (ir6) peptide of borrelia burgdorferi after successful antibiotic treatment of lyme disease".ti. and "2003".yr. (1)

638	"Cloning of the gene encoding the decorin-binding protein b (dbpb) in borrelia burgdorferi sensu lato and characterisation of the antibody responses to dbpb in lyme borreliosis".ti. and "2002".yr. (1)

639	"Comparative reactivity of human sera to recombinant vlse and other borrelia burgdorferi antigens in class-specific enzyme-linked immunosorbent assays for lyme borreliosis".ti. and "2002".yr. (1)

640	"The chemokine cxcl13 in acute neuroborreliosis".ti. and "2010".yr. (1)

641	"Regional cerebral blood flow and cognitive deficits in chronic lyme disease".ti. and "2003".yr. (1)

642	"Lyme arthritis presenting as acute septic arthritis in children".ti. and "2003".yr. (1)

643	"Infections preceding early arthritis in southern sweden: A prospective population-based study".ti. and "2003".yr. (1)

644	"Serologic markers of lyme disease in children with autism".ti. and "2013".yr. (1)

645	"Epidemiological features and clinical manifestations of lyme borreliosis in korea during the period 2005-2012".ti. and "2015".yr. (1)

646	"Using risk group profiles as a lightweight qualitative approach for intervention development: An example of prevention of tick bites and lyme disease".ti. and "2013".yr. (1)

647	"Diagnostic usefulness of routine lyme serology in patients with early inflammatory arthritis in nonendemic areas".ti. and "2003".yr. (1)

648	"Good outcomes of lyme arthritis in 24 patients in an endemic area of switzerland".ti. and "2004".yr. (1)

649	"Igm deficiency: A retrospective chart review of clinical and immunologic features".ti. and "2012".yr. (0)

650	"Lyme neuroborreliosis and dementia".ti. and "2014".yr. (1)

651	("Efficacy of antibiotic prophylaxis for the prevention of lyme disease: An updated systematic review and meta-analysis".ti. and "2010".yr.) not comment.pt. (1)

652	"Beekeepers in central europe are at high risk for contracting lyme borreliosis".ti. and "2012".yr. (0)

653	"Potential uses of modafinil in psychiatric disorders".ti. and "2004".yr. (0)

654	("Predictive factors for differentiating between septic arthritis and lyme disease of the knee in children".ti. and "2016".yr.) not comment.pt. (1)

655	"Lyme arthritis in children presenting with joint effusions.[erratum appears in j bone joint surg am. 2011 feb;93(3):E11]".ti. and "2011".yr. (0)

656	"Lyme arthritis of the pediatric lower extremity in the setting of polyarticular disease".ti. and "2014".yr. (1)

657	"Information value of detection of borrelia antibodies in the healthy blood donors and in a population at risk".ti. and "2007".yr. (0)

658	"Chronic fatigue syndrome patients subsequently diagnosed with lyme disease borrelia burgdorferi: Evidence for mycoplasma species coinfections".ti. and "2008".yr. (0)

659	"Evaluation of two commercial systems for automated processing, reading, and interpretation of lyme borreliosis western blots".ti. and "2008".yr. (1)

660	"Validation of cultivation and pcr methods for diagnosis of lyme neuroborreliosis".ti. and "2008".yr. (1)

661	"Lyme disease in oregon".ti. and "2008".yr. (1)

662	"Improvement of lyme borreliosis serodiagnosis by a newly developed recombinant immunoglobulin g (igg) and igm line immunoblot assay and addition of vlse and dbpa homologues".ti. and "2005".yr. (1)

663	"Development of a multiantigen panel for improved detection of borrelia burgdorferi infection in early lyme disease".ti. and "2015".yr. (1)

664	"Collection and characterization of samples for establishment of a serum repository for lyme disease diagnostic test development and evaluation".ti. and "2014".yr. (1)

665	"Discriminating lyme neuroborreliosis from other neuroinflammatory diseases by levels of cxcl13 in cerebrospinal fluid".ti. and "2011".yr. (1)

666	"Differences and similarities between culture-confirmed human granulocytic anaplasmosis and early lyme disease".ti. and "2013".yr. (1)

667	"Comparative cost-effectiveness of two-tiered testing strategies for serodiagnosis of lyme disease with noncutaneous manifestations".ti. and "2013".yr. (1)

668	"Motion-onset and pattern-reversal visual evoked potentials in diagnostics of neuroborreliosis".ti. and "2006".yr. (1)

669	"Pattern and motion-related visual-evoked potentials in neuroborreliosis: Follow-up study".ti. and "2012".yr. (1)

670	"More specific bands in the igg western blot in sera from scottish patients with suspected lyme borreliosis".ti. and "2010".yr. (1)

671	"Development of real time pcr to detect toxoplasma gondii and borrelia burgdorferi infections in postal samples".ti. and "2008".yr. (1)

672	"Local borrelia burgdorferi sensu stricto and borrelia afzelii strains in a single mixed antigen improves western blot sensitivity".ti. and "2009".yr. (1)

673	"Provider knowledge, attitudes, and practices regarding lyme disease in arkansas".ti. and "2015".yr. (1)

674	"Provider knowledge, attitudes, and practices regarding lyme disease in arkansas".ti. and "2015".yr. (1)

675	"Borrelia burgdorferi-associated lymphocytoma cutis: Clinicopathologic, immunophenotypic, and molecular study of 106 cases".ti. and "2004".yr. (1)

676	"Granuloma annulare--a manifestation of infection with borrelia".ti. and "2008".yr. (1)

677	"Borrelia in granuloma annulare, morphea and lichen sclerosus: A pcr-based study and review of the literature".ti. and "2010".yr. (1)

678	"Enteroviral testing and length of hospital stay for children evaluated for lyme meningitis".ti. and "2013".yr. (1)

679	"Organisational intentions and responses: Presenting the risk of lyme disease to countryside users".ti. and "2013".yr. (0)

680	"Epidemics of vector-borne diseases observed in infectious disease surveillance in japan, 2000-2005".ti. and "2007".yr. (1)

681	"Level of awareness regarding some zoonotic diseases, among dog owners of ithaca, new york".ti. and "2014".yr. (1)

682	"Diagnostic novelties of chronic lyme/ neuroborreliosis".ti. and "2012".yr. (0)

683	"Behavioral and attitudes survey about lyme disease among a brazilian population in the endemic area of martha's vineyard, massachusetts".ti. and "2010".yr. (1)

684	"Diagnostic performance of cerebrospinal fluid chemokine cxcl13 and antibodies to the c6-peptide in lyme neuroborreliosis".ti. and "2011".yr. (1)

685	"Susceptibility, likelihood to be diagnosed, worry and fear for contracting lyme disease".ti. and "2017".yr. (1)

686	"Safety, immunogenicity, and efficacy of borrelia burgdorferi outer surface protein a (ospa) vaccine: A meta-analysis".ti. and "2017".yr. (1)

687	"Effectiveness of residential acaricides to prevent lyme and other tick-borne diseases in humans".ti. and "2016".yr. (1)

688	"Borrelia-specific interferon-gamma and interleukin-4 secretion in cerebrospinal fluid and blood during lyme borreliosis in humans: Association with clinical outcome".ti. and "2004".yr. (1)

689	"Primary cutaneous marginal zone lymphoma: Studies on borrelia infection".ti. and "2010".yr. (0)

690	"The epidemiology of lyme disease in the state of massachusetts, 2001-2011".ti. and "2014".yr. (0)

691	"Possible involvement of borrelia burgdorferi and hcv in the aethiology of mycosis fungoides".ti. and "2006".yr. (0)

692	"Epidemiology of tick bites and borreliosis in children attending kindergarten or so-called forest kindergarten in southwest germany".ti. and "2006".yr. (1)

693	"Isolation and detection of borrelia burgdorferi from human vaginal and seminal secretions".ti. and "2014".yr. (0)

694	"Morgellons disease: Analysis of a population with clinically confirmed microscopic subcutaneous fibers of unknown etiology".ti. and "2010".yr. (1)

695	"Safety of intravenous antibiotic therapy in chronic lyme disease".ti. and "2008".yr. (0)

696	"Clinical and seroepidemiologic characteristics of babesia wa-1 coinfection in patients with lyme disease in california".ti. and "2003".yr. (0)

697	"Clinical and immunologic evidence for transmission of lyme disease through intimate human contact".ti. and "2004".yr. (0)

698	"Lyme neuroborreliosis in hiv-1 positive men successfully treated with oral doxycycline: A case series and literature review".ti. and "2011".yr. (1)

699	"Spatiotemporal bayesian analysis of lyme disease in new york state, 1990-2000".ti. and "2006".yr. (1)

700	"Epidemic and spatial dynamics of lyme disease in new york state, 1990-2000".ti. and "2005".yr. (1)

701	"A prospective cross-over field trial shows protection of lemon eucalyptus extract against tick bites".ti. and "2004".yr. (1)

702	"Effects of reduced deer density on the abundance of ixodes scapularis (acari: Ixodidae) and lyme disease incidence in a northern new jersey endemic area".ti. and "2007".yr. (1)

703	"The relationship between deer density, tick abundance, and human cases of lyme disease in a residential community".ti. and "2014".yr. (1)

704	"Human behaviors elevating exposure to ixodes pacificus (acari: Ixodidae) nymphs and their associated bacterial zoonotic agents in a hardwood forest".ti. and "2004".yr. (1)

705	"The effects of bird feeders on lyme disease prevalence and density of ixodes scapularis (acari: Ixodidae) in a residential area of dutchess county, new york".ti. and "2003".yr. (1)

706	"Geographical and genospecies distribution of borrelia burgdorferi sensu lato dna detected in humans in the usa".ti. and "2014".yr. (1)

707	"Diagnostic accuracy and comparison of two assays for borrelia-specific igg and igm antibodies: Proposals for statistical evaluation methods, cut-off values and standardization".ti. and "2013".yr. (1)

708	"Audit of the laboratory diagnosis of lyme disease in scotland".ti. and "2005".yr. (1)

709	"Comparative evaluation of three different elisa methods for the diagnosis of early culture-confirmed lyme disease in italy".ti. and "2005".yr. (1)

710	"The use of local isolates in western blots improves serological diagnosis of lyme disease in scotland".ti. and "2007".yr. (1)

711	"Performance of a multiplexed serological microarray for the detection of antibodies against central nervous system pathogens".ti. and "2014".yr. (1)

712	"Community partnership designed to promote lyme disease prevention and engagement in citizen science".ti. and "2016".yr. (1)

713	"Cellular immune reactivity to recombinant borrellia-specific antigens for improved laboratory diagnosis of lyme borreliosis".ti. and "2004".yr. (0)

714	"Differentiating lyme arthritis from septic arthritis: Clues uncovered".ti. and "2011".yr. (0)

715	"Reactive lyme serology in optic neuritis".ti. and "2005".yr. (1)

716	"Cxcl13 and neopterin concentrations in cerebrospinal fluid of patients with lyme neuroborreliosis and other diseases that cause neuroinflammation".ti. and "2014".yr. (1)

717	"Neuroborreliosis-associated cerebral vasculitis: Long-term outcome and health-related quality of life".ti. and "2013".yr. (1)

718	"Quality of life, fatigue, depression and cognitive impairment in lyme neuroborreliosis".ti. and "2015".yr. (1)

719	"Prevalence and spectrum of residual symptoms in lyme neuroborreliosis after pharmacological treatment: A systematic review".ti. and "2016".yr. (1)

720	"Cerebrospinal fluid findings in adults with acute neuroborreliosis".ti. and "2011".yr. (0)

721	"Prevalence of taste disorders in idiopathic and b. Burgdorferi-associated facial palsy".ti. and "2009".yr. (1)

722	"lymphocyte chemoattractant (cxcl13) in the early diagnosis of acute lyme neuroborreliosis".ti. and "2008".yr. and published erratum.pt. (1)

723	"Acute peripheral facial palsy in adults".ti. and "2005".yr. (1)

724	"Double infection with encephalitis and lyme borreliosis transmitted by tick bite".ti. and "2005".yr. (0)

725	"Diagnosis of lyme neuroborreliosis with antibodies to recombinant proteins dbpa, bbk32, and ospc, and vlse ir6 peptide".ti. and "2003".yr. (1)

726	"Lyme disease-related intracranial hypertension in children: Clinical and imaging findings".ti. and "2016".yr. (1)

727	"The chemokine cxcl13 as a diagnostic and therapy response marker in acute neuroborreliosis".ti. and "2010".yr. (0)

728	"Neuroborreliosis in the south west of england".ti. and "2006".yr. (0)

729	"Clinical features of double infection with tick-borne encephalitis and lyme borreliosis transmitted by tick bite".ti. and "2006".yr. (1)

730	"Neuroborelliosis in south west england".ti. and "2014".yr. (0)

731	"Pediatric lyme arthritis of the hip: The great imitator?".ti. and "2015".yr. (0)

732	"Primary lyme arthritis of the pediatric hip".ti. and "2011".yr. (1)

733	"Evaluating the child with acute hip pain (irritable hip ) in a lyme endemic region".ti. and "2015".yr. (1)

734	"Reportable bacterial infections among new york city-born infants, 2001-2009".ti. and "2016".yr. (1)

735	"Chronic lyme disease: A survey of connecticut primary care physicians".ti. and "2010".yr. (1)

736	"False positive lyme disease igm immunoblots in children".ti. and "2016".yr. (1)

737	"Transient prolonged corrected qt interval in lyme disease".ti. and "2006".yr. (1)

738	"New insights into stages of lyme disease symptoms from a novel hospital-based registry".ti. and "2014".yr. (1)

739	"An estimate of lyme borreliosis incidence in western europe+".ti. and "2016".yr. (0)

740	"Preventive behaviors and knowledge of tick-borne illnesses: Results of a survey from an endemic area".ti. and "2015".yr. (1)

741	"Outcomes of children treated for lyme arthritis: Results of a large pediatric cohort".ti. and "2010".yr. (1)

742	"Borrelia burgdorferi persists in the gastrointestinal tract of children and adolescents with lyme disease".ti. and "2002".yr. (0)

743	"Lyme vaccination safety".ti. and "2002".yr. (0)

744	"Cerebrovascular events in lyme neuroborreliosis".ti. and "2015".yr. (1)

745	"Cutaneous borreliosis associated with t cell-predominant infiltrates: A diagnostic challenge".ti. and "2015".yr. (1)

746	"Factors influencing the utilization of lyme disease-prevention behaviors in a high-risk population".ti. and "2004".yr. (1)

747	"Using administrative medical claims data to supplement state disease registry systems for reporting zoonotic infections".ti. and "2013".yr. (1)

748	"Pharmacist initiation of postexposure doxycycline for lyme disease prophylaxis".ti. and "2014".yr. (1)

749	"Wais-iii and wms-iii performance in chronic lyme disease".ti. and "2006".yr. (1)

750	"Acute neuroborreliosis with involvement of the central nervous system".ti. and "2010".yr. (1)

751	"Long-term cognitive deficits after treated lyme neuroborreliosis-neuropsychological profile and demographical characteristics".ti. and "2013".yr. (0)

752	"Facial palsy as an initial symptom of lyme neuroborreliosis in an austrian endemic area".ti. and "2013".yr. (0)

753	"Clinical and epidemiological peculiarities of mixt infection (mi) of lyme disease (ld) and tick-borne encephalitis (tbe)".ti. and "2005".yr. (0)

754	"Laboratory diagnosis of lyme neuroborreliosis is influenced by the test used: Comparison of two elisas, immunoblot and cxcl13 testing".ti. and "2014".yr. (1)

755	"Electrocardiograph abnormalities in children with lyme meningitis".ti. and "2012".yr. (1)

756	"Neuropathy and cognitive impairment following vaccination with the ospa protein of borrelia burgdorferi".ti. and "2004".yr. (1)

757	"Diagnostic value of sural nerve biopsy in patients with suspected borrelia neuropathy".ti. and "2008".yr. (1)

758	"Distribution and presentation of lyme borreliosis in scotland - analysis of data from a national testing laboratory".ti. and "2015".yr. (1)

759	"Lyme borreliosis in scotland during two peak periods".ti. and "2009".yr. (0)

760	"Is tayside becoming a scottish hotspot for lyme borreliosis?".ti. and "2011".yr. (1)

761	"Application of nanotrap technology for high sensitivity measurement of urinary outer surface protein a carboxyl-terminus domain in early stage lyme borreliosis".ti. and "2015".yr. (1)

762	"Lyme risk for immigrants to the united states: The role of an educational tool".ti. and "2005".yr. (1)

763	"Imported disease in emergency departments: An undiscovered country".ti. and "2006".yr. (1)

764	"Seroprevalence of west nile virus, crimean-congo hemorrhagic fever virus, francisella tularensis and borrelia burgdorferi in rural population of manisa, western turkey".ti. and "2016".yr. (1)

765	"Implications of gender in chronic lyme disease".ti. and "2009".yr. (1)

766	"The clinical and laboratory characteristics of early lyme disease differ between men and women".ti. and "2010".yr. (0)

767	"Lyme neuroborreliosis: Aetiology and diagnosis of facial palsy in children from tyrol".ti. and "2010".yr. (1)

768	"Identification of a novel pathogenic borrelia species causing lyme borreliosis with unusually high spirochaetaemia: A descriptive study".ti. and "2016".yr. (1)

769	"Safety and immunogenicity of a novel multivalent ospa vaccine against lyme borreliosis in healthy adults: A double-blind, randomised, dose-escalation phase 1/2 trial".ti. and "2013".yr. (1)

770	"Oral doxycycline versus intravenous ceftriaxone for european lyme neuroborreliosis: A multicentre, non-inferiority, double-blind, randomised trial".ti. and "2008".yr. (1)

771	"Etiology, diagnosis, and management of facial palsy: 2000 patients at a facial nerve center".ti. and "2014".yr. (1)

772	"Sudden deafness and lyme disease".ti. and "2003".yr. (1)

773	"Clinicopathological features of primary cutaneous b-cell lymphomas from an academic regional hospital in central italy: No evidence of borrelia burgdorferi association".ti. and "2007".yr. (1)

774	"Demonstration of b. Burgdorferi-dna in two cases of nodal lymphoma".ti. and "2004".yr. (1)

775	"Macrolide therapy of chronic lyme disease".ti. and "2003".yr. (1)

776	"Comparative study of early lyme disease: Erythema migrans in new york state and northeastern poland".ti. and "2002".yr. (1)

777	"Efficacy of a long-term antibiotic treatment in patients with a chronic tick associated poly-organic syndrome (tapos)".ti. and "2009".yr. (1)

778	"Feedback on difficulties raised by the interpretation of serological tests for the diagnosis of lyme disease".ti. and "2014".yr. (1)

779	"Impact of serodiagnosis on the management of lyme borreliosis at angers university hospital".ti. and "2014".yr. (1)

780	"The potential use of cholestyramine to reduce the risk of developing clostridium difficile-associated diarrhoea in patients receiving long-term intravenous ceftriaxone".ti. and "2015".yr. (1)

781	"Seroprevalence of borrelia burgdorferi in north eastern india".ti. and "2008".yr. (1)

782	"Presence of borrelia burgdorferi in endomyocardial biopsies in patients with new-onset unexplained dilated cardiomyopathy".ti. and "2010".yr. (1)

783	"Intrathecally produced igg and igm antibodies to recombinant vlse, vlse peptide, recombinant ospc and whole cell extracts in the diagnosis of lyme neuroborreliosis".ti. and "2014".yr. (1)

784	"Utility of serodiagnostics designed for use in the united states for detection of lyme borreliosis acquired in europe and vice versa".ti. and "2014".yr. (1)

785	"An ospa-polymerase chain reaction/restriction fragment length polymorphism-based method for sensitive detection and reliable differentiation of all european borrelia burgdorferi sensu lato species and ospa types".ti. and "2004".yr. (1)

786	"Diagnosis of lyme borreliosis using enzyme immunoanalysis".ti. and "2005".yr. (1)

787	"Bacterial tick-borne diseases caused by bartonella spp., borrelia burgdorferi sensu lato, coxiella burnetii, and rickettsia spp. Among patients with cataract surgery".ti. and "2014".yr. (1)

788	"The reportable events monthly report (remr)".ti. and "2013".yr. (1)

789	"The disease among u.S. Military members, active and reserve component, 2001-2008".ti. and "2009".yr. (0)

790	"[incidence of lyme disease in spain]".ti. and "2016".yr. (1)

791	"Estimating the incidence of new onset lyme disease in rhode island".ti. and "2008".yr. (1)

792	"Promoting specialty care for lyme disease: Lessons learned".ti. and "2006".yr. (1)

793	"Supervised resistance exercise for patients with persistent symptoms of lyme disease".ti. and "2015".yr. (1)

794	"Occupational diseases among farmers in poland".ti. and "2016".yr. (1)

795	"Outer surface protein e antibody response and its effect on complement factor h binding to ospe in lyme borreliosis".ti. and "2008".yr. (1)

796	"Epidemiologic and ecologic characteristics of some diseases transmitted by arthropods on the littoral of the republic of croatia".ti. and "2002".yr. (1)

797	"Lyme disease reporting for navy and marine corps (1997-2000)".ti. and "2003".yr. (1)

798	"Epidemiological characteristics and clinical manifestations of lyme borreliosis in croatia".ti. and "2006".yr. (1)

799	"Severity of lyme disease with persistent symptoms. Insights from a double-blind placebo-controlled clinical trial".ti. and "2008".yr. (1)

800	"Safety of intravenous antibiotic therapy in patients referred for treatment of neurologic lyme disease".ti. and "2010".yr. (1)

801	"Lyme disease--united states, 2001-2002".ti. and "2004".yr. (1)

802	"Effect of electronic laboratory reporting on the burden of lyme disease surveillance--new jersey, 2001-2006".ti. and "2008".yr. (1)

803	"Notes from the field: Update on lyme carditis, groups at high risk, and frequency of associated sudden cardiac death--united states".ti. and "2014".yr. (1)

804	"Lyme disease - united states, 2000".ti. and "2002".yr. and Marshall.ax. (0)

805	"Morphoea and borrelia burgdorferi: Results from the scottish highlands in the context of the world literature".ti. and "2002".yr. (1)

806	"Improvement in the laboratory recognition of lyme borreliosis with the combination of culture and pcr methods".ti. and "2003".yr. (1)

807	"Surveillance for lyme disease--united states, 1992-2006".ti. and "2008".yr. (1)

808	"Lyme disease - united states, 2003-2005".ti. and "2007".yr. (1)

809	"Lyme disease - united states, 2001-2002".ti. and "2004".yr. (1)

810	("Lyme disease - united states, 2000".ti. and "2002".yr.) not MMWR - Morbidity & Mortality Weekly Report.jn. (1)

811	"Reported vectorborne and zoonotic diseases, u.S. Air force, 2000-2011".ti. and "2012".yr. (1)

812	"Surveillance snapshot: Lyme disease among beneficiaries of the military health system, 2001-2012".ti. and "2013".yr. (1)

813	"The geographic distribution of incident lyme disease among active component service members stationed in the continental united states, 2004-2013".ti. and "2014".yr. (1)

814	"Characterizing the relationship between tick bites and lyme disease in active component u.S. Armed forces in the eastern united states".ti. and "2015".yr. (1)

815	"Lyme disease trends--dutchess county, new york, 1992-2000".ti. and "2003".yr. (1)

816	"Relapsing inflammatory optic neuropathy: Is it a new autoimmune disease?".ti. and "2011".yr. (0)

817	"Pilot study into borrelia seroprevelence in new brunswick multiple sclerosis patients".ti. and "2016".yr. (0)

818	"Differential aspects of primary progressive multiple sclerosis and neuroborreliosis".ti. and "2010".yr. (0)

819	"Differential diagnosis of multiple sclerosis and chronic progressive borrelial encephalomyelitis".ti. and "2011".yr. (0)

820	"Borrelia burgdorferi - infection agent - risk assessment in ms".ti. and "2014".yr. (0)

821	"Lyme disease serology in amyotrophic lateral sclerosis".ti. and "2009".yr. (1)

822	"Facial nerve palsy associated with lyme disease".ti. and "2003".yr. (1)

823	"Serological testing for borrelia: It's a mess".ti. and "2011".yr. (0)

824	"General practitioner reported incidence of lyme carditis in the netherlands".ti. and "2015".yr. (1)

825	"Pediatric lyme neuroborreliosis: Different clinical presentations of the same agent; single center experience".ti. and "2016".yr. (1)

826	"Clinical evaluation and oct optic disc and macular analysis in 6 cases of neuroretinitis".ti. and "2011".yr. (0)

827	"Incidence of neuroborreliosis-associated cerebral vasculitis in germany".ti. and "2013".yr. (0)

828	"Relevance of the antibody index to diagnose lyme neuroborreliosis among seropositive patients".ti. and "2007".yr. (1)

829	"Lyme neuroborreliosis and dementia".ti. and "2013".yr. (0)

830	"European neuroborreliosis: Neuropsychological findings 30 months post treatment".ti. and "2011".yr. (0)

831	"A randomized, placebo-controlled trial of repeated iv antibiotic therapy for lyme encephalopathy".ti. and "2008".yr. (1)

832	"Practice parameter: Treatment of nervous system lyme disease".ti. and "2007".yr. (1)

833	("Lyme disease and optic neuritis: Long-term follow-up of seropositive patients".ti. and "2003".yr.) not comment.pt. (1)

834	"Cognitive function in post-treatment lyme disease: Do additional antibiotics help?".ti. and "2003".yr. (1)

835	"Lyme disease and dementia".ti. and "2007".yr. (0)

836	("Study and treatment of post lyme disease" and "A randomized double masked clinical trial").ti. and "2003".yr. (1)

837	"Diagnostic accuracy of csfb-lymphocyte chemoattractant in lyme neuroborreliosis".ti. and "2007".yr. (0)

838	"A prospective study on the role of cxcl13 in lyme neuroborreliosis".ti. and "2011".yr. (1)

839	("Lyme disease and optic neuritis: Long-term follow-up of seropositive patients".ti. and "2003".yr.) not comment.pt. (1)

840	"Intrusive symptoms and infectious encephalopathies".ti. and "2016".yr. (0)

841	"Tick-borne infections and pediatric bipolar disorder".ti. and "2016".yr. (0)

842	"Diagnosis of neuroborreliosis - improved knowledge base for qualified antibody analysis and cerebrospinal fluid data pattern related interpretations".ti. and "2013".yr. (0)

843	"Treatment of north american lyme neuroborrelliosis with oral doxycycline and intravenous ceftriaxone: A comparative case series".ti. and "2015".yr. (0)

844	"Amyotrophic lateral sclerosis (als) with laboratory abnormalities of unknown significance (laus)-where does it begin and where does it end?".ti. and "2015".yr. (0)

845	"Amyotrophic lateral sclerosis and lyme borreliosis".ti. and "2006".yr. (0)

846	"Neuropsychological profile of children after an episode of neuroborreliosis".ti. and "2013".yr. (1)

847	"Lyme neuroborreliosis: Aetiology and diagnosis of facial palsy in children from tyrol".ti. and "2013".yr. (0)

848	"Mycoplasma pneumoniae and borrelia burgdorferi as cause for facial paralysis in children: Single-center experience of 10 years".ti. and "2013".yr. (0)

849	"Observation of children of mothers who underwent borreliosis in the course of pregnancy".ti. and "2009".yr. (0)

850	"Randomized trial of longer-term therapy for symptoms attributed to lyme disease".ti. and "2016".yr. (1)

851	"Antinuclear antibodies in patients with lyme disease".ti. and "2010".yr. (0)

852	"Presence and indigenous nature of lyme disease in southern italy".ti. and "2003".yr. (1)

853	"Molecular and microscopical evidence of ehrlichia spp. And borrelia burgdorferi sensu lato in patients, animals and ticks in the czech republic".ti. and "2002".yr. (1)

854	"Evaluation of the c6 enzyme-linked immunoadsorbent assay for the serodiagnosis of lyme borreliosis in north-eastern italy".ti. and "2006".yr. (1)

855	"Seroprevalence of borrelia burgdorferi infection among forestry workers and farmers in duzce, north-western turkey".ti. and "2008".yr. (1)

856	"Comparative evaluation of two enzyme linked immunosorbent assay methods and three western blot methods for the diagnosis of culture-confirmed early lyme borreliosis in italy".ti. and "2005".yr. (1)

857	"National surveillance 12-monthly data notification rate changes".ti. and "2009".yr. (0)

858	"No association of seropositivity for anti-borrelia igg antibody with mental and physical complaints".ti. and "2008".yr. (1)

859	"Cerebral metabolic changes associated with lyme disease".ti. and "2002".yr. (1)

860	"Difficulties of interpreting borrelia serology in patients with uveitis".ti. and "2011".yr. (1)

861	"Prevalence of borrelia burgdorferi infection in a series of 98 primary cutaneous lymphomas".ti. and "2011".yr. (1)

862	"Lyme disease in maine: A comparison of nedss surveillance data and maine health data organization hospital discharge data".ti. and "2014".yr. (1)

863	"Minocycline as a substitute for doxycycline in targeted scenarios: A systematic review".ti. and "2015".yr. (1)

864	"Geographic expansion of lyme disease in the southeastern united states, 2000-2014".ti. and "2015".yr. (1)

865	"The lymphocyte transformation test for borrelia detects active lyme borreliosis and verifies effective antibiotic treatment".ti. and "2012".yr. (1)

866	"Analysis of the influence of ascaris, borrelia, toxocara, toxoplasma and mycoplasma infections on the incidence and course of perthes disease - pilot study".ti. and "2013".yr. (1)

867	"Serological detection of lyme borreliosis agents in patients from korea, 2005-2009".ti. and "2011".yr. (1)

868	"Spirochetes in otology: Are we testing for the right pathogens?".ti. and "2008".yr. (1)

869	"No evidence for the diagnostic value of borrelia serology in patients with sudden hearing loss".ti. and "2012".yr. (1)

870	"Long-term outcome of facial palsy in neuroborreliosis".ti. and "2005".yr. (1)

871	"Microbiologic findings in acute facial palsy in children".ti. and "2013".yr. (1)

872	("Lyme disease associated with sudden sensorineural hearing loss: Case report and literature review".ti. and "2013".yr.) not letter.pt. (1)

873	"The vlse (ir6) peptide elisa in the serodiagnosis of lyme facial paralysis".ti. and "2004".yr. (1)

874	"Clinical characteristics, treatment and outcome of children with lyme arthritis in nova scotia".ti. and "2015".yr. (1)

875	"Surveillance for childhood lyme disease by the canadian pediatric surveillance program".ti. and "2015".yr. (0)

876	"Treatment of intractable headaches associated with lyme disease using neuromodulation techniques".ti. and "2010".yr. (0)

877	("Physician reported incidence of early and late lyme borreliosis".ti. and "2015".yr.) not published erratum.pt. (1)

878	"Sensitivity of a point of care tick-test for the development of lyme borreliosis".ti. and "2013".yr. (1)

879	"Diagnosis and management of human borreliosis in romania".ti. and "2014".yr. (0)

880	"Pilot study assessing the effectiveness of factory-treated, long-lasting permethrin-impregnated clothing for the prevention of tick bites during occupational tick exposure in highly infested military training areas, germany".ti. and "2015".yr. (1)

881	"Epidemiological situation of lyme borreliosis in germany: Surveillance data from six eastern german states, 2002 to 2006".ti. and "2008".yr. (1)

882	"The association between infectious burden and parkinson's disease: A case-control study".ti. and "2015".yr. (1)

883	"Concordance of four commercial enzyme immunoassay and three immunoblot formats for the detection of lyme borreliosis antibodies in human serum: The two-tier approach remains".ti. and "2016".yr. (1)

884	"Overdiagnosis and overtreatment of lyme disease in children".ti. and "2002".yr. (1)

885	"Synovial fluid findings in children with knee monoarthritis in lyme disease endemic areas".ti. and "2014".yr. (1)

886	"Causes of febrile illnesses after a tick bite in slovenian children".ti. and "2003".yr. (1)

887	"Lyme borreliosis and acute peripheral facial palsy in slovenian children".ti. and "2010".yr. (1)

888	"Borrelia burgdorferi sensu lato bacteremia in slovenian children with solitary and multiple erythema migrans".ti. and "2011".yr. (1)

889	"Borrelial lymphocytoma in children".ti. and "2015".yr. (1)

890	"Azithromycin is equally effective as amoxicillin in children with solitary erythema migrans".ti. and "2015".yr. (1)

891	"Diagnostic utility of borrelia burgdorferi cerebrospinal fluid polymerase chain reaction in children with lyme meningitis".ti. and "2005".yr. (1)

892	"One-year follow-up of tick-borne central nervous system infections in childhood".ti. and "2012".yr. (1)

893	"Prevalence of lyme meningitis in children with aseptic meningitis in a lyme disease-endemic region".ti. and "2011".yr. (1)

894	"New antigens for serologic diagnosis of neuroborreliosis in children".ti. and "2005".yr. (1)

895	"The positive predictive value of lyme elisa for the diagnosis of lyme disease in children".ti. and "2015".yr. (1)

896	"Early differentiation of lyme from enteroviral meningitis".ti. and "2005".yr. (1)

897	"Improved laboratory diagnostics of lyme neuroborreliosis in children by detection of antibodies to new antigens in cerebrospinal fluid".ti. and "2008".yr. (1)

898	"Lyme neuroborreliosis in children: A prospective study of clinical features, prognosis, and outcome".ti. and "2008".yr. (1)

899	"Treatment complications in children with lyme meningitis".ti. and "2012".yr. (1)

900	"Successful treatment of neuroborreliosis with ten day regimens".ti. and "2002".yr. (1)

901	"Prediction of lyme meningitis based on a logistic regression model using clinical and cerebrospinal fluid analysis: A european study".ti. and "2009".yr. (1)

902	"Etiology of aseptic meningitis, peripheral facial nerve palsy, and a combination of both in children".ti. and "2010".yr. (1)

903	"Accuracy of reporting of lyme disease in connecticut".ti. and "2003".yr. (0)

904	"Effectiveness of personal preventive measures for lyme disease".ti. and "2003".yr. (0)

905	"Lyme arthritis outcomes in children: A single center cohort study".ti. and "2012".yr. (0)

906	"Infectious agents in juvenile scleroderma".ti. and "2014".yr. (0)

907	"Prediction of lyme meningitis in children from a lyme disease-endemic region: A logistic-regression model using history, physical, and laboratory findings".ti. and "2006".yr. (1)

908	"Validation of a clinical prediction rule to distinguish lyme meningitis from aseptic meningitis".ti. and "2012".yr. (1)

909	"Lyme carditis in children: Presentation, predictive factors, and clinical course".ti. and "2009".yr. (1)

910	"Distinguishing lyme from septic knee monoarthritis in lyme disease-endemic areas".ti. and "2013".yr. (1)

911	"Comparative study of cefuroxime axetil versus amoxicillin in children with early lyme disease".ti. and "2002".yr. (1)

912	"Prospective validation of a clinical prediction model for lyme meningitis in children".ti. and "2009".yr. (1)

913	"Clinical predictors of lyme disease among children with a peripheral facial palsy at an emergency department in a lyme disease-endemic area".ti. and "2008".yr. (1)

914	"Long-term clinical outcome after lyme neuroborreliosis in childhood".ti. and "2012".yr. (1)

915	"Acute pediatric monoarticular arthritis: Distinguishing lyme arthritis from other etiologies".ti. and "2009".yr. (1)

916	"Long-term neuropsychologic and health outcomes of children with facial nerve palsy attributable to lyme disease".ti. and "2003".yr. (1)

917	"Severity of chronic lyme disease compared to other chronic conditions: A quality of life survey".ti. and "2014".yr. (1)

918	"Development of a public health assessment tool to prevent lyme disease: Tool construction and validation".ti. and "2005".yr. (1)

919	"Bilateral facial paralysis: A 13 year experience".ti. and "2016".yr. (1)

920	"Health care costs, utilization and patterns of care following lyme disease".ti. and "2015".yr. (1)

921	"Adaptation and evaluation of a multi-criteria decision analysis model for lyme disease prevention".ti. and "2015".yr. (1)

922	"Seropositivity of lyme borreliosis and associated risk factors: A population-based study in children and adolescents in germany (kiggs)".ti. and "2012".yr. (1)

923	"Direct molecular detection and genotyping of borrelia burgdorferi from whole blood of patients with early lyme disease".ti. and "2012".yr. (1)

924	"Integrated assessment of behavioral and environmental risk factors for lyme disease infection on block island, rhode island".ti. and "2014".yr. (1)

925	"A prospective study among patients presenting at the general practitioner with a tick bite or erythema migrans in the netherlands".ti. and "2013".yr. (1)

926	"Quantitative detection of borrelia burgdorferi sensu lato in erythema migrans skin lesions using internally controlled duplex real time pcr".ti. and "2013".yr. (1)

927	"C6 peptide-based multiplex phosphorescence analysis (phosphan) for serologic confirmation of lyme borreliosis".ti. and "2015".yr. (1)

928	"Are patients with erythema migrans who have leukopenia and/or thrombocytopenia coinfected with anaplasma phagocytophilum or tick-borne encephalitis virus?".ti. and "2014".yr. (1)

929	"Clinical characteristics associated with borrelia burgdorferi sensu lato skin culture results in patients with erythema migrans".ti. and "2013".yr. (1)

930	"Gender disparity between cutaneous and non-cutaneous manifestations of lyme borreliosis".ti. and "2013".yr. (1)

931	"Correlation of culture positivity, pcr positivity, and burden of borrelia burgdorferi sensu lato in skin samples of erythema migrans patients with clinical findings".ti. and "2015".yr. (1)

932	"The degree of damage in the peripheral facial nerve palsy in children depending on the cause: The role of lyme neuroborreliosis".ti. and "2014".yr. (0)

933	"Workplace health threats and their effects in podlasie region".ti. and "2008".yr. (0)

934	"Tick-borne infections as a cause of heart transplantation".ti. and "2011".yr. (1)

935	"Comparison of detection of borrelia burgdorferi dna and anti-borrelia burgdorferi antibodies in patients with erythema migrans in north-eastern poland".ti. and "2015".yr. (1)

936	"Simultaneous gastrointestinal infections in children and adolescents".ti. and "2004".yr. (0)

937	"Lyme borreliosis infection as a cause of dilated cardiomyopathy".ti. and "2006".yr. (1)

938	"Significance of borrelia infection in development of dilated cardiomypathy (a pilot study)".ti. and "2007".yr. (1)

939	"Lyme borreliosis--waiting for lyme carditis? A long-term prospective study".ti. and "2005".yr. (1)

940	"Prevention of lyme disease in dutch children: Analysis of determinants of tick inspection by parents".ti. and "2002".yr. (1)

941	"Pulse therapy with ceftriaxone on lyme neuroborreliosis".ti. and "2005".yr. (0)

942	"Lyme borreliosis in patients over 60-years-old".ti. and "2010".yr. (0)

943	"Human serum reactivity against borrelia ospc, ospa, flab and vlse protein antigens in early and disseminated lyme borreliosis".ti. and "2010".yr. (0)

944	"Tick-bite preventative behavior of people on recreational areas in warsaw [conference poster]".ti. and "2014".yr. (0)

945	"Deer, predators, and the emergence of lyme disease".ti. and "2012".yr. (1)

946	"Invasion of two tick-borne diseases across new england: Harnessing human surveillance data to capture underlying ecological invasion processes".ti. and "2016".yr. (1)

947	"Epidemiology of borreliosis in workers of the district forestry offices in lower silesia".ti. and "2007".yr. (1)

948	"Lyme disease with effusion either in hip or knee in children from podlaskie region treated in clinic in 2004-2010".ti. and "2014".yr. (1)

949	"Lyme disease in poland in 2013".ti. and "2015".yr. (1)

950	"Lyme disease in poland in 2011".ti. and "2013".yr. (1)

951	"Lyme disease in poland in 2012".ti. and "2014".yr. (1)

952	"Cns lyme disease manifestation in children".ti. and "2007".yr. (1)

953	"Correlates of perceived health-related quality of life in post-treatment lyme encephalopathy".ti. and "2013".yr. (1)

954	"Patients with organically unexplained symptoms presenting to a borreliosis clinic: Clinical and psychobehavioral characteristics and quality of life".ti. and "2013".yr. (1)

955	"Lyme disease and post-treatment lyme disease syndrome: The neglected disease in our own backyard".ti. and "2014".yr. (1)

956	"Lyme disease - what is the cost for scotland?".ti. and "2003".yr. (1)

957	"A qualitative approach to understanding patients' diagnosis of lyme disease".ti. and "2006".yr. (1)

958	"Evaluation of deer-targeted interventions on lyme disease incidence in connecticut".ti. and "2011".yr. (1)

959	"Lyme disease in a british referral clinic".ti. and "2012".yr. (1)

960	"Management of suspected lyme borreliosis: Experience from an outpatient parenteral antibiotic therapy service".ti. and "2013".yr. (1)

961	"Living in limbo: Contested narratives of patients with chronic symptoms following lyme disease".ti. and "2015".yr. (0)

962	"Post-treatment lyme disease syndrome symptomatology and the impact on life functioning: Is there something here?".ti. and "2013".yr. (1)

963	"Neuro-lyme disease: Mr imaging findings".ti. and "2009".yr. (1)

964	"Reduction in lyme disease risk as an economic benefit of habitat restoration".ti. and "2012".yr. (0)

965	"Prevalence of yersinia spp., chlamydia trachomatis, chlamydophila pneumoniae and borrelia burgdorferi antibodies in healthy blood donors' sera".ti. and "2013".yr. (0)

966	"Antibody profile to borrelia burgdorferi in veterinarians from nuevo leon, mexico, a non-endemic area of this zoonosis".ti. and "2016".yr. (1)

967	"Articular manifestations in patients with lyme disease".ti. and "2015".yr. (0)

968	"Do infections trigger juvenile idiopathic arthritis?".ti. and "2011".yr. (1)

969	"Evaluation of skin thickness lesions in patients with lyme disease measured by modified rodnan total skin score".ti. and "2012".yr. (1)

970	"Seroprevalence of borrelia burgdorferi in patients with behcet's disease".ti. and "2003".yr. (1)

971	"Rhode island tick-borne disease surveillance summary 2012-2013".ti. and "2014".yr. (1)

972	"Diagnostic impact of routine lyme serology in recent-onset arthritis: Results from the espoir cohort".ti. and "2016".yr. (1)

973	"Serological and clinical one year follow-up of patients with erythema migrans treated in a romanian infectious disease hospital".ti. and "2014".yr. (0)

974	"Serological, clinical and epidemiological aspects of lyme borreliosis in mures county, romania".ti. and "2014".yr. (0)

975	"Reinfection with lyme borreliosis: A retrospective follow-up study in southern sweden".ti. and "2002".yr. (1)

976	"Clinical outcome of erythema migrans after treatment with phenoxymethyl penicillin".ti. and "2003".yr. (1)

977	"5-y follow-up study of patients with neuroborreliosis".ti. and "2002".yr. (1)

978	"Human granulocytic ehrlichiosis as a common cause of tick-associated fever in southeast sweden: Report from a prospective clinical study".ti. and "2002".yr. (1)

979	"Characteristics of erythema migrans in borrelia afzelii and borrelia garinii infections".ti. and "2003".yr. (1)

980	"Three major lyme borrelia genospecies (borrelia burgdorferi sensu stricto, b. Afzelii and b. Garinii) identified by pcr in cerebrospinal fluid from patients with neuroborreliosis in sweden".ti. and "2002".yr. (1)

981	"Risk of acquiring tick bites in south-eastern sweden".ti. and "2002".yr. (1)

982	"Intravenous ceftriaxone compared with oral doxycycline for the treatment of lyme neuroborreliosis".ti. and "2005".yr. (1)

983	"Seroprevalence of rickettsia spp. Infection among tick-bitten patients and blood donors in sweden".ti. and "2008".yr. (1)

984	"Serological and molecular evidence of rickettsia helvetica in denmark".ti. and "2004".yr. (1)

985	"Clinical characteristics of childhood lyme neuroborreliosis in an endemic area of northern europe".ti. and "2009".yr. (1)

986	"Cerebrospinal fluid chemokine cxcl13 in the diagnosis of neuroborreliosis in children".ti. and "2013".yr. (1)

987	"Acute facial nerve palsy in children: How often is it lyme borreliosis?".ti. and "2007".yr. (1)

988	"Laboratory data in children with lyme neuroborreliosis, relation to clinical presentation and duration of symptoms".ti. and "2009".yr. (1)

989	"Lyme disease and schizophrenia: Case studies from an adjunctive minocycline study".ti. and "2013".yr. (0)

990	"Tick bites and erythema migrans in transylvania".ti. and "2013".yr. (0)

991	"Diagnosis and treatment in patients with chronic tick associated poly-organic syndrome (tapos) - a case series".ti. and "2010".yr. (0)

992	"Situating risk in the context of a woodland visit: A case study on lyme borreliosis".ti. and "2012".yr. (0)

993	"Urban and rural risks of lyme disease in the scottish highlands".ti. and "2009".yr. (1)

994	"Borreliosis - cause of obstructive sleep apnea syndrome. Comparison with normal osas".ti. and "2009".yr. (0)

995	"Socio-economic characteristics in notified erythema migrans patients".ti. and "2015".yr. (1)

996	"Borrelia burgdorferi (strain b. Afzelii) antibodies among malaysian blood donors and patients".ti. and "2002".yr. (1)

997	"Comparing spatio-temporal clusters of arthropod-borne infections using administrative medical claims and state reported surveillance data".ti. and "2012".yr. (1)

998	"Landscape risk factors for lyme disease in the eastern broadleaf forest province of the hudson river valley and the effect of explanatory data classification resolution".ti. and "2015".yr. (1)

999	"Tick related diseases in switzerland, 2008 to 2011".ti. and "2013".yr. (1)

1000	"Intentions to receive a potentially available lyme disease vaccine in an urban sample".ti. and "2016".yr. (1)

1001	"Clinical features of 705 borrelia burgdorferi seropositive patients in an endemic area of northern italy".ti. and "2014".yr. (1)

1002	"An mr brain images classifier system via particle swarm optimization and kernel support vector machine".ti. and "2013".yr. (1)

1003	"Putative association between anti-borrelia igg and autoimmune thyroid disease?".ti. and "2005".yr. (1)

1004	"Incidence and antibiotic treatment of erythema migrans in norway 2005-2009".ti. and "2016".yr. (0)

1005	"U.S. Healthcare providers' experience with lyme and other tick-borne diseases".ti. and "2014".yr. (1)

1006	"Tick-borne disease preventive practices and perceptions in an endemic area".ti. and "2016".yr. (1)

1007	"Human tick infestation pattern, tick-bite rate, and associated borrelia burgdorferi s.L. Infection risk during occupational tick exposure at the seedorf military training area, northwestern germany".ti. and "2014".yr. (1)

1008	"Epidemiology of lyme disease in low-incidence states".ti. and "2015".yr. (1)

1009	"Continuing increase of tick bites and lyme disease between 1994 and 2009".ti. and "2015".yr. (1)

1010	"U.S. Public's experience with ticks and tick-borne diseases: Results from national healthstyles surveys".ti. and "2015".yr. (1)

1011	"Prospective study on the incidence of infection by borrelia burgdorferi sensu lato after a tick bite in a highly endemic area of switzerland".ti. and "2011".yr. (1)

1012	"Functional outcomes in patients with borrelia burgdorferi reinfection".ti. and "2014".yr. (1)

1013	"Serological and molecular evidence for spotted fever group rickettsia and borrelia burgdorferi sensu lato co-infections in the netherlands".ti. and "2016".yr. (1)

1014	"Epidemiology and cost of hospital care for lyme borreliosis in germany: Lessons from a health care utilization database analysis".ti. and "2015".yr. (1)

1015	"Climate change influences on the annual onset of lyme disease in the united states".ti. and "2015".yr. (1)

1016	"Predictors of protective behaviour against ticks in the uk: A mixed methods study".ti. and "2014".yr. (1)

1017	"Human seroprevalence against borrelia burgdorferi sensu lato in two comparable regions of the eastern alps is not correlated to vector infection rates".ti. and "2015".yr. (1)

1018	"Geographical differences in seroprevalence of borrelia burgdorferi antibodies in norway, 2011-2013".ti. and "2016".yr. (1)

1019	"A prospective study on the incidence of borrelia burgdorferi sensu lato infection after a tick bite in sweden and on the aland islands, finland (2008-2009)".ti. and "2016".yr. (1)

1020	"Trends in surveillance data of human lyme borreliosis from six federal states in eastern germany, 2009-2012".ti. and "2014".yr. (1)

1021	"Spatial distribution of a population at risk: An important factor for understanding the recent rise in tick-borne diseases (lyme borreliosis and tick-borne encephalitis in the czech republic)".ti. and "2013".yr. (1)

1022	"Peri-urbanisation, counter-urbanisation, and an extension of residential exposure to ticks: A clue to the trends in lyme borreliosis incidence in the czech republic?".ti. and "2014".yr. (1)

1023	"Evidence of rapid changes in lyme disease awareness in canada".ti. and "2016".yr. (1)

1024	"Knowledge and prevention of tick-borne diseases vary across an urban-to-rural human land-use gradient".ti. and "2013".yr. (1)

1025	"Risk factors identified in blood donors screening positive or inconclusive for babesia microti".ti. and "2014".yr. (0)

1026	"Prevalence of lyme borreliosis in scottish blood donors".ti. and "2011".yr. (0)

1027	"Seroprevalence of antibodies to borrelia burgdorferi in a healthy swiss blood donor population".ti. and "2011".yr. (0)

1028	"Diagnosis of febrile illnesses in returned travelers using the pc software gideon".ti. and "2005".yr. (1)

1029	"An analysis of imported infections over a 5-year period at a teaching hospital in the united kingdom".ti. and "2003".yr. (1)

1030	"Incidence rates of selected infectious diseases in the most populated counties, united states, 2004".ti. and "2008".yr. (0)

1031	"Investigation of borrelia burgdorferi seroprevalence in van region of turkey".ti. and "2008".yr. (0)

1032	"Analysis of borrelia burgdorferi antibody seroprevalence in patients with behcet's disease using 3 different serological assays".ti. and "2004".yr. (0)

1033	"Evaluation of borrelia burgdorferi sensu lato seroprevalence in the province of bolu, turkey".ti. and "2016".yr. (0)

1034	"The seroprevalence of anaplasma phagocytophilum in humans from two different climatic regions of turkey and its co-seroprevalence rate with borrelia burgdorferi turkiyenin iki farkli".ti. and "2011".yr. (0)

1035	"Adverse event reports following vaccination for lyme disease: December 1998-july 2000".ti. and "2002".yr. (1)

1036	"Antibiotic treatment patterns associated with persons newly-diagnosed with lyme disease in the united states".ti. and "2014".yr. (0)

1037	"Assessing the incidence and treatment of post-treatment lyme disease syndrome in an administrative claims database".ti. and "2014".yr. (0)

1038	"The geographic correlation between lyme disease incidence and degenerative neurological disease mortality: An ecological study".ti. and "2013".yr. (0)

1039	"Assessing lyme disease prevalence in the united states medicaid population".ti. and "2014".yr. (0)

1040	"Effect of gender on clinical and epidemiologic features of lyme borreliosis".ti. and "2007".yr. (1)

1041	"Lyme borreliosis incidence in two french departments: Correlation with infection of ixodes ricinus ticks by borrelia burgdorferi sensu lato".ti. and "2007".yr. (1)

1042	"Knowledge, attitudes, and behaviors regarding lyme disease prevention among connecticut residents, 1999-2004".ti. and "2008".yr. (1)

1043	"How big is the lyme problem? Using novel methods to estimate the true number of lyme disease cases in british columbia residents from 1997 to 2008".ti. and "2011".yr. (1)

1044	"Passive tick surveillance, dog seropositivity, and incidence of human lyme disease".ti. and "2004".yr. (1)

1045	"Empiric antibiotic treatment of erythema migrans-like skin lesions as a function of geography: A clinical and cost effectiveness modeling study".ti. and "2013".yr. (1)

1046	"Evaluation of recombinant line immunoblot for detection of lyme disease in slovakia: Comparison with two other immunoassays".ti. and "2008".yr. (1)

1047	"Long-term lyme disease antibiotic therapy beliefs among new england residents".ti. and "2011".yr. (1)

1048	"A serological survey about zoonoses in the verkhoyansk area, northeastern siberia (sakha republic, russian federation)".ti. and "2016".yr. (1)

1049	"Seroepidemiology of nine zoonoses in viljujsk, republic of sakha (northeastern siberia, russian federation)".ti. and "2011".yr. (1)

1050	"Estimated incidence of erythema migrans in five regions of france and ecological correlations with environmental characteristics".ti. and "2013".yr. (1)

1051	"Precipitation and the occurrence of lyme disease in the northeastern united states".ti. and "2004".yr. (1)

1052	"The occurrence of spotted fever rickettsioses and other tick-borne infections in forest workers in poland".ti. and "2011".yr. (1)

1053	"A school-based intervention to increase lyme disease preventive measures among elementary school-aged children".ti. and "2016".yr. (1)

1054	"Comparison of erythema migrans caused by borrelia burgdorferi and borrelia garinii".ti. and "2011".yr. (1)

1055	"Comparison of post-lyme borreliosis symptoms in erythema migrans patients with positive and negative borrelia burgdorferi sensu lato skin culture".ti. and "2011".yr. (1)

1056	"Using nation-wide health insurance claims data to augment lyme disease surveillance".ti. and "2015".yr. (1)

1057	"Two introductions of lyme disease into connecticut: A geospatial analysis of human cases from 1984 to 2012".ti. and "2015".yr. (1)

1058	"Is localized scleroderma caused by borrelia burgdorferi?".ti. and "2016".yr. (1)

1059	"Tick-borne encephalitis and lyme disease in hungary: The epidemiological situation between 1998 and 2008".ti. and "2013".yr. (1)

1060	"Ticking all the boxes? A systematic review of education and communication interventions to prevent tick-borne disease".ti. and "2012".yr. (1)

1061	"Subset-directed antiviral treatment of 142 herpesvirus patients with chronic fatigue syndrome".ti. and "2010".yr. (0)

1062	"The pilot studies for lyme disease (lyme borreliosis) in blood donors in rckik in poznan".ti. and "2015".yr. (0)

1063	"Tick exposure and lyme disease at a summer camp in maryland".ti. and "2005".yr. (1)

1064	"Children with multiple erythema migrans: Are there any pre-treatment symptoms and/or signs suggestive for central nervous system involvement?".ti. and "2002".yr. (1)

1065	"Solitary erythema migrans in children: Comparison of treatment with azithromycin and phenoxymethylpenicillin".ti. and "2002".yr. (1)

1066	"Development and laboratory evaluation of a new recombinant elisa for the serodiagnosis of lyme disease".ti. and "2002".yr. (1)

1067	"Solitary borrelial lymphocytoma in adult patients".ti. and "2002".yr. (1)

1068	"Development of erythema migrans in spite of treatment with antibiotics after a tick bite".ti. and "2002".yr. (1)

1069	"Electrocardiographic findings in patients with erythema migrans".ti. and "2002".yr. (1)

1070	"Evaluation of immunofluorescence test (ift) and immuno (western) blot (wb) test in patients with erythema migrans".ti. and "2002".yr. (1)

1071	"Clinical and epidemiological findings for patients with erythema migrans. Comparison of cohorts from the years 1993 and 2000".ti. and "2002".yr. (1)

1072	"Limitation of serological testing for lyme borreliosis: Evaluation of elisa and western blot in comparison with pcr and culture methods".ti. and "2002".yr. (1)

1073	"Clinical appearance of erythema migrans caused by borrelia afzelii and borrelia garinii--effect of the patient's sex".ti. and "2006".yr. (1)

1074	"Comparison of immunofluorescence assay (ifa) and liaison in patients with different clinical manifestations of lyme borreliosis".ti. and "2006".yr. (1)

1075	"Paradigm burgenland: Risk of borrelia burgdorferi sensu lato infection indicated by variable seroprevalence rates in hunters".ti. and "2006".yr. (1)

1076	"Clinical and epidemiological features of lyme borreliosis in bulgaria".ti. and "2004".yr. (1)

1077	"A comparison of two treatment regimens of ceftriaxone in late lyme disease".ti. and "2005".yr. (1)

1078	"Peripheral facial palsy as an initial symptom of lyme neuroborreliosis in an austrian endemic area".ti. and "2015".yr. (0)

1079	"Contact with horses is a risk factor for tick-borne lymphadenopathy".ti. and "2012".yr. (1)

1080	"Electrocardiographic findings in children with erythema migrans".ti. and "2006".yr. (1)

1081	"Solitary erythema migrans in children: Comparison of treatment with clarithromycin and amoxicillin".ti. and "2012".yr. (1)

1082	"Doxycycline versus ceftriaxone for the treatment of patients with chronic lyme borreliosis".ti. and "2006".yr. (1)

1083	"Prevalence and incidence of lyme borreliosis among slovene forestry workers during the period of tick activity".ti. and "2005".yr. (1)

1084	"Appalachian trail hikers' ability to recognize lyme disease by visual stimulus photographs".ti. and "2014".yr. (1)

1085	"Deer hunting in a residential community: The community's perspective".ti. and "2003".yr. (0)

1086	"Lyme disease testing in children in an endemic area".ti. and "2011".yr. (1)

1087	"Our experience with enzyme-linked immunospot assay in the laboratory diagnostics of lyme borreliosis".ti. and "2015".yr. (0)

1088	"Seroprevalence of anti-toxoplasma gondii and anti-borrelia species antibodies in patients with schizophrenia: A case-control study from western turkey".ti. and "2015".yr. (1)

1089	"Respiratory modulation of cardiac vagal tone in lyme disease".ti. and "2014".yr. (1)

1090	"Lyme disease in west virginia: An assessment of distribution and clinicians' knowledge of disease and surveillance".ti. and "2016".yr. (1)

1091	"The importance of the size of erythema migrans (em) for diagnosis of lyme borreliosis in slovenian children o pomembnosti velikosti erythema migrans (em) za diagnozo lymske borelioze pri otrocih v sloveniji".ti. and "2012".yr. (0)

1092	"Serologic diagnosis of lyme disease with indirect immunofluorescence assay, enzyme-linked immunosorbent assay and western blot".ti. and "2011".yr. (0)

1093	"Preliminary survey of ticks and tick-borne pathogens in tianjin, china".ti. and "2013".yr. (0)

1094	"Confirmed exposure to tick-borne encephalitis virus and probable human cases of tick-borne encephalitis in central/northern anatolia, turkey".ti. and "2011".yr. (1)

1095	"Lyme disease surveillance using sampling estimation: Evaluation of an alternative methodology in new york state".ti. and "2016".yr. (0)

1096	"Tick bite prophylaxis: Results from a 2012 survey of healthcare providers".ti. and "2015".yr. (1)

1097	"Characterizing lyme disease surveillance in an endemic state".ti. and "2016".yr. (0)

1098	"Underreporting of lyme and other tick-borne diseases in residents of a high-incidence county, minnesota, 2009".ti. and "2016".yr. (0)

1099	or/1-1098 (795)

1100	1099 (795)

1101	limit 1100 to english language (793)

1102	1100 not 1101 (2)

1103	limit 1100 to (english language or no language specified) (793)

1104	1100 not 1103 (2)

1105	limit 1100 to (english language or no language specified or multilingual or undetermined) (793)

1106	1100 not 1105 (2)

1107	((afrikaans or albanian or arabic or armenian or azerbaijani or belorussian or bengali or bosnian or bulgarian or burmese or catalan or chinese or croatian or czech or danish or dutch or esperanto or estonian or finnish or flemish or french or gaelic, scots or georgian or german or greek or hausa or hebrew or hindi or hungarian or icelandic or indonesian or italian or japanese or kinyarwanda or kirghiz or korean or latin or latvian or lithuanian or macedonian or malay or malayalam or marathi or maori or masai or norwegian or persian or polish or portuguese or pushto or romanian or rumanian or russian or serbian or slovak or slovene or spanish or swahili or swedish or tagalog or tamil or telugu or thai or turkish or ukrainian or urdu or vietnamese or welsh) not english).lg. (4603029)

1108	1100 not 1107 (793)

1109	1100 and 1107 (2)

1110	((afrikaans or albanian or arabic or armenian or azerbaijani or belorussian or bengali or bosnian or bulgarian or burmese or catalan or chinese or croatian or czech or danish or dutch or esperanto or estonian or finnish or flemish or french or gaelic, scots or georgian or german or greek or hausa or hebrew or hindi or hungarian or icelandic or indonesian or italian or japanese or kinyarwanda or kirghiz or korean or latin or latvian or lithuanian or macedonian or malay or malayalam or marathi or maori or masai or norwegian or persian or polish or portuguese or pushto or romanian or rumanian or russian or serbian or slovak or slovene or spanish or swahili or swedish or tagalog or tamil or telugu or thai or turkish or ukrainian or urdu or vietnamese or welsh) not (english or undetermined or multilingual)).lg. (4602987)

1111	1100 not 1110 (793)

1112	1100 and 1110 (2)

**Includes: Embase** via Ovid http://ovidsp.ovid.com/

Date range searched: <1974 to 2024 February 01>

Date searched: 2nd February 2024

1	"Exposure and preventive behaviours toward ticks and lyme disease in canada: Results from a first national survey".ti. and "2017".yr. (1)

2	"Lyme and tick-borne infections patient survey".ti. and "2012".yr. (0)

3	"An internet survey of lyme patients experience of access to care".ti. and "2013".yr. (0)

4	"A patient survey of a community: Lyme patients experiences of health care".ti. and "2013".yr. (0)

5	"Lyme disease on the nhs: Patient survey results 2016".ti. and "2016".yr. (0)

6	"Lyme disease, data and statistics".ti. and "2016".yr. (0)

7	"Patient narratives and an umwelten-based account of the more-than-human".ti. and "2016".yr. (0)

8	"Stress, despair, and quality of life: The lived experience of families with children below 26 years of age diagnosed with lyme disease".ti. and "2016".yr. (0)

9	"Zoonoses report united kingdom 2003".ti. and "2004".yr. (0)

10	"Zoonoses summary report: Uk 2014".ti. and "2015".yr. (0)

11	"National lyme disease surveillance in canada 2013: Web report".ti. and "2015".yr. (0)

12	"National lyme disease surveillance in canada 2009-2012".ti. and "2015".yr. (0)

13	"Surveillance of lyme disease in canada".ti. and "2016".yr. (0)

14	"Epidemiology and risks of tick-borne lyme borreliosis".ti. and "2009".yr. (0)

15	"Lyme neuroborreliosis in ireland 2012 and 2013".ti. and "2013".yr. (0)

16	"Foreign travel associated illness, england, wales and northern ireland: 2007 report".ti. and "2007".yr. (0)

17	"Pediatric lyme disease prevention".ti. and "2015".yr. (0)

18	"Tick-borne diseases: Assessing the knowledge, attitudes, and behaviors of college students".ti. and "2015".yr. (0)

19	"Fight lyme now survey".ti. and "2017".yr. (0)

20	"Common animal associated infections quarterly report (england and wales) - third quarter 2014".ti. and "2014".yr. (0)

21	"Lyme disease in ireland survey results 2016".ti. and "2016".yr. (0)

22	"Nhs testing and investigation of lyme borreliosis – patient experiences survey".ti. and "2014".yr. (0)

23	"German patient survey 2015 results".ti. and "2015".yr. (0)

24	"Waterborne zoonoses".ti. and "2004".yr. (0)

25	"How to design work related information for low-literate employees?".ti. and "2014".yr. (0)

26	"Multi-center validation of a clinical decision rule to distinguish lyme from aseptic meningitis in children in lyme disease endemic areas".ti. and "2011".yr. (1)

27	"Lyme disease as a cause of aseptic meningitis in children in a lyme-endemic region...2007 society for academic emergency medicine annual meeting".ti. and "2007".yr. (0)

28	"Clinical predictors of lyme disease among pediatric patients with acute monoarticular arthritis...2007 society for academic emergency medicine annual meeting".ti. and "2007".yr. (0)

29	"Acute polyradiculoneuritis syndrome: Clinical observations and differential diagnosis".ti. and "2012".yr. (1)

30	"Course of borrelia burgdorferi dna shedding in urine after treatment".ti. and "2007".yr. (1)

31	"The many faces of solitary and multiple erythema migrans".ti. and "2013".yr. (1)

32	"Clinical spectrum of skin manifestations of lyme borreliosis in 204 children in austria".ti. and "2015".yr. (1)

33	"Detection of spirochaetal dna simultaneously in skin biopsies, peripheral blood and urine from patients with erythema migrans".ti. and "2004".yr. (1)

34	"Granuloma annulare displaying pseudorosettes in borelia infection".ti. and "2008".yr. (1)

35	"Laboratory diagnostic methods in suspected disseminated lyme disease: A comparison of different techniques".ti. and "2011".yr. (1)

36	"Peripheral neuropathy in acrodermatitis chronica atrophicans - effect of treatment".ti. and "2002".yr. (1)

37	"Chronic symptoms are common in patients with neuroborreliosis -- a questionnaire follow-up study".ti. and "2002".yr. (1)

38	"European neuroborreliosis: Quality of life 30 months after treatment".ti. and "2011".yr. (0)

39	"Risk factors for a non-favorable outcome after treated european neuroborreliosis".ti. and "2013".yr. (1)

40	"Pcr in lyme neuroborreliosis: A prospective study".ti. and "2005".yr. (1)

41	"Multiple sclerosis in vest-agder county, norway".ti. and "2011".yr. (1)

42	"Cxcl13 chemokine in pediatric and adult neuroborreliosis".ti. and "2011".yr. (1)

43	"Use of electrocochleography for assessing endolymphatic hydrops in patients with lyme disease and meniere's disease".ti. and "2002".yr. (1)

44	"Brain magnetic resonance imaging does not contribute to the diagnosis of chronic neuroborreliosis".ti. and "2007".yr. (1)

45	"The analysis of the course of early localized lyme disease among patients of the infectious diseases clinic in szczecin".ti. and "2013".yr. (0)

46	"Borrelia burgdorferi sensu lato in brazil: Occurrence confirmed by immunohistochemistry and focus floating microscopy".ti. and "2010".yr. (1)

47	"Lyme neuroborreliosis".ti. and "2011".yr. and Acta Veterinaria.jn. (0)

48	"Serological markers of borrelia burgdorferi infection among forestry workers in west pomerania during a five-year period".ti. and "2004".yr. (0)

49	"Lyme borreliosis--the utility of improved real-time pcr assay in the detection of borrelia burgdorferi infections".ti. and "2015".yr. (1)

50	"Non-specific arthralgia as the main manifestation of borrelia burgdorferi sensu lato infection".ti. and "2008".yr. (1)

51	"Rational design of antigens to improve the serodiagnosis of tick-borne borreliosis in central regions of russia".ti. and "2014".yr. (1)

52	"Lyme disease in poland - a serious problem?".ti. and "2016".yr. (1)

53	"Recent lesson from a clinical and seroepidemiological survey: Low positive predictive value of borrelia burgdorferi antibody testing in a high risk population".ti. and "2012".yr. (1)

54	"Atovaquone plus cholestyramine in patients coinfected with babesia microti and borrelia burgdorferi refractory to other treatment".ti. and "2006".yr. (1)

55	"Mr imaging assessment of brain and cervical cord damage in patients with neuroborreliosis".ti. and "2006".yr. (1)

56	"Mri features of lyme arthritis in children".ti. and "2005".yr. (1)

57	"Immunological aspects of lyme borreliosis co-infection".ti. and "2011".yr. (1)

58	"Longitudinal evaluation of an educational intervention for preventing tick bites in an area with endemic lyme disease in baltimore county, maryland".ti. and "2003".yr. (1)

59	"Lyme disease incidence in wisconsin: A comparison of state-reported rates and rates from a population-based cohort".ti. and "2002".yr. (1)

60	"Effects of climate on variability in lyme disease incidence in the northeastern united states".ti. and "2003".yr. (1)

61	"Medical risks of wilderness hiking".ti. and "2003".yr. (1)

62	"Long-term follow-up of patients with culture-confirmed lyme disease".ti. and "2003".yr. (1)

63	"Systemic symptoms without erythema migrans as the presenting picture of early lyme disease".ti. and "2003".yr. (1)

64	"Lyme borreliosis and facial paralysis--a prospective analysis of risk factors and outcome".ti. and "2002".yr. (1)

65	"Higher prevalence of antibodies to borrelia burgdorferi in psychiatric patients than in healthy subjects".ti. and "2002".yr. (1)

66	"Borrelia-associated primary cutaneous malt lymphoma in a nonendemic region".ti. and "2003".yr. (1)

67	"Fourteen-year seroepidemiological study of zoonoses in a greek village".ti. and "2002".yr. (1)

68	"Lyme disease testing by multiplexed bead analysis using vlse1 and pepc10 antigens compared with 2-tiered testing".ti. and "2010".yr. (1)

69	"Focus floating microscopy: Gold standard for cutaneous borreliosis?".ti. and "2007".yr. (2)

70	"Cutaneous lymphoid hyperplasia and other lymphoid infiltrates of the breast nipple: A retrospective clinicopathologic study of fifty-six patients".ti. and "2005".yr. (1)

71	"Acrodermatitis chronica atrophicans with pseudolymphomatous infiltrates".ti. and "2013".yr. (1)

72	"Association between lyme disease and weather types in new york state".ti. and "2013".yr. (1)

73	"An evaluation of a lyme disease prevention program in a working population".ti. and "2006".yr. (1)

74	"Emergence of a persistent symptomatology in a cohort of patients reporting previous diagnosis and treatment for lyme disease".ti. and "2015".yr. (0)

75	"Subjective symptoms after treatment of early lyme disease".ti. and "2010".yr. (1)

76	"Etiology of pericarditis in a prospective cohort of 1162 cases".ti. and "2015".yr. (1)

77	"Psychiatric comorbidity and other psychological factors in patients with chronic lyme disease ".ti. and "2009".yr. (1)

78	"Long-term assessment of fatigue in patients with culture-confirmed lyme disease".ti. and "2015".yr. (1)

79	"No value for routine serologic screening for borrelia burgdorferi in patients with uveitis in the netherlands".ti. and "2016".yr. (1)

80	"Cardiac tropism of borrelia burgdorferi: An autopsy study of sudden cardiac death associated with lyme carditis".ti. and "2016".yr. (1)

81	"Peridomestic lyme disease prevention: Results of a population-based case-control study".ti. and "2009".yr. (1)

82	"Risk perceptions regarding ticks and lyme disease: A national survey".ti. and "2004".yr. (1)

83	"Disparities in reportable communicable disease incidence by census tract-level poverty, new york city, 2006-2013".ti. and "2015".yr. (1)

84	"Hyperplasia of hair follicles and other adnexal structures in cutaneous lymphoproliferative disorders: A study of 53 cases, including so-called pseudolymphomatous folliculitis and overt lymphomas".ti. and "2008".yr. (1)

85	"Spatial patterns of lyme disease risk in california based on disease incidence data and modeling of vector-tick exposure".ti. and "2006".yr. (1)

86	"Reinfection and relapse in early lyme disease".ti. and "2006".yr. (1)

87	"Lyme disease risk influences human settlement in the wildland-urban interface: Evidence from a longitudinal analysis of counties in the northeastern united states".ti. and "2014".yr. (1)

88	"Spatial and temporal emergence pattern of lyme disease in virginia".ti. and "2014".yr. (1)

89	"Meteorological influences on the seasonality of lyme disease in the united states".ti. and "2014".yr. (1)

90	"Survey for tick-borne zoonoses in the state of espirito santo, southeastern brazil".ti. and "2010".yr. (1)

91	"Human behavioral and ecological risk factors for lyme disease infection on block island, rhode island".ti. and "2013".yr. (1)

92	"Linking acarological risk and lyme disease incidence in the usa".ti. and "2011".yr. (1)

93	"Lyme disease and convergence insufficiency: Is it a near fit?".ti. and "2006".yr. (0)

94	"Epidemiological, clinical and paraclinical aspects of lyme disease in bihor county between 2007-2011".ti. and "2011".yr. (0)

95	"Particular aspects of lyme disease in bihor county".ti. and "2014".yr. (0)

96	"Borrelia burgdorferi infection among forestry workers - assessed with an immunoenzymatic method (elisa), pcr and correlated with the clinical state of the patients".ti. and "2003".yr. (1)

97	"Clinical characteristics and treatment outcome of early lyme disease in patients with microbiologically confirmed erythema migrans".ti. and "2002".yr. (1)

98	"Duration of antibiotic therapy for early lyme disease. A randomized, double-blind, placebo-controlled trial".ti. and "2003".yr. (1)

99	"Risk of tick-borne bacterial diseases in humans in the florence area, tuscany".ti. and "2003".yr. (1)

100	"Epidemiological and clinical differences among rickettsia slovaca rickettsiosis and other tick-borne diseases in spain".ti. and "2003".yr. (1)

101	"Bartonella henselae and borrelia burgdorferi infections of the central nervous system".ti. and "2003".yr. (1)

102	"Lyme borreliosis in south-eastern poland: Relationships with environmental factors and medical attention standards".ti. and "2011".yr. (0)

103	"Occurrence of cases of borreliosis certified as an occupational disease in the province of wielkopolska (poland)".ti. and "2009".yr. (1)

104	"Seroepidemiological study of lyme borreliosis among forestry workers in southern poland".ti. and "2009".yr. (1)

105	"Correlation of tests for detection of borrelia burgdorferi sensu lato infection in patients with diagnosed borreliosis".ti. and "2006".yr. (1)

106	"Serological survey in persons occupationally exposed to tick-borne pathogens in cases of co-infections with borrelia burgdorferi, anaplasma phagocytophilum, bartonella spp. And babesia microti".ti. and "2012".yr. (1)

107	"Risk of tick-borne bacterial diseases among workers of roztocze national park (south-eastern poland)".ti. and "2005".yr. (1)

108	"Study on lyme borreliosis focus in the lublin region (eastern poland)".ti. and "2008".yr. (1)

109	"Risk of tick-borne diseases in various categories of employment among forestry workers in eastern poland".ti. and "2012".yr. (1)

110	"Incidence and prevalence of infection with anaplasma phagocytophilum. Prospective study in healthy individuals exposed to ticks".ti. and "2004".yr. (1)

111	"Human anaplasmosis in north-eastern poland: Seroprevalence in humans and prevalence in ixodes ricinus ticks".ti. and "2004".yr. (1)

112	"Detecting dnas of anaplasma phagocytophilum and babesia in the blood of patients suspected of lyme disease".ti. and "2004".yr. (1)

113	"Occurrence of igg antibodies to anaplasma phagocytophilum in humans suspected of lyme borreliosis in eastern slovakia".ti. and "2009".yr. (1)

114	"Molecular and serological diagnosis of borrelia burgdorferi infection among patients with diagnosed erythema migrans".ti. and "2007".yr. (1)

115	"Epidemiology of lyme disease among workers of forest inspectorates in poland".ti. and "2013".yr. (1)

116	"Exposure to ticks and seroprevalence of borrelia burgdorferi among a healthy young population living in the area of southern podlasie, poland".ti. and "2014".yr. (1)

117	"Occupational exposure to zoonotic agents among agricultural workers in lombardy region, northern italy".ti. and "2013".yr. (1)

118	"Association between incidence of lyme disease and spring-early summer season temperature changes in hungary--1998-2010".ti. and "2013".yr. (1)

119	"Sero-epidemiological study of lyme disease among high-risk population groups in eastern slovakia".ti. and "2015".yr. (1)

120	"Risk compensation and vaccination: Can getting vaccinated cause people to engage in risky behaviors?".ti. and "2007".yr. (1)

121	"Risk perceptions and their relation to risk behavior".ti. and "2004".yr. (1)

122	"Prophylaxis with single-dose doxycycline for the prevention of lyme disease after an ixodes scapularis tick bite".ti. and "2002".yr. (0)

123	"Potential misdiagnoses of bell's palsy in the emergency department".ti. and "2014".yr. (1)

124	"Appropriateness of lyme disease serologic testing".ti. and "2004".yr. (1)

125	"Brief communication: Hematogenous dissemination in early lyme disease.[summary for patients in ann intern med. 2005 may 3;142(9):I48; pmid: 15867401]".ti. and "2005".yr. (0)

126	"Effect of iv antibiotic therapy on antibody profile in patients with post-treatment lyme encephalopathy".ti. and "2010".yr. (1)

127	"Anti-flagellin antibodies in patients with pandas-like illnesses".ti. and "2010".yr. (1)

128	"Prevalence of chlamydophila psittaci and borrelia burgdorferi infections in a series of 108 primary cutaneous lymphomas".ti. and "2011".yr. (1)

129	"Human exposure to anaplasma phagocytophilum in portugal".ti. and "2006".yr. (1)

130	"Joint involvement in russian patients with lyme borreliosis presenting with acrodermatitis chronica atrophicans".ti. and "2003".yr. (0)

131	"Is there an association between spondyloarthritis and antibodies towards borrelia, ehrlichia and chlamydia species?".ti. and "2015".yr. (1)

132	"Antibodies to borrelia burgdorferi in sera and synovial fluid in patients with lyme arthritis".ti. and "2006".yr. (0)

133	"Juvenile lyme arthritis".ti. and "2003".yr. (0)

134	"Serologic validation of four western blots for diagnosis of borrelia burgdorferi infection".ti. and "2009".yr. (1)

135	"Simultaneous use of serum igg and igm for risk scoring of suspected early lyme borreliosis: Graphical and bivariate analyses".ti. and "2010".yr. (1)

136	"Lyme borreliosis in sweden--diagnostic performance of five commercial borrelia serology kits using sera from well-defined patient groups".ti. and "2004".yr. (1)

137	"Seroprevalence of antibodies to borrelia burgdorferi sensu lato in healthy adults from western norway: Risk factors and methodological aspects".ti. and "2014".yr. (1)

138	"Improved serodiagnosis of early lyme borreliosis: Immunoblot with local borrelia afzelii strain".ti. and "2003".yr. (0)

139	"C6-peptide serology as diagnostic tool in neuroborreliosis".ti. and "2008".yr. (1)

140	"Memory and executive functions in adolescents with posttreatment lyme disease".ti. and "2008".yr. (1)

141	"Possible role of borrelia burgdorferi sensu lato infection in lichen sclerosus".ti. and "2008".yr. (1)

142	"How accurate is a clinical diagnosis of erythema chronicum migrans? Prospective study comparing the diagnostic accuracy of general practitioners and dermatologists in an area where lyme borreliosis is endemic".ti. and "2004".yr. (1)

143	"Lyme disease - united states, 2000".ti. and "2002".yr. and MMWR - Morbidity & Mortality Weekly Report.jn. (0)

144	"Clinical presentation of childhood neuroborreliosis; neurological examination may be normal".ti. and "2010".yr. (1)

145	"Seroprevalence of borrelia igg antibodies among young swedish children in relation to reported tick bites, symptoms and previous treatment for lyme borreliosis: A population-based survey".ti. and "2010".yr. (1)

146	"Lyme meningitis, the major cause of childhood meningitis in an endemic area: A population based study".ti. and "2012".yr. (0)

147	"Integrating spatial epidemiology into a decision model for evaluation of facial palsy in children".ti. and "2011".yr. (1)

148	"Epidemiological situation of lyme borreliosis in belgium, 2003 to 2012".ti. and "2015".yr. (0)

149	"Serodiagnosis of borreliosis: Indirect immunofluorescence assay, enzyme-linked immunosorbent assay and immunoblotting".ti. and "2011".yr. (1)

150	"Hyperosmia in lyme disease".ti. and "2014".yr. (1)

151	"Role of psychiatric comorbidity in chronic lyme disease".ti. and "2008".yr. (1)

152	"Burden and viability of borrelia burgdorferi in skin and joints of patients with erythema migrans or lyme arthritis".ti. and "2011".yr. (1)

153	"A82: Lyme arthritis: An emerging clinical problem in nova scotia, canada".ti. and "2014".yr. (0)

154	"Brief report: Long-term assessment of fibromyalgia in patients with culture-confirmed lyme disease".ti. and "2015".yr. (1)

155	"The utility of borrelia burgdorferi specific pcr of synovial fluid as a diagnostic tool for lyme arthritis".ti. and "2002".yr. (0)

156	"Prolonged synovitis in pediatric lyme disease: A study on clinical and laboratory predictors".ti. and "2002".yr. (0)

157	"A comparison of the presentation, synovial fluid analysis, and treatment course in children and adults with lyme arthritis".ti. and "2010".yr. (1)

158	"Prevalence and clinical presentation of lyme arthritis in a large cohort of patients with recent-onset arthritis".ti. and "2013".yr. (1)

159	"Sleep disturbances, fatigue and memory complaints attributed to chronic lyme disease".ti. and "2003".yr. (0)

160	"Maladaptive schemas in chronic lyme disease".ti. and "2004".yr. (0)

161	"Co-morbid mental disorders in chronic lyme disease: The first 50 patients".ti. and "2003".yr. (0)

162	"A prospective, longitudinal cohort study evaluating psychosocial risk and protective factors for post lyme disease syndrome".ti. and "2010".yr. (1)

163	"A lyme disease education program changes knowledge, attitudes and behaviors in elementary school children living in an endemic area".ti. and "2004".yr. (0)

164	("Diminished health-related quality-of-life improves over time in lyme disease" and "12 year followup from the nantucket lyme disease cohort study").ti. and "2002".yr. (0)

165	"Components of the health belief model are determinants of lyme disease preventive behavior among school-aged children in an endemic area of massachusetts".ti. and "2006".yr. (0)

166	"Interest of systematic lyme serology in context of recent onset arthritis".ti. and "2014".yr. (1)

167	"Lyme arthritis: A comparison of presentation, synovial fluid analysis, and treatment course in children and adults".ti. and "2013".yr. (1)

168	"Cost-effectiveness analysis of the lyme disease vaccine".ti. and "2002".yr. (1)

169	"Investigation of anaplasmosis in yiyuan county, shandong province, china".ti. and "2011".yr. (1)

170	"Seropositivity for anti-borrelia igg antibody is independently associated with carotid atherosclerosis".ti. and "2006".yr. (1)

171	"Gender differences in childhood lyme neuroborreliosis".ti. and "2015".yr. (1)

172	"Evaluation of a new euroimmun anti-borrelia select elisa".ti. and "2013".yr. (1)

173	"Lyme disease in the differential diagnosis of psychiatric patients".ti. and "2003".yr. (0)

174	"Seroepidemiological investigation of lyme disease and human granulocytic anaplasmosis among people living in forest areas of eight provinces in china".ti. and "2013".yr. (1)

175	"Evaluation of six recombinant proteins for serological diagnosis of lyme borreliosis in china".ti. and "2016".yr. (1)

176	"A study of the technique of western blot for diagnosis of lyme disease caused by borrelia afzelii in china".ti. and "2013".yr. (1)

177	"Absence of borrelia burgdorferi in the myocardium of subjects with normal left ventricular systolic function: A study using pcr and electron microscopy".ti. and "2016".yr. (1)

178	"Ethic of consensibility, subaltern ethicality: The clinical application of embryonic stem cells in india".ti. and "2013".yr. (0)

179	"Tick-borne infections and pediatric bipolar disorder".ti. and "2015".yr. (1)

180	"Personal and family history of immune-related conditions increase the risk of plasma cell disorders: A population-based study".ti. and "2011".yr. (1)

181	"Borrelia infection and risk of non-hodgkin lymphoma".ti. and "2008".yr. (1)

182	"P05.22. Health beliefs and experiences of patients with chronic lyme disease: A qualitative study...International research congress on integrative medicine and health 2012 portland, oregon, usa. 15-18 may 2012".ti. and "2012".yr. (0)

183	"Exploring the association between morgellons disease and lyme disease: Identification of borrelia burgdorferi in morgellons disease patients".ti. and "2015".yr. (1)

184	"Experiences of patients identifying with chronic lyme disease in the healthcare system: A qualitative study".ti. and "2014".yr. (1)

185	"Diagnostic challenges of early lyme disease: Lessons from a community case series".ti. and "2009".yr. (1)

186	"Probable late lyme disease: A variant manifestation of untreated borrelia burgdorferi infection".ti. and "2012".yr. (1)

187	"Clinical characteristics and cerebrospinal fluid parameters in patients with peripheral facial palsy caused by lyme neuroborreliosis compared with facial palsy of unknown origin (bell's palsy)".ti. and "2011".yr. (1)

188	"Utilization of serology for the diagnosis of suspected lyme borreliosis in denmark: Survey of patients seen in general practice".ti. and "2010".yr. (1)

189	"Lyme arthritis in southern norway--an endemic area for lyme borreliosis".ti. and "2014".yr. (1)

190	"Public health impact of strain specific immunity to borrelia burgdorferi".ti. and "2015".yr. (1)

191	"The diagnostic accuracy of serological tests for lyme borreliosis in europe: A systematic review and meta-analysis".ti. and "2016".yr. (1)

192	"Doxycycline-mediated effects on persistent symptoms and systemic cytokine responses post-neuroborreliosis: A randomized, prospective, cross-over study".ti. and "2012".yr. (1)

193	"Mapping of hormones and cortisol responses in patients after lyme neuroborreliosis".ti. and "2010".yr. (0)

194	"Efficacy and safety of pharmacological treatments for lyme neuroborreliosis in children: A systematic review".ti. and "2015".yr. (0)

195	"Neurocognitive functions and brain atrophy after proven neuroborreliosis: A case-control study".ti. and "2015".yr. (1)

196	"The nebop score - a clinical prediction test for evaluation of children with lyme neuroborreliosis in europe".ti. and "2015".yr. (1)

197	"Multi-criteria decision analysis as an innovative approach to managing zoonoses: Results from a study on lyme disease in canada".ti. and "2013".yr. (1)

198	"Factors associated with preventive behaviors regarding lyme disease in canada and switzerland: A comparative study".ti. and "2015".yr. (1)

199	"Acceptability of tick control interventions to prevent lyme disease in switzerland and canada: A mixed-method study".ti. and "2016".yr. (1)

200	"From lyme disease emergence to endemicity: A cross sectional comparative study of risk perceptions in different populations".ti. and "2014".yr. (1)

201	"Education on tick bite and lyme borreliosis prevention, aimed at schoolchildren in the netherlands: Comparing the effects of an online educational video game versus a leaflet or no intervention".ti. and "2016".yr. (1)

202	"Study on public perceptions and protective behaviors regarding lyme disease among the general public in the netherlands: Implications for prevention programs".ti. and "2013".yr. (1)

203	"Comparing the effect of a leaflet and a movie in preventing tick bites and lyme disease in the netherlands".ti. and "2016".yr. (1)

204	"Determinants and protective behaviours regarding tick bites among school children in the netherlands: A cross-sectional study".ti. and "2013".yr. (1)

205	"Subjective health complaints are not associated with tick bites or antibodies to borrelia burgdorferi sensu lato in blood donors in western norway: A cross-sectional study".ti. and "2015".yr. (1)

206	"Decrease in tick bite consultations and stabilization of early lyme borreliosis in the netherlands in 2014 after 15 years of continuous increase".ti. and "2016".yr. (1)

207	"Are the current notification criteria for lyme borreliosis in norway suitable? Results of an evaluation of lyme borreliosis surveillance in norway, 1995-2013".ti. and "2016".yr. (1)

208	"Experience of lyme disease and preferences for precautions: A cross-sectional survey of uk patients".ti. and "2013".yr. (1)

209	"Exploratory spatial analysis of lyme disease in texas -what can we learn from the reported cases?".ti. and "2015".yr. (1)

210	"Early lyme disease with spirochetemia - diagnosed by dna sequencing".ti. and "2010".yr. (0)

211	"Long-term prognosis of patients treated for erythema migrans in france".ti. and "2002".yr. (1)

212	"Borreliosis - a cause of obstructive sleep apnea syndrome".ti. and "2012".yr. (1)

213	"Contribution to laboratory diagnostics of neuroborreliosis".ti. and "2003".yr. (1)

214	"Reported incidence of lyme disease in slovakia and antibodies to b. Burgdorferi antigens detected in healthy population".ti. and "2005".yr. (1)

215	"Our experience with examination of antibodies against antigens of borrelia burgdorferi in patients with suspected lyme disease".ti. and "2010".yr. (1)

216	"Antibodies to borrelia burgdorferi in erythema migrans patients".ti. and "2007".yr. (1)

217	"Prevalence of antibodies to borrelia burgdorferi sensu stricto in humans from a cuban village".ti. and "2012".yr. (1)

218	"Zoonoses in humans from small rural properties in jataizinho, parana, brazil".ti. and "2013".yr. (0)

219	"Borrelia burgdorferi sensu lato in humans in a rural area of parana state, brazil".ti. and "2015".yr. (1)

220	"Seroprevalence of borrelia burgdorferi in occupationally exposed persons in the belgrade area, serbia".ti. and "2015".yr. (1)

221	"Laboratory diagnosis of lyme borreliosis in scottish patients: A novel approach".ti. and "2014".yr. (2)

222	"Mycosis fungoides: Is it a borrelia burgdorferi-associated disease?".ti. and "2006".yr. (1)

223	"Morphoea: A manifestation of infection with borrelia species?".ti. and "2007".yr. (1)

224	"Lyme neuroborreliosis: A treatable cause of acute ocular motor disturbances in children".ti. and "2015".yr. (1)

225	"Intermediate uveitis in children and young adults: Differences in clinical course, associations and visual outcome".ti. and "2014".yr. (1)

226	"Clinico-biological aspects in human neuroborreliosis".ti. and "2013".yr. (0)

227	"Awareness that dogs can be carriers for ticks that transmit lyme disease".ti. and "2016".yr. (0)

228	"Surveillance for lyme disease in canada, 2009 to 2012".ti. and "2015".yr. (0)

229	"Descriptive epidemiology of lyme disease in ontario: 1999-2004".ti. and "2006".yr. (1)

230	"Lyme disease: Knowledge, beliefs, and practices of physicians in a low-endemic area".ti. and "2012".yr. (1)

231	"Lyme disease: Knowledge and practices of family practitioners in southern quebec".ti. and "2015".yr. (1)

232	"Challenges with lyme disease diagnostics".ti. and "2012".yr. (1)

233	"Evaluation of a new health canada-approved lyme immunoblot assay for confirmation of positive eia screen results".ti. and "2011".yr. (1)

234	"Immune complexes in early lyme disease".ti. and "2007".yr. (1)

235	"A systematic review and meta-analysis for the adverse effects, immunogenicity and efficacy of lyme disease vaccines: Guiding novel vaccine development".ti. and "2017".yr. (1)

236	"Seroprevalence study of tick-borne encephalitis, borrelia burgdorferi, dengue and toscana virus in turin province".ti. and "2007".yr. (1)

237	"An enhanced elispot assay for sensitive detection of antigen-specific t cell responses to borrelia burgdorferi".ti. and "2013".yr. (1)

238	"Can elispot be applied to a clinical setting as a diagnostic utility for neuroborreliosis?".ti. and "2012".yr. (1)

239	"Study of awareness of tick-borne diseases among children and young people in the czech republic".ti. and "2003".yr. (1)

240	"Long term and repeated electron microscopy and pcr detection of borrelia burgdorferi sensu lato after an antibiotic treatment".ti. and "2004".yr. (1)

241	"Direct detection of borrelia burgdorferi spirochetes in patients with early disseminated lyme borreliosis".ti. and "2009".yr. (1)

242	"Pilot cross-sectional study of three zoonoses (lyme disease, tularaemia, leptospirosis) among healthy blood donors in eastern slovakia".ti. and "2015".yr. (1)

243	"Tick-borne encephalitis: Course and complications - our observations from 2009 to 2012".ti. and "2014".yr. (1)

244	"Standard criteria of western blot for the diagnosis of lyme disease caused by borrelia garinii in china".ti. and "2005".yr. (1)

245	"Diagnosis of lyme borreliosis by polymerase chain reaction".ti. and "2002".yr. (1)

246	"Ability of the borreliacidal antibody test to confirm lyme disease in clinical practice".ti. and "2002".yr. (1)

247	"Detection of borrelia burgdorferi by species-specific and broad-range pcr of synovial fluid and synovial tissue of lyme arthritis patients before and after antibiotic treatment".ti. and "2003".yr. (1)

248	"Disease-specific diagnosis of coinfecting tickborne zoonoses: Babesiosis, human granulocytic ehrlichiosis, and lyme disease".ti. and "2002".yr. (1)

249	"Prospective study of coinfection in patients with erythema migrans".ti. and "2003".yr. (1)

250	"Asymptomatic infection with borrelia burgdorferi".ti. and "2003".yr. (1)

251	"Parental knowledge of and attitudes toward lymerix (recombinant osp--a lyme vaccine)".ti. and "2002".yr. (1)

252	"An open-label, nonrandomized, single-center, prospective extension, clinical trial of booster dose schedules to assess the safety profile and immunogenicity of recombinant outer-surface protein a (ospa) lyme disease vaccine".ti. and "2003".yr. (1)

253	"Serodiagnosis of lyme borreliosis infection using surface plasmon resonance".ti. and "2008".yr. (1)

254	"Long-term effects of immunization with recombinant lipoprotein outer surface protein a on serologic test for lyme disease".ti. and "2004".yr. (1)

255	"Comparison of western immunoblotting and the c6 lyme antibody test for laboratory detection of lyme disease".ti. and "2004".yr. (1)

256	"Ccl19 as a chemokine risk factor for post-treatment lyme disease syndrome: A prospective clinical cohort study".ti. and "2016".yr. (0)

257	"Misdiagnosis of late-onset lyme arthritis by inappropriate use of borrelia burgdorferi immunoblot testing with synovial fluid".ti. and "2012".yr. (1)

258	"Rapid, simple, quantitative, and highly sensitive antibody detection for lyme disease".ti. and "2010".yr. (1)

259	"Lack of serum antibodies against borrelia burgdorferi in children with autism".ti. and "2013".yr. (1)

260	"Humoral immune responses in patients with lyme neuroborreliosis".ti. and "2010".yr. (1)

261	"Study of a cohort of 1,886 persons to determine changes in antibody reactivity to borrelia burgdorferi 3 months after a tick bite".ti. and "2015".yr. (1)

262	"Simple objective detection of human lyme disease infection using immuno-pcr and a single recombinant hybrid antigen".ti. and "2014".yr. (1)

263	"Significantly improved accuracy of diagnosis of early lyme disease by peptide enzyme-linked immunosorbent assay based on the borreliacidal antibody epitope of borrelia burgdorferi ospc".ti. and "2008".yr. (1)

264	"Evaluation of the recombinant vlse-based liaison chemiluminescence immunoassay for detection of borrelia burgdorferi and diagnosis of lyme disease".ti. and "2008".yr. (1)

265	"A decrease in the immunoglobulin g antibody response against the vlse protein of borrelia burgdorferi sensu lato correlates with the resolution of clinical signs in antibiotic-treated patients with early lyme disease".ti. and "2006".yr. (1)

266	"Multiplex immunoassay for lyme disease using vlse1-igg and pepc10-igm antibodies: Improving test performance through bioinformatics".ti. and "2011".yr. (1)

267	"Evaluation of selected borrelia burgdorferi lp54 plasmid-encoded gene products expressed during mammalian infection as antigens to improve serodiagnostic testing for early lyme disease".ti. and "2015".yr. (1)

268	"Impact of clinical variables on borrelia burgdorferi-specific antibody seropositivity in acute-phase sera from patients in north america with culture-confirmed early lyme disease".ti. and "2008".yr. (1)

269	"A novel multivalent ospa vaccine against lyme borreliosis is safe and immunogenic in an adult population previously infected with borrelia burgdorferi sensu lato".ti. and "2014".yr. (1)

270	"Evaluating frequency, diagnostic quality, and cost of lyme borreliosis testing in germany: A retrospective model analysis".ti. and "2012".yr. (1)

271	("The impact of immunosuppression on erythema migrans. A retrospective study of clinical presentation, response to treatment and production of borrelia antibodies in 33 patients".ti. and "2006".yr.) not erratum.pt. (1)

272	"Frequency and clinical features of intraocular inflammation in tokyo".ti. and "2009".yr. (1)

273	"Uveitis in children: Single center experience [p064]".ti. and "2011".yr. (0)

274	"Automated cerebrospinal fluid cell count--new reference ranges and evaluation of its clinical use in central nervous system infections".ti. and "2014".yr. (1)

275	"Liaison (r) borrelia assay: A recombinant antigen based, automated chemiluminescence immunoassay for the diagnosis of lyme disease".ti. and "2006".yr. (0)

276	"Clinical comparison of two commercially available lyme screening elisa assays".ti. and "2007".yr. (0)

277	"Development and validation of a novel ifn-gamma elispot assay for sensitive and specific detection of antigen-specific t cell response to borrelia burgdorferi".ti. and "2013".yr. (1)

278	"Validation and correlation of vidas iggii and igmii lyme antibodies vs vidas lyme igg/igm in the laboratory evaluation of lyme disease: Clinical implications".ti. and "2015".yr. (1)

279	"Method comparison of two elisa platforms for the detection of b.Burgdorferi (lyme) antibodies; c6 specific peptide vs. Whole cell sonicate".ti. and "2013".yr. (1)

280	"Bead based multiplex immunoassays for the laboratory neuroborreliosis".ti. and "2012".yr. (0)

281	"Detection of borrelia burgdorferi in formalin fixed paraffin embedded skin biopsies".ti. and "2012".yr. (1)

282	("Enzyme-linked immunospot assay in the laboratory diagnostics of lyme borreliosis".ti. and "2015".yr.) not "Our experience with".ti. (1)

283	"2-tiered antibody testing for early and late lyme disease using only an immunoglobulin g blot with the addition of a vlse band as the second-tier test".ti. and "2010".yr. (1)

284	"Two-tiered antibody testing for lyme disease with use of 2 enzyme immunoassays, a whole-cell sonicate enzyme immunoassay followed by a vlse c6 peptide enzyme immunoassay".ti. and "2011".yr. (1)

285	"Performance of united states serologic assays in the diagnosis of lyme borreliosis acquired in europe".ti. and "2013".yr. (1)

286	"A comparison of lyme disease serologic test results from 4 laboratories in patients with persistent symptoms after antibiotic treatment".ti. and "2014".yr. (1)

287	"Lyme disease testing by large commercial laboratories in the united states".ti. and "2014".yr. (1)

288	"Lyme disease and human granulocytic anaplasmosis coinfection: Impact of case definition on coinfection rates and illness severity".ti. and "2013".yr. (1)

289	"Higher mrna levels of chemokines and cytokines associated with macrophage activation in erythema migrans skin lesions in patients from the united states than in patients from austria with lyme borreliosis".ti. and "2008".yr. (1)

290	"Antibiotic treatment duration and long-term outcomes of patients with early lyme disease from a lyme disease-hyperendemic area".ti. and "2010".yr. (1)

291	"Poor positive predictive value of lyme disease serologic testing in an area of low disease incidence".ti. and "2015".yr. (1)

292	"Evaluation of the c6 lyme enzyme immunoassay for the diagnosis of lyme disease in children and adolescents".ti. and "2016".yr. (1)

293	"Borrelial lymphocytoma in adult patients".ti. and "2016".yr. (1)

294	"Course and outcome of early lyme borreliosis in patients with hematological malignancies".ti. and "2015".yr. (1)

295	"Xenodiagnosis to detect borrelia burgdorferi infection: A first-in-human study".ti. and "2014".yr. (1)

296	"Development of a metabolic biosignature for detection of early lyme disease".ti. and "2015".yr. (1)

297	"Blood cultures for patients with extracutaneous manifestations of lyme disease in the united states".ti. and "2009".yr. (1)

298	"Suspected early lyme neuroborreliosis in patients with erythema migrans".ti. and "2013".yr. (1)

299	("Course and outcome of early european lyme neuroborreliosis" and "Clinical and laboratory findings").ti. and "2016".yr. (1)

300	"Lyme disease diagnosed by alternative methods: A phenotype similar to that of chronic fatigue syndrome".ti. and "2015".yr. (1)

301	"Prospective study of serologic tests for lyme disease".ti. and "2008".yr. (1)

302	"Comparison of findings for patients with borrelia garinii and borrelia afzelii isolated from cerebrospinal fluid".ti. and "2006".yr. (1)

303	"Treatment of erythema migrans with doxycycline for 10 days versus 15 days".ti. and "2012".yr. (1)

304	"Incidence and patterns of extended-course antibiotic therapy in patients evaluated for lyme disease".ti. and "2015".yr. (1)

305	"Long-term assessment of post-treatment symptoms in patients with culture-confirmed early lyme disease".ti. and "2015".yr. (1)

306	"Long-term follow-up of patients with lyme disease: Longitudinal analysis of clinical and quality-of-life measures".ti. and "2016".yr. (1)

307	"Extensive limb swelling after immunization: Reports to the vaccine adverse event reporting system".ti. and "2003".yr. (1)

308	"Effect of borrelia burgdorferi genotype on the sensitivity of c6 and 2-tier testing in north american patients with culture-confirmed lyme disease".ti. and "2008".yr. (1)

309	"Long-term assessment of health-related quality of life in patients with culture-confirmed early lyme disease".ti. and "2015".yr. and Wormser.au. (0)

310	"Evaluation of a new multiparametric microspot array for serodiagnosis of lyme borreliosis".ti. and "2015".yr. (2)

311	"Lyme disease in the u.K.: Clinical and laboratory features and response to treatment".ti. and "2010".yr. (1)

312	"Comparison of pcr methods and culture for the detection of borrelia spp. In patients with erythema migrans".ti. and "2008".yr. (1)

313	"Seroprevalence of tick-borne infections in forestry rangers from northeastern italy".ti. and "2004".yr. (1)

314	"Ticking the right boxes: Classification of patients suspected of lyme borreliosis at an academic referral center in the netherlands".ti. and "2015".yr. (1)

315	"Anaplasma phagocytophilum infection in patients with early lyme borreliosis, erythema migrans, in north-eastern poland".ti. and "2009".yr. (1)

316	"Neuroborreliosis--an epidemiological, clinical and healthcare cost study from an endemic area in the south-east of sweden".ti. and "2010".yr. (1)

317	"Analysis of borrelia burgdorferi igg antibodies with a combination of igg elisa and vlse c6 peptide elisa".ti. and "2005".yr. (1)

318	"Peripheral facial palsy in patients with tick-borne encephalitis".ti. and "2012".yr. (1)

319	"Testing patients with non-specific symptoms for antibodies against borrelia burgdorferi sensu lato does not provide useful clinical information about their aetiology".ti. and "2015".yr. (1)

320	"Lyme borreliosis caused by diverse genospecies of borrelia burgdorferi sensu lato in northeastern china".ti. and "2014".yr. (1)

321	"Seroprevalence of seven pathogens transmitted by the ixodes ricinus tick in forestry workers in france".ti. and "2016".yr. (1)

322	"Comparison of isolation rate of borrelia burgdorferi sensu lato in two different culture media, mkp and bsk-h".ti. and "2014".yr. (1)

323	"High frequency of false positive igm immunoblots for borrelia burgdorferi in clinical practice".ti. and "2012".yr. (1)

324	"Comparison of five different immunoassays for the detection of borrelia burgdorferi igm and igg antibodies".ti. and "2006".yr. (1)

325	"High sensitivity and specificity of the c6-peptide elisa on cerebrospinal fluid in lyme neuroborreliosis patients".ti. and "2011".yr. (1)

326	"Performance of four commercial immunoblots for serological confirmation of lyme borreliosis in rhone-alpes (france)".ti. and "2009".yr. (1)

327	"Lyme borreliosis: Descriptive study of 151 patients followed up in a university hospital, lyon (france)".ti. and "2009".yr. (1)

328	"Value of the lymphocyte transformation test to determine the acuity of neuroborreliosis".ti. and "2012".yr. (1)

329	"Evaluation of two automated tests on the vidas instrument to detect anti-lyme disease antibodies igg and igm in human serum, plasma and csf".ti. and "2010".yr. (1)

330	"Molecular identification of borrelia spirochetes isolated from skin biopsies of patients characterised with an unusual dermal manifestation (prurigo pigmentosa) in taiwan".ti. and "2012".yr. (1)

331	"Preliminary study on immunological reactivity in people occupationally exposed to tick-transmitted pathogens".ti. and "2009".yr. (1)

332	"Methodology in reporting diagnostic accuracy for borrelia specific igg and igm antibodies. Tests often differ but performance of the test antigens may be quite similar anyway".ti. and "2012".yr. (1)

333	"Direct detection of early lyme borreliosis from whole blood".ti. and "2011".yr. (1)

334	"German investigation of lyme borreliosis: Evaluation of therapeutic and diagnostic cost (gilead)".ti. and "2011".yr. (1)

335	"Detection of borrelia burgdorferi sensu lato in urine specimens from patients with early and late lyme borreliosis".ti. and "2012".yr. (1)

336	"Preferential reactivity towards borrelia afzelii in the sera of patients with lyme borreliosis from the north-east of poland".ti. and "2010".yr. (1)

337	"Combining c6 elisa and igm immunoblot for the detection of antibodies in early lyme infection".ti. and "2010".yr. (1)

338	"Lyme borreliosis and tick-borne encephalitis in latvia: A comparative study of cases reported in 2007-2010".ti. and "2012".yr. (1)

339	"Course and outcome of erythema migrans in patients with underlying rheumatological disease".ti. and "2011".yr. (1)

340	"Neuroborreliosis: Detection of intrathecally produced antibodies against borrelia in cerebrospinal fluid using the line blot immunoassay anti-borrelia euroline-rn-at".ti. and "2011".yr. (1)

341	"Correlation between indirect immunofluorescence and western blot, serological techniques used for the diagnosis of lyme disease".ti. and "2012".yr. (1)

342	"Spectrum of clinical forms and seasonal distribution of lyme borreliosis in children".ti. and "2010".yr. (1)

343	"Co-infection with borrelia burgdorferi and tick-borne encephalitis virus in humans, ticks and tick cells - analysis of clinical cases, literature and experimental possibilities".ti. and "2012".yr. (1)

344	"Performance evaluation of the new vidas lyme igm and igg assays compared to the previous vidas lyme total antibodies assay on fresh prospective sera".ti. and "2012".yr. (1)

345	"Dimeric ospcadv - advanced recombinant ospc antigens for the sensitive and specific detection of class igm antibodies in early stages of borrelia infections".ti. and "2011".yr. (1)

346	"Evaluation of different immunoassays on liaison".ti. and "2009".yr. (1)

347	"Infectious diseases among eufor military personnel".ti. and "2011".yr. (1)

348	"Childhood neuroborreliosis does not lead to long-term cognitive disturbances".ti. and "2011".yr. (1)

349	"Patients after proven neuroborreliosis - how severe is the persisting neuropsychological damage?".ti. and "2009".yr. (1)

350	"Neuroborreliosis in bulgaria - clinical manifestation and serological findings".ti. and "2011".yr. (1)

351	"Clinical manifestations of lyme borreliosis in bulgaria - comparison with tick studies".ti. and "2010".yr. (1)

352	"Evaluation of the new vidas lyme igm and vidas lyme igg kits as screening test for the serological diagnosis of lyme borreliosis".ti. and "2012".yr. (1)

353	"Evaluation of two line immunoassays for the detection of borrelia antibodies".ti. and "2009".yr. (1)

354	"Attention and learning in children with lyme disease and children with attention deficit hyperactivity disorder".ti. and "2013".yr. (0)

355	"Personality function in chronic lyme disease".ti. and "2008".yr. (0)

356	"Spect brain imaging in chronic lyme disease".ti. and "2012".yr. (1)

357	"Lyme arthritis in 20 children residing in a non-endemic area".ti. and "2005".yr. (1)

358	"Prolonged synovitis in pediatric lyme arthritis cannot be predicted by clinical or laboratory parameters".ti. and "2009".yr. (1)

359	"Effect of latitude on the rate of change in incidence of lyme disease in the united states".ti. and "2013".yr. (0)

360	"Treatment for the neurological complications of lyme disease".ti. and "2008".yr. (1)

361	"Antibiotics for the neurological complications of lyme disease".ti. and "2016".yr. (1)

362	"A comparison of analysis units for associating lyme disease with forest-edge habitat".ti. and "2006".yr. (0)

363	"Serologically detected new tick-borne zoonoses in eastern croatia".ti. and "2003".yr. (1)

364	"Systematic analysis of wide-field fundus autofluorescence (faf) imaging in posterior uveitis".ti. and "2014".yr. (1)

365	"Season is an unreliable predictor of lyme neuroborreliosis".ti. and "2015".yr. (1)

366	"Lyme disease trends: Delaware, 2000 - 2004".ti. and "2007".yr. (1)

367	"Comparison of a two- or three-week regimen and a review of treatment of erythema migrans with phenoxymethylpenicillin".ti. and "2006".yr. (1)

368	"Mini erythema migrans--a sign of early lyme borreliosis".ti. and "2006".yr. (1)

369	"Gabapentin for the symptomatic treatment of chronic neuropathic pain in patients with late-stage lyme borreliosis: A pilot study".ti. and "2005".yr. (1)

370	"Bull's-eye and nontarget skin lesions of lyme disease: An internet survey of identification of erythema migrans".ti. and "2012".yr. (1)

371	"Utility of a commercial immunoblot kit (bag-borrelia blot) in the diagnosis of the preliminary stages of lyme disease".ti. and "2003".yr. (1)

372	"A dutch nationwide evaluation of serological assays for detection of borrelia antibodies in clinically well-defined patients".ti. and "2015".yr. (1)

373	"Percent positive rate of lyme real-time polymerase chain reaction in blood, cerebrospinal fluid, synovial fluid, and tissue".ti. and "2008".yr. (1)

374	"Evaluation of commercial screening tests and blot assays for the diagnosis of lyme borreliosis".ti. and "2012".yr. (1)

375	"Presence of anti-borrelia burgdorferi antibodies and borrelia burgdorferi sensu lato dna in samples of subjects in an area of the northern italy in the period 2002-2008".ti. and "2011".yr. (1)

376	"Antigen biochips verify and extend the scope of antibody detection in lyme borreliosis".ti. and "2007".yr. (1)

377	"A lightcycler taqman assay for detection of borrelia burgdorferi sensu lato in clinical samples".ti. and "2007".yr. (1)

378	"Comparison of five diagnostic modalities for direct detection of borrelia burgdorferi in patients with early lyme disease".ti. and "2012".yr. (1)

379	"Pregnant women with erythema migrans and isolation of borreliae from blood: Course and outcome after treatment with ceftriaxone".ti. and "2011".yr. (1)

380	"Cross-reactivity between lyme and syphilis screening assays: Lyme disease does not cause false-positive syphilis screens".ti. and "2016".yr. (1)

381	"The c6 lyme antibody test has low sensitivity for antibody detection in cerebrospinal fluid".ti. and "2009".yr. (1)

382	"Comparison of males versus females with culture-confirmed early lyme disease at presentation and at 11-20 years after diagnosis".ti. and "2016".yr. (1)

383	"Single-tier testing with the c6 peptide elisa kit compared with two-tier testing for lyme disease".ti. and "2013".yr. (1)

384	"Tick-borne diseases: An evaluation of a lyme disease prevention education program for eight, nine, and ten year olds".ti. and "2009".yr. (0)

385	"Understanding lyme disease: Illness experience, prevention, and the health belief model".ti. and "2008".yr. (0)

386	"Quaternary lyme disease: Symptomatology, epidemiology and anthropology of an emerging disease".ti. and "2015".yr. (0)

387	"Opportunities and barriers in the control and prevention of lyme disease: Implications for practice".ti. and "2015".yr. (0)

388	"A comparison of memory and executive functioning in children with lyme disease and children with attention-deficit/hyperactivity disorder".ti. and "2014".yr. (0)

389	"A national survey of risk perceptions and practices to prevent tick-borne lyme disease and mosquito-borne viral encephalitis".ti. and "2003".yr. (0)

390	"Spatial analysis of human lyme disease risk in an endemic county".ti. and "2015".yr. (0)

391	"Memory and executive functions in adolescents with lyme disease".ti. and "2007".yr. (0)

392	"Personality traits, perceived stress, and coping styles in patients with chronic lyme disease and fibromyalgia".ti. and "2009".yr. (0)

393	"Coping, depression, and quality of life of individuals with chronic lyme disease".ti. and "2015".yr. (0)

394	"Audiologic manifestations of patients with post-treatment lyme disease syndrome".ti. and "2003".yr. (1)

395	"Disease risk in a dynamic environment: The spread of tick-borne pathogens in minnesota, usa".ti. and "2015".yr. (1)

396	"An examination of the demographic and environmental variables correlated with lyme disease emergence in virginia".ti. and "2015".yr. (1)

397	"Effects of landscape fragmentation and climate on lyme disease incidence in the northeastern united states".ti. and "2013".yr. (1)

398	"Increasing residential proximity of lyme borreliosis cases to high-risk habitats: A retrospective study in central bohemia, the czech republic, 1987-2010".ti. and "2015".yr. (1)

399	"Mapping lyme disease incidence for diagnostic and preventive decisions, maryland".ti. and "2002".yr. (1)

400	"Infections with candidatus neoehrlichia mikurensis and cytokine responses in 2 persons bitten by ticks, sweden".ti. and "2015".yr. (1)

401	"Lyme disease, virginia, usa, 2000-2011".ti. and "2014".yr. (1)

402	"Hypersensitivity to ticks and lyme disease risk".ti. and "2005".yr. (1)

403	"Enhancing lyme disease surveillance by using administrative claims data, tennessee, usa".ti. and "2015".yr. (1)

404	"Effect of surveillance method on reported characteristics of lyme disease, connecticut, 1996-2007".ti. and "2012".yr. (1)

405	"No geographic correlation between lyme disease and death due to 4 neurodegenerative disorders, united states, 2001-2010".ti. and "2015".yr. (1)

406	"Epidemiology of lyme disease, nova scotia, canada, 2002-2013".ti. and "2015".yr. (1)

407	"Borrelia miyamotoi sensu lato seroreactivity and seroprevalence in the northeastern united states".ti. and "2014".yr. (1)

408	"Geographic distribution and expansion of human lyme disease, united states".ti. and "2015".yr. (1)

409	"Bartonella spp. Bacteremia and rheumatic symptoms in patients from lyme disease-endemic region".ti. and "2012".yr. and Maggi.au. (0)

410	"Incidence of clinician-diagnosed lyme disease, united states, 2005-2010".ti. and "2015".yr. (1)

411	"Lyme disease in hispanics, united states, 2000-2013".ti. and "2016".yr. (1)

412	"Human babesiosis, maine, usa, 1995-2011".ti. and "2014".yr. (1)

413	"Effectiveness of personal protective measures to prevent lyme disease".ti. and "2008".yr. (1)

414	"Antibodies against borrelia burgdorferi sensu lato among adults, germany, 2008-2011".ti. and "2015".yr. (1)

415	"Economic impact of lyme disease".ti. and "2006".yr. (1)

416	"Spatio-temporal patterns in county-level incidence and reporting of lyme disease in the northeastern united states, 1990-2000".ti. and "2007".yr. (0)

417	"Temporal and spatial distribution of tick-borne disease cases among humans and canines in illinois (2000-2009)".ti. and "2014".yr. (0)

418	"Validating negative binomial lyme disease regression model with bootstrap resampling".ti. and "2016".yr. (0)

419	"Variability in results from negative binomial models for lyme disease measured at different spatial scales".ti. and "2015".yr. (1)

420	"Lyme borreliosis--epidemiological analysis of incidence in the northern region of slovakia".ti. and "2012".yr. (1)

421	"Improvement of diagnostic approach to lyme neuroborreliosis in children by using recombinant antigens in detection of intrathecally produced igm/igg".ti. and "2016".yr. (1)

422	"Synoptic spatial classification and lyme disease in new york state (1991-2006)".ti. and "2012".yr. (1)

423	"Lyme borreliosis in southern united kingdom and a case for a new syndrome, chronic arthropod-borne neuropathy".ti. and "2015".yr. (1)

424	"Lyme disease in france: A primary care-based prospective study".ti. and "2005".yr. (1)

425	"Neuroborreliosis in the south west of england".ti. and "2008".yr. (1)

426	"Tick-borne encephalitis and lyme borreliosis in belarus republic, 1998-2007".ti. and "2008".yr. (0)

427	"Epidemiology of tick-borne encephalitis and lyme disease in the republic of belarus, 1998-2007".ti. and "2009".yr. (0)

428	"Epidemiological situation for tick-borne encephalitis and lyme disease in the republic of karelia, russia, in 2002-2011".ti. and "2012".yr. (0)

429	"Incidence of tick-borne encephalitis and lyme disease in leninggrad oblast".ti. and "2012".yr. (0)

430	"Serological evidence for human granulocytic ehrlichiosis in western austria".ti. and "2003".yr. (1)

431	"The phenomenon of chronic lyme ; an observational study".ti. and "2012".yr. (1)

432	"Clinical data and cerebrospinal fluid findings in lyme meningitis versus aseptic meningitis".ti. and "2003".yr. (1)

433	"Improving national surveillance of lyme neuroborreliosis in denmark through electronic reporting of specific antibody index testing from 2010 to 2012".ti. and "2015".yr. (3)

434	"Lyme borreliosis in the netherlands: Strong increase in gp consultations and hospital admissions in past 10 years".ti. and "2006".yr. (1)

435	"Laboratory diagnosis of lyme borreliosis at the portuguese national institute of health (1990-2004)".ti. and "2006".yr. (1)

436	["Surveillance of lyme borreliosis in germany, 2002 and 2003".ti. and "2005".yr. and Maggi.ax.] (0)

437	"Disseminated and chronic lyme borreliosis in norway, 1995 - 2004".ti. and "2005".yr. and borreliosis.kw. (0)

438	"Lyme borreliosis: Europe-wide coordinated surveillance and action needed?".ti. and "2006".yr. (1)

439	"Incidence and hospitalisation rates of lyme borreliosis, france, 2004 to 2012".ti. and "2014".yr. (1)

440	"Lv and rv function in patients with lyme disease diagnosed with endomyocardial biopsy-a long term follow-up".ti. and "2012".yr. (1)

441	"Is improvement of the left ventricular systolic function in patients with new-onset dilated cardiomyopathy related to the presence of virus or inflammation in endomycardial biopsy?".ti. and "2012".yr. (1)

442	"Has serological testing of antibodies to cardiotropic agents any role in the diagnosis of inflammatory cardiomyopathy?".ti. and "2010".yr. (1)

443	("Large differences between test strategies for the detection of anti-borrelia antibodies are revealed by comparing eight elisas and five immunoblots".ti. and "2011".yr.) not letter.pt. (1)

444	"Increased incidence of lyme borreliosis in southern sweden following mild winters and during warm, humid summers".ti. and "2006".yr. (1)

445	"Borrelia burgdorferi sensu lato serology in the netherlands: Guidelines versus daily practice".ti. and "2014".yr. (1)

446	"Risk of acquiring tick-borne infections in forestry workers from lazio, italy".ti. and "2010".yr. (1)

447	"Management of paediatric lyme disease in non-endemic and endemic areas: Data from the registry of the italian society for pediatric infectious diseases".ti. and "2013".yr. (1)

448	"Pre-treatment and post-treatment assessment of the c(6) test in patients with persistent symptoms and a history of lyme borreliosis".ti. and "2004".yr. (0)

449	"Laboratory diagnosis of lyme neuroborreliosis: A comparison of three csf anti-borrelia antibody assays".ti. and "2014".yr. (1)

450	"Seroreactivity for spotted fever rickettsiae and co-infections with other tick-borne agents among habitants in central and southern sweden".ti. and "2013".yr. (1)

451	"Borrelia burgdorferi vlse antigen for the serological diagnosis of lyme borreliosis".ti. and "2008".yr. (1)

452	"Co-infections with borrelia species, anaplasma phagocytophilum and babesia spp. In patients with tick-borne encephalitis".ti. and "2014".yr. (1)

453	"Risk of borrelia burgdorferi infection in western switzerland following a tick bite".ti. and "2004".yr. (1)

454	"Duration of antibiotic treatment in disseminated lyme borreliosis: A double-blind, randomized, placebo-controlled, multicenter clinical study".ti. and "2007".yr. (1)

455	"Comparison of an automated borrelia indirect chemiluminescent immunoassay (clia) with a vlse/c6 elisa and immunoblot".ti. and "2007".yr. (1)

456	"Lyme meningoradiculitis: Prospective evaluation of biological diagnosis methods".ti. and "2007".yr. (1)

457	"Differential diagnoses of suspected lyme borreliosis or post-lyme-disease syndrome".ti. and "2007".yr. (1)

458	"Sensitivity and specificity of a commercial c6 peptide enzyme immuno assay in diagnosis of acute lyme neuroborreliosis".ti. and "2007".yr. (1)

459	"Evaluation of two commercially available rapid diagnostic tests for lyme borreliosis".ti. and "2015".yr. (1)

460	"C6 peptide elisa test in the serodiagnosis of lyme borreliosis in sweden".ti. and "2007".yr. (1)

461	"Prevalence and incidence of antibodies to borrelia burgdorferi and to tick-borne encephalitis virus in agricultural and forestry workers from tuscany, italy".ti. and "2005".yr. (1)

462	"Incidence and management of presumption of lyme borreliosis in belgium: Recent data from the sentinel network of general practitioners".ti. and "2012".yr. (1)

463	"The improvement of left ventricular systolic function induced by antibiotic therapy in patients with inflammatory cardiomyopathy caused by borrelia burgdorferi infection".ti. and "2010".yr. (1)

464	"Hypothalamic-pituitary insufficiency following infectious diseases of the central nervous system".ti. and "2008".yr. (1)

465	"North atlantic weather oscillation and human infectious diseases in the czech republic, 1951-2003".ti. and "2005".yr. (1)

466	"Screening of the prevalence of antibodies to borrelia burgdorferi in madrid province, spain".ti. and "2004".yr. (1)

467	"Detection of borrelia burgdorferi sensu lato in endomyocardial biopsy specimens in individuals with recent-onset dilated cardiomyopathy".ti. and "2012".yr. (1)

468	"Comprehensive assessment of endomyocardial biopsy specimens in patienswith recent-onset dilated cardiomyopathy".ti. and "2011".yr. (1)

469	"Borrelia burgdorferi infection in iranian patients with multiple sclerosis".ti. and "2009".yr. (1)

470	"Clinical effects of fluconazole in patients with neuroborreliosis".ti. and "2004".yr. (1)

471	"Parkinson's syndrome in chronic neuroborreliosis".ti. and "2012".yr. (1)

472	"Oral doxycycline for lyme neuroborreliosis with symptoms of encephalitis, myelitis, vasculitis or intracranial hypertension".ti. and "2014".yr. (1)

473	"A study on the association between infectious burden and alzheimer's disease".ti. and "2015".yr. (1)

474	"Borreliosis. Cause of obstructive sleep apnoea syndrome".ti. and "2008".yr. (0)

475	"The prevalence of neuropsychiatric symptoms in lyme borreliosis patients".ti. and "2015".yr. (1)

476	"Efficacy and safety of pharmacological treatments for acute lyme neuroborreliosis - a systematic review".ti. and "2015".yr. (1)

477	"The diagnostic spectrum in patients with suspected chronic lyme neuroborreliosis--the experience from one year of a university hospital's lyme neuroborreliosis outpatients clinic".ti. and "2011".yr. (1)

478	"European neuroborreliosis: Neuropsychological findings 30 months post-treatment".ti. and "2012".yr. (0)

479	"Clinical and immunological characteristics of multiple sclerosis patients with b.Burgdorferi antibodies".ti. and "2008".yr. (0)

480	"Clinical and serological features of borrelial infection in multiple sclerosis patients".ti. and "2010".yr. (1)

481	"Antibodies against ebv, cmv, vzv, hsv-1, hsv-2, hhv-6b and borrelia burgdorferi and viral nucleic acids in serum and csf samples of patients with multiple sclerosis".ti. and "2012".yr. (1)

482	"Borreliosis and associated autoimmunity".ti. and "2016".yr. (1)

483	"Oral doxycycline vs. Iv centriaxone for european lume neuro-borreliosis. A double-blind, randomized controlled clinical trial".ti. and "2008".yr. (0)

484	"Remaining complaints 1 year after treatment for acute lyme neuroborreliosis; frequency, pattern and risk factors".ti. and "2010".yr. (1)

485	"Clinical usefulness of intrathecal antibody testing in acute lyme neuroborreliosis".ti. and "2007".yr. (1)

486	"Borrelia burgdorferi: Risk factor in alzheimer's disease".ti. and "2011".yr. (1)

487	"Borrelia burgdorferi: Risk factor in alzheimer's disease p1021.[abstract]".ti. and "2012".yr. (0)

488	"Chronic polyneuropathy and lyme disease".ti. and "2006".yr. (1)

489	"Antibiotic therapy in patients with the combination of definite multiple sclerosis and chronic lyme disease".ti. and "2012".yr. (1)

490	"Peripheral facial palsy associated with viral infections in children".ti. and "2011".yr. (1)

491	"Outcome of peripheral facial palsy in children - a catamnestic study".ti. and "2013".yr. (1)

492	"Canalicular magnetic stimulation lacks specificity to differentiate idiopathic facial palsy from borreliosis in children".ti. and "2008".yr. (1)

493	"Facial palsy: Etiology, outcome and management in children".ti. and "2011".yr. (1)

494	"Uncommon synovial cysts in children".ti. and "2006".yr. (1)

495	"Pediatric tick-borne infections of the central nervous system in an endemic region of sweden: A prospective evaluation of clinical manifestations".ti. and "2012".yr. (0)

496	"Tick bites, erythema migrans and lyme borreliosis: Incidence and management by the gp in belgium".ti. and "2004".yr. (0)

497	"The burden of lyme borreliosis expressed in disability-adjusted life years".ti. and "2015".yr. (1)

498	"Cognitive functioning in lyme borreliosis".ti. and "2004".yr. (0)

499	"Mental disorders in patients with lyme disease".ti. and "2003".yr. (0)

500	"Clinical and demographic characteristics of psychiatric patients seropositive for borrelia burgdorferi".ti. and "2006".yr. (1)

501	("Two great masqueraders" and "in a psychiatry clinic-case series of neurosyphilis and neuroborreliosis").ti. and "2012".yr. (1)

502	"Analysis of weather effects on variability in lyme disease incidence in the northeastern united states".ti. and "2002".yr. (1)

503	"Evaluation of clinical specimens for rickettsia, bartonella, borrelia, coxiella, anaplasma, franciscella and diplorickettsia positivity using serological and molecular biology methods".ti. and "2012".yr. (0)

504	"Detection of borrelia burgdorferi sensu lato dna by pcr in serum of patients with clinical symptoms of lyme borreliosis".ti. and "2008".yr. (1)

505	"Cerebrospinal-fluid profile in neuroborreliosis and its diagnostic significance".ti. and "2006".yr. (1)

506	"Short-term stability of borrelia garinii in cerebrospinal fluid".ti. and "2016".yr. (1)

507	"Dna persistence after treatment of lyme borreliosis".ti. and "2014".yr. (1)

508	"Lyme borreliosis--analysis of the trends in slovakia, 1999-2008".ti. and "2011".yr. (1)

509	"Lipid peroxidation in the pathogenesis of neuroborreliosis".ti. and "2016".yr. (1)

510	"Sex differences in the clinical and serologic presentation of early lyme disease: Results from a retrospective review".ti. and "2010".yr. (1)

511	"A determination of the spatial concordance between lyme disease incidence and habitat probability of its primary vector ixodes scapularis (black-legged tick)".ti. and "2014".yr. (1)

512	"Diluting the dilution effect: A spatial lyme model provides evidence for the importance of habitat fragmentation with regard to the risk of infection".ti. and "2009".yr. (1)

513	"Molecular survey of five tick-borne pathogens (ehrlichia chaffeensis, ehrlichia ewingii, anaplasma phagocytophilum, borrelia burgdorferi sensu lato and babesia microti) in egyptian farmers".ti. and "2011".yr. (0)

514	"Recurrent early pregnancy loss, intrauterine death-causes, diagnosis and therapeutical approach".ti. and "2012".yr. (1)

515	"Making sense of unfamiliar risks in the countryside: The case of lyme disease".ti. and "2011".yr. (1)

516	"Whose lyme is it anyway? Subject positions and the construction of responsibility for managing the health risks from lyme disease".ti. and "2012".yr. (1)

517	"A controlled trial of a novel primary prevention program for lyme disease and other tick-borne illnesses".ti. and "2007".yr. (1)

518	"Developing requirements for a mobile app to support citizens in dealing with ticks and tick bites via end-user profiling".ti. and "2015".yr. (1)

519	"Healthcare access and burden of care for patients with lyme disease: A large united states survey".ti. and "2011".yr. (1)

520	"Increasing burden of lyme carditis in us children's hospitals".ti. and "2015".yr. (1)

521	"Suspected inflammatory cardiomyopathy. Prevalence of borrelia burgdorferi in endomyocardial biopsies with positive serological evidence".ti. and "2015".yr. (1)

522	"Recent-onset dilated cardiomyopathy associated with borrelia burgdorferi infection".ti. and "2015".yr. (1)

523	"Detection of spirochaetal microorganisms by focus floating microscopy in necrobiosis lipoidica in patients from central europe".ti. and "2008".yr. (1)

524	"Lyme borreliosis: Differences between the patients who are tested and those with the infection".ti. and "2008".yr. (0)

525	"Fabric impregnation using acaricides: Effective and safe method for the prevention of tick-infestation and tick-borne diseases".ti. and "2008".yr. (0)

526	"Etiology of tick-borne febrile illnesses in adult residents of north-eastern poland: Report from a prospective clinical study".ti. and "2006".yr. (1)

527	"Disappearance of specific immune response after successful therapy of chronic lyme borreliosis".ti. and "2004".yr. (1)

528	"Serological description of estonian patients with lyme disease, a comparison with control sera from endemic and non-endemic areas".ti. and "2004".yr. (1)

529	"Comparison of different borrelia burgdorferi sensu lato strains for detection of immune response in patients with erythema migrans".ti. and "2008".yr. (1)

530	"Significant improvement of the recombinant borrelia igg immunoblot for serodiagnosis of early neuroborreliosis".ti. and "2004".yr. (1)

531	"Immune responses to borrelial vlse ir6 peptide variants".ti. and "2007".yr. (1)

532	"Antibody responses to borrelia ir(6) peptide variants and the c6 peptide in swedish patients with erythema migrans".ti. and "2009".yr. (0)

533	"Serological evidence for tick-borne encephalitis, borreliosis, and human granulocytic anaplasmosis in mongolia".ti. and "2006".yr. (1)

534	"Lyme borreliosis reinfection: Might it be explained by a gender difference in immune response?".ti. and "2006".yr. (1)

535	"Could co-infection with anaplasma play a role in borrelia-associated primary cutaneous marginal zone b-cell lymphomas?".ti. and "2016".yr. (1)

536	"Factors affecting duration of chronic fatigue syndrome in pediatric patients".ti. and "2012".yr. (1)

537	"Validate or falsify: Lessons learned from a microscopy method claimed to be useful for detecting borrelia and babesia organisms in human blood".ti. and "2016".yr. (1)

538	"Solitary and multiple erythema migrans in children: Comparison of demographic, clinical and laboratory findings".ti. and "2003".yr. (1)

539	"Borrelia antibodies in children evaluated for lyme neuroborreliosis".ti. and "2008".yr. (1)

540	"Oral antibiotic treatment and long-term outcomes of lyme facial nerve palsy".ti. and "2011".yr. (1)

541	"Comparison of erythema migrans caused by borrelia afzelii and borrelia garinii".ti. and "2004".yr. (1)

542	"Isolation of borrelia burgdorferi sensu lato from blood of adult patients with borrelial lymphocytoma, lyme neuroborreliosis, lyme arthritis and acrodermatitis chronica atrophicans".ti. and "2011".yr. (1)

543	"Comparative analysis of the infectivity rate of both borrelia burgdorferi and anaplasma phagocytophilum in humans and dogs in a new jersey community".ti. and "2014".yr. (1)

544	"Evidence for strain-specific immunity in patients treated for early lyme disease".ti. and "2014".yr. (1)

545	"Lyme borreliosis: A european perspective on diagnosis and clinical management".ti. and "2009".yr. (1)

546	"The positive predictive value of borrelia burgdorferi serology in the light of symptoms of patients sent to an outpatient service for tick-borne diseases".ti. and "2010".yr. (1)

547	"Commercial test kits for detection of lyme borreliosis: A meta-analysis of test accuracy".ti. and "2016".yr. (1)

548	"Risk factors in lyme borreliosis: A german case-control study".ti. and "2002".yr. (1)

549	"Seroprevalence of infection with borrelia burgdorferi s. L. In two adjacent regions of eastern france and southwestern germany".ti. and "2002".yr. (1)

550	"Foci of tick-borne diseases in southwest germany".ti. and "2002".yr. (1)

551	"Seroprevalence of borrelia infection in occupational tick-exposed people in bavaria (germany)".ti. and "2002".yr. (1)

552	"Seroprevalence of antibodies against borrelia burgdorferi s. L. And tick-borne encephalitis virus in the bergische land region of northrhine westfalia".ti. and "2002".yr. (1)

553	"Acute facial palsy in children--a 2-year follow-up study with focus on lyme neuroborreliosis".ti. and "2003".yr. (1)

554	"Project management of lyme disease through monitoring of seroprevalence of anti-borrelia antibodies in macedonian patients".ti. and "2014".yr. (0)

555	"Retrospective study of lyme borreliosis positive serologies in 2003".ti. and "2004".yr. (0)

556	"Evaluation of plateliatm lyme igm and plateliatm lyme igg assays for lyme borreliosis diagnosis".ti. and "2007".yr. (0)

557	"Comparison between enzygnost((r)) lyme link vlse/igg and liaison((r)) borrelia igg for the laboratory diagnosis of lyme disease".ti. and "2007".yr. (0)

558	"Evaluation of five commercial screening assays and two commercial immunoblots for the serological diagnosis of lyme borreliosis".ti. and "2007".yr. (0)

559	"Trends in blacklegged tick submissions and lyme disease serology at ontario public health laboratories".ti. and "2009".yr. (1)

560	"Neuroborreliosis and tick-borne encephalitis - same vector, but different clinical course".ti. and "2007".yr. (0)

561	"Usefulness of c6 peptide antibody assay in early localised lyme borreliosis".ti. and "2007".yr. (0)

562	"The level of morbidity of lyme borreliosis after antibiotic preventive treatment".ti. and "2007".yr. (0)

563	"Seroprevalence of lyme disease and associated risk factors in rural population of beijing".ti. and "2015".yr. (1)

564	"Diagnostic use of the lymphocyte transformation test-memory lymphocyte immunostimulation assay in confirming active lyme borreliosis in clinically and serologically ambiguous cases".ti. and "2014".yr. (1)

565	"Test of 259 serums from patients with arthritis or neurological symptoms confirmed existence of lyme disease in hainan province, china".ti. and "2015".yr. (1)

566	"Examination of specific dna by pcr in patients with different forms of lyme borreliosis".ti. and "2008".yr. (1)

567	"Antibody reactivity to borrelia burgdorferi sensu stricto antigens in patients from the brazilian amazon region with skin diseases not related to lyme disease".ti. and "2010".yr. (1)

568	"Presence of borrelia burgdorferi sensu lato in patients with morphea from the amazonic region in brazil".ti. and "2011".yr. (0)

569	"Relating land cover and spatial distribution of nephropathia epidemica and lyme borreliosis in belgium".ti. and "2013".yr. (1)

570	"The paradox of the binomial ixodes ricinus activity and the observed unimodal lyme borreliosis season in hungary".ti. and "2014".yr. (1)

571	"A seventeen-year epidemiological surveillance study of borrelia burgdorferi infections in two provinces of northern spain".ti. and "2014".yr. (1)

572	"Gis and remote sensing use in the exploration of lyme disease epidemiology".ti. and "2015".yr. (1)

573	"Towards landscape design guidelines for reducing lyme disease risk".ti. and "2006".yr. (1)

574	"Relevance of chronic lyme disease to family medicine as a complex multidimensional chronic disease construct: A systematic review".ti. and "2014".yr. (0)

575	"Emerging incidence of lyme borreliosis, babesiosis, bartonellosis, and granulocytic ehrlichiosis in australia".ti. and "2011".yr. (0)

576	"Clinical determinants of lyme borreliosis, babesiosis, bartonellosis, anaplasmosis, and ehrlichiosis in an australian cohort".ti. and "2015".yr. (0)

577	"Benefit of intravenous antibiotic therapy in patients referred for treatment of neurologic lyme disease".ti. and "2011".yr. (0)

578	"Determinants of the geographic distribution of puumala virus and lyme borreliosis infections in belgium".ti. and "2007".yr. (1)

579	"Enzyme-linked immunosorbent assay, immunofluorescent assay, and recombinant immunoblotting in the serodiagnosis of early lyme borreliosis".ti. and "2003".yr. (0)

580	"Multicentric study of seroprevalence of borrelia burgdorferi and anaplasma phagocytophila in high-risk groups in regions of central and southern italy".ti. and "2004".yr. (1)

581	"Prevalence of igg antibodies against borrelia burgdorferi s.L. And ehrlichia phagocytophila in sera of patients presenting symptoms of lyme disease in a central region of italy".ti. and "2002".yr. (1)

582	"Detection of different borrelia burgdorferi genospecies in serum of people with different occupational risks: Short report".ti. and "2009".yr. (1)

583	"Diagnostic evaluation of human babesiosis in montenegro".ti. and "2012".yr. (1)

584	"Development of a foundation for a case definition of post-treatment lyme disease syndrome".ti. and "2013".yr. (1)

585	"Ultrasonographic evaluation of knee joints in patients with lyme disease".ti. and "2012".yr. (0)

586	"Serological diagnosis of lyme disease in valencia (spain)".ti. and "2010".yr. (1)

587	"Low risk of developing borrelia burgdorferi infection in the south-east of sweden after being bitten by a borrelia burgdorferi-infected tick".ti. and "2011".yr. (1)

588	"Epidemiological, clinical and zoonotic evidences for the existence of lyme disease in central of mexico".ti. and "2010".yr. (1)

589	"Maternal lyme borreliosis and pregnancy outcome".ti. and "2010".yr. (1)

590	"Clinical aspects of neuroborreliosis in bydgoszcz, poland".ti. and "2010".yr. (1)

591	"Antibodies to decorin-binding protein b (dbpb) in the diagnosis of lyme neuroborreliosis in children".ti. and "2014".yr. (1)

592	"Study of lyme borreliosis in ukraine".ti. and "2008".yr. (1)

593	"Comparison of immunofluorescence assay (ifa) and enzyme immuno assay (eia) for borrelial antibody detection in patients with signs and symptoms suggesting neuroborreliosis".ti. and "2006".yr. (0)

594	"The influence of meteorological conditions of the preceding winter on the incidences of tick-borne encephalitis and lyme borreliosis in the czech republic".ti. and "2008".yr. (1)

595	"Diversity of borrelia burgdorferi sensu lato in natural foci of novosibirsk region".ti. and "2008".yr. (1)

596	"A new recombinant igg and igm line immuno blot improves serodiagnosis of lyme borreliosis".ti. and "2004".yr. (0)

597	"Improvement of lyme borreliosis serodiagnosis by a new recombinant igg and igm line immunoblot".ti. and "2006".yr. (0)

598	"Comparison of a new line assay using purified and recombinant antigens with a european lysate blot for serodiagnosis of lyme borreliosis".ti. and "2006".yr. (0)

599	"Serological survey of bartonella spp., borrelia burgdorferi, brucella spp., coxiella burnetii, francisella tularensis, leptospira spp., echinococcus, hanta-,tick-borne encephalitis (tbe)-and xenotropic murine leukemia virus-related virus (xmrv) infection in employees of two forestry enterprises in north rhine-westphalia (nrw), germany, 2011-2013".ti. and "2015".yr. (1)

600	"Lydi-sentinel - a surveillance network on the incidence of lyme disease in bavaria".ti. and "2013".yr. (1)

601	"Serologic evidence of borrelia burgdorferi and anaplasma phagocytophilum infection among patients hospitalized with tick-borne encephalitis (tbe)".ti. and "2006".yr. (0)

602	"Lyme borreliosis: Prevalence of antibodies against non-proteinic (lipid) antigens".ti. and "2008".yr. (0)

603	"Quality of lyme disease serology. Insights and trends from the german proficiency testing program 2000-2005".ti. and "2006".yr. (0)

604	"Clinical assessment of patients with suspected lyme borreliosis".ti. and "2008".yr. (1)

605	"Evaluation of a new lyme borreliosis immunoblot including ebv and syphilis antigens as differential diagnosis tools".ti. and "2009".yr. (1)

606	"Pcr-detection and molecular-genetic analysis of tick-transmitted pathogens in patients of novosibirsk region, russia".ti. and "2008".yr. (1)

607	"The usefulness of 'in vivo' antigens in the diagnosis of human lyme borreliosis".ti. and "2008".yr. (1)

608	"Laboratory diagnosis of early lyme borreliosis - comparison of elisa, western blot (ecoline), and pcr results".ti. and "2006".yr. (0)

609	"Improved culture conditions for the growth and detection of borrelia from human serum".ti. and "2013".yr. (1)

610	"Detection of borreliae in archived sera from patients with clinically suspect lyme disease".ti. and "2014".yr. (1)

611	"A novel isothermal assay of borrelia burgdorferi by recombinase polymerase amplification with lateral flow detection".ti. and "2016".yr. (1)

612	"Psychiatric symptoms in the course of lyme borreliosis and tick borne encephalitis".ti. and "2004".yr. (0)

613	"Knowledge on lyme disease among foresters".ti. and "2011".yr. (1)

614	"Neurological complications of vaccination with outer surface protein a (ospa)".ti. and "2011".yr. (1)

615	"Urinary bladder detrusor dysfunction symptoms in lyme disease".ti. and "2013".yr. (1)

616	"The association of lyme disease with loss of sexual libido and the role of urinary bladder detrusor dysfunction".ti. and "2014".yr. (1)

617	"Absence of borrelia burgdorferi antibodies in the sera of venezuelan patients with localized scleroderma (morphea)".ti. and "2006".yr. (1)

618	"The clinical spectrum of lyme neuroborreliosis".ti. and "2010".yr. (1)

619	"Interstitial granulomatous dermatitis with histiocytic pseudorosettes: A new histopathologic pattern in cutaneous borreliosis. Detection of borrelia burgdorferi dna sequences by a highly sensitive pcr-elisa".ti. and "2003".yr. (1)

620	"Lyme disease knowledge, beliefs, and practices of new hampshire primary care physicians".ti. and "2002".yr. (1)

621	("Lyme arthritis in children presenting with joint effusions".ti. and "2011".yr.) not comment.pt. (2)

622	"Seroprevalence of antibodies that react with anaplasma phagocytophila, the agent of human granulocytic ehrlichiosis, in different populations in westchester county, new york".ti. and "2002".yr. (1)

623	"Importance of sample preparation for molecular diagnosis of lyme borreliosis from urine".ti. and "2002".yr. (1)

624	"Two-year evaluation of borrelia burgdorferi culture and supplemental tests for definitive diagnosis of lyme disease".ti. and "2005".yr. (1)

625	"Recombinant assay for serodiagnosis of lyme disease regardless of ospa vaccination status".ti. and "2002".yr. (1)

626	"Recombinant bbk32 protein in serodiagnosis of early and late lyme borreliosis".ti. and "2002".yr. (1)

627	"Comparison of the borrelia dotblot g, mardx, and vidas enzyme immunoassays for detecting immunoglobulin g antibodies to borrelia burgdorferi in human serum".ti. and "2002".yr. (1)

628	("Assessment of new culture method for detection of borrelia species from serum of lyme disease patients".ti. and "2014".yr.) not erratum.pt. (1)

629	"Quantitative detection of borrelia burgdorferi in 2-millimeter skin samples of erythema migrans lesions: Correlation of results with clinical and laboratory findings".ti. and "2002".yr. (1)

630	"Evaluation of the c6 peptide enzyme-linked immunosorbent assay for individuals vaccinated with the recombinant ospa vaccine".ti. and "2002".yr. (1)

631	"Significant improvement of the recombinant borrelia-specific immunoglobulin g immunoblot test by addition of vlse and a dbpa homologue derived from borrelia garinii for diagnosis of early neuroborreliosis".ti. and "2003".yr. (1)

632	"Varicella zoster and borrelia burgdorferi are the main agents associated with facial paresis, especially in children".ti. and "2003".yr. (1)

633	"Patterns of lyme disease diagnosis and treatment by family physicians in a southeastern state".ti. and "2002".yr. (2)

634	"Role of experience and context in learning to diagnose lyme disease".ti. and "2002".yr. (1)

635	"Specific cutaneous infiltrates of b-cell chronic lymphocytic leukemia (b-cll) at sites typical for borrelia burgdorferi infection".ti. and "2002".yr. (1)

636	"Serodiagnosis of lyme disease by kinetic enzyme-linked immunosorbent assay using recombinant vlse1 or peptide antigens of borrelia burgdorferi compared with 2-tiered testing using whole-cell lysates".ti. and "2003".yr. (1)

637	"Persistence of the antibody response to the vlse sixth invariant region (ir6) peptide of borrelia burgdorferi after successful antibiotic treatment of lyme disease".ti. and "2003".yr. (1)

638	"Cloning of the gene encoding the decorin-binding protein b (dbpb) in borrelia burgdorferi sensu lato and characterisation of the antibody responses to dbpb in lyme borreliosis".ti. and "2002".yr. (1)

639	"Comparative reactivity of human sera to recombinant vlse and other borrelia burgdorferi antigens in class-specific enzyme-linked immunosorbent assays for lyme borreliosis".ti. and "2002".yr. (0)

640	("The chemokine cxcl13 in acute neuroborreliosis".ti. and "2010".yr.) not erratum.pt. (1)

641	"Regional cerebral blood flow and cognitive deficits in chronic lyme disease".ti. and "2003".yr. (1)

642	"Lyme arthritis presenting as acute septic arthritis in children".ti. and "2003".yr. (1)

643	"Infections preceding early arthritis in southern sweden: A prospective population-based study".ti. and "2003".yr. (1)

644	"Serologic markers of lyme disease in children with autism".ti. and "2013".yr. (1)

645	"Epidemiological features and clinical manifestations of lyme borreliosis in korea during the period 2005-2012".ti. and "2015".yr. (1)

646	"Using risk group profiles as a lightweight qualitative approach for intervention development: An example of prevention of tick bites and lyme disease".ti. and "2013".yr. (0)

647	"Diagnostic usefulness of routine lyme serology in patients with early inflammatory arthritis in nonendemic areas".ti. and "2003".yr. (1)

648	"Good outcomes of lyme arthritis in 24 patients in an endemic area of switzerland".ti. and "2004".yr. (1)

649	"Igm deficiency: A retrospective chart review of clinical and immunologic features".ti. and "2012".yr. (1)

650	"Lyme neuroborreliosis and dementia".ti. and "2014".yr. (1)

651	("Efficacy of antibiotic prophylaxis for the prevention of lyme disease: An updated systematic review and meta-analysis".ti. and "2010".yr.) not comment.pt. not letter.pt. (1)

652	"Beekeepers in central europe are at high risk for contracting lyme borreliosis".ti. and "2012".yr. (0)

653	"Potential uses of modafinil in psychiatric disorders".ti. and "2004".yr. (1)

654	("Predictive factors for differentiating between septic arthritis and lyme disease of the knee in children".ti. and "2016".yr.) not comment.pt. (1)

655	"Lyme arthritis in children presenting with joint effusions.[erratum appears in j bone joint surg am. 2011 feb;93(3):E11]".ti. and "2011".yr. (0)

656	"Lyme arthritis of the pediatric lower extremity in the setting of polyarticular disease".ti. and "2014".yr. (0)

657	"Information value of detection of borrelia antibodies in the healthy blood donors and in a population at risk".ti. and "2007".yr. (0)

658	"Chronic fatigue syndrome patients subsequently diagnosed with lyme disease borrelia burgdorferi: Evidence for mycoplasma species coinfections".ti. and "2008".yr. (1)

659	"Evaluation of two commercial systems for automated processing, reading, and interpretation of lyme borreliosis western blots".ti. and "2008".yr. (1)

660	"Validation of cultivation and pcr methods for diagnosis of lyme neuroborreliosis".ti. and "2008".yr. (1)

661	"Lyme disease in oregon".ti. and "2008".yr. (1)

662	"Improvement of lyme borreliosis serodiagnosis by a newly developed recombinant immunoglobulin g (igg) and igm line immunoblot assay and addition of vlse and dbpa homologues".ti. and "2005".yr. (1)

663	"Development of a multiantigen panel for improved detection of borrelia burgdorferi infection in early lyme disease".ti. and "2015".yr. (1)

664	"Collection and characterization of samples for establishment of a serum repository for lyme disease diagnostic test development and evaluation".ti. and "2014".yr. (1)

665	"Discriminating lyme neuroborreliosis from other neuroinflammatory diseases by levels of cxcl13 in cerebrospinal fluid".ti. and "2011".yr. (1)

666	"Differences and similarities between culture-confirmed human granulocytic anaplasmosis and early lyme disease".ti. and "2013".yr. (1)

667	"Comparative cost-effectiveness of two-tiered testing strategies for serodiagnosis of lyme disease with noncutaneous manifestations".ti. and "2013".yr. (1)

668	"Motion-onset and pattern-reversal visual evoked potentials in diagnostics of neuroborreliosis".ti. and "2006".yr. (1)

669	"Pattern and motion-related visual-evoked potentials in neuroborreliosis: Follow-up study".ti. and "2012".yr. (1)

670	"More specific bands in the igg western blot in sera from scottish patients with suspected lyme borreliosis".ti. and "2010".yr. (1)

671	"Development of real time pcr to detect toxoplasma gondii and borrelia burgdorferi infections in postal samples".ti. and "2008".yr. (1)

672	"Local borrelia burgdorferi sensu stricto and borrelia afzelii strains in a single mixed antigen improves western blot sensitivity".ti. and "2009".yr. (1)

673	"Provider knowledge, attitudes, and practices regarding lyme disease in arkansas".ti. and "2015".yr. (1)

674	"Provider knowledge, attitudes, and practices regarding lyme disease in arkansas".ti. and "2015".yr. (1)

675	"Borrelia burgdorferi-associated lymphocytoma cutis: Clinicopathologic, immunophenotypic, and molecular study of 106 cases".ti. and "2004".yr. (1)

676	"Granuloma annulare--a manifestation of infection with borrelia".ti. and "2008".yr. (1)

677	"Borrelia in granuloma annulare, morphea and lichen sclerosus: A pcr-based study and review of the literature".ti. and "2010".yr. (1)

678	"Enteroviral testing and length of hospital stay for children evaluated for lyme meningitis".ti. and "2013".yr. (1)

679	"Organisational intentions and responses: Presenting the risk of lyme disease to countryside users".ti. and "2013".yr. (0)

680	"Epidemics of vector-borne diseases observed in infectious disease surveillance in japan, 2000-2005".ti. and "2007".yr. (1)

681	"Level of awareness regarding some zoonotic diseases, among dog owners of ithaca, new york".ti. and "2014".yr. (0)

682	"Diagnostic novelties of chronic lyme/ neuroborreliosis".ti. and "2012".yr. and Romanian Society of Gastroenterology.af. (1)

683	"Behavioral and attitudes survey about lyme disease among a brazilian population in the endemic area of martha's vineyard, massachusetts".ti. and "2010".yr. (1)

684	"Diagnostic performance of cerebrospinal fluid chemokine cxcl13 and antibodies to the c6-peptide in lyme neuroborreliosis".ti. and "2011".yr. (1)

685	"Susceptibility, likelihood to be diagnosed, worry and fear for contracting lyme disease".ti. and "2017".yr. (1)

686	"Safety, immunogenicity, and efficacy of borrelia burgdorferi outer surface protein a (ospa) vaccine: A meta-analysis".ti. and "2017".yr. (1)

687	"Effectiveness of residential acaricides to prevent lyme and other tick-borne diseases in humans".ti. and "2016".yr. (1)

688	"Borrelia-specific interferon-gamma and interleukin-4 secretion in cerebrospinal fluid and blood during lyme borreliosis in humans: Association with clinical outcome".ti. and "2004".yr. (1)

689	"Primary cutaneous marginal zone lymphoma: Studies on borrelia infection".ti. and "2010".yr. (1)

690	"The epidemiology of lyme disease in the state of massachusetts, 2001-2011".ti. and "2014".yr. (1)

691	"Possible involvement of borrelia burgdorferi and hcv in the aethiology of mycosis fungoides".ti. and "2006".yr. (0)

692	"Epidemiology of tick bites and borreliosis in children attending kindergarten or so-called forest kindergarten in southwest germany".ti. and "2006".yr. (1)

693	"Isolation and detection of borrelia burgdorferi from human vaginal and seminal secretions".ti. and "2014".yr. (1)

694	"Morgellons disease: Analysis of a population with clinically confirmed microscopic subcutaneous fibers of unknown etiology".ti. and "2010".yr. (1)

695	"Safety of intravenous antibiotic therapy in chronic lyme disease".ti. and "2008".yr. (0)

696	"Clinical and seroepidemiologic characteristics of babesia wa-1 coinfection in patients with lyme disease in california".ti. and "2003".yr. (0)

697	"Clinical and immunologic evidence for transmission of lyme disease through intimate human contact".ti. and "2004".yr. (0)

698	"Lyme neuroborreliosis in hiv-1 positive men successfully treated with oral doxycycline: A case series and literature review".ti. and "2011".yr. (1)

699	"Spatiotemporal bayesian analysis of lyme disease in new york state, 1990-2000".ti. and "2006".yr. (1)

700	"Epidemic and spatial dynamics of lyme disease in new york state, 1990-2000".ti. and "2005".yr. (1)

701	"A prospective cross-over field trial shows protection of lemon eucalyptus extract against tick bites".ti. and "2004".yr. (1)

702	"Effects of reduced deer density on the abundance of ixodes scapularis (acari: Ixodidae) and lyme disease incidence in a northern new jersey endemic area".ti. and "2007".yr. (1)

703	"The relationship between deer density, tick abundance, and human cases of lyme disease in a residential community".ti. and "2014".yr. (1)

704	"Human behaviors elevating exposure to ixodes pacificus (acari: Ixodidae) nymphs and their associated bacterial zoonotic agents in a hardwood forest".ti. and "2004".yr. (1)

705	"The effects of bird feeders on lyme disease prevalence and density of ixodes scapularis (acari: Ixodidae) in a residential area of dutchess county, new york".ti. and "2003".yr. (1)

706	"Geographical and genospecies distribution of borrelia burgdorferi sensu lato dna detected in humans in the usa".ti. and "2014".yr. (1)

707	"Diagnostic accuracy and comparison of two assays for borrelia-specific igg and igm antibodies: Proposals for statistical evaluation methods, cut-off values and standardization".ti. and "2013".yr. (1)

708	"Audit of the laboratory diagnosis of lyme disease in scotland".ti. and "2005".yr. (1)

709	"Comparative evaluation of three different elisa methods for the diagnosis of early culture-confirmed lyme disease in italy".ti. and "2005".yr. (1)

710	"The use of local isolates in western blots improves serological diagnosis of lyme disease in scotland".ti. and "2007".yr. (1)

711	"Performance of a multiplexed serological microarray for the detection of antibodies against central nervous system pathogens".ti. and "2014".yr. (1)

712	"Community partnership designed to promote lyme disease prevention and engagement in citizen science".ti. and "2016".yr. (0)

713	"Cellular immune reactivity to recombinant borrellia-specific antigens for improved laboratory diagnosis of lyme borreliosis".ti. and "2004".yr. (0)

714	"Differentiating lyme arthritis from septic arthritis: Clues uncovered".ti. and "2011".yr. (0)

715	"Reactive lyme serology in optic neuritis".ti. and "2005".yr. (1)

716	"Cxcl13 and neopterin concentrations in cerebrospinal fluid of patients with lyme neuroborreliosis and other diseases that cause neuroinflammation".ti. and "2014".yr. (1)

717	"Neuroborreliosis-associated cerebral vasculitis: Long-term outcome and health-related quality of life".ti. and "2013".yr. (1)

718	"Quality of life, fatigue, depression and cognitive impairment in lyme neuroborreliosis".ti. and "2015".yr. (1)

719	"Prevalence and spectrum of residual symptoms in lyme neuroborreliosis after pharmacological treatment: A systematic review".ti. and "2016".yr. (1)

720	"Cerebrospinal fluid findings in adults with acute neuroborreliosis".ti. and "2011".yr. (1)

721	"Prevalence of taste disorders in idiopathic and b. Burgdorferi-associated facial palsy".ti. and "2009".yr. (1)

722	"lymphocyte chemoattractant (cxcl13) in the early diagnosis of acute lyme neuroborreliosis".ti. and "2008".yr. and erratum.pt. (1)

723	"Acute peripheral facial palsy in adults".ti. and "2005".yr. (1)

724	"Double infection with encephalitis and lyme borreliosis transmitted by tick bite".ti. and "2005".yr. (0)

725	"Diagnosis of lyme neuroborreliosis with antibodies to recombinant proteins dbpa, bbk32, and ospc, and vlse ir6 peptide".ti. and "2003".yr. (0)

726	"Lyme disease-related intracranial hypertension in children: Clinical and imaging findings".ti. and "2016".yr. (1)

727	"The chemokine cxcl13 as a diagnostic and therapy response marker in acute neuroborreliosis".ti. and "2010".yr. (1)

728	"Neuroborreliosis in the south west of england".ti. and "2006".yr. (0)

729	"Clinical features of double infection with tick-borne encephalitis and lyme borreliosis transmitted by tick bite".ti. and "2006".yr. (1)

730	"Neuroborelliosis in south west england".ti. and "2014".yr. (1)

731	"Pediatric lyme arthritis of the hip: The great imitator?".ti. and "2015".yr. (0)

732	"Primary lyme arthritis of the pediatric hip".ti. and "2011".yr. (1)

733	"Evaluating the child with acute hip pain (irritable hip ) in a lyme endemic region".ti. and "2015".yr. (1)

734	"Reportable bacterial infections among new york city-born infants, 2001-2009".ti. and "2016".yr. (1)

735	"Chronic lyme disease: A survey of connecticut primary care physicians".ti. and "2010".yr. (1)

736	"False positive lyme disease igm immunoblots in children".ti. and "2016".yr. (1)

737	"Transient prolonged corrected qt interval in lyme disease".ti. and "2006".yr. (1)

738	"New insights into stages of lyme disease symptoms from a novel hospital-based registry".ti. and "2014".yr. (1)

739	"An estimate of lyme borreliosis incidence in western europe+".ti. and "2016".yr. (0)

740	"Preventive behaviors and knowledge of tick-borne illnesses: Results of a survey from an endemic area".ti. and "2015".yr. (1)

741	"Outcomes of children treated for lyme arthritis: Results of a large pediatric cohort".ti. and "2010".yr. (1)

742	"Borrelia burgdorferi persists in the gastrointestinal tract of children and adolescents with lyme disease".ti. and "2002".yr. (1)

743	"Lyme vaccination safety".ti. and "2002".yr. (1)

744	"Cerebrovascular events in lyme neuroborreliosis".ti. and "2015".yr. (1)

745	"Cutaneous borreliosis associated with t cell-predominant infiltrates: A diagnostic challenge".ti. and "2015".yr. (1)

746	"Factors influencing the utilization of lyme disease-prevention behaviors in a high-risk population".ti. and "2004".yr. (1)

747	"Using administrative medical claims data to supplement state disease registry systems for reporting zoonotic infections".ti. and "2013".yr. (1)

748	"Pharmacist initiation of postexposure doxycycline for lyme disease prophylaxis".ti. and "2014".yr. (1)

749	"Wais-iii and wms-iii performance in chronic lyme disease".ti. and "2006".yr. (1)

750	"Acute neuroborreliosis with involvement of the central nervous system".ti. and "2010".yr. (1)

751	"Long-term cognitive deficits after treated lyme neuroborreliosis-neuropsychological profile and demographical characteristics".ti. and "2013".yr. (1)

752	"Facial palsy as an initial symptom of lyme neuroborreliosis in an austrian endemic area".ti. and "2013".yr. (1)

753	"Clinical and epidemiological peculiarities of mixt infection (mi) of lyme disease (ld) and tick-borne encephalitis (tbe)".ti. and "2005".yr. (0)

754	"Laboratory diagnosis of lyme neuroborreliosis is influenced by the test used: Comparison of two elisas, immunoblot and cxcl13 testing".ti. and "2014".yr. (1)

755	"Electrocardiograph abnormalities in children with lyme meningitis".ti. and "2012".yr. (1)

756	"Neuropathy and cognitive impairment following vaccination with the ospa protein of borrelia burgdorferi".ti. and "2004".yr. (1)

757	"Diagnostic value of sural nerve biopsy in patients with suspected borrelia neuropathy".ti. and "2008".yr. (1)

758	"Distribution and presentation of lyme borreliosis in scotland - analysis of data from a national testing laboratory".ti. and "2015".yr. (1)

759	"Lyme borreliosis in scotland during two peak periods".ti. and "2009".yr. (1)

760	"Is tayside becoming a scottish hotspot for lyme borreliosis?".ti. and "2011".yr. (1)

761	"Application of nanotrap technology for high sensitivity measurement of urinary outer surface protein a carboxyl-terminus domain in early stage lyme borreliosis".ti. and "2015".yr. (1)

762	"Lyme risk for immigrants to the united states: The role of an educational tool".ti. and "2005".yr. (1)

763	"Imported disease in emergency departments: An undiscovered country".ti. and "2006".yr. (1)

764	"Seroprevalence of west nile virus, crimean-congo hemorrhagic fever virus, francisella tularensis and borrelia burgdorferi in rural population of manisa, western turkey".ti. and "2016".yr. (1)

765	"Implications of gender in chronic lyme disease".ti. and "2009".yr. (1)

766	"The clinical and laboratory characteristics of early lyme disease differ between men and women".ti. and "2010".yr. (0)

767	"Lyme neuroborreliosis: Aetiology and diagnosis of facial palsy in children from tyrol".ti. and "2010".yr. (1)

768	"Identification of a novel pathogenic borrelia species causing lyme borreliosis with unusually high spirochaetaemia: A descriptive study".ti. and "2016".yr. (1)

769	"Safety and immunogenicity of a novel multivalent ospa vaccine against lyme borreliosis in healthy adults: A double-blind, randomised, dose-escalation phase 1/2 trial".ti. and "2013".yr. (1)

770	"Oral doxycycline versus intravenous ceftriaxone for european lyme neuroborreliosis: A multicentre, non-inferiority, double-blind, randomised trial".ti. and "2008".yr. (1)

771	"Etiology, diagnosis, and management of facial palsy: 2000 patients at a facial nerve center".ti. and "2014".yr. (1)

772	"Sudden deafness and lyme disease".ti. and "2003".yr. (1)

773	"Clinicopathological features of primary cutaneous b-cell lymphomas from an academic regional hospital in central italy: No evidence of borrelia burgdorferi association".ti. and "2007".yr. (1)

774	"Demonstration of b. Burgdorferi-dna in two cases of nodal lymphoma".ti. and "2004".yr. (1)

775	"Macrolide therapy of chronic lyme disease".ti. and "2003".yr. (1)

776	"Comparative study of early lyme disease: Erythema migrans in new york state and northeastern poland".ti. and "2002".yr. (1)

777	"Efficacy of a long-term antibiotic treatment in patients with a chronic tick associated poly-organic syndrome (tapos)".ti. and "2009".yr. (1)

778	"Feedback on difficulties raised by the interpretation of serological tests for the diagnosis of lyme disease".ti. and "2014".yr. (1)

779	"Impact of serodiagnosis on the management of lyme borreliosis at angers university hospital".ti. and "2014".yr. (1)

780	"The potential use of cholestyramine to reduce the risk of developing clostridium difficile-associated diarrhoea in patients receiving long-term intravenous ceftriaxone".ti. and "2015".yr. (1)

781	"Seroprevalence of borrelia burgdorferi in north eastern india".ti. and "2008".yr. (1)

782	"Presence of borrelia burgdorferi in endomyocardial biopsies in patients with new-onset unexplained dilated cardiomyopathy".ti. and "2010".yr. (1)

783	"Intrathecally produced igg and igm antibodies to recombinant vlse, vlse peptide, recombinant ospc and whole cell extracts in the diagnosis of lyme neuroborreliosis".ti. and "2014".yr. (1)

784	"Utility of serodiagnostics designed for use in the united states for detection of lyme borreliosis acquired in europe and vice versa".ti. and "2014".yr. (1)

785	"An ospa-polymerase chain reaction/restriction fragment length polymorphism-based method for sensitive detection and reliable differentiation of all european borrelia burgdorferi sensu lato species and ospa types".ti. and "2004".yr. (1)

786	"Diagnosis of lyme borreliosis using enzyme immunoanalysis".ti. and "2005".yr. (1)

787	"Bacterial tick-borne diseases caused by bartonella spp., borrelia burgdorferi sensu lato, coxiella burnetii, and rickettsia spp. Among patients with cataract surgery".ti. and "2014".yr. (1)

788	"The reportable events monthly report (remr)".ti. and "2013".yr. (2)

789	"The disease among u.S. Military members, active and reserve component, 2001-2008".ti. and "2009".yr. (0)

790	"[incidence of lyme disease in spain]".ti. and "2016".yr. (1)

791	"Estimating the incidence of new onset lyme disease in rhode island".ti. and "2008".yr. (1)

792	"Promoting specialty care for lyme disease: Lessons learned".ti. and "2006".yr. (1)

793	"Supervised resistance exercise for patients with persistent symptoms of lyme disease".ti. and "2015".yr. (1)

794	"Occupational diseases among farmers in poland".ti. and "2016".yr. (1)

795	"Outer surface protein e antibody response and its effect on complement factor h binding to ospe in lyme borreliosis".ti. and "2008".yr. (1)

796	"Epidemiologic and ecologic characteristics of some diseases transmitted by arthropods on the littoral of the republic of croatia".ti. and "2002".yr. (1)

797	"Lyme disease reporting for navy and marine corps (1997-2000)".ti. and "2003".yr. (1)

798	"Epidemiological characteristics and clinical manifestations of lyme borreliosis in croatia".ti. and "2006".yr. (1)

799	"Severity of lyme disease with persistent symptoms. Insights from a double-blind placebo-controlled clinical trial".ti. and "2008".yr. (1)

800	"Safety of intravenous antibiotic therapy in patients referred for treatment of neurologic lyme disease".ti. and "2010".yr. (1)

801	"Lyme disease--united states, 2001-2002".ti. and "2004".yr. (1)

802	"Effect of electronic laboratory reporting on the burden of lyme disease surveillance--new jersey, 2001-2006".ti. and "2008".yr. (1)

803	"Notes from the field: Update on lyme carditis, groups at high risk, and frequency of associated sudden cardiac death--united states".ti. and "2014".yr. (1)

804	"Lyme disease - united states, 2000".ti. and "2002".yr. and Marshall.au. (0)

805	"Morphoea and borrelia burgdorferi: Results from the scottish highlands in the context of the world literature".ti. and "2002".yr. (1)

806	"Improvement in the laboratory recognition of lyme borreliosis with the combination of culture and pcr methods".ti. and "2003".yr. (1)

807	"Surveillance for lyme disease--united states, 1992-2006".ti. and "2008".yr. (1)

808	"Lyme disease - united states, 2003-2005".ti. and "2007".yr. (1)

809	"Lyme disease - united states, 2001-2002".ti. and "2004".yr. (1)

810	("Lyme disease - united states, 2000".ti. and "2002".yr.) not MMWR - Morbidity & Mortality Weekly Report.jn. (4)

811	"Reported vectorborne and zoonotic diseases, u.S. Air force, 2000-2011".ti. and "2012".yr. (1)

812	"Surveillance snapshot: Lyme disease among beneficiaries of the military health system, 2001-2012".ti. and "2013".yr. (1)

813	"The geographic distribution of incident lyme disease among active component service members stationed in the continental united states, 2004-2013".ti. and "2014".yr. (1)

814	"Characterizing the relationship between tick bites and lyme disease in active component u.S. Armed forces in the eastern united states".ti. and "2015".yr. (1)

815	"Lyme disease trends--dutchess county, new york, 1992-2000".ti. and "2003".yr. (0)

816	"Relapsing inflammatory optic neuropathy: Is it a new autoimmune disease?".ti. and "2011".yr. (1)

817	"Pilot study into borrelia seroprevelence in new brunswick multiple sclerosis patients".ti. and "2016".yr. (1)

818	"Differential aspects of primary progressive multiple sclerosis and neuroborreliosis".ti. and "2010".yr. (1)

819	"Differential diagnosis of multiple sclerosis and chronic progressive borrelial encephalomyelitis".ti. and "2011".yr. (1)

820	"Borrelia burgdorferi - infection agent - risk assessment in ms".ti. and "2014".yr. (1)

821	"Lyme disease serology in amyotrophic lateral sclerosis".ti. and "2009".yr. (1)

822	"Facial nerve palsy associated with lyme disease".ti. and "2003".yr. (1)

823	"Serological testing for borrelia: It's a mess".ti. and "2011".yr. (0)

824	"General practitioner reported incidence of lyme carditis in the netherlands".ti. and "2015".yr. (1)

825	"Pediatric lyme neuroborreliosis: Different clinical presentations of the same agent; single center experience".ti. and "2016".yr. (1)

826	"Clinical evaluation and oct optic disc and macular analysis in 6 cases of neuroretinitis".ti. and "2011".yr. (1)

827	"Incidence of neuroborreliosis-associated cerebral vasculitis in germany".ti. and "2013".yr. (1)

828	"Relevance of the antibody index to diagnose lyme neuroborreliosis among seropositive patients".ti. and "2007".yr. (1)

829	"Lyme neuroborreliosis and dementia".ti. and "2013".yr. (1)

830	"European neuroborreliosis: Neuropsychological findings 30 months post treatment".ti. and "2011".yr. (0)

831	"A randomized, placebo-controlled trial of repeated iv antibiotic therapy for lyme encephalopathy".ti. and "2008".yr. (1)

832	"Practice parameter: Treatment of nervous system lyme disease".ti. and "2007".yr. (1)

833	("Lyme disease and optic neuritis: Long-term follow-up of seropositive patients".ti. and "2003".yr.) not comment.pt. not letter.pt. (1)

834	"Cognitive function in post-treatment lyme disease: Do additional antibiotics help?".ti. and "2003".yr. (1)

835	"Lyme disease and dementia".ti. and "2007".yr. (0)

836	("Study and treatment of post lyme disease" and "A randomized double masked clinical trial").ti. and "2003".yr. (1)

837	"Diagnostic accuracy of csfb-lymphocyte chemoattractant in lyme neuroborreliosis".ti. and "2007".yr. (0)

838	"A prospective study on the role of cxcl13 in lyme neuroborreliosis".ti. and "2011".yr. (1)

839	("Lyme disease and optic neuritis: Long-term follow-up of seropositive patients".ti. and "2003".yr.) not comment.pt. (2)

840	"Intrusive symptoms and infectious encephalopathies".ti. and "2016".yr. (1)

841	"Tick-borne infections and pediatric bipolar disorder".ti. and "2016".yr. (1)

842	"Diagnosis of neuroborreliosis - improved knowledge base for qualified antibody analysis and cerebrospinal fluid data pattern related interpretations".ti. and "2013".yr. (1)

843	"Treatment of north american lyme neuroborrelliosis with oral doxycycline and intravenous ceftriaxone: A comparative case series".ti. and "2015".yr. (1)

844	"Amyotrophic lateral sclerosis (als) with laboratory abnormalities of unknown significance (laus)-where does it begin and where does it end?".ti. and "2015".yr. (1)

845	"Amyotrophic lateral sclerosis and lyme borreliosis".ti. and "2006".yr. (0)

846	"Neuropsychological profile of children after an episode of neuroborreliosis".ti. and "2013".yr. (1)

847	"Lyme neuroborreliosis: Aetiology and diagnosis of facial palsy in children from tyrol".ti. and "2013".yr. (1)

848	"Mycoplasma pneumoniae and borrelia burgdorferi as cause for facial paralysis in children: Single-center experience of 10 years".ti. and "2013".yr. (1)

849	"Observation of children of mothers who underwent borreliosis in the course of pregnancy".ti. and "2009".yr. (1)

850	"Randomized trial of longer-term therapy for symptoms attributed to lyme disease".ti. and "2016".yr. (1)

851	"Antinuclear antibodies in patients with lyme disease".ti. and "2010".yr. (0)

852	"Presence and indigenous nature of lyme disease in southern italy".ti. and "2003".yr. (1)

853	"Molecular and microscopical evidence of ehrlichia spp. And borrelia burgdorferi sensu lato in patients, animals and ticks in the czech republic".ti. and "2002".yr. (1)

854	"Evaluation of the c6 enzyme-linked immunoadsorbent assay for the serodiagnosis of lyme borreliosis in north-eastern italy".ti. and "2006".yr. (1)

855	"Seroprevalence of borrelia burgdorferi infection among forestry workers and farmers in duzce, north-western turkey".ti. and "2008".yr. (1)

856	"Comparative evaluation of two enzyme linked immunosorbent assay methods and three western blot methods for the diagnosis of culture-confirmed early lyme borreliosis in italy".ti. and "2005".yr. (1)

857	"National surveillance 12-monthly data notification rate changes".ti. and "2009".yr. (1)

858	"No association of seropositivity for anti-borrelia igg antibody with mental and physical complaints".ti. and "2008".yr. (1)

859	"Cerebral metabolic changes associated with lyme disease".ti. and "2002".yr. (1)

860	"Difficulties of interpreting borrelia serology in patients with uveitis".ti. and "2011".yr. (1)

861	"Prevalence of borrelia burgdorferi infection in a series of 98 primary cutaneous lymphomas".ti. and "2011".yr. (1)

862	"Lyme disease in maine: A comparison of nedss surveillance data and maine health data organization hospital discharge data".ti. and "2014".yr. (0)

863	"Minocycline as a substitute for doxycycline in targeted scenarios: A systematic review".ti. and "2015".yr. (1)

864	"Geographic expansion of lyme disease in the southeastern united states, 2000-2014".ti. and "2015".yr. (1)

865	"The lymphocyte transformation test for borrelia detects active lyme borreliosis and verifies effective antibiotic treatment".ti. and "2012".yr. (1)

866	"Analysis of the influence of ascaris, borrelia, toxocara, toxoplasma and mycoplasma infections on the incidence and course of perthes disease - pilot study".ti. and "2013".yr. (1)

867	"Serological detection of lyme borreliosis agents in patients from korea, 2005-2009".ti. and "2011".yr. (1)

868	"Spirochetes in otology: Are we testing for the right pathogens?".ti. and "2008".yr. (1)

869	"No evidence for the diagnostic value of borrelia serology in patients with sudden hearing loss".ti. and "2012".yr. (1)

870	"Long-term outcome of facial palsy in neuroborreliosis".ti. and "2005".yr. (1)

871	"Microbiologic findings in acute facial palsy in children".ti. and "2013".yr. (2)

872	("Lyme disease associated with sudden sensorineural hearing loss: Case report and literature review".ti. and "2013".yr.) not letter.pt. (2)

873	"The vlse (ir6) peptide elisa in the serodiagnosis of lyme facial paralysis".ti. and "2004".yr. (1)

874	"Clinical characteristics, treatment and outcome of children with lyme arthritis in nova scotia".ti. and "2015".yr. (1)

875	"Surveillance for childhood lyme disease by the canadian pediatric surveillance program".ti. and "2015".yr. (1)

876	"Treatment of intractable headaches associated with lyme disease using neuromodulation techniques".ti. and "2010".yr. (1)

877	("Physician reported incidence of early and late lyme borreliosis".ti. and "2015".yr.) not erratum.pt. not article in press.pt. (3)

878	"Sensitivity of a point of care tick-test for the development of lyme borreliosis".ti. and "2013".yr. (1)

879	"Diagnosis and management of human borreliosis in romania".ti. and "2014".yr. (1)

880	"Pilot study assessing the effectiveness of factory-treated, long-lasting permethrin-impregnated clothing for the prevention of tick bites during occupational tick exposure in highly infested military training areas, germany".ti. and "2015".yr. (1)

881	"Epidemiological situation of lyme borreliosis in germany: Surveillance data from six eastern german states, 2002 to 2006".ti. and "2008".yr. (1)

882	"The association between infectious burden and parkinson's disease: A case-control study".ti. and "2015".yr. (0)

883	"Concordance of four commercial enzyme immunoassay and three immunoblot formats for the detection of lyme borreliosis antibodies in human serum: The two-tier approach remains".ti. and "2016".yr. (1)

884	"Overdiagnosis and overtreatment of lyme disease in children".ti. and "2002".yr. (1)

885	"Synovial fluid findings in children with knee monoarthritis in lyme disease endemic areas".ti. and "2014".yr. (1)

886	"Causes of febrile illnesses after a tick bite in slovenian children".ti. and "2003".yr. (1)

887	"Lyme borreliosis and acute peripheral facial palsy in slovenian children".ti. and "2010".yr. (1)

888	"Borrelia burgdorferi sensu lato bacteremia in slovenian children with solitary and multiple erythema migrans".ti. and "2011".yr. (1)

889	"Borrelial lymphocytoma in children".ti. and "2015".yr. (1)

890	"Azithromycin is equally effective as amoxicillin in children with solitary erythema migrans".ti. and "2015".yr. (1)

891	"Diagnostic utility of borrelia burgdorferi cerebrospinal fluid polymerase chain reaction in children with lyme meningitis".ti. and "2005".yr. (1)

892	"One-year follow-up of tick-borne central nervous system infections in childhood".ti. and "2012".yr. (1)

893	"Prevalence of lyme meningitis in children with aseptic meningitis in a lyme disease-endemic region".ti. and "2011".yr. (1)

894	"New antigens for serologic diagnosis of neuroborreliosis in children".ti. and "2005".yr. (1)

895	"The positive predictive value of lyme elisa for the diagnosis of lyme disease in children".ti. and "2015".yr. (1)

896	"Early differentiation of lyme from enteroviral meningitis".ti. and "2005".yr. (1)

897	"Improved laboratory diagnostics of lyme neuroborreliosis in children by detection of antibodies to new antigens in cerebrospinal fluid".ti. and "2008".yr. (1)

898	"Lyme neuroborreliosis in children: A prospective study of clinical features, prognosis, and outcome".ti. and "2008".yr. (1)

899	"Treatment complications in children with lyme meningitis".ti. and "2012".yr. (1)

900	"Successful treatment of neuroborreliosis with ten day regimens".ti. and "2002".yr. (1)

901	"Prediction of lyme meningitis based on a logistic regression model using clinical and cerebrospinal fluid analysis: A european study".ti. and "2009".yr. (1)

902	"Etiology of aseptic meningitis, peripheral facial nerve palsy, and a combination of both in children".ti. and "2010".yr. (1)

903	"Accuracy of reporting of lyme disease in connecticut".ti. and "2003".yr. (0)

904	"Effectiveness of personal preventive measures for lyme disease".ti. and "2003".yr. (0)

905	"Lyme arthritis outcomes in children: A single center cohort study".ti. and "2012".yr. (1)

906	"Infectious agents in juvenile scleroderma".ti. and "2014".yr. (1)

907	"Prediction of lyme meningitis in children from a lyme disease-endemic region: A logistic-regression model using history, physical, and laboratory findings".ti. and "2006".yr. (1)

908	"Validation of a clinical prediction rule to distinguish lyme meningitis from aseptic meningitis".ti. and "2012".yr. (1)

909	"Lyme carditis in children: Presentation, predictive factors, and clinical course".ti. and "2009".yr. (1)

910	"Distinguishing lyme from septic knee monoarthritis in lyme disease-endemic areas".ti. and "2013".yr. (1)

911	"Comparative study of cefuroxime axetil versus amoxicillin in children with early lyme disease".ti. and "2002".yr. (1)

912	"Prospective validation of a clinical prediction model for lyme meningitis in children".ti. and "2009".yr. (1)

913	"Clinical predictors of lyme disease among children with a peripheral facial palsy at an emergency department in a lyme disease-endemic area".ti. and "2008".yr. (1)

914	"Long-term clinical outcome after lyme neuroborreliosis in childhood".ti. and "2012".yr. (1)

915	"Acute pediatric monoarticular arthritis: Distinguishing lyme arthritis from other etiologies".ti. and "2009".yr. (1)

916	"Long-term neuropsychologic and health outcomes of children with facial nerve palsy attributable to lyme disease".ti. and "2003".yr. (1)

917	"Severity of chronic lyme disease compared to other chronic conditions: A quality of life survey".ti. and "2014".yr. (1)

918	"Development of a public health assessment tool to prevent lyme disease: Tool construction and validation".ti. and "2005".yr. (0)

919	"Bilateral facial paralysis: A 13 year experience".ti. and "2016".yr. (1)

920	"Health care costs, utilization and patterns of care following lyme disease".ti. and "2015".yr. (1)

921	"Adaptation and evaluation of a multi-criteria decision analysis model for lyme disease prevention".ti. and "2015".yr. (1)

922	"Seropositivity of lyme borreliosis and associated risk factors: A population-based study in children and adolescents in germany (kiggs)".ti. and "2012".yr. (1)

923	"Direct molecular detection and genotyping of borrelia burgdorferi from whole blood of patients with early lyme disease".ti. and "2012".yr. (1)

924	"Integrated assessment of behavioral and environmental risk factors for lyme disease infection on block island, rhode island".ti. and "2014".yr. (1)

925	"A prospective study among patients presenting at the general practitioner with a tick bite or erythema migrans in the netherlands".ti. and "2013".yr. (1)

926	"Quantitative detection of borrelia burgdorferi sensu lato in erythema migrans skin lesions using internally controlled duplex real time pcr".ti. and "2013".yr. (1)

927	"C6 peptide-based multiplex phosphorescence analysis (phosphan) for serologic confirmation of lyme borreliosis".ti. and "2015".yr. (1)

928	"Are patients with erythema migrans who have leukopenia and/or thrombocytopenia coinfected with anaplasma phagocytophilum or tick-borne encephalitis virus?".ti. and "2014".yr. (1)

929	"Clinical characteristics associated with borrelia burgdorferi sensu lato skin culture results in patients with erythema migrans".ti. and "2013".yr. (1)

930	"Gender disparity between cutaneous and non-cutaneous manifestations of lyme borreliosis".ti. and "2013".yr. (1)

931	"Correlation of culture positivity, pcr positivity, and burden of borrelia burgdorferi sensu lato in skin samples of erythema migrans patients with clinical findings".ti. and "2015".yr. (1)

932	"The degree of damage in the peripheral facial nerve palsy in children depending on the cause: The role of lyme neuroborreliosis".ti. and "2014".yr. (1)

933	"Workplace health threats and their effects in podlasie region".ti. and "2008".yr. (0)

934	("Tick-borne infections as a cause of heart transplantation".ti. and "2011".yr.) not article.pt. (1)

935	"Comparison of detection of borrelia burgdorferi dna and anti-borrelia burgdorferi antibodies in patients with erythema migrans in north-eastern poland".ti. and "2015".yr. (1)

936	"Simultaneous gastrointestinal infections in children and adolescents".ti. and "2004".yr. (1)

937	"Lyme borreliosis infection as a cause of dilated cardiomyopathy".ti. and "2006".yr. (1)

938	"Significance of borrelia infection in development of dilated cardiomypathy (a pilot study)".ti. and "2007".yr. (1)

939	"Lyme borreliosis--waiting for lyme carditis? A long-term prospective study".ti. and "2005".yr. (1)

940	"Prevention of lyme disease in dutch children: Analysis of determinants of tick inspection by parents".ti. and "2002".yr. (1)

941	"Pulse therapy with ceftriaxone on lyme neuroborreliosis".ti. and "2005".yr. (1)

942	"Lyme borreliosis in patients over 60-years-old".ti. and "2010".yr. (1)

943	"Human serum reactivity against borrelia ospc, ospa, flab and vlse protein antigens in early and disseminated lyme borreliosis".ti. and "2010".yr. (1)

944	"Tick-bite preventative behavior of people on recreational areas in warsaw [conference poster]".ti. and "2014".yr. (0)

945	"Deer, predators, and the emergence of lyme disease".ti. and "2012".yr. (1)

946	"Invasion of two tick-borne diseases across new england: Harnessing human surveillance data to capture underlying ecological invasion processes".ti. and "2016".yr. (1)

947	"Epidemiology of borreliosis in workers of the district forestry offices in lower silesia".ti. and "2007".yr. (1)

948	"Lyme disease with effusion either in hip or knee in children from podlaskie region treated in clinic in 2004-2010".ti. and "2014".yr. (1)

949	"Lyme disease in poland in 2013".ti. and "2015".yr. (1)

950	"Lyme disease in poland in 2011".ti. and "2013".yr. (1)

951	"Lyme disease in poland in 2012".ti. and "2014".yr. (1)

952	"Cns lyme disease manifestation in children".ti. and "2007".yr. (1)

953	"Correlates of perceived health-related quality of life in post-treatment lyme encephalopathy".ti. and "2013".yr. (1)

954	"Patients with organically unexplained symptoms presenting to a borreliosis clinic: Clinical and psychobehavioral characteristics and quality of life".ti. and "2013".yr. (1)

955	"Lyme disease and post-treatment lyme disease syndrome: The neglected disease in our own backyard".ti. and "2014".yr. (1)

956	"Lyme disease - what is the cost for scotland?".ti. and "2003".yr. (1)

957	"A qualitative approach to understanding patients' diagnosis of lyme disease".ti. and "2006".yr. (1)

958	"Evaluation of deer-targeted interventions on lyme disease incidence in connecticut".ti. and "2011".yr. (1)

959	"Lyme disease in a british referral clinic".ti. and "2012".yr. (1)

960	"Management of suspected lyme borreliosis: Experience from an outpatient parenteral antibiotic therapy service".ti. and "2013".yr. (1)

961	"Living in limbo: Contested narratives of patients with chronic symptoms following lyme disease".ti. and "2015".yr. (0)

962	"Post-treatment lyme disease syndrome symptomatology and the impact on life functioning: Is there something here?".ti. and "2013".yr. (2)

963	"Neuro-lyme disease: Mr imaging findings".ti. and "2009".yr. (1)

964	"Reduction in lyme disease risk as an economic benefit of habitat restoration".ti. and "2012".yr. (0)

965	"Prevalence of yersinia spp., chlamydia trachomatis, chlamydophila pneumoniae and borrelia burgdorferi antibodies in healthy blood donors' sera".ti. and "2013".yr. (1)

966	"Antibody profile to borrelia burgdorferi in veterinarians from nuevo leon, mexico, a non-endemic area of this zoonosis".ti. and "2016".yr. (1)

967	"Articular manifestations in patients with lyme disease".ti. and "2015".yr. (0)

968	"Do infections trigger juvenile idiopathic arthritis?".ti. and "2011".yr. (1)

969	"Evaluation of skin thickness lesions in patients with lyme disease measured by modified rodnan total skin score".ti. and "2012".yr. (1)

970	"Seroprevalence of borrelia burgdorferi in patients with behcet's disease".ti. and "2003".yr. (1)

971	"Rhode island tick-borne disease surveillance summary 2012-2013".ti. and "2014".yr. (1)

972	"Diagnostic impact of routine lyme serology in recent-onset arthritis: Results from the espoir cohort".ti. and "2016".yr. (0)

973	"Serological and clinical one year follow-up of patients with erythema migrans treated in a romanian infectious disease hospital".ti. and "2014".yr. (0)

974	"Serological, clinical and epidemiological aspects of lyme borreliosis in mures county, romania".ti. and "2014".yr. (0)

975	"Reinfection with lyme borreliosis: A retrospective follow-up study in southern sweden".ti. and "2002".yr. (1)

976	"Clinical outcome of erythema migrans after treatment with phenoxymethyl penicillin".ti. and "2003".yr. (1)

977	"5-y follow-up study of patients with neuroborreliosis".ti. and "2002".yr. (1)

978	"Human granulocytic ehrlichiosis as a common cause of tick-associated fever in southeast sweden: Report from a prospective clinical study".ti. and "2002".yr. (1)

979	"Characteristics of erythema migrans in borrelia afzelii and borrelia garinii infections".ti. and "2003".yr. (1)

980	"Three major lyme borrelia genospecies (borrelia burgdorferi sensu stricto, b. Afzelii and b. Garinii) identified by pcr in cerebrospinal fluid from patients with neuroborreliosis in sweden".ti. and "2002".yr. (1)

981	"Risk of acquiring tick bites in south-eastern sweden".ti. and "2002".yr. (1)

982	"Intravenous ceftriaxone compared with oral doxycycline for the treatment of lyme neuroborreliosis".ti. and "2005".yr. (1)

983	"Seroprevalence of rickettsia spp. Infection among tick-bitten patients and blood donors in sweden".ti. and "2008".yr. (1)

984	"Serological and molecular evidence of rickettsia helvetica in denmark".ti. and "2004".yr. (1)

985	"Clinical characteristics of childhood lyme neuroborreliosis in an endemic area of northern europe".ti. and "2009".yr. (1)

986	"Cerebrospinal fluid chemokine cxcl13 in the diagnosis of neuroborreliosis in children".ti. and "2013".yr. (1)

987	"Acute facial nerve palsy in children: How often is it lyme borreliosis?".ti. and "2007".yr. (1)

988	"Laboratory data in children with lyme neuroborreliosis, relation to clinical presentation and duration of symptoms".ti. and "2009".yr. (1)

989	"Lyme disease and schizophrenia: Case studies from an adjunctive minocycline study".ti. and "2013".yr. (1)

990	"Tick bites and erythema migrans in transylvania".ti. and "2013".yr. (0)

991	"Diagnosis and treatment in patients with chronic tick associated poly-organic syndrome (tapos) - a case series".ti. and "2010".yr. (0)

992	"Situating risk in the context of a woodland visit: A case study on lyme borreliosis".ti. and "2012".yr. (0)

993	"Urban and rural risks of lyme disease in the scottish highlands".ti. and "2009".yr. (1)

994	"Borreliosis - cause of obstructive sleep apnea syndrome. Comparison with normal osas".ti. and "2009".yr. (0)

995	"Socio-economic characteristics in notified erythema migrans patients".ti. and "2015".yr. (0)

996	"Borrelia burgdorferi (strain b. Afzelii) antibodies among malaysian blood donors and patients".ti. and "2002".yr. (1)

997	"Comparing spatio-temporal clusters of arthropod-borne infections using administrative medical claims and state reported surveillance data".ti. and "2012".yr. (1)

998	"Landscape risk factors for lyme disease in the eastern broadleaf forest province of the hudson river valley and the effect of explanatory data classification resolution".ti. and "2015".yr. (1)

999	"Tick related diseases in switzerland, 2008 to 2011".ti. and "2013".yr. (1)

1000	"Intentions to receive a potentially available lyme disease vaccine in an urban sample".ti. and "2016".yr. (1)

1001	"Clinical features of 705 borrelia burgdorferi seropositive patients in an endemic area of northern italy".ti. and "2014".yr. (1)

1002	"An mr brain images classifier system via particle swarm optimization and kernel support vector machine".ti. and "2013".yr. (1)

1003	"Putative association between anti-borrelia igg and autoimmune thyroid disease?".ti. and "2005".yr. (1)

1004	"Incidence and antibiotic treatment of erythema migrans in norway 2005-2009".ti. and "2016".yr. (0)

1005	"U.S. Healthcare providers' experience with lyme and other tick-borne diseases".ti. and "2014".yr. (1)

1006	"Tick-borne disease preventive practices and perceptions in an endemic area".ti. and "2016".yr. (1)

1007	"Human tick infestation pattern, tick-bite rate, and associated borrelia burgdorferi s.L. Infection risk during occupational tick exposure at the seedorf military training area, northwestern germany".ti. and "2014".yr. (1)

1008	"Epidemiology of lyme disease in low-incidence states".ti. and "2015".yr. (1)

1009	"Continuing increase of tick bites and lyme disease between 1994 and 2009".ti. and "2015".yr. (1)

1010	"U.S. Public's experience with ticks and tick-borne diseases: Results from national healthstyles surveys".ti. and "2015".yr. (1)

1011	"Prospective study on the incidence of infection by borrelia burgdorferi sensu lato after a tick bite in a highly endemic area of switzerland".ti. and "2011".yr. (1)

1012	"Functional outcomes in patients with borrelia burgdorferi reinfection".ti. and "2014".yr. (1)

1013	"Serological and molecular evidence for spotted fever group rickettsia and borrelia burgdorferi sensu lato co-infections in the netherlands".ti. and "2016".yr. (1)

1014	"Epidemiology and cost of hospital care for lyme borreliosis in germany: Lessons from a health care utilization database analysis".ti. and "2015".yr. (1)

1015	"Climate change influences on the annual onset of lyme disease in the united states".ti. and "2015".yr. (1)

1016	"Predictors of protective behaviour against ticks in the uk: A mixed methods study".ti. and "2014".yr. (1)

1017	"Human seroprevalence against borrelia burgdorferi sensu lato in two comparable regions of the eastern alps is not correlated to vector infection rates".ti. and "2015".yr. (1)

1018	"Geographical differences in seroprevalence of borrelia burgdorferi antibodies in norway, 2011-2013".ti. and "2016".yr. (1)

1019	"A prospective study on the incidence of borrelia burgdorferi sensu lato infection after a tick bite in sweden and on the aland islands, finland (2008-2009)".ti. and "2016".yr. (1)

1020	"Trends in surveillance data of human lyme borreliosis from six federal states in eastern germany, 2009-2012".ti. and "2014".yr. (1)

1021	"Spatial distribution of a population at risk: An important factor for understanding the recent rise in tick-borne diseases (lyme borreliosis and tick-borne encephalitis in the czech republic)".ti. and "2013".yr. (1)

1022	"Peri-urbanisation, counter-urbanisation, and an extension of residential exposure to ticks: A clue to the trends in lyme borreliosis incidence in the czech republic?".ti. and "2014".yr. (1)

1023	"Evidence of rapid changes in lyme disease awareness in canada".ti. and "2016".yr. (1)

1024	"Knowledge and prevention of tick-borne diseases vary across an urban-to-rural human land-use gradient".ti. and "2013".yr. (1)

1025	"Risk factors identified in blood donors screening positive or inconclusive for babesia microti".ti. and "2014".yr. (1)

1026	"Prevalence of lyme borreliosis in scottish blood donors".ti. and "2011".yr. (1)

1027	"Seroprevalence of antibodies to borrelia burgdorferi in a healthy swiss blood donor population".ti. and "2011".yr. (1)

1028	"Diagnosis of febrile illnesses in returned travelers using the pc software gideon".ti. and "2005".yr. (1)

1029	"An analysis of imported infections over a 5-year period at a teaching hospital in the united kingdom".ti. and "2003".yr. (1)

1030	"Incidence rates of selected infectious diseases in the most populated counties, united states, 2004".ti. and "2008".yr. (0)

1031	"Investigation of borrelia burgdorferi seroprevalence in van region of turkey".ti. and "2008".yr. (1)

1032	"Analysis of borrelia burgdorferi antibody seroprevalence in patients with behcet's disease using 3 different serological assays".ti. and "2004".yr. (1)

1033	"Evaluation of borrelia burgdorferi sensu lato seroprevalence in the province of bolu, turkey".ti. and "2016".yr. (1)

1034	"The seroprevalence of anaplasma phagocytophilum in humans from two different climatic regions of turkey and its co-seroprevalence rate with borrelia burgdorferi turkiyenin iki farkli".ti. and "2011".yr. (0)

1035	"Adverse event reports following vaccination for lyme disease: December 1998-july 2000".ti. and "2002".yr. (1)

1036	"Antibiotic treatment patterns associated with persons newly-diagnosed with lyme disease in the united states".ti. and "2014".yr. (1)

1037	"Assessing the incidence and treatment of post-treatment lyme disease syndrome in an administrative claims database".ti. and "2014".yr. (1)

1038	"The geographic correlation between lyme disease incidence and degenerative neurological disease mortality: An ecological study".ti. and "2013".yr. (1)

1039	"Assessing lyme disease prevalence in the united states medicaid population".ti. and "2014".yr. (1)

1040	"Effect of gender on clinical and epidemiologic features of lyme borreliosis".ti. and "2007".yr. (1)

1041	"Lyme borreliosis incidence in two french departments: Correlation with infection of ixodes ricinus ticks by borrelia burgdorferi sensu lato".ti. and "2007".yr. (1)

1042	"Knowledge, attitudes, and behaviors regarding lyme disease prevention among connecticut residents, 1999-2004".ti. and "2008".yr. (1)

1043	"How big is the lyme problem? Using novel methods to estimate the true number of lyme disease cases in british columbia residents from 1997 to 2008".ti. and "2011".yr. (1)

1044	"Passive tick surveillance, dog seropositivity, and incidence of human lyme disease".ti. and "2004".yr. (1)

1045	"Empiric antibiotic treatment of erythema migrans-like skin lesions as a function of geography: A clinical and cost effectiveness modeling study".ti. and "2013".yr. (1)

1046	"Evaluation of recombinant line immunoblot for detection of lyme disease in slovakia: Comparison with two other immunoassays".ti. and "2008".yr. (1)

1047	"Long-term lyme disease antibiotic therapy beliefs among new england residents".ti. and "2011".yr. (1)

1048	"A serological survey about zoonoses in the verkhoyansk area, northeastern siberia (sakha republic, russian federation)".ti. and "2016".yr. (1)

1049	"Seroepidemiology of nine zoonoses in viljujsk, republic of sakha (northeastern siberia, russian federation)".ti. and "2011".yr. (1)

1050	"Estimated incidence of erythema migrans in five regions of france and ecological correlations with environmental characteristics".ti. and "2013".yr. (1)

1051	"Precipitation and the occurrence of lyme disease in the northeastern united states".ti. and "2004".yr. (1)

1052	"The occurrence of spotted fever rickettsioses and other tick-borne infections in forest workers in poland".ti. and "2011".yr. (1)

1053	"A school-based intervention to increase lyme disease preventive measures among elementary school-aged children".ti. and "2016".yr. (1)

1054	"Comparison of erythema migrans caused by borrelia burgdorferi and borrelia garinii".ti. and "2011".yr. (1)

1055	"Comparison of post-lyme borreliosis symptoms in erythema migrans patients with positive and negative borrelia burgdorferi sensu lato skin culture".ti. and "2011".yr. (1)

1056	"Using nation-wide health insurance claims data to augment lyme disease surveillance".ti. and "2015".yr. (1)

1057	"Two introductions of lyme disease into connecticut: A geospatial analysis of human cases from 1984 to 2012".ti. and "2015".yr. (1)

1058	"Is localized scleroderma caused by borrelia burgdorferi?".ti. and "2016".yr. (1)

1059	"Tick-borne encephalitis and lyme disease in hungary: The epidemiological situation between 1998 and 2008".ti. and "2013".yr. (1)

1060	"Ticking all the boxes? A systematic review of education and communication interventions to prevent tick-borne disease".ti. and "2012".yr. (1)

1061	"Subset-directed antiviral treatment of 142 herpesvirus patients with chronic fatigue syndrome".ti. and "2010".yr. (1)

1062	"The pilot studies for lyme disease (lyme borreliosis) in blood donors in rckik in poznan".ti. and "2015".yr. (1)

1063	"Tick exposure and lyme disease at a summer camp in maryland".ti. and "2005".yr. (1)

1064	"Children with multiple erythema migrans: Are there any pre-treatment symptoms and/or signs suggestive for central nervous system involvement?".ti. and "2002".yr. (1)

1065	"Solitary erythema migrans in children: Comparison of treatment with azithromycin and phenoxymethylpenicillin".ti. and "2002".yr. (1)

1066	"Development and laboratory evaluation of a new recombinant elisa for the serodiagnosis of lyme disease".ti. and "2002".yr. (1)

1067	"Solitary borrelial lymphocytoma in adult patients".ti. and "2002".yr. (1)

1068	"Development of erythema migrans in spite of treatment with antibiotics after a tick bite".ti. and "2002".yr. (1)

1069	"Electrocardiographic findings in patients with erythema migrans".ti. and "2002".yr. (1)

1070	"Evaluation of immunofluorescence test (ift) and immuno (western) blot (wb) test in patients with erythema migrans".ti. and "2002".yr. (1)

1071	"Clinical and epidemiological findings for patients with erythema migrans. Comparison of cohorts from the years 1993 and 2000".ti. and "2002".yr. (1)

1072	"Limitation of serological testing for lyme borreliosis: Evaluation of elisa and western blot in comparison with pcr and culture methods".ti. and "2002".yr. (1)

1073	"Clinical appearance of erythema migrans caused by borrelia afzelii and borrelia garinii--effect of the patient's sex".ti. and "2006".yr. (1)

1074	"Comparison of immunofluorescence assay (ifa) and liaison in patients with different clinical manifestations of lyme borreliosis".ti. and "2006".yr. (1)

1075	"Paradigm burgenland: Risk of borrelia burgdorferi sensu lato infection indicated by variable seroprevalence rates in hunters".ti. and "2006".yr. (1)

1076	"Clinical and epidemiological features of lyme borreliosis in bulgaria".ti. and "2004".yr. (1)

1077	"A comparison of two treatment regimens of ceftriaxone in late lyme disease".ti. and "2005".yr. (1)

1078	"Peripheral facial palsy as an initial symptom of lyme neuroborreliosis in an austrian endemic area".ti. and "2015".yr. (0)

1079	"Contact with horses is a risk factor for tick-borne lymphadenopathy".ti. and "2012".yr. (1)

1080	"Electrocardiographic findings in children with erythema migrans".ti. and "2006".yr. (1)

1081	"Solitary erythema migrans in children: Comparison of treatment with clarithromycin and amoxicillin".ti. and "2012".yr. (1)

1082	"Doxycycline versus ceftriaxone for the treatment of patients with chronic lyme borreliosis".ti. and "2006".yr. (1)

1083	"Prevalence and incidence of lyme borreliosis among slovene forestry workers during the period of tick activity".ti. and "2005".yr. (1)

1084	"Appalachian trail hikers' ability to recognize lyme disease by visual stimulus photographs".ti. and "2014".yr. (1)

1085	"Deer hunting in a residential community: The community's perspective".ti. and "2003".yr. (0)

1086	"Lyme disease testing in children in an endemic area".ti. and "2011".yr. (1)

1087	"Our experience with enzyme-linked immunospot assay in the laboratory diagnostics of lyme borreliosis".ti. and "2015".yr. (1)

1088	"Seroprevalence of anti-toxoplasma gondii and anti-borrelia species antibodies in patients with schizophrenia: A case-control study from western turkey".ti. and "2015".yr. (1)

1089	"Respiratory modulation of cardiac vagal tone in lyme disease".ti. and "2014".yr. (1)

1090	"Lyme disease in west virginia: An assessment of distribution and clinicians' knowledge of disease and surveillance".ti. and "2016".yr. (1)

1091	"The importance of the size of erythema migrans (em) for diagnosis of lyme borreliosis in slovenian children o pomembnosti velikosti erythema migrans (em) za diagnozo lymske borelioze pri otrocih v sloveniji".ti. and "2012".yr. (0)

1092	"Serologic diagnosis of lyme disease with indirect immunofluorescence assay, enzyme-linked immunosorbent assay and western blot".ti. and "2011".yr. (0)

1093	"Preliminary survey of ticks and tick-borne pathogens in tianjin, china".ti. and "2013".yr. (0)

1094	"Confirmed exposure to tick-borne encephalitis virus and probable human cases of tick-borne encephalitis in central/northern anatolia, turkey".ti. and "2011".yr. (1)

1095	"Lyme disease surveillance using sampling estimation: Evaluation of an alternative methodology in new york state".ti. and "2016".yr. (0)

1096	"Tick bite prophylaxis: Results from a 2012 survey of healthcare providers".ti. and "2015".yr. (1)

1097	"Characterizing lyme disease surveillance in an endemic state".ti. and "2016".yr. (0)

1098	"Underreporting of lyme and other tick-borne diseases in residents of a high-incidence county, minnesota, 2009".ti. and "2016".yr. (0)

1099	or/1-1098 (920)

1100	1099 not (69 or 282 or 310 or 633 or 871 or 872 or 962) (907)

1101	69 or 282 or 310 or 633 or 871 or 872 or 962 (13)

1102	remove duplicates from 1101 (7)

1103	1100 or 1102 (914)

1104	1103 (914)

1105	limit 1104 to english language (913)

1106	1104 not 1105 (1)

1107	limit 1104 to (english language or no language specified) (913)

1108	1104 not 1107 (1)

1109	limit 1104 to (english language or no language specified or polyglot) (913)

1110	1104 not 1109 (1)

1111	((afrikaans or albanian or arabic or armenian or azerbaidzhani or azerbaijani or basque or belorussian or bengali or bulgarian or burmese or bosnian or catalan or catalan; valencian or chinese or croatian or czech or danish or dutch or dutch; flemish or esperanto or estonian or finnish or french or gallegan or georgian or german or greek or "greek, modern (1453-)" or hebrew or hindi or hungarian or icelandic or indonesian or italian or japanese or korean or latvian or lithuanian or macedonian or malay or maori or mongolian or norwegian or persian or polish or portuguese or pushto or romanian or russian or serbian or sinhalese or slovak or slovene or slovenian or spanish or spanish; castilian or swedish or tagalog or thai or turkish or ukrainian or urdu or uzbek or vietnamese) not english).lg. (4690568)

1112	1104 not 1111 (913)

1113	1104 and 1111 (1)

1114	((afrikaans or albanian or arabic or armenian or azerbaidzhani or azerbaijani or basque or belorussian or bengali or bulgarian or burmese or bosnian or catalan or catalan; valencian or chinese or croatian or czech or danish or dutch or dutch; flemish or esperanto or estonian or finnish or french or gallegan or georgian or german or greek or "greek, modern (1453-)" or hebrew or hindi or hungarian or icelandic or indonesian or italian or japanese or korean or latvian or lithuanian or macedonian or malay or maori or mongolian or norwegian or persian or polish or portuguese or pushto or romanian or russian or serbian or sinhalese or slovak or slovene or slovenian or spanish or spanish; castilian or swedish or tagalog or thai or turkish or ukrainian or urdu or uzbek or vietnamese) not (english or polyglot)).lg. (4690568)

1115	1104 not 1114 (913)

1116	1104 and 1114 (1)

**Excludes: MEDLINE® ALL** via Ovid http://ovidsp.ovid.com/

Date range searched: <1946 to January 29, 2024>

Date searched: 30 January 2024

1	"[duration of antibiotic therapy for lyme disease]".ti. and "Swedish Council".af. (0)

2	"[lyme-borreliosis: Diagnosis, symptoms and treatment]".ti. (1)

3	"Dossier: Lyme disease".ti. (0)

4	"16th consensus conference on anti-infectious therapy of the spilf. Lyme borreliosis: Diagnostic, therapeutic and preventive steps. Long text. Paris, france, 13 december 2006".ti. (0)

5	"[isolated intracranial hypertension due to lyme's disease]".ti. (1)

6	"[neuroborreliosis or borrelia hysteria. This case becomes a nightmare!]".ti. (1)

7	"[the patient is worried about tick bite. Is it usefull to test for borrelia?]".ti. (1)

8	"[trigeminal neuralgia, neuroborreliosis, and herpes: Finding the intruder]".ti. (1)

9	"A case of lyme disease with facial nerve palsy. [japanese]".ti. (0)

10	"[tick-borne diseases exposure of forestry workers and preventive methods usage]".ti. (1)

11	"[sequelae of tick-born encephalitis in cases treated in the infection observatory ward of regional hospital in gizycko]".ti. (1)

12	"[clinical thinking in neurological diagnosis (french experience)]".ti. (1)

13	"[evaluation of 10 cases of lyme disease presenting with erythema migrans in istanbul, turkey]".ti. (1)

14	"Lyme disease, first reported case in ecuador. [spanish] enfermedad de lyme. Primer caso reportado en ecuador".ti. (0)

15	"Guillain-barre syndrome after neuroborreliosis? Similar syndrome, different disease".ti. (0)

16	"Erratum: Diagnostic utility of serology to b. Burgdorferi in cartagena".ti. (0)

17	"Communicable diseases in croatia in 2007. [croatian] zarazne bolesti u hrvatskoj 2007".ti. (0)

18	"A fourteen-year-old boy with facial paresis caused by lyme disease. [dutch] een 14-jarige jongen met een facialisparese door de ziekte van lyme".ti. (0)

19	"An investigation of the prevalence of borrelia burgdorferi antibodies in north cyprus".ti. (0)

20	"[facial palsy: Diagnosis and management by primary care physicians]".ti. (1)

21	"[risk of human infection with ixodes tick-borne relapsing fever in different regional subzones of the perm region]".ti. (1)

22	"[use of indirect micromodification of enzyme immune (imei) reaction for mass screening serological studies for ixodes tick-borne borreliosis]".ti. (1)

23	"How to treat lyme borreliosis in a pregnant woman".ti. (0)

24	"[articular lesions in ixodes tick-borne borreliosis (lyme disease)]".ti. (1)

25	"[use of western-immunoblotting to determine serum igg-antibodies to antigens of different genotypes of borrelia--borrelia burgdorferi sensu stricto, borrelia afzelii, and borrelia garinii]".ti. (1)

26	"[detection of anti-borrelia antibodies by immunoblotting in lyme borreliosis]".ti. (1)

27	"[rapid development of dementia of an elderly person, diagnosis and successful treatment]".ti. (1)

28	"Creutzfeldt-jakob disease in clinical practice: A case report. [bosnian] creutzfeldt-jakobova bolest u klinickoj praksi: Prikaz bolesnice".ti. (0)

29	"Difficulty in analysis of serological tests results for spirochete infection in children".ti. (0)

30	"Skin manifestations of spirochete infection in chldren - own observations skorna manifestacja zakazenia kre{ogonek}tkiem borrelia burgdorferi u dzieci - doswiadczenia wlasne".ti. (0)

31	"[clinical course of borreliosis in children]".ti. (1)

32	"Analysis of the clinical course of infection caused by borrelia burgdorferi in children. [polish] analiza przebiegu klinicznego zakazenia kretkiem borrelia burgdorferi u dzieci".ti. (0)

33	"[specific features of borrelia burgdorferi infection in children]".ti. (1)

34	"Arthritis in the course of borrelia burgdorferi infection in children".ti. (0)

35	"[diagnostic problems with borrelia burgdorferi infection in children]".ti. (1)

36	"Non-specific symptoms of borrelia burgdorferi infection in children. [polish] nietypowe objawy zakazenia kretkiem borrelia burgdorferi u dzieci".ti. (0)

37	"[clinical characteristics of lyme disease in 12 cases]".ti. (1)

38	"News in pediatric practice 2001-2002: Infection. [italian] novita pediatria pratica 2001-2002: Infezione".ti. (0)

39	"References: Effective prevention of tick stings. [german] tipps: So beugen sie zeckenstichen wirksam vor".ti. (0)

40	"Advances in the treatment and prevention of lyme borreliosis. [german]".ti. (0)

41	"[ticks and borreliosis--an underestimated danger]".ti. (1)

42	"Lyme borreliose: Diagnostic, therapeutic and preventive approaches. Short text".ti. (1)

43	"The 16th consensus conference in anti-infectious therapeutics of the spilf (french language-speaking society of infectious pathology). Lyme borreliosis: Diagnostic, therapeutic and preventive steps. Long text - wednesday, december 13, 2006".ti. (0)

44	"Suspected borreliosis: What is wrong with the patient?".ti. (0)

45	"[the proof of antineural antibodies in patients with neuroborreliosis]".ti. (1)

46	"Rivm examines antibiotics against lyme: Possibly prophylactic treatment helps. [dutch] rivm onderzoekt antibiotica tegen lyme".ti. (0)

47	"Tug of war to new guideline on lyme disease: Patients associations fall off the cbo. [dutch] touwtrekken om nieuwe richtlijn lyme: Patientenverenigingen vallen cbo af".ti. (0)

48	"Difficult to tame schwer zu zahmen".ti. (0)

49	"[role of transcranial magnetic stimulation in clinical diagnosis: Facial nerve neurography]".ti. (1)

50	"[lyme's serology]".ti. (1)

51	"[perception du risque de maladie de lyme par les résidents d' une communauté infestée par des tiques étoilées américaines]".ti. (0)

52	"[lyme carditis]".ti. and Zimmermann.ax. (1)

53	"[abdominal pain and wall distension as the onset form of neuroborreliosis]".ti. (1)

54	"Seronegative lyme disease: Report of two cases confirmed with pcr".ti. (0)

55	"[diplopia as the first manifestation of lyme disease]".ti. (1)

56	"[seroprevalence of borrelia burgdorferi and tick-borne encephalitis virus in a rural area of samsun, turkey]".ti. (1)

57	"Diagnose in beeld: A boy with nail abnormalities. [dutch] diagn ose in beeld: Een jongen met nagelafwijkingen".ti. (0)

58	"Accident ischemique transitoire revelant une neuroborreliose de lyme. [french] tia revealing lyme neuroborreliosis".ti. (0)

59	"Facial paralysis revealing a neuro-borreliosis. [french] paralysie faciale revelatrice d'une neuroborreliose".ti. (0)

60	"[doxycycline]".ti. and Aupee.ax. (1)

61	"[complete atrioventricular block as the first clinical manifestation of a tick bite (lyme disease)]".ti. (1)

62	"[serology of lyme borreliosis and human granulocytic ehrlichiosis in 2005-2010]".ti. (1)

63	"[upper limb pain in an adolescent]".ti. (1)

64	"[lyme disease in patients with multiple sclerosis: Clinical, diagnostic and therapeutic features]".ti. (1)

65	"[clinical and instrumental characteristics of chronic neuroborreliosis]".ti. (1)

66	"Performance of four commercial immunoblots for serological confirmation of lyme borreliosis in rhone-alpes, france. [french] evaluation des performances de quatre immunoblots commercialises pour la confirmation de la borreliose de lyme dans une population de patients suivis en rhone-alpes".ti. (0)

67	"The role of ceftriaxone in modern antimicrobial therapy - a systematic review of recent clinical trials. [croatian] mjesto ceftriaksona u suvremenoj antimikrobnoj terapiji - sustavni pregled novijih klinickih ispitivanja".ti. (0)

68	"[the public health risk of ticks and tick-borne diseases in the south-eastern poland]".ti. (1)

69	"[neurologic problems in tick-borne diseases--clinical and diagnostic aspects]".ti. (1)

70	"[the information value of detection of borrelia antibodies in the healthy blood donors and in the population at risk]".ti. (1)

71	"Many faces of neuroborreliosis. Case report. [polish] rozne oblicza neuroboreliozy. Opis przypadku".ti. (0)

72	"[fever after a tick bite: Clinical manifestations and diagnosis of acute tick bite-associated infections in northeastern switzerland]".ti. (1)

73	"[optimization of serologic diagnosis of lime borreliosis]".ti. (1)

74	"[lyme borreliosis and demyelinating disease of the central nervous system]".ti. (1)

75	"Antibodies against the causative agents of some natural focal infections in blood donor sera from western slovakia. [czech] protilatky proti povodcom niektorych prirodne ohniskovych nakaz v serach darcov krvi zo zapadneho slovenska".ti. (0)

76	"[therapy-resistant depression with fatigue. A case of presumed streptococcal-associated autoimmune disorder]".ti. (1)

77	"Specifics of cytokine regulation in tick-borne encephalitis and lyme borreliosis".ti. (1)

78	"[5-yr-old with borrelial lymphocytoma]".ti. (1)

79	"[eosinophilic fasciitis associated with borrelia burgdorferi infection]".ti. (1)

80	"Ceffriaxone and pseudocholelithiasis in children. [croatian] ceftriaxon i pseudoholelitijaza u djece".ti. (0)

81	"Progressive facial hemiatrophy without changes in central nervous system and linear scleroderma en coup de sabre with extracerebral tumour - report of two cases. [polish] polowiczy zanik twarzy bez zmian w osrodkowym ukladzie nerwowym i twardzina en coup de sabre z guzem zewnatrzmozgowym - opis przypadkow".ti. (0)

82	"[case reports of borrelia infection in lakartidningen in 1909]".ti. (1)

83	"[pain and neuroborreliosis]".ti. and Berthele.ax. (1)

84	"[two cases of proliferation of monoclonal and monotypic lymphocytes and plasma cells corresponding to acrodermatitis chronica atrophicans]".ti. (1)

85	"Surveillance system for assessing central nervous infections in lower saxony 1998-2000. [german] surveillance-system zur erfassung zentralnervoser infektionen in niedersachsen 1998-2000".ti. (0)

86	"Photoonycholysis caused by doxycycline. [czech] fotoonycholyza zpusobena doxycyklinem".ti. (0)

87	"[neopterin in serum and cerebrospinal fluid in lyme disease]".ti. (1)

88	"[the analysis of epidemiology, clinical symptoms, serological tests in the course of borreliosis]".ti. (1)

89	"[analysis of borrelia burgdorferi genostrains among patients with lyme disease]".ti. (1)

90	"[outbreak of lyme borreliosis with joint manifestation--case report]".ti. (1)

91	"Differential diagnosis of acrodermatitis chronica atrophicans with emphasis on chronic venous insufficiency. [german] differenzialdiagnose der acrodermatitis chronica atrophicans unter besonderer berucksichtigung der chronisch-venosen insuffizienz".ti. (0)

92	"Until a vaccine has been developed against lyme borreliosis, early antibiotic therapy is recommendable. [german] gegen lyme-borreliose bald auch impfen: Bis dahin: Moglichst fruh antibiotika!".ti. (0)

93	"[against lyme borreliosis vaccination also, soon. Until then: Antibiotics as early as possible!]".ti. (1)

94	"Real-time pcr-based in vitro susceptibility testing of borrelia burgdorferi against antimicrobial agents in the presence of eukariotic cells".ti. (0)

95	"[synovectomy of the knee joint]".ti. and Blahut.ax. (1)

96	"[lyme disease acrodermitis chronica atrophicans: Misleading vascular signs]".ti. (1)

97	"Lyme neuroborreliosis: How to not underestimate neurological forms of lyme disease. [french] neuroborreliose de lyme: Comment ne pas meconnaitre une maladie de lyme neurologique".ti. (0)

98	"[acute myelitis and lyme disease]".ti. (1)

99	"[neurosyphilis and neuroborreliosis. Retrospective evaluation of 22 cases]".ti. (1)

100	"[lyme carditis]".ti. and Olszowska.ax. (1)

101	"Multiannual fluctuations of some climate related diseases in poland and their possible relation to climate conditions".ti. (0)

102	"[a reprospective analysis of 973 patients with lyme borreliosis in kuyavian-pomeranian voivodship in 2000-2005]".ti. (1)

103	"[should a lumbar puncture be performed in any child with acute peripheral facial palsy and clinical suspicion of lyme borreliosis?]".ti. (1)

104	"The 46th annual meeting of the german society of occupational and environmental medicine (dgaum)".ti. (0)

105	"Faut-il traiter les symptômes post-borréliose de lyme par des antibiotiques au long cours?".ti. (0)

106	"[the combination of laryngeal paresis in the patients presenting with systemic tick-borne borreliosis (lyme disease)]".ti. (1)

107	"[ocular manifestations of lyme disease]".ti. and Bodaghi.ax. (1)

108	"Rheumatoid arthritis and lyme disease: A case report".ti. (0)

109	"[lyme's disease presenting as increased gamma-gt activity]".ti. (1)

110	"Acrodermatitis chronica atrophicans. Proximal involvement of a lower limb. [french] acrodermatite chronique atrophiante atteinte proximale d'un membre inferieur".ti. (0)

111	"[a curious diving accident happened to a forest walk lover]".ti. (1)

112	"Problemen in de diagnose en behandeling van lyme-borreliose in nederland".ti. (0)

113	"[multiple erythema migrans and facial nerve paralysis: Clinical manifestations of early disseminated lyme borreliosis]".ti. (1)

114	"[inflammatory cardiac diseases by primary extracardial diseases]".ti. (1)

115	"Contagious and parasitic occupational diseases in the czech republic. [czech] nemoci z povolani prenosne a parazitarni v ceske republice".ti. (0)

116	"[occupational infectious diseases in the czech republic]".ti. (1)

117	"Lyme borreliosis of the musculoskeletal system".ti. (0)

118	"[multiple erythema]".ti. and Brunner.ax. (1)

119	"[human granulocytic ehrlichiosis co-incident with lyme borreliosis in pregnant woman--a case study]".ti. (1)

120	"[an overlooked disease of tick bites: Lyme disease]".ti. (1)

121	"[features of cytokine levels in serum of patients with tick-borne borreliosis with different clinical signs]".ti. (1)

122	"Recurring focal neurologic events and cognitive impairment in a 52-years-old male".ti. (0)

123	"[lyme disease in childhood: An imported disease?]".ti. (1)

124	"[prevalence of antibodies to borrelia burgdorferi in sardinian teen-agers]".ti. (1)

125	"[lyme carditis--rare cause of dilated cardiomyopathy and rhythm disturbances]".ti. (1)

126	"Rheumatology and dermatology: Many interconnections, immense border areas. [italian] reumatologia e dermatologia: Molte le interconnessioni, vasti i grenzgebiete ".ti. (0)

127	"Acute polyradiculoneuropathy and lyme disease. [portuguese] polirradiculoneurite aguda e doenca de lyme-simile".ti. (0)

128	"Lyme disease. [turkish] lyme hastaligi".ti. (0)

129	"[back pain without radiculitis as an initial manifestation of lyme disease: Two cases]".ti. (1)

130	"[the new possibilities in serologic diagnostic of ixodes mite-borne borreliosis using immunochip]".ti. (1)

131	"[radiculalgia revealing lyme disease]".ti. (1)

132	"[clinical and laboratory characterization of ixodes tick-borne borreliosis in the baikal area]".ti. (1)

133	"Vaccine recommendations for infants and children travelers. [spanish] vacunacion del nino en viajes internacionales".ti. (0)

134	"[prevalence of borrelia burgdorferi antibodies in healthy population in poland]".ti. (1)

135	"Lyme borreliosis: Diagnostic, therapeutic and preventive steps".ti. (0)

136	"16th consensus conference in anti-infectious therapy. Lyme borreliosis: Preventive, therapeutic and diagnosis management. [french] 16<sup>e</sup> conference de consensus en therapeutique anti-infectieuse borreliose de lyme: Demarches diagnostiques, therapeutiques et preventives".ti. (0)

137	"Lyme borreliosis and co-infections. Place of anaplasma phagocytophilum and bartonella henselae".ti. (1)

138	"Epidemiological data on the spread of lyme disease in abruzzo. [italian] dati epidemiologici sulla diffusione in abruzzo della malattia di lyme in personale a rischio".ti. (0)

139	"Rapid diagnostic tests for infectious diseases. [turkish] enfeksiyon hastaliklarinda hizli tani testleri".ti. (0)

140	"[arthritis and infections]".ti. and Cimaz.ax. (1)

141	"[incidence of tick-borne encephalitis virus and borrelia burgdorferi infections in farmers of the lublin province]".ti. (1)

142	"Lyme borreliosis in people in slovakia during 1999-2001".ti. (0)

143	"The incidence of some zoonoses in the slovak republic in 2009".ti. (0)

144	"[neuroborreliosis in patient with aplastic anaemia secondary to therapy with ticlopidine]".ti. (1)

145	"[borrelia-associated lymphocytoma cutis]".ti. and Corcostegui.ax. (1)

146	"[neuroborreliosis after an early lyme disease treated by azithromycin]".ti. (1)

147	"Antibodies specific to borrelia burgdorferi in wild animal workers".ti. (0)

148	"Polyneuritis cranialis: Portuguese hospital case series. [portuguese] polineuropatia craniana - uma serie hospitalar".ti. (0)

149	"[a case of neuroborelliosis complicated by acute adrenal insufficiency]".ti. (1)

150	"A lyme patient about her disease".ti. (0)

151	"[neuroborreliosis]".ti. and Cuvelier.ax. (1)

152	"[ultrasonography in diagnosing lyme arthritis of the knee joints in correlation with anti-ccp antibodies]".ti. (1)

153	"[usefulness of examinations of serum levels of matrix metalloproteinases 1, mmp-3, mmp-9, tissue inhibitor of metalloproteinases 1, hyaluronic acid and antibodies against cyclic citrullinated peptide in lyme arthritis, rheumatoid arthritis and patients with arthritic complaints]".ti. (1)

154	"[neuroborreliosis--a disease with many faces]".ti. (1)

155	"[clinical forms of neuroborreliosis among hospitalized patients in the years 2000-2005]".ti. (1)

156	"Clinical and evolutionary characteristics of four patients with pulmonary histoplasmosis reported in the paraiba paulista valley region. [portuguese] caracteristicas clinicas evolutivas de quatro pacientes com histoplasmose pulmonar encontrado na regiao do vale do paraiba".ti. (0)

157	"Lyme borreliosis - differential diagnostic difficulties in the interpretation of serological results. [croatian] lyme borelioza - diferencijalno dijagnosticke poteskoce u interpretaciji seroloskih rezultata".ti. (0)

158	"Difficulties in neuroborreliosis diagnostics".ti. (0)

159	"Lyme borreliosis as a cause of bilateral neuroretinitis with pronounced unilateral stellate maculopathy in a 8-year old girl. [slovak] lymeska borelioza jako pricina bilateralni neuroretinitidy s vyraznou jednostrannou hvezdicovitou makulopatii u osmilete divky".ti. (0)

160	"[lyme borreliosis in bosnia and herzegovina--clinical, laboratory and epidemiological research]".ti. (1)

161	"[borrelial erythema of the face]".ti. (1)

162	"Histopathological study of erythema chronicum migrans cases diagnosed in manaus. [portuguese, english] estudo histopatologico de casos de eritema cronico migratorio diagnosticados em manaus".ti. (0)

163	"Two specific manifestations of lyme disease. [dutch] twee bijzondere uitingsvormen van de ziekte van lyme".ti. (0)

164	"The significance of borrelia burgdorferi dna investigation in a diagnostic algorithm of lyme arthritis. [czech] vyznam vysetreni dna borrelia burgdorferi v diagnostickem algoritmu lymeske artritidy".ti. (0)

165	"[doubling of the number of cases of tick bites and lyme borreliosis seen by general practitioners in the netherlands]".ti. (1)

166	"[the lyme carditis as a rare differential diagnosis to an anterior myocardial infarction]".ti. (1)

167	"[sero-epidemiological studies of zoonotic infections in hunters--comparative analysis with veterinarians, farmers, and abattoir workers]".ti. (1)

168	"[seroepidemiological studies of zoonotic infections in hunters in southeastern austria--prevalences, risk factors, and preventive methods]".ti. (1)

169	"Lyme disease (lyme borreliosis)".ti. and Cabadaj.ax. (0)

170	"Linear and localized scleroderma in plaques in a patient. [spanish] esclerodermia localizada en placas y lineal en un paciente".ti. (0)

171	"[erythema chronicum migrans. A case report]".ti. (1)

172	"Multiple gyrated erythemas in stage iii borreliosis".ti. (0)

173	"[tick-borne infections]".ti. and Dobler.ax. (1)

174	"[epidemiology of borreliosis in workers of the district forestry offices in lower silesia]".ti. (1)

175	"A small turn then goes back.. [french] un petit tour et puis revient".ti. (0)

176	"[acute severe leukoencephalitis with posterior lesions due to borrelia burgdorferi infection]".ti. (1)

177	"[tick-borne borreliosis in the iaroslavl region]".ti. (1)

178	"[natural focal infections transmitted by ixodes ticks in the iaroslavl region: Epidemiological aspects]".ti. (1)

179	"Detection of serum antibodies against flagellin with a protein biochip in patients with lyme borreliosis".ti. (0)

180	"Serological epidemiology investigation of lyme disease in henan province".ti. (0)

181	"New knowledge about treatment of disseminated lyme borreliosis. [slovak] novsie poznatky o liecbe diseminovanej formy lymskej boreliozy".ti. (0)

182	"[borreliosis in children - clinical manifestation, diagnosis and treatment]".ti. (1)

183	"[seroprevalence of anaplasma phagocytophilum in patients with suspected lyme borreliosis]".ti. (1)

184	"[borreliosis--increasing clinical problem]".ti. (1)

185	"[prevalence of borreliosis among forestry workers in kujawsko-pomorskie voivodeship]".ti. (1)

186	"Zoonoses in the republic of croatia".ti. (0)

187	"Value in practice of the cerebrospinal fluid in treponema pallidum hemagglutination test for confirmation of neurosyphilis".ti. (0)

188	"[when is a diagnosis of borreliosis advisable?]".ti. (1)

189	"[complete heart block and lyme disease: Report of two different cases and literature review]".ti. (1)

190	"[diseases produced by borrelia]".ti. (1)

191	"[using the polymerase chain reaction to borrelia burgdorferi infection in localized scleroderma injure (morphea), in venezuelan patients]".ti. (1)

192	"[erythema migrans and lyme disease]".ti. and Fain.ax. (1)

193	"The importance of serologic testing for lyme disease, in case of atrioventricular heart block. [french] interet des serologies de lyme devant un bloc auriculoventriculaire".ti. (0)

194	"[in fibromyalgia antibiotic therapy is not indicated]".ti. (1)

195	"[child fever in tropical area: Don't forget borreliosis]".ti. (1)

196	"Antibiotic therapy of head and neck infections. [german] antibiotikatherapie der infektionen an kopf und hals".ti. (0)

197	"Lyme's disease in persons occupationally exposed to ticks - an evaluation according to occupational medical and accident insurance legislation on the basis of a case report. [german] lyme-borreliose bei beruflich zecken-exponierten personen - eine arbeitsmedizinisch-unfallversicherungsrechtliche bewertung anhand einer kasuistik".ti. (0)

198	"[unilateral mydriasis in lyme neuroborreliosis]".ti. (1)

199	"[erythema migrans due to borrelia afzelii]".ti. (1)

200	"Neuroborreliosis in a patient with presumed cryptogenic ischaemic strokes. [german] neuroborreliose bei einer patientin mit vermeintlich kryptogenen zerebralen infarkten".ti. (0)

201	"[occurrence of erythema migrans in hajnowka district in years 2004-2009]".ti. (1)

202	"[combined manifestations of natural foci in the territory of the vologda province]".ti. (1)

203	"Lyme borreliosis in children. Epidemiology, diagnosis, clinical treatment, and therapy".ti. (1)

204	"Lyme borreliosis in infancy and adolescence".ti. (0)

205	"[tick removed. Sending on to examination?]".ti. (1)

206	"Diagnosis and therapy of borreliosis. [german] diagnose und therapie der borreliose".ti. (0)

207	"Is chronic fatigue syndrome in reality sometimes chronic lyme disease?. [dutch] is het chronischevermoeidheidssyndroom in werkelijkheid soms chronische lymeziekte?".ti. (0)

208	"Analysis of the clinical course of lyme borreliosis in respect to the initial disease manifestation in białowieża residents".ti. (0)

209	"[detection of dna of borrelia circulating in novosibirsk region]".ti. (1)

210	"[detection of borrelia dna in the borrelia burgdorferi sensu lato complex in the blood of patients with ixodes tick-borne borrelios]".ti. (1)

211	"[concentration of svap in serum of patients with different forms of lyme borreliosis]".ti. (1)

212	"[lyme carditis and symptomatic sinus node dysfunction]".ti. (1)

213	"Increased intracranial pressure due to neuroborreliosis. [german] pseudotumor cerebri bei neuroborreliose".ti. (0)

214	"A cutaneous manifestation of borreliosis. [french] une manifestation cutanee de la borreliose".ti. (0)

215	"Differential diagnosis of attention-deficit hyperactivity disorder in adolescents and adults - case report of an 18-year-old woman with neuroborreliosis. [german] differenzialdiagnostische abklarung der aufmerksamkeitsdefizit-/hyperaktivitatsstorung im jugendlichen- und erwachsenenalter - fallbeispiel einer 18 jahre alten patientin mit neuroborreliose".ti. (0)

216	"Assumed and real consequences of a tick bite: One willingly considers as borreliosis what one cannot explain. [german] vermeintliche und echte folgen eines zeckenstichs: Was man nicht erklaren kann, sieht man gern als borreliose an ".ti. (0)

217	"Mental problems or infection: Is there a chronic borreliosis despite therapy?. [german] psychische probleme oder infektion: Gibt es eine chronische borreliose trotz therapie?".ti. (0)

218	"[after the tick bite: Update borreliosis 2012]".ti. (1)

219	"Chronic borreliosis. Do combinations with hydroxychloroquine help?. [german] chronische borreliose. Helfen kombinationen mit hydroxychioroquin?".ti. (0)

220	"[bilateral tonic pupil as the only remaining ophthalmic sign of lyme disease: Case report]".ti. (1)

221	"[cutaneous marginal zone lymphoma (salt) and infection with borrelia burgdorferi]".ti. (1)

222	"[lyme borreliosis--diagnostic difficulties in interpreting serological results]".ti. (1)

223	"[early recurrence of neuroborreliosis with a fatal outcome]".ti. (1)

224	"[the swollen knee - differential diagnosis and therapy concepts from the pediatric rheumatologic perspective]".ti. (1)

225	"Clinical forms of tick bite. [spanish] picadura de garrapata: Formas clinicas".ti. (0)

226	"Ecopathological evidence and diagnosis of zoonotic diseases in the elderly".ti. (0)

227	"[pseudotumour cerebri in a patient with lyme disease and hypothyroidism]".ti. (1)

228	"Epidemiology of newly emerging and re-emerging zoonoses in bulgaria in the last decade".ti. (0)

229	"[diagnosis and treatment of lyme arthritis. Recommendations of the pharmacotherapy commission of the deutsche gesellschaft fur rheumatologie (german society for rheumatology)]".ti. (1)

230	"[acute transverse myelitis and lyme borreliosis: A case report]".ti. (1)

231	"Epidemiological investigation on lyme disease in changbai and tonghua county, jilin province".ti. (0)

232	"Anti-borrelia burgdorferi antibody response in 827 patients with suspected lyme disease".ti. (0)

233	"Borrelia burgdorferi antibody levels in 105 patients with suspected lyme disease".ti. (0)

234	"The first case of lyme disease in shanxi province, china: A case study".ti. (0)

235	"Tick vector-borne diseases: Human risk".ti. (0)

236	"[diseases and pathogenic agents transmitted by ticks in switzerland]".ti. (1)

237	"Neuroborreliose bij kinderen, herkenning en diagnostiek. [dutch] neuroborreliosis in children: Recognizing and diagnosing".ti. (0)

238	"[optic neuritis]".ti. and Wermund.ax. (1)

239	"[alcohol and epilepsy: A case report between alcohol withdrawal seizures and neuroborreliosis]".ti. (1)

240	"Cooperation against work-related zoonoses".ti. (0)

241	"[a short journey through spirochetal infections]".ti. (1)

242	"[facial nerve paresis]".ti. and Glocker.ax. (1)

243	"[ocular manifestations of lyme borreliosis in northwest croatia]".ti. (1)

244	"[diaphragmatic paralysis and arthromyalgia caused by lyme disease]".ti. (1)

245	"[interferon inductors in treatment of associative forms of tick-borne infection in children]".ti. (1)

246	"[seroepidemiologic study of lyme's borreliosis in mexico city and the northeast of the mexican republic]".ti. (1)

247	"Cranial polyneuritis in a case of neuroborreliosis with asymptomatic enhancement of cranial nerves. [german] polyneuritis cranialis: Neuroborreliose mit asymptomatischem hirnnervenenhancement".ti. (0)

248	"Facial palsies and neuroborreliosis in childhood. [german] fazialisparesen bei neuroborreliosen im kindesalter".ti. (0)

249	"Chronic lyme-disease as a chameleon of medicine: Burden of a special patient group".ti. (0)

250	"[idiopathic facial palsy]".ti. and Grosheva.ax. (1)

251	"[re-infection with borrelia burgdorferi s.L in a patient with a history of neuroborreliosis--case report]".ti. (1)

252	"[failures of antibiotic treatment in lyme arthritis]".ti. (1)

253	"[delusional disorders in the course of tick-born encephalitis and borreliosis in patients with hemophilia a and posttraumatic epilepsy--diagnostic and therapeutic difficulties]".ti. (1)

254	"Seroepidemiological investigation of lyme disease in a part of forest region of xiao hinggan mountains".ti. (0)

255	"[reversible complete heart block by re-infection with borrelia burgdorferi with negative igm-antibodies]".ti. (1)

256	"[lyme borreliosis: Census of adult patients hospitalized in indre-et-loire (france), from the hospital discharge data (1999-2006)]".ti. (1)

257	"[seroprevalence of tick-borne encephalitis virus (tbev) among the residents of rural areas in sinop, central black-sea region, turkey]".ti. (1)

258	"Clinical characteristics of patients with intermediate uveitis".ti. (0)

259	"Cervical radiculitis as initial clinical feature of neuroborreliosis - case reports. [slovak] cervikalna radikulitida ako inicialny klinicky prejav neuroboreliozy - kazuistiky".ti. (0)

260	"[chronic fatigue syndrome following tick-borne diseases]".ti. (1)

261	"[lyme disease: Basis for treatment strategy, primary preventive care and secondary preventive care]".ti. (1)

262	"Epidemiological situation as to selected zoonoses in the slovak republic in 2007".ti. (0)

263	("Practice parameter: Treatment of nervous system lyme disease" and "report of the quality standards subcommittee of the american academy of neurology").ti. and Halperin.ax. (1)

264	"[juvenile idiopathic arthritis and lyme disease: A case report]".ti. (1)

265	"[suspected borreliosis - what's behind it?]".ti. (1)

266	"[diagnosing lyme disease: The role of clinical and complementary examination]".ti. (1)

267	"[borrelia burgdorferi]".ti. and Hashimoto.ax. and "2010".yr. (1)

268	"Stage-directed therapy of lyme borreliosis".ti. (0)

269	"[a case of suspected borreliosis]".ti. (1)

270	"Treatment after tick bites - problematics of borrelia infection".ti. (0)

271	"[purplish-erythematous plaque of the lower limb]".ti. (1)

272	"[polymorphic mental disorders in the course of lyme borreliosis--case study]".ti. (1)

273	"Lyme borreliosis during pregnancy: Consequences for the newborn?. [dutch] lyme-borreliose in de zwangerschap: Gevolgen voor de pasgeborene?".ti. (0)

274	"[borrelia burgdorferi myocarditis]".ti. (1)

275	"A case with an edge. [german] ein fall mit biss".ti. (0)

276	"Borrelia infections: Lyme disease and recurrent fever. [spanish] infecciones producidas por borrelias: Enfermedad de lyme y fiebre recurrente".ti. (0)

277	"[lyme borreliosis]".ti. and Herzer.ax. and "2005".yr. (1)

278	"[45-year-old woman with joint pain and suspected lyme arthritis]".ti. (1)

279	"[lyme borreliosis]".ti. and Herzer.ax. and "2014".yr. (1)

280	"Report from usa. [german] bericht aus usa".ti. (0)

281	"Clinical and therapeutic aspects of primary cutaneous marginal zone b-cell lymphoma. [dutch] klinische en therapeutische aspecten van het primair cutane marginale zone b-cel lymfoom".ti. (0)

282	"[early diagnosis of lyme borreliosis. Do not look only for erythema migrans]".ti. (1)

283	("[early diagnosis of lyme borreliosis]".ti. and Hofmann.ax.) not "Do not look only for erythema migrans".ti. (1)

284	"[the variable spectrum of cutaneous lyme borreliosis. Diagnosis and therapy]".ti. (1)

285	"[early summer meningoencephalitis and lyme borreliosis. Relevant health risks in north bavaria]".ti. (1)

286	"[comatose state--think different]".ti. (1)

287	"Neuro-ehrlichiosis: A new tick-borne disease with nervous system involvement?".ti. (0)

288	"Analysis of imported lyme disease during beijing olympics".ti. (0)

289	"Human granulocytic anaplasmosis in bulgaria - confirmed cases and review of the literature".ti. (0)

290	"Prevalence of borrelia burgdorferi sensu lato species among patients in the czech republic; direct sequencing analysis and real-time polymerase chain reaction. [czech] prevalence druhu borrelia burgdorferi sensu lato u pacientu v ceske republice; prima sekvenc ni analyza a polymerazova rete zova reakce v realnem case".ti. (0)

291	"Erratum: Haufigste spate manifestation der lyme-borreliose (monatsschrift fur kinderheilkunde b 151:11 (1156-1162)). [german]".ti. (0)

292	"Lyme borreliosis (borrelia burgdorferi). [german] lyme-borreliose (borrelia burgdorferi)".ti. (0)

293	"One sided reddish-livid swelling of the earlobe - atheroma?".ti. (0)

294	"Rational diagnostic strategies for lyme borreliosis. [german] rationale diagnostik der lyme-borreliose".ti. (0)

295	("[lyme borreliosis in children and adolescents]".ti. and Huppertz.ax.) not practice guideline.pt. (1)

296	"[lyme borreliosis]".ti. and Huppertz.ax. and "2003".yr. (1)

297	"[clinical analysis of patients with peripheral facial palsy]".ti. (1)

298	"[a case of lyme disease requiring over 1 year to diagnose at an infectious-disease clinic]".ti. (1)

299	"Pediyatrik morfea (lokalize skleroderma) 14 olguya ait epidemiyolojik klinik ve laboratuvar bulguları".ti. (0)

300	"[inf-gamma and gm-csf concentrations in serum of patients with lyme disease]".ti. (1)

301	"[reddish-blue color lower leg with distinct vessel design. Venous insufficiency was lyme borreliosis]".ti. (1)

302	"[small risk of developing lyme borreliosis following a tick bite on ameland: Research in a general practice]".ti. (1)

303	"Nurse activities in accompanying children with lyme borreliosis".ti. (0)

304	"[synovitis score: Value of histopathological diagnostics in unclear arthritis. Case reports from rheumatological pathological practice]".ti. (1)

305	"[the role of pcr for the diagnosis of lyme arthritis]".ti. (1)

306	"Problems of false positive laboratory tests during qualification to the program of severe plaque psoriasis treatment on the basis of two cases. [polish] problemy falszywie dodatnich testow laboratoryjnych podczas kwalifikacji do programu leczenie ciezkiej postaci luszczycy plackowatej na podstawie dwoch przypadkow".ti. (0)

307	"[multilocular large erythemas and fever]".ti. (1)

308	"Unusual cutaneous borreliosis in alsace region. [french] la borreliose cutanee en alsace, dans tous ses etats".ti. (0)

309	"Bell palsy. Diagnostic and therapy. [german] idiopathische fazialisparese. Diagnostik und therapie".ti. (0)

310	"[cardiac arrest due to torsades de pointes ventricular tachycardia in a patient with lyme carditis.]".ti. (1)

311	"A multiplex fluorescent quantitative pcr method for detection of three rodent-carrying pathogens".ti. (0)

312	"Lyme disease in alsace: Incidence of a re-emerging disease. [french] incidence d'une maladie reemergente: La borreliose de lyme en alsace".ti. (0)

313	"The course of lyme disease in children - own observation. [polish] przebieg boreliozy u dzieci - obserwacje wlasne".ti. (0)

314	"[mental disorders in the course of lyme borreliosis and tick borne encephalitis]".ti. (1)

315	"[screening for antibodies against zoonotic agents among employees of the zoological garden of vienna, schonbrunn, austria]".ti. (1)

316	"Antibiotic therapy of lyme disease is criticized: Long-term therapy has no benefits, only side effects arzneimittel und therapie: Antibiose bei borreliose in der kritik: Langzeittherapie hat keinen nutzen, dafur nebenwirkungen".ti. (0)

317	"[a man with acutely starting pain in the right hip and thigh]".ti. (1)

318	"[complete atrio-ventricular block as a first symptom of borreliosis: Antibiotic treatment instead of pacemaker implantation]".ti. (1)

319	("Case reports" and "other association").ti. (0)

320	"[lyme disease from an ophthalmological point of view]".ti. (1)

321	"[psoriasis as koebner phenomenon in erythma migrans]".ti. (1)

322	"[clinical courses of acute and chronic neuroborreliosis following treatment with ceftriaxone]".ti. (1)

323	"[neuroborreliosis and diphasic meningoencephalitis -- common features and differences]".ti. (1)

324	"Cardiac arrest as atypical manifestation of early borreliosis - case report. [polish] nagle zatrzymanie krazenia jako nietypowy objaw wczesnej boreliozy - opis przypadku".ti. (0)

325	"Lyme borreliosis - not only endemic area disease - case report. [polish] borelioza - choroba nie tylko obszarow endemicznych - opis przypadku".ti. (0)

326	"Epidemiological aspects of the incidence of lyme borreliosis in košice region".ti. (0)

327	"Observation of the incidence of anti-anaplasma phagocytophilum antibodies in the risky groups of patients".ti. (0)

328	"Occurrence of zoonoses in the norwegian population".ti. (0)

329	"[usage of real time polymerase chain reaction for diagnostics of different tick-borne infections]".ti. (1)

330	"[isolation of the recombinant proteins ospc and the fragment flaa (f-flaa) from the western-siberian borrelia garinii nt29 isolates and the study of their immunochemical properties]".ti. (1)

331	"[lyme disease]".ti. and Kawabata.ax. and "2003".yr. (1)

332	"A case of lyme disease with erythema migrans. [japanese]".ti. (0)

333	"[case of borrelia brainstem encephalitis presenting with severe dysphagia]".ti. (1)

334	"[evaluation of the usefulness cerebrospinal fluid myelin basic protein (mbp) concentration examination in patients with lyme neuroborreliosis--preliminary study]".ti. (1)

335	"[evaluation of cerebrospinal fluid serotonin (5-ht) concentration in patients with post-lyme disease syndrome--preliminary study]".ti. (1)

336	"[cranial neuritis with enhancement on post-contrast mri due to lyme disease]".ti. (1)

337	"[exposure to ticks and erythema chronicum migrans among borreliosis patients in lower silesia]".ti. (1)

338	"[eosinophilic fasciitis associated with borrelia afzelii infection]".ti. (1)

339	"Peripheral retinal vasculitis associated with lyme disease".ti. (0)

340	"A case of lyme disease with unusual cutaneous manifestations. [korean]".ti. (0)

341	"[acute urinary retention in a young woman with lyme borreliosis.]".ti. (1)

342	"A case of lyme disease. [japanese]".ti. (0)

343	"[preliminary studies on presence of antineuronal antibodies in serum of patients with lyme borreliosis]".ti. (1)

344	"[ambiguous swelling of the earlobe]".ti. (1)

345	"[prevalence of lyme disease among forestry workers]".ti. (1)

346	"[picture of the month: Erythema]".ti. and Kofoed.ax. (1)

347	"Occupational infectious diseases in the west bohemian region in 1996-2000. [czech] profesionalni infekcni nemoci v zapadoceskem regionu v letech 1996-2000".ti. (0)

348	"[neuroimmune aspects of the pathogenesis and nitric oxide negative effects modifying the pathogenetic treatment of tick-borne infections]".ti. (1)

349	"[the polymerase chain reaction evaluation of borrelia burgdorferi dna presence in peripheral blood of patients with lyme disease]".ti. (1)

350	"[ehrlichia chaffeensis antibodies prevalence among patients bitten by ticks]".ti. (1)

351	"[effect of antibiotic therapy on levels of proinflammatory cytokines: Interleukin il-1, il-6 and tumor necrosis factor tnf-alpha in serum of patients with lyme borreliosis]".ti. (1)

352	"Fibrous nodules in lyme borreliosis. [czech] fibrozni uzly u lymeske boreliozy".ti. (0)

353	"[unclassified cardiomyopathy or lyme carditis? A three year follow-up]".ti. (1)

354	"[severe diarrhoea with invasive intestinal spirochaetosis]".ti. (1)

355	"Lyme disease in the ivory coast: A serological survey in abidjan. [french] borreliose de lyme en cote-d'ivoire: Mythe ou realite? Une enquete transversale de seropre valence a abidjan".ti. (0)

356	"[a japanese case of neuroborreliosis with papillitis]".ti. (1)

357	"Inflammatory dysfunctions of the facial nerve. [slovak] zapalove dysfunkcie tvaroveho nervu".ti. (0)

358	"Bullous localized scleroderma - presentation of two cases, therapeutic difficulties. [polish] twardzina ograniczona pecherzowa - przedstawienie dwoch przypadkow, trudnosci terapeutyczne".ti. (0)

359	"[neuroborreliosis in north jutland]".ti. (1)

360	"[early diagnosis of lyme arthritis]".ti. (1)

361	"[comparison of eia borrelia recombinant igm of the 3rd generation with eia test based on whole cell antigen in the diagnosis of lyme borreliosis]".ti. (1)

362	"[eye involvement of borrelia aetiology]".ti. (1)

363	"[multilocular erythema migrans in borreliosis]".ti. (1)

364	"[laboratory diagnosis of lyme borreliosis]".ti. and Kristiansen.ax. (1)

365	"[the rate of detection and diagnostic significance of antibodies to borrelia burgdorferri in patients with eyes diseases of inflammatory nature]".ti. (1)

366	"[acrodermatitis chronica atrophicans: Late manifestation of lyme borreliosis]".ti. (1)

367	"[a man with a swollen nipple]".ti. (1)

368	"[risk for infection by lyme disease cause in green surfaces maintenance workers in belgrade]".ti. (1)

369	"Multiple foci of recurrent benign lymphoplasia in lyme disease".ti. (0)

370	"Early disseminated lyme borreliosis".ti. and Kryazheva.ax. (0)

371	"[coexistence of two occupational infectious diseases in one patient--borreliosis and tick-borne encephalitis--a case report]".ti. (1)

372	"Comparison of results of measurements of visual evoked potentials in patients with multiple sclerosis and neuroborreliosis".ti. (0)

373	"Comprehensive biopsy-based diagnosis of inflammatory cardiomyopathy as a cause of new-onset heart failure of unclear etiology: Initial experience. [czech] komplexni biopticka diagnostika zane tlive kardiomyopatie jako priciny nove vznikleho srdecniho selhani nejasne etiologie: Prvni zkusenosti".ti. (0)

374	"Inflammatory cardiomyopathy secondary to borrelia infection, without a systemic serologic response".ti. (0)

375	"[clinical spectrum and incidence of neuro-borreliosis in the netherlands]".ti. (1)

376	"Early recurrence of a late lyme carditis: Successful treatment with roxithromycin and co-trimoxazole".ti. (0)

377	"Lyme disease: Current diagnosis and therapy".ti. (0)

378	"[comparative analysis of methods for laboratory diagnosis of lyme's disease in early stages of infection]".ti. (1)

379	"Role of borrelia burgdorferi in the development of morphea".ti. (0)

380	"Nipple disorders in children. [dutch] tepelafwijkingen bij kinderen".ti. (0)

381	"[the recurrence of lyme arthritis]".ti. (1)

382	"Dynamics of the polymerase chain reaction (pcr) and its value in lyme borreliosis. [czech] dynamika polymerazove retezove reakce (pcr) a jeji vyznam v diagnostice lymeske neuroboreliozy".ti. (0)

383	"[neuro-ophthamologic manifestations as unusual presentation of lyme disease]".ti. (1)

384	"Unusual finding in peripheral blood. [spanish] hallazgo infrecuente en sangre periferica".ti. (0)

385	"[lyme borreliosis--the most frequent vector-borne infection in denmark]".ti. (1)

386	"[lyme carditis--a bitter lesson or a delayed diagnostic success--a case report]".ti. (1)

387	"[borreliosis--simultaneous lyme carditis and psychiatric disorders--case report]".ti. (1)

388	"[lyme arthritis in children: A diagnostic trap]".ti. (1)

389	"[facial diplegia as the presenting feature of lyme disease]".ti. (1)

390	"[electromagnetic fields of extremely high frequency and radon baths in the treatment of lyme disease with nervous system lesions]".ti. (1)

391	"A case of selective paresis of the deep stabilization system as a result of borreliosis. [czech] pripad selektivni parezy hlubokeho stabilizacniho systemu jako nasledek boreliozy".ti. (0)

392	"Research advancement in action of cytokines in lyme disease. [chinese]".ti. (0)

393	"Study on tick-borne disease in northwest of china".ti. (0)

394	"Serological survey of lyme disease in some forest areas in shannxi province, china".ti. (0)

395	"Investigation of lyme disease natural epidemic focus in shaanxi province".ti. (0)

396	"Frequent incidence of lyme neuroborreliosis in children in the czech republic. [czech] casty vyskyt lymeske neuroboreliozy u deti v ceske republice".ti. (0)

397	"[the level of knowledge of general practitioners regarding the early phase of lyme borreliosis. Survey conducted among 106 general practitioners]".ti. (1)

398	"[ixodes tick-borne borreliosis and tick-borne encephalitis in the udmurt republic: Retrospective analysis of their spread]".ti. (1)

399	"[long-term morbidity and spatial distribution of tick-borne encephalitis and ixodes tick-borne borreliosis in udmurtia]".ti. (1)

400	"Serum epidemiological investigation on anaplasma phagocytophilum in population of the north forest area in fujian province".ti. (0)

401	"Surveillance and study of tick-borne diseases in zhejiang province, china".ti. (0)

402	"[invasion by trichinae in the patient hospitalized with suspicion of occupational borreliosis: A case report]".ti. (1)

403	"Anetoderma - a predictor of the occurrence of systemic autoimmunologic diseases. [polish] anetodermia - zwiastun wystapienia ukladowych shorzen autoimmunologicznych".ti. (0)

404	"Preliminary epidemiological investigation of lyme disease in hunan province".ti. (0)

405	"A seroepidemiological investigation of lyme disease in the shawan forest region of xinjiang, china".ti. (0)

406	"[study on the relation of transmission of lyme disease and ecological protection from western regions in china]".ti. (1)

407	"[ticks bite in foresters]".ti. (1)

408	"[neuroborreliosis in vest-agder]".ti. (1)

409	"Rare cutaneous signes of lyme disease in lorraine region".ti. (0)

410	"[differential diagnostics of neuroinfections: Lyme disease]".ti. (1)

411	"Cold, erythematous-violaceous macules on both breasts of a 66-year-old woman. [spanish] maculas eritemato-violaceas frias en ambas mamas de una paciente de 66 anos".ti. (0)

412	"Htlv-1 infection and alzheimer-type dementia. [spanish] infeccion por htlv-1 y demencia tipo alzheimer".ti. (0)

413	"[lyme disease. A tick bite, no panic! Emerging zoonoses in quebec]".ti. (1)

414	"[once again lyme disease]".ti. (1)

415	"Development of an indicator-based early warning system for lyme disease outbreaks".ti. (0)

416	"Neuroborreliosis. [croatian] neuroborelioza".ti. (0)

417	"[ usefulness of laboratory investigations for the diagnosis of acrodermatitis chronica atrophicans. ]".ti. (1)

418	"[onset of leber's hereditary optic neuropathy in association with borreliosis]".ti. (1)

419	"[atypical form of lyme disease]".ti. (1)

420	"[epidemiologic situation on dangerous infectious diseases on the territory of republic of abkhazia]".ti. (1)

421	"[lyme disease disclosed by pseudoneoplastic weight loss]".ti. (1)

422	"[preparations for serodiagnosis of diseases due to causative agents of ixode tick-borne borreliosis (lyme disease). Communication 1. Comparative study of the 1st generation enzyme immunoassay test-systems for detection of antibodies to borrelia burkdorferi sensu lato]".ti. (1)

423	"[preparations for serodiagnosis of diseases due to causative agents of ixodes tick-borne borreliosis (lyme disease). Communication 3. Comparative study of enzyme immunoassay test-systems for detection of igm to borrelia burgdorferi sensu lato]".ti. (1)

424	"[preparations for serodiagnosis of diseases due to causative agents of ixode tick-borne borreliosis (lyme disease). Communication 2. Comparative study of recombinant enzyme immunoassay test-systems (relisa) for serological diagnosis of ixode tick-borne borreliosis]".ti. (1)

425	"Acrodermatitis chronica atrophicans".ti. and Bottlaender.ax. (0)

426	"First isolation of borrelia sp. (borrelia afzelii) from cerebrospinal fluid in a patient with neuroborreliosis in croatia. [croatian] prva izolacija borelije (borrelia afzelii) iz likvora bolesnika s neuroboreliozom u hrvatskoj".ti. (0)

427	"First isolation of borreliae garinii in croatia. [croatian] prva izolacija borreliae garinii u hrvatskoj".ti. (0)

428	"Azithromycin in the treatment of erythema migrans. [croatian] azitromycin u lijecenju erythema migrans".ti. (0)

429	"Fast onset cognitive impairment: Neuroborreliosis and positive 14-3-3 protein deterioro cognitivo de rapida instauracion: Neuroborreliosis y proteina 14-3-3 positiva".ti. (0)

430	"[red lower limb]".ti. (1)

431	"[psoriasiform exanthem in a patient with men-i]".ti. (1)

432	"[may lyme borreliosis lead to heart transplantation? - a case report]".ti. (1)

433	"[recurrent nerve palsy due to lyme disease: Report of two cases]".ti. (1)

434	"Risks of biotic agents".ti. (0)

435	"Characteristics of borrelia associated with lyme disease diagnosed in okinawa where lyme disease was not expected".ti. (0)

436	"[lyme uveitis: 2 case reports]".ti. (1)

437	"[red circles over the whole body]".ti. (1)

438	"[erythema chronicum migrans]".ti. and Maurin.ax. (1)

439	"A case of a 14-year-old-girl with atrophoderma pasini-pierini. [polish] przypadek atrophodermii pasini-pierini u 14-letniej dziewczynki".ti. (0)

440	"[infectious diseases in poland in 2000]".ti. (1)

441	"[the onset of chronic lyme-borreliosis after the cure of tick-borne rickettsia infection in simultaneous infection]".ti. (1)

442	"[sensorineural loss of hearing in lower registers as the main symptom of lyme disease]".ti. (1)

443	"[spinal cord stimulation in teenager with complex regional pain syndrome for lymes disease. Case report and review of the literature]".ti. (1)

444	"[diagnosis, treatment and prevention of ixodes tick-borne borreliosis in the ul'ianovsk region]".ti. (1)

445	"[natural focal tick-borne transmissive infections in the children of perm territory: Differential diagnosis algorithm]".ti. (1)

446	"66-year-old patient with necrolytic migratory erythema, weight loss, anemia and diabetes mellitus. [german] 66-jahriger patient mit erythema necrolyticum migrans, gewichtsverlust, anamie und diabetes mellitus".ti. (0)

447	"Neuroborreliosis versus sclerosis multiplex. [czech]".ti. (0)

448	"[clinical and laboratory predictors for forecasting the outcomes of ixodes tick-borne borreliosis]".ti. (1)

449	"[seroprevalence of lyme borreliosis in workers from cordoba, colombia]".ti. (1)

450	"Prognosis of severity of tick-borne encephalitis based on selected parameters. [croatian] predvicrossed d signanje tezine krpeljnog meningoencefalitisa na temelju izabranih pokazatelja".ti. (0)

451	"[knowledge and prevention of tick-bite borreliosis: Survey of the population in alsace, an endemic area]".ti. (1)

452	"On an epidemiological study of lyme disease in hokkaido, japan".ti. (0)

453	"[risk of lyme disease development after a tick bite]".ti. (1)

454	"[borrelia infection with shoulder pain as the primary symptom]".ti. (1)

455	"[concentration of soluble forms of selectins in serum and in cerebrospinal fluid in group of patients with neuroborreliosis--a preliminary study]".ti. (1)

456	"Correlation between environmental and/or occupational risk and seropositivity for lyme disease. Analysis of data from liguria region. [italian] correlazione tra esposizione ambientale e/o professionale a rischio e sierologia nella borreliosi di lyme. Analisi del data base ligure".ti. (0)

457	"[detection of borrelia dna from patients with neuroborreliosis and erythema migrans]".ti. (1)

458	"Single test-tube nested polymerase chain reaction in the detection of borrelia burgdorferi in body fluids. [czech] dvoustupnova polymerazova retezova reakce (nested pcr) v jedne zkumavce na prukaz borrelia burgdorferi sensu lato v biologickych tekutinach".ti. (0)

459	"[prevalence of antiborrelia antibodies in two populations: Various interpretations of the same data]".ti. (1)

460	"[lyme disease with hepatitis and corticosteroids: A case report]".ti. (1)

461	"[long-term antibiotics are senseless]".ti. (1)

462	"[serological survey of the population in the territory endemic for ixodes tick-borne borreliosis]".ti. (1)

463	"[ecological and parasitological prerequisites for the spread of ixodes tick-borne borrelioses in the foci with various landscapes]".ti. (1)

464	"[causes affecting the incidence of ixodes tick-borne borreliosis]".ti. (1)

465	"[a late revealing of a case of tick-born borreliosis]".ti. (1)

466	"[clinical manifestations of neuroborreliosis in the volga region]".ti. (1)

467	"[cutaneous manifestations of the late stage of lyme's disease]".ti. (1)

468	"[a case of lyme neuroborreliosis coexistent with t-cell lymphoma]".ti. (1)

469	"Detection of anti-borrelia and anti-babesia antibodies in the serum of children with clinical manifestation and compatible epidemiology with lyme-like disease in the state of mato grosso do sul. [portuguese] resquisa de anticorpos anti-borrelia e anti-babesia em soro de criancas com manifestac-tioes cliinicas e epidemiologia compativeis com a doenc a de lyme-simile no estado de mato grosso do sul".ti. (0)

470	"[lyme borreliosis hepatitis]".ti. (1)

471	"[bilateral anterior uveiopapillitis, suspicious of lyme disease--case report]".ti. (1)

472	"[lyme disease--case report]".ti. and Nicula.ax. and "2008".yr. (1)

473	"Multiple erythema migrans-like lesions".ti. (0)

474	("[tick-bite in penile skin" and "A case report]").ti. (1)

475	"[all physicians must be capable to diagnose borrelia infection. A case report, or how to be a patient]".ti. (1)

476	"[ixodes ticks of the town of cherkessk and its environs]".ti. (1)

477	"[lyme disease]".ti. and Weis.ax. and Zeitschrift fur Gastroenterologie.jn. (1)

478	"Emergent zoonotic diseases in bosnia and herzegovina during 2007 to 2011 year".ti. (0)

479	"[two cases of rare unilateral sensorineural hearing loss]".ti. (1)

480	"[ticks and the pediatrician]".ti. (1)

481	"[coexisting of borreliosis, depression and psoriasis--case report]".ti. (1)

482	"[clinical manifestation of lyme borreliosis in children with positive and negatiwe western blot results]".ti. (1)

483	"[the significance of immunoblot tests in diagnosis of lyme borreliosis in children]".ti. (1)

484	"[unusual clinical manifestation of lyme disease--report of 2 cases]".ti. (1)

485	"[contribution of electronic health record in surveillance of notifiable diseases]".ti. (1)

486	"[improvement of federal epidemiologic surveillance, provision of biologic security of population of russian federation]".ti. (1)

487	"[no work disability in supposed post-borreliosis syndrome. On the decision of the olg saarbrucken of 19 may 2010]".ti. (1)

488	"Lyme borreliosis in children. [polish] borelioza z lyme u dzieci".ti. (0)

489	"[seroprevalence of lyme disease in navarra, spain]".ti. (1)

490	"[increase in childhood neuroborreliosis]".ti. (1)

491	"Hatay yöresinde kene kaynaklı lyme borelyozu seroprevalansı".ti. (0)

492	"[borrelia-associated lymphocytoma cutis]".ti. and Palmen.ax. (1)

493	"[concentrations of pro-inflammatory cytokines ifn-gamma, il-6, il-12 and il-15 in serum and cerebrospinal fluid in patients with neuroborreliosis undergoing antibiotic treatment]".ti. (1)

494	"[anti yersinia enterocolitica antybodies titer in serum in patients with chronic lyme arthritis]".ti. (1)

495	"Borrelia and rickettsia in skin bioptates of erythema migrans patients borelije i rikecije u bioptatima koze bolesnika s erythema migrans".ti. (0)

496	"[tick bites, lyme disease and early summer meningoencephalitis. Current indications concerning infection risks, prevention and therapy]".ti. (1)

497	"[lyme borreliosis in poland in 2009]".ti. (1)

498	"[lyme borreliosis in poland in 2010]".ti. (1)

499	"Van bölgesindeki riskli gruplarda borrelia burgdorferi seropozitifliği".ti. (0)

500	"[seropositivity of borrelia burgdorferi in risky groups in van region, turkey]".ti. (1)

501	"A woman in her 50s with manic psychosis".ti. (1)

502	"[atrioventricular heart block in lyme disease]".ti. (1)

503	"[epidemiological characteristics of lyme-like disease in children]".ti. (1)

504	"[recurrent tick-borne fever: A possible diagnosis in patients returning from senegal]".ti. (1)

505	"borrelia infection diagnostic challenge. Manage when suspected also without serologic findings".ti. (1)

506	"Is diagnosis of lyme borreliosis still difficult?".ti. (0)

507	"Pediatric facial palsy. [spanish] paralisis facial infantil".ti. (0)

508	"[epidemiologic analysis of infectious diseases identified as occupational diseases in poland in the years 1998-2002]".ti. (1)

509	"[slowly enlarging erythematous macule]".ti. and Canas.ax. (1)

510	"[facial palsy in a 7-year-old german tourist]".ti. (1)

511	"[results of etiologic diagnosis in clinical syndrome consistent with acute and chronic borreliosis]".ti. (1)

512	"[lyme borreliosis: From the erythema migrans to lyme arthritis]".ti. (1)

513	"Possibilities for prevention neuroinfections. [czech] moznosti prevence neuroinfekci".ti. (0)

514	"[the use of electron microscopy and virus isolation in the diagnosis of aseptic neuroinfections]".ti. (1)

515	"Diagnosis and treatment of neuroborreliosis: What is relevant and what is unnecessary".ti. (0)

516	"[infectious neurological diseases. Diagnostic confirmation from the cerebrospinal fluid]".ti. (1)

517	"[bell's palsy: Idiopathic or infectious?]".ti. (1)

518	"A contribution to the current in post-borrelia syndrome. [czech] prispevek k problematice postboreliove ho syndromu".ti. (0)

519	"[role of interleukin-18, interleukin-1beta and its soluble receptor (sil-1rii) in early and late lyme borreliosis]".ti. (1)

520	"[concentration of interleukin-18, interleukin-1beta, soluble receptor for interleukin-1 (sil-1rii) and c-reactive protein in patients with neuroborreliosis]".ti. (1)

521	"[the application of decision trees for constructing an algorithm for the differential diagnosis of zoonotic infections]".ti. (1)

522	"[the blood coagulation system and microcirculatory disorders in ixodid tick-borne borreliosis caused by borrelia miyamotoi]".ti. (1)

523	"[inflammatory borrelia - associated dilated cardiomyopathy]".ti. (1)

524	"[first report of three culture confirmed human lyme cases in turkey]".ti. (1)

525	"[choosing the threshold level in the detecting of antibodies to the causative agents of ixodes tick-borne borreliosis by lanthanide immonofluorescence assay]".ti. (1)

526	"[the clinical features of the acute period of mixed infection of tick-borne encephalitis and ixodes tick-borne borreliosis in children]".ti. (1)

527	"[epidemiologic situation by natural-foci infections in the crimea federal district in 2014-2015]".ti. (1)

528	"[isolated intracranial hypertension as the presenting sign of lyme disease]".ti. (1)

529	"[epidemiology and therapy of lyme arthritis and other manifestations of lyme borreliosis in germany: Results of a nation-wide survey]".ti. (1)

530	"Annual meeting and 1st symposium of the borreliosis society: Deplorable state of affairs in advanced lyme borreliosis. [german]".ti. (0)

531	"[clinical manifestations of 30 cases with lyme disease]".ti. (1)

532	"Establishment of a technique for rapid detection of the ospa outer membrane protein of borrelia burgdorferi via a gold-immuno-chromatographic assay".ti. (0)

533	"A serological study on tick-borne infectious diseases among residents in changbai mountainous area".ti. (0)

534	"[erythema migrans. Duration of antibiotic therapy?]".ti. (1)

535	"Recognition of the threats caused by infectious diseases in the netherlands in 2002 and 2003 by the weekly meetings of the early warning committee. [dutch]".ti. (0)

536	"[diagnosis of lyme borreliosis]".ti. and Rampini.ax. (1)

537	"[red swollen helix, facial erythema a week later. Borrelia lymphocytoma]".ti. (1)

538	"[humoral response to selected antigens of yersinia enterocolitica and yersinia pseudotuberculosis in the course of yersiniosis in humans. Iv. Specificity of antigens]".ti. (1)

539	"[winged scapula in lyme borreliosis]".ti. (1)

540	"Anti-bodies against lyme's disease (borreliosis) in persons occupationally exposed to ticks in bavaria - what are the risk factors?. [german]".ti. (0)

541	"[borreliosis as the cause of disability pensions in norway]".ti. (1)

542	"[lyme borreliosis and pregnancy]".ti. and Relic.ax. (1)

543	"Early clinical presentation following ticks' bite: Etiological diagnosis?. [french]".ti. (0)

544	"[acute hemiparesis revealing a neuroborreliosis in a child]".ti. (1)

545	"[physical factors in rehabilitation treatment of patients with ixodes tick-borne borreliosis with primary lesions of the joints]".ti. (1)

546	"[multiple ischemic strokes due to borrelia garinii meningovasculitis]".ti. (1)

547	"Lyme disease revealed by an atrio-ventricular block. [french, english]".ti. (0)

548	"[results of a prospective standardized study of 30 patients with chronic neurological and cognitive disorders after tick bites]".ti. (1)

549	"[lyme disease with cardiac involvement]".ti. and Rodionova.ax. (1)

550	"[lyme disease in cuba? Report of possible cases]".ti. (1)

551	"The chronic atrophic acrodermatitis".ti. and Romashkina.ax. (0)

552	"Lyme disease in the area of bjelovarsko-bilogorska county - negligible or neglected disease lymeska bolest na podrucju bjelovarsko-bilogorske zupanije - zanemariva ili zanemarena bolest".ti. (0)

553	"[growing boreliosis anxiety and its reasons]".ti. (1)

554	"[acrodermatitis chronica atrophicans can be difficult to diagnose]".ti. (1)

555	"[lyme carditis presenting as acute coronary syndrome: A case report]".ti. (1)

556	"[the organic disorders in the course of lyme disease]".ti. (1)

557	"[mental problems in lyme disease]".ti. (1)

558	"[clinical reasoning and decision-making in practice. An older man with prostate carcinoma and a painless paraparesis of the legs]".ti. (1)

559	"[peripheral facial palsy caused by lyme disease]".ti. (1)

560	"[cytokine cxcl13--a possible early csf marker for neuroborreliosis]".ti. (1)

561	"[why is my foot swollen and red?]".ti. (1)

562	"Lyme disease and juvenile idiopathic arthritis - a pediatric case report".ti. and Moreira.ax. and case report.pt. (0)

563	"About the central control of breathing: Becuase of a woman with obstructive sleep apnea, lyme disease and chronic consumption of opiods. [spanish] sobre el control central de la respiracion: A proposito de una mujer con apnea obstructiva del sueno, enfermedad de lyme y consumo cronico de opioides".ti. (0)

564	"Lyme disease in childhood: Data from an endemic area in sicily. [italian] la borreliosi di lyme in eta pediatrica: Individuata un'area endemica in sicilia".ti. (0)

565	"[acquired nystagmus in a 12-year-old boy as initial presentation of lyme disease]".ti. (1)

566	"[relapsing (recurrent) disease caused by borrelia miyamotoi]".ti. (1)

567	"[ocular lyme disease occurring during childhood: Five case reports]".ti. (1)

568	"Lyme disease".ti. and Nyman.ax. (1)

569	"Borrelial lymphocytoma. [dutch] borrelia-lymfocytoom".ti. (0)

570	"[tick-borne encephalitis in the north part of the canton de vaud]".ti. (1)

571	"[en route in switzerland - tick-borne and hantavirus infections]".ti. (1)

572	"Epidemiological data on lyme disease in alsace, limousin and rhône-alpes".ti. (0)

573	"Lyme borreliosis during pregnancy. [german] lymeborreliose wahrend der schwangerschaft".ti. (0)

574	"Differential diagnosis of hyperpigmentation. What's behind the blemishes on the skin".ti. (0)

575	"[evidence of borrelia burgdorferi spirochetes in patients with early disseminated lyme borreliosis]".ti. (1)

576	"[a curious case of livedo]".ti. (1)

577	"Red knee - inflammation without trauma - lyme disease".ti. (0)

578	"[isolated left abducens nerve palsy secondary to lyme disease in an 11-year-old boy]".ti. (1)

579	"Reversible dementias and treatable dementias".ti. (1)

580	"Children lyme borreliosis: Retrospective study of 16 cases. [french] borreliose de lyme chez l'enfant: Etude retrospective de 16 cas".ti. (0)

581	"[children lyme borreliosis: Retrospective study of 16 cases]".ti. (1)

582	"[serological verification of tick-borne encephalitis cases in the altai republic]".ti. (1)

583	"[epidemiological characterization of ixodes tick-borne borreliosis in the krasnoyarsk territory]".ti. (1)

584	"Analysis of 92 patients with tick-borne diseases in mudanjiang forestry central hospital".ti. (0)

585	"Tick-borne infection: Doubtful borreliosis test".ti. (0)

586	"[case of the month: A boy with a notable skin rash]".ti. (1)

587	"[arthritis and clinical history]".ti. (1)

588	"[ixodes tick-borne borreliosis with erythema nodosum]".ti. (1)

589	"[cardiovascular manifestations of lyme disease]".ti. and Sinski.ax. (1)

590	"Borrelia infection in swedish children - effective treatment and low risk of being affected by residual problems".ti. (0)

591	"[pcr sensitivity in detection of dna of borrelia burgdorferi sensu lato in different isolates]".ti. (1)

592	"[neuroboreliosis with motoric disturbations in the developmental age]".ti. (1)

593	"Signs and significance of a tick-bite: Psychiatric disorders associated with lyme disease".ti. (1)

594	"Cutaneous pseudolymphoma. An example of the relevance of minor surgery in pc. [spanish] seudolinfoma cutaneo. Un ejemplo de la pertinencia de la cirugia menor en atencion primaria".ti. (0)

595	"Epidemic of lyme disease research in jixian county of tianjin".ti. (0)

596	"Lyme disease: Which tests to prescribe to children?. [french] la maladie de lyme: Que prescrire comme examens complementaires chez l'enfant?".ti. (0)

597	"[recurrent ischemic strokes revealing lyme meningovascularitis]".ti. (1)

598	"[differential diagnosis of late-stage neuroborreliosis with affection of the central nervous system]".ti. (1)

599	"[differential aspects of multiple sclerosis and chronic borrelial encephalomyelitis]".ti. (1)

600	"[papilledema associated with visual field defects and motility disorders]".ti. (1)

601	"Nested-pcr real time as alternative molecular tool for detection of borrelia burgdorferi compared to the classical serological diagnosis of the blood".ti. (1)

602	("[psoriasis migrans" and "Erythema migrans as koebner phenomenon in psoriasis]").ti. (1)

603	"[lymphadenopathy and absences]".ti. (1)

604	"Lyme panniculitis. [dutch] lyme panniculitis".ti. (0)

605	"[epidemiology of tick-borne diseases in poland]".ti. (1)

606	"[lymphocytoma cutis benigna. Case report of a 9 year old boy]".ti. (1)

607	"[epidemiology of co-existing opisthorchiasis with ixodes tick-borne infections on tiumen region]".ti. (1)

608	"[even in fever, joint pain and erythema: Borrelia antibodies do not prove lyme disease]".ti. (1)

609	"[a non-itching lesion on the stomach]".ti. (1)

610	"Epidemiologic reports. [croatian] epidemioloska izvjesca".ti. (0)

611	"[the influence of child's behaviour on the course of neurological diagnosis]".ti. (1)

612	"[borrelia lymphocytoma in the breast]".ti. (1)

613	"[anetoderma with positive borrelia serology]".ti. (1)

614	"Erythema migrans and tetanus recall vaccination".ti. (0)

615	"Advances in the etiology of lichen sclerosus et atrophicus".ti. (0)

616	"[lyme disease in pediatrics: Different faces, the same disease. Cases series]".ti. (1)

617	"[the study of adaptation syndrome in mixed-infection of tick-borne encephalitis and borreliosis in children]".ti. (1)

618	"An investigation on lyme disease in xinjiang prospecting bureau of henan oilfield".ti. (0)

619	"[woman in her 70s with rapidly increasing cognitive impairment]".ti. (1)

620	"[anterior optic neuropathy in lyme disease, monosymptomatic form]".ti. (1)

621	"[occurrence of occupational diseases in poland, 2014]".ti. (1)

622	"Meningoradiculitis: A well-known but often missed expression of neuroborreliosis. [french]".ti. (0)

623	"A case of acute hepatitis due to lyme disease. [japanese]".ti. (0)

624	"[successful treatment of neuroborreliosis with combined administration of antibiotics and steroids: A case report]".ti. (1)

625	"[diagnostic difficulties in neuroborreliosis in children]".ti. (1)

626	"Clinical features of neurological manifestation of lyme disease in xinjing".ti. (0)

627	"Surveillance and analysis on the natural foci of lyme disease in southern mountainous area of urumqi".ti. (0)

628	"Molecular epidemiological study of borrelia burgdorferi infection among population in xinjiang".ti. (0)

629	"Seroprevalence of ehrlichiosis and lyme disease".ti. (0)

630	"Ocular involvement during rheumatic diseases in children".ti. (0)

631	"Neuroretinitis associated with interstitial keratitis: The first case report of lyme disease in the state of parana. [portuguese]".ti. (0)

632	"[suspected borreliosis: Therapy is incompatible with the french sun]".ti. (1)

633	"[seroprevalence of lyme borreliosis and tick-borne encephalitis in workers at risk, in eastern france]".ti. (1)

634	"[ataxic sensory neuropathy and lyme disease]".ti. (1)

635	"Cutaneous and systemic lyme disease: Pathogenesis - diagnosis - treatment. [german]".ti. (0)

636	"Serologic survey of lyme disease in mentougou district of beijing".ti. (0)

637	"An outbreak of lyme disease combined with scrub typhus".ti. (0)

638	"[significance of circulating immune complexes (cic) in the diagnosis of infections with borrelia burgdorferi]".ti. (1)

639	("monoinfections caused by borrelia burgdorferi and borrelia burgdorferi" and "anaplasma phagocytophilum co-infections in forestry workers and farmers").ti. (1)

640	"[risk of occupational infections caused by borrelia burgdorferi among forestry workers and farmers]".ti. (1)

641	"Seroprevalence of lyme borreliosis in forestry workers from lazio".ti. (0)

642	"Borreliosis. A proposito de un caso de enfermedad de lyme".ti. (0)

643	"Anosmia. [dutch] anosmie".ti. (0)

644	"[evaluation and prognosis of epidemiologic situation for lyme borreliosis in vladimir region]".ti. (1)

645	"[retrospective study of lyme borreliosis positive serologies in 2003]".ti. (1)

646	("Meningitis".ti. and Tsokos.ax. and "2001".yr.) not case report.pt. (0)

647	"[cellular borrelia tests]".ti. (1)

648	"Tick-borne zoonoses occurring in poland".ti. (0)

649	"[the lyme borreliosis controversy]".ti. (1)

650	"[false positive serological test in suspected borreliosis]".ti. (1)

651	"[diagnosis at a glance]".ti. and Ushiki.ax. (1)

652	"[neuroimaging in lyme disease]".ti. (1)

653	"[epidemiology of ixodes tick-borne borreliosis in the north of the volga-viatka region]".ti. (1)

654	"Seropositivity of lyme disease in erzurum province, turkey".ti. (0)

655	"[torticollis and seizures due to neuroborreliosis in a child]".ti. (1)

656	"Nueroborreliosis in belgian children. [dutch] neuroborreliose in een populatie belgische kinderen".ti. (0)

657	"Persistent symptoms after treatment of lyme disease: The lyme prospect study persisterende klachten na behandeling van lyme-borreliose: De lymeprospect-studie".ti. (0)

658	"[working together on occupational zoonoses]".ti. (1)

659	"Borrelial lymphocytome: About three cases. [french]".ti. (0)

660	"[central nervous system infection--tick-borne encephalitis, neuroborreliosis or both?]".ti. (1)

661	"Formation of multiplier educators for surveillance and zoonoses prevention in a medium city in brazil: From the analysis of health needs to the practical application of results".ti. (0)

662	"[the impact of climatic changes on ixodes (ixodidae) ticks and their related natural and focal infections]".ti. (1)

663	"[epidemiology of lyme disease in a healthcare area in north-west spain]".ti. (1)

664	"[lyme disease in paediatrics]".ti. and Diaz.ax. (1)

665	"[clinical polymorphism of neuroborreliosis at a late stage of the disease]".ti. (1)

666	"[total atrioventricular block following a tick bite]".ti. and Verbunt.ax. (1)

667	"Benign cutan lymphoid hyperplasia - atypical localisation: Case report".ti. (0)

668	"Ticks of public health importance in republic of croatia and protection measures".ti. (0)

669	("Tick bites and malaria" and "tekenbeten en malaria").ti. (0)

670	"[lyme-arthritis--a case report]".ti. and "von Ameln-Mayerhofer".ax. (1)

671	"Evaluation of the diagnostic significance of the lymphocyte proliferation test in patients with lyme borreliosis".ti. (0)

672	"New lyme disease pathogen discovered neuer borreliose-erreger entdeckt".ti. (0)

673	"[clinical manifestations in acute lyme diseases]".ti. (1)

674	"[peripheral facial palsy in children: Test for lyme borreliosis only in the presence of other clinical signs]".ti. (1)

675	"Epidemiology of occupational infectious diseases in health care workers. [polish] epidemiologia chorob zawodowych o etiologii zakaznej u pracownikow sluzby zdrowia regionu wroclawskiego".ti. (0)

676	"[acute atrioventricular block in chronic lyme disease]".ti. (1)

677	("prevalence of antibodies against anaplasma phagocytophilum" and "babesia microti i borrelia burgdorferi in adults in north-eastern poland").ti. (1)

678	"lyme disease--a reason for sudden sensorineural hearing loss and vestibular neuronitis".ti. (1)

679	"[bilateral facial nerve paralysis-diagnosis and treatment]".ti. (1)

680	"Seroepidemiological study of lyme disease in shandong province".ti. (0)

681	"[lyme disease complicated with human granulocytic anaplasmosis in a case]".ti. (1)

682	"Lyme borreliosis - the difficult search for an appropiate prophylaxis".ti. (0)

683	"[subacute proximal entrapment neuropathy of the radial nerve at the hiatus radialis]".ti. (1)

684	"[lyme disease could mimic dermatomyositis]".ti. (1)

685	"[lyme disease]".ti. and Weiss.ax. and Zeitschrift fur Gastroenterologie.jn. (0)

686	"[lyme disease]. [german] die lyme-borreliose".ti. (0)

687	"[tick bites and borreliosis in children attending forest kindergarten: A challenge for prevention]".ti. (1)

688	"[activity of lysosomal exoglycosidases in serum of patients with chronic borrelia arthritis]".ti. (1)

689	"Homeopathy in borreliosis: Plea for an unconventional therapeutic option. [german] homoopathie bei borreliose: Pladoyer fur eine unkonventionelle therapeutische option".ti. (0)

690	"[occupational diseases in poland]".ti. and Wilczynska.ax. and "2005".ti. (1)

691	"[what is your diagnosis? Erythema migrans (bull eye ). Initial stage of lyme borreliosis]".ti. (1)

692	"[exacerbation of chronic lower back pain with an unexpected twist]".ti. (1)

693	"[extrapulmonary complications of mycoplasma pneumoniae infections]".ti. and Sobanska.ax. (1)

694	"[guillain-barre syndrome in a course of early cutaneous type of lyme borreliosis: Diagnostic and therapeutic difficulties]".ti. (1)

695	"Borreliosis in humans in the region of warmia and mazury in 1999-2008".ti. (0)

696	"[acrodermatitis chronica atrophicans]".ti. and Wittmann.ax. (1)

697	"Lesions in tappers' hands in a hevea brasiliensis variety farm in the southwest of ivory-coast: Epidemiologic and forensic aspects".ti. (0)

698	"[presence of the antinuclear antibodies and antibodies to borrelia burgdorferi among patients with morphea en plaque, deep linear scleroderma and atrophoderma pasini-pierini]".ti. (1)

699	"Neuroborreliosis or schizophrenia coexisting with a neuroinfectious disease - case report".ti. (0)

700	"Progress in epidemiological research of lyme disease".ti. (0)

701	"Study of the natural foci of lyme disease in the city of meizhou".ti. (0)

702	"Investigation of lyme disease in changping district, beijing".ti. (0)

703	("Forty cases of human infestations with hard ticks" and "japan").ti. (0)

704	"Signal regulation on interspecies spread of tick-borne borrelia burgdorferii sensu lato".ti. (0)

705	"[spirochetal pericarditis]".ti. and Yano.ax. and "2007".yr. (1)

706	"Interpretation of the microbiologic tests used in the diagnosis of lyme borreliosis".ti. (0)

707	"Epidemiological investigation of lyme disease in parts of forest areas in qinghai province".ti. (0)

708	"[mesangioproliferative iga-nephritis in a patient with lyme borreliosis]".ti. (1)

709	"[clinical forms of neuroborreliosis--the analysis of patients diagnosed in department of infectious diseases and neuroinfection medical academy in bialystok between 2000-2005]".ti. (1)

710	"[comparison of two types of diagnostic test detecting antibodies against borrelia burgdorferi: Eia based on one genospecies antigens and elisa based on recombinant antigens]".ti. (1)

711	"[western-blot with vlse protein and in vivo antigens in lyme borreliosis diagnosis]".ti. (1)

712	"[encephalomeningitis caused by listeria monocytogenes in patient infected by tbe virus--case report]".ti. (1)

713	"[examination of intrathecal immunological response to borrelia burgdorferi antigens in patients with suspected neuroborreliosis--a preliminary communication]".ti. (1)

714	"Cases of cancer diagnosed at the department of infectious diseases and neuroinfections at the medical university of bialystok".ti. (0)

715	"[comparison of test with antigen vlse (c6) with tests with recombinant antigens in patients with lyme borreliosis]".ti. (1)

716	"[peripheral neuropathies in lyme borreliosis]".ti. (1)

717	"[presence of antibodies against gangliosides among patients with lyme borreliosis--preliminary study]".ti. (1)

718	"[effect of exposure to tick-bites on the course of lyme borreliosis in bialowieza residents]".ti. (1)

719	"Frequency of lyme arthritis in patients with unknown subacute arthritis".ti. (0)

720	"Neuroborreliosis at the universitiy hospital for infectious diseases".ti. (0)

721	"A piquant diagnosis. [french] un diagnostic piquant".ti. (0)

722	"Clinical analysis and diagnosis of 28 cases with neurological lyme disease".ti. (0)

723	"Nested pcr for testing serum samples of suspected lyme disease patients".ti. (0)

724	"[prediction of potential geographic distribution of lyme disease in qinghai province with maximum entropy model]".ti. (1)

725	"[infectious diseases in poland in 2001]".ti. (1)

726	"[infectious diseases in poland in 2002]".ti. (1)

727	"[enlarged lymph nodes, fever, abdominal cramps, numbness in arm and leg. Deciding evidence in the gluteal fold]".ti. (1)

728	"[prevalence of antibodies to anaplasma phagocytophilum and borrelia burgdorferi in forestry workers from the lublin region]".ti. (1)

729	"[lyme disease and general medicine, a survey of a sample of 133 instilled voluntary general practitioners in haute-savoie: A state of the art and a comparison with the recommendations of the 2006 consensus conference]".ti. (0)

730	"Advocate from vejen now leads denmark's first case of recognition of borrelia diagnoses from abroad. A woman was denied sickness benefits because her municipality would not recognize foreign medical records if she had borrelia".ti. (0)

731	"Usikkerhed om danske borrelia-diagnoser".ti. (0)

732	or/1-731 (448)

733	732 (448)

734	limit 733 to english language (4)

735	733 not 734 (444)

736	limit 733 to (english language or no language specified) (4)

737	733 not 736 (444)

738	limit 733 to (english language or no language specified or multilingual or undetermined) (4)

739	733 not 738 (444)

740	((afrikaans or albanian or arabic or armenian or azerbaijani or belorussian or bengali or bosnian or bulgarian or burmese or catalan or chinese or croatian or czech or danish or dutch or esperanto or estonian or finnish or flemish or french or gaelic, scots or georgian or german or greek or hausa or hebrew or hindi or hungarian or icelandic or indonesian or italian or japanese or kinyarwanda or kirghiz or korean or latin or latvian or lithuanian or macedonian or malay or malayalam or marathi or maori or masai or norwegian or persian or polish or portuguese or pushto or romanian or rumanian or russian or serbian or slovak or slovene or spanish or swahili or swedish or tagalog or tamil or telugu or thai or turkish or ukrainian or urdu or vietnamese or welsh) not english).lg. (4602920)

741	733 not 740 (4)

742	733 and 740 (444)

743	((afrikaans or albanian or arabic or armenian or azerbaijani or belorussian or bengali or bosnian or bulgarian or burmese or catalan or chinese or croatian or czech or danish or dutch or esperanto or estonian or finnish or flemish or french or gaelic, scots or georgian or german or greek or hausa or hebrew or hindi or hungarian or icelandic or indonesian or italian or japanese or kinyarwanda or kirghiz or korean or latin or latvian or lithuanian or macedonian or malay or malayalam or marathi or maori or masai or norwegian or persian or polish or portuguese or pushto or romanian or rumanian or russian or serbian or slovak or slovene or spanish or swahili or swedish or tagalog or tamil or telugu or thai or turkish or ukrainian or urdu or vietnamese or welsh) not (english or undetermined or multilingual)).lg. (4602878)

744	733 not 743 (4)

745	733 and 743 (444)

**Excludes: Embase** via Ovid http://ovidsp.ovid.com/

Date range searched: <1974 to 2024 January 29>

Date searched: 30 January 2024

1	"[duration of antibiotic therapy for lyme disease]".ti. and "Swedish Council".af. (0)

2	"[lyme-borreliosis: Diagnosis, symptoms and treatment]".ti. (1)

3	"Dossier: Lyme disease".ti. (0)

4	"16th consensus conference on anti-infectious therapy of the spilf. Lyme borreliosis: Diagnostic, therapeutic and preventive steps. Long text. Paris, france, 13 december 2006".ti. (0)

5	"[isolated intracranial hypertension due to lyme's disease]".ti. (1)

6	"[neuroborreliosis or borrelia hysteria. This case becomes a nightmare!]".ti. (1)

7	"[the patient is worried about tick bite. Is it usefull to test for borrelia?]".ti. (1)

8	"[trigeminal neuralgia, neuroborreliosis, and herpes: Finding the intruder]".ti. (0)

9	"A case of lyme disease with facial nerve palsy. [japanese]".ti. (0)

10	"[tick-borne diseases exposure of forestry workers and preventive methods usage]".ti. (1)

11	"[sequelae of tick-born encephalitis in cases treated in the infection observatory ward of regional hospital in gizycko]".ti. (1)

12	"[clinical thinking in neurological diagnosis (french experience)]".ti. (1)

13	"[evaluation of 10 cases of lyme disease presenting with erythema migrans in istanbul, turkey]".ti. (1)

14	"Lyme disease, first reported case in ecuador. [spanish] enfermedad de lyme. Primer caso reportado en ecuador".ti. (0)

15	"Guillain-barre syndrome after neuroborreliosis? Similar syndrome, different disease".ti. (1)

16	"Erratum: Diagnostic utility of serology to b. Burgdorferi in cartagena".ti. (1)

17	"Communicable diseases in croatia in 2007. [croatian] zarazne bolesti u hrvatskoj 2007".ti. (0)

18	"A fourteen-year-old boy with facial paresis caused by lyme disease. [dutch] een 14-jarige jongen met een facialisparese door de ziekte van lyme".ti. (0)

19	"An investigation of the prevalence of borrelia burgdorferi antibodies in north cyprus".ti. (0)

20	"[facial palsy: Diagnosis and management by primary care physicians]".ti. (1)

21	"[risk of human infection with ixodes tick-borne relapsing fever in different regional subzones of the perm region]".ti. (1)

22	"[use of indirect micromodification of enzyme immune (imei) reaction for mass screening serological studies for ixodes tick-borne borreliosis]".ti. (1)

23	"How to treat lyme borreliosis in a pregnant woman".ti. (1)

24	"[articular lesions in ixodes tick-borne borreliosis (lyme disease)]".ti. (1)

25	"[use of western-immunoblotting to determine serum igg-antibodies to antigens of different genotypes of borrelia--borrelia burgdorferi sensu stricto, borrelia afzelii, and borrelia garinii]".ti. (1)

26	"[detection of anti-borrelia antibodies by immunoblotting in lyme borreliosis]".ti. (1)

27	"[rapid development of dementia of an elderly person, diagnosis and successful treatment]".ti. (1)

28	"Creutzfeldt-jakob disease in clinical practice: A case report. [bosnian] creutzfeldt-jakobova bolest u klinickoj praksi: Prikaz bolesnice".ti. (0)

29	"Difficulty in analysis of serological tests results for spirochete infection in children".ti. (1)

30	"Skin manifestations of spirochete infection in chldren - own observations skorna manifestacja zakazenia kre{ogonek}tkiem borrelia burgdorferi u dzieci - doswiadczenia wlasne".ti. (0)

31	"[clinical course of borreliosis in children]".ti. (1)

32	"Analysis of the clinical course of infection caused by borrelia burgdorferi in children. [polish] analiza przebiegu klinicznego zakazenia kretkiem borrelia burgdorferi u dzieci".ti. (0)

33	"[specific features of borrelia burgdorferi infection in children]".ti. (1)

34	"Arthritis in the course of borrelia burgdorferi infection in children".ti. (1)

35	"[diagnostic problems with borrelia burgdorferi infection in children]".ti. (1)

36	"Non-specific symptoms of borrelia burgdorferi infection in children. [polish] nietypowe objawy zakazenia kretkiem borrelia burgdorferi u dzieci".ti. (0)

37	"[clinical characteristics of lyme disease in 12 cases]".ti. (0)

38	"News in pediatric practice 2001-2002: Infection. [italian] novita pediatria pratica 2001-2002: Infezione".ti. (0)

39	"References: Effective prevention of tick stings. [german] tipps: So beugen sie zeckenstichen wirksam vor".ti. (0)

40	"Advances in the treatment and prevention of lyme borreliosis. [german]".ti. (0)

41	"[ticks and borreliosis--an underestimated danger]".ti. (1)

42	"Lyme borreliose: Diagnostic, therapeutic and preventive approaches. Short text".ti. (1)

43	"The 16th consensus conference in anti-infectious therapeutics of the spilf (french language-speaking society of infectious pathology). Lyme borreliosis: Diagnostic, therapeutic and preventive steps. Long text - wednesday, december 13, 2006".ti. (1)

44	"Suspected borreliosis: What is wrong with the patient?".ti. (1)

45	"[the proof of antineural antibodies in patients with neuroborreliosis]".ti. (1)

46	"Rivm examines antibiotics against lyme: Possibly prophylactic treatment helps. [dutch] rivm onderzoekt antibiotica tegen lyme".ti. (0)

47	"Tug of war to new guideline on lyme disease: Patients associations fall off the cbo. [dutch] touwtrekken om nieuwe richtlijn lyme: Patientenverenigingen vallen cbo af".ti. (0)

48	"Difficult to tame schwer zu zahmen".ti. (0)

49	"[role of transcranial magnetic stimulation in clinical diagnosis: Facial nerve neurography]".ti. (1)

50	"[lyme's serology]".ti. (1)

51	"[perception du risque de maladie de lyme par les résidents d' une communauté infestée par des tiques étoilées américaines]".ti. (0)

52	"[lyme carditis]".ti. and Zimmermann.au. (0)

53	"[abdominal pain and wall distension as the onset form of neuroborreliosis]".ti. (1)

54	"Seronegative lyme disease: Report of two cases confirmed with pcr".ti. (0)

55	"[diplopia as the first manifestation of lyme disease]".ti. (1)

56	"[seroprevalence of borrelia burgdorferi and tick-borne encephalitis virus in a rural area of samsun, turkey]".ti. (1)

57	"Diagnose in beeld: A boy with nail abnormalities. [dutch] diagn ose in beeld: Een jongen met nagelafwijkingen".ti. (0)

58	"Accident ischemique transitoire revelant une neuroborreliose de lyme. [french] tia revealing lyme neuroborreliosis".ti. (0)

59	"Facial paralysis revealing a neuro-borreliosis. [french] paralysie faciale revelatrice d'une neuroborreliose".ti. (0)

60	"[doxycycline]".ti. and Aupee.au. (0)

61	"[complete atrioventricular block as the first clinical manifestation of a tick bite (lyme disease)]".ti. (1)

62	"[serology of lyme borreliosis and human granulocytic ehrlichiosis in 2005-2010]".ti. (1)

63	"[upper limb pain in an adolescent]".ti. (1)

64	"[lyme disease in patients with multiple sclerosis: Clinical, diagnostic and therapeutic features]".ti. (1)

65	"[clinical and instrumental characteristics of chronic neuroborreliosis]".ti. (2)

66	"Performance of four commercial immunoblots for serological confirmation of lyme borreliosis in rhone-alpes, france. [french] evaluation des performances de quatre immunoblots commercialises pour la confirmation de la borreliose de lyme dans une population de patients suivis en rhone-alpes".ti. (0)

67	"The role of ceftriaxone in modern antimicrobial therapy - a systematic review of recent clinical trials. [croatian] mjesto ceftriaksona u suvremenoj antimikrobnoj terapiji - sustavni pregled novijih klinickih ispitivanja".ti. (0)

68	"[the public health risk of ticks and tick-borne diseases in the south-eastern poland]".ti. (1)

69	"[neurologic problems in tick-borne diseases--clinical and diagnostic aspects]".ti. (1)

70	"[the information value of detection of borrelia antibodies in the healthy blood donors and in the population at risk]".ti. (1)

71	"Many faces of neuroborreliosis. Case report. [polish] rozne oblicza neuroboreliozy. Opis przypadku".ti. (0)

72	"[fever after a tick bite: Clinical manifestations and diagnosis of acute tick bite-associated infections in northeastern switzerland]".ti. (0)

73	"[optimization of serologic diagnosis of lime borreliosis]".ti. (1)

74	"[lyme borreliosis and demyelinating disease of the central nervous system]".ti. (1)

75	"Antibodies against the causative agents of some natural focal infections in blood donor sera from western slovakia. [czech] protilatky proti povodcom niektorych prirodne ohniskovych nakaz v serach darcov krvi zo zapadneho slovenska".ti. (0)

76	"[therapy-resistant depression with fatigue. A case of presumed streptococcal-associated autoimmune disorder]".ti. (1)

77	"Specifics of cytokine regulation in tick-borne encephalitis and lyme borreliosis".ti. (1)

78	"[5-yr-old with borrelial lymphocytoma]".ti. (1)

79	"[eosinophilic fasciitis associated with borrelia burgdorferi infection]".ti. (1)

80	"Ceffriaxone and pseudocholelithiasis in children. [croatian] ceftriaxon i pseudoholelitijaza u djece".ti. (0)

81	"Progressive facial hemiatrophy without changes in central nervous system and linear scleroderma en coup de sabre with extracerebral tumour - report of two cases. [polish] polowiczy zanik twarzy bez zmian w osrodkowym ukladzie nerwowym i twardzina en coup de sabre z guzem zewnatrzmozgowym - opis przypadkow".ti. (0)

82	"[case reports of borrelia infection in lakartidningen in 1909]".ti. (0)

83	"[pain and neuroborreliosis]".ti. and Berthele.au. (0)

84	"[two cases of proliferation of monoclonal and monotypic lymphocytes and plasma cells corresponding to acrodermatitis chronica atrophicans]".ti. (1)

85	"Surveillance system for assessing central nervous infections in lower saxony 1998-2000. [german] surveillance-system zur erfassung zentralnervoser infektionen in niedersachsen 1998-2000".ti. (0)

86	"Photoonycholysis caused by doxycycline. [czech] fotoonycholyza zpusobena doxycyklinem".ti. (0)

87	"[neopterin in serum and cerebrospinal fluid in lyme disease]".ti. (1)

88	"[the analysis of epidemiology, clinical symptoms, serological tests in the course of borreliosis]".ti. (1)

89	"[analysis of borrelia burgdorferi genostrains among patients with lyme disease]".ti. (1)

90	"[outbreak of lyme borreliosis with joint manifestation--case report]".ti. (1)

91	"Differential diagnosis of acrodermatitis chronica atrophicans with emphasis on chronic venous insufficiency. [german] differenzialdiagnose der acrodermatitis chronica atrophicans unter besonderer berucksichtigung der chronisch-venosen insuffizienz".ti. (0)

92	"Until a vaccine has been developed against lyme borreliosis, early antibiotic therapy is recommendable. [german] gegen lyme-borreliose bald auch impfen: Bis dahin: Moglichst fruh antibiotika!".ti. (0)

93	"[against lyme borreliosis vaccination also, soon. Until then: Antibiotics as early as possible!]".ti. (0)

94	"Real-time pcr-based in vitro susceptibility testing of borrelia burgdorferi against antimicrobial agents in the presence of eukariotic cells".ti. (0)

95	"[synovectomy of the knee joint]".ti. and Blahut.au. (0)

96	"[lyme disease acrodermitis chronica atrophicans: Misleading vascular signs]".ti. (1)

97	"Lyme neuroborreliosis: How to not underestimate neurological forms of lyme disease. [french] neuroborreliose de lyme: Comment ne pas meconnaitre une maladie de lyme neurologique".ti. (0)

98	"[acute myelitis and lyme disease]".ti. (1)

99	"[neurosyphilis and neuroborreliosis. Retrospective evaluation of 22 cases]".ti. (1)

100	"[lyme carditis]".ti. and Olszowska.au. (0)

101	"Multiannual fluctuations of some climate related diseases in poland and their possible relation to climate conditions".ti. (0)

102	"[a reprospective analysis of 973 patients with lyme borreliosis in kuyavian-pomeranian voivodship in 2000-2005]".ti. (1)

103	"[should a lumbar puncture be performed in any child with acute peripheral facial palsy and clinical suspicion of lyme borreliosis?]".ti. (1)

104	"The 46th annual meeting of the german society of occupational and environmental medicine (dgaum)".ti. (1)

105	"Faut-il traiter les symptômes post-borréliose de lyme par des antibiotiques au long cours?".ti. (0)

106	"[the combination of laryngeal paresis in the patients presenting with systemic tick-borne borreliosis (lyme disease)]".ti. (1)

107	"[ocular manifestations of lyme disease]".ti. and Bodaghi.au. (0)

108	"Rheumatoid arthritis and lyme disease: A case report".ti. (1)

109	"[lyme's disease presenting as increased gamma-gt activity]".ti. (1)

110	"Acrodermatitis chronica atrophicans. Proximal involvement of a lower limb. [french] acrodermatite chronique atrophiante atteinte proximale d'un membre inferieur".ti. (0)

111	"[a curious diving accident happened to a forest walk lover]".ti. (1)

112	"Problemen in de diagnose en behandeling van lyme-borreliose in nederland".ti. (0)

113	"[multiple erythema migrans and facial nerve paralysis: Clinical manifestations of early disseminated lyme borreliosis]".ti. (1)

114	"[inflammatory cardiac diseases by primary extracardial diseases]".ti. (1)

115	"Contagious and parasitic occupational diseases in the czech republic. [czech] nemoci z povolani prenosne a parazitarni v ceske republice".ti. (0)

116	"[occupational infectious diseases in the czech republic]".ti. (1)

117	"Lyme borreliosis of the musculoskeletal system".ti. (1)

118	"[multiple erythema]".ti. and Brunner.au. (0)

119	"[human granulocytic ehrlichiosis co-incident with lyme borreliosis in pregnant woman--a case study]".ti. (1)

120	"[an overlooked disease of tick bites: Lyme disease]".ti. (1)

121	"[features of cytokine levels in serum of patients with tick-borne borreliosis with different clinical signs]".ti. (1)

122	"Recurring focal neurologic events and cognitive impairment in a 52-years-old male".ti. (1)

123	"[lyme disease in childhood: An imported disease?]".ti. (1)

124	"[prevalence of antibodies to borrelia burgdorferi in sardinian teen-agers]".ti. (1)

125	"[lyme carditis--rare cause of dilated cardiomyopathy and rhythm disturbances]".ti. (1)

126	"Rheumatology and dermatology: Many interconnections, immense border areas. [italian] reumatologia e dermatologia: Molte le interconnessioni, vasti i grenzgebiete ".ti. (0)

127	"Acute polyradiculoneuropathy and lyme disease. [portuguese] polirradiculoneurite aguda e doenca de lyme-simile".ti. (0)

128	"Lyme disease. [turkish] lyme hastaligi".ti. (0)

129	"[back pain without radiculitis as an initial manifestation of lyme disease: Two cases]".ti. (1)

130	"[the new possibilities in serologic diagnostic of ixodes mite-borne borreliosis using immunochip]".ti. (1)

131	"[radiculalgia revealing lyme disease]".ti. (1)

132	"[clinical and laboratory characterization of ixodes tick-borne borreliosis in the baikal area]".ti. (1)

133	"Vaccine recommendations for infants and children travelers. [spanish] vacunacion del nino en viajes internacionales".ti. (0)

134	"[prevalence of borrelia burgdorferi antibodies in healthy population in poland]".ti. (1)

135	"Lyme borreliosis: Diagnostic, therapeutic and preventive steps".ti. (1)

136	"16th consensus conference in anti-infectious therapy. Lyme borreliosis: Preventive, therapeutic and diagnosis management. [french] 16<sup>e</sup> conference de consensus en therapeutique anti-infectieuse borreliose de lyme: Demarches diagnostiques, therapeutiques et preventives".ti. (0)

137	"Lyme borreliosis and co-infections. Place of anaplasma phagocytophilum and bartonella henselae".ti. (1)

138	"Epidemiological data on the spread of lyme disease in abruzzo. [italian] dati epidemiologici sulla diffusione in abruzzo della malattia di lyme in personale a rischio".ti. (0)

139	"Rapid diagnostic tests for infectious diseases. [turkish] enfeksiyon hastaliklarinda hizli tani testleri".ti. (0)

140	"[arthritis and infections]".ti. and Cimaz.au. (0)

141	"[incidence of tick-borne encephalitis virus and borrelia burgdorferi infections in farmers of the lublin province]".ti. (1)

142	"Lyme borreliosis in people in slovakia during 1999-2001".ti. (0)

143	"The incidence of some zoonoses in the slovak republic in 2009".ti. (0)

144	"[neuroborreliosis in patient with aplastic anaemia secondary to therapy with ticlopidine]".ti. (1)

145	"[borrelia-associated lymphocytoma cutis]".ti. and Corcostegui.au. (0)

146	"[neuroborreliosis after an early lyme disease treated by azithromycin]".ti. (1)

147	"Antibodies specific to borrelia burgdorferi in wild animal workers".ti. (0)

148	"Polyneuritis cranialis: Portuguese hospital case series. [portuguese] polineuropatia craniana - uma serie hospitalar".ti. (0)

149	"[a case of neuroborelliosis complicated by acute adrenal insufficiency]".ti. (1)

150	"A lyme patient about her disease".ti. (1)

151	"[neuroborreliosis]".ti. and Cuvelier.au. (0)

152	"[ultrasonography in diagnosing lyme arthritis of the knee joints in correlation with anti-ccp antibodies]".ti. (1)

153	"[usefulness of examinations of serum levels of matrix metalloproteinases 1, mmp-3, mmp-9, tissue inhibitor of metalloproteinases 1, hyaluronic acid and antibodies against cyclic citrullinated peptide in lyme arthritis, rheumatoid arthritis and patients with arthritic complaints]".ti. (1)

154	"[neuroborreliosis--a disease with many faces]".ti. (1)

155	"[clinical forms of neuroborreliosis among hospitalized patients in the years 2000-2005]".ti. (1)

156	"Clinical and evolutionary characteristics of four patients with pulmonary histoplasmosis reported in the paraiba paulista valley region. [portuguese] caracteristicas clinicas evolutivas de quatro pacientes com histoplasmose pulmonar encontrado na regiao do vale do paraiba".ti. (0)

157	"Lyme borreliosis - differential diagnostic difficulties in the interpretation of serological results. [croatian] lyme borelioza - diferencijalno dijagnosticke poteskoce u interpretaciji seroloskih rezultata".ti. (0)

158	"Difficulties in neuroborreliosis diagnostics".ti. (1)

159	"Lyme borreliosis as a cause of bilateral neuroretinitis with pronounced unilateral stellate maculopathy in a 8-year old girl. [slovak] lymeska borelioza jako pricina bilateralni neuroretinitidy s vyraznou jednostrannou hvezdicovitou makulopatii u osmilete divky".ti. (0)

160	"[lyme borreliosis in bosnia and herzegovina--clinical, laboratory and epidemiological research]".ti. (1)

161	"[borrelial erythema of the face]".ti. (1)

162	"Histopathological study of erythema chronicum migrans cases diagnosed in manaus. [portuguese, english] estudo histopatologico de casos de eritema cronico migratorio diagnosticados em manaus".ti. (0)

163	"Two specific manifestations of lyme disease. [dutch] twee bijzondere uitingsvormen van de ziekte van lyme".ti. (0)

164	"The significance of borrelia burgdorferi dna investigation in a diagnostic algorithm of lyme arthritis. [czech] vyznam vysetreni dna borrelia burgdorferi v diagnostickem algoritmu lymeske artritidy".ti. (0)

165	"[doubling of the number of cases of tick bites and lyme borreliosis seen by general practitioners in the netherlands]".ti. (1)

166	"[the lyme carditis as a rare differential diagnosis to an anterior myocardial infarction]".ti. (1)

167	"[sero-epidemiological studies of zoonotic infections in hunters--comparative analysis with veterinarians, farmers, and abattoir workers]".ti. (0)

168	"[seroepidemiological studies of zoonotic infections in hunters in southeastern austria--prevalences, risk factors, and preventive methods]".ti. (0)

169	"Lyme disease (lyme borreliosis)".ti. and Cabadaj.au. (0)

170	"Linear and localized scleroderma in plaques in a patient. [spanish] esclerodermia localizada en placas y lineal en un paciente".ti. (0)

171	"[erythema chronicum migrans. A case report]".ti. not conference abstract.pt. (2)

172	"Multiple gyrated erythemas in stage iii borreliosis".ti. (1)

173	"[tick-borne infections]".ti. and Dobler.au. (0)

174	"[epidemiology of borreliosis in workers of the district forestry offices in lower silesia]".ti. (1)

175	"A small turn then goes back.. [french] un petit tour et puis revient".ti. (0)

176	"[acute severe leukoencephalitis with posterior lesions due to borrelia burgdorferi infection]".ti. (1)

177	"[tick-borne borreliosis in the iaroslavl region]".ti. (1)

178	"[natural focal infections transmitted by ixodes ticks in the iaroslavl region: Epidemiological aspects]".ti. (1)

179	"Detection of serum antibodies against flagellin with a protein biochip in patients with lyme borreliosis".ti. (0)

180	"Serological epidemiology investigation of lyme disease in henan province".ti. (0)

181	"New knowledge about treatment of disseminated lyme borreliosis. [slovak] novsie poznatky o liecbe diseminovanej formy lymskej boreliozy".ti. (0)

182	"[borreliosis in children - clinical manifestation, diagnosis and treatment]".ti. (1)

183	"[seroprevalence of anaplasma phagocytophilum in patients with suspected lyme borreliosis]".ti. (1)

184	"[borreliosis--increasing clinical problem]".ti. (1)

185	"[prevalence of borreliosis among forestry workers in kujawsko-pomorskie voivodeship]".ti. (1)

186	"Zoonoses in the republic of croatia".ti. (1)

187	"Value in practice of the cerebrospinal fluid in treponema pallidum hemagglutination test for confirmation of neurosyphilis".ti. (0)

188	"[when is a diagnosis of borreliosis advisable?]".ti. (0)

189	"[complete heart block and lyme disease: Report of two different cases and literature review]".ti. (1)

190	"[diseases produced by borrelia]".ti. (1)

191	"[using the polymerase chain reaction to borrelia burgdorferi infection in localized scleroderma injure (morphea), in venezuelan patients]".ti. (1)

192	"[erythema migrans and lyme disease]".ti. and Fain.au. (0)

193	"The importance of serologic testing for lyme disease, in case of atrioventricular heart block. [french] interet des serologies de lyme devant un bloc auriculoventriculaire".ti. (0)

194	"[in fibromyalgia antibiotic therapy is not indicated]".ti. (0)

195	"[child fever in tropical area: Don't forget borreliosis]".ti. (1)

196	"Antibiotic therapy of head and neck infections. [german] antibiotikatherapie der infektionen an kopf und hals".ti. (0)

197	"Lyme's disease in persons occupationally exposed to ticks - an evaluation according to occupational medical and accident insurance legislation on the basis of a case report. [german] lyme-borreliose bei beruflich zecken-exponierten personen - eine arbeitsmedizinisch-unfallversicherungsrechtliche bewertung anhand einer kasuistik".ti. (0)

198	"[unilateral mydriasis in lyme neuroborreliosis]".ti. (2)

199	"[erythema migrans due to borrelia afzelii]".ti. (1)

200	"Neuroborreliosis in a patient with presumed cryptogenic ischaemic strokes. [german] neuroborreliose bei einer patientin mit vermeintlich kryptogenen zerebralen infarkten".ti. (0)

201	"[occurrence of erythema migrans in hajnowka district in years 2004-2009]".ti. (1)

202	"[combined manifestations of natural foci in the territory of the vologda province]".ti. (1)

203	"Lyme borreliosis in children. Epidemiology, diagnosis, clinical treatment, and therapy".ti. and "2006".yr. (1)

204	"Lyme borreliosis in infancy and adolescence".ti. (0)

205	"[tick removed. Sending on to examination?]".ti. (0)

206	"Diagnosis and therapy of borreliosis. [german] diagnose und therapie der borreliose".ti. (0)

207	"Is chronic fatigue syndrome in reality sometimes chronic lyme disease?. [dutch] is het chronischevermoeidheidssyndroom in werkelijkheid soms chronische lymeziekte?".ti. (0)

208	"Analysis of the clinical course of lyme borreliosis in respect to the initial disease manifestation in białowieża residents".ti. (0)

209	"[detection of dna of borrelia circulating in novosibirsk region]".ti. (1)

210	"[detection of borrelia dna in the borrelia burgdorferi sensu lato complex in the blood of patients with ixodes tick-borne borrelios]".ti. (0)

211	"[concentration of svap in serum of patients with different forms of lyme borreliosis]".ti. (1)

212	"[lyme carditis and symptomatic sinus node dysfunction]".ti. (1)

213	"Increased intracranial pressure due to neuroborreliosis. [german] pseudotumor cerebri bei neuroborreliose".ti. (0)

214	"A cutaneous manifestation of borreliosis. [french] une manifestation cutanee de la borreliose".ti. (0)

215	"Differential diagnosis of attention-deficit hyperactivity disorder in adolescents and adults - case report of an 18-year-old woman with neuroborreliosis. [german] differenzialdiagnostische abklarung der aufmerksamkeitsdefizit-/hyperaktivitatsstorung im jugendlichen- und erwachsenenalter - fallbeispiel einer 18 jahre alten patientin mit neuroborreliose".ti. (0)

216	"Assumed and real consequences of a tick bite: One willingly considers as borreliosis what one cannot explain. [german] vermeintliche und echte folgen eines zeckenstichs: Was man nicht erklaren kann, sieht man gern als borreliose an ".ti. (0)

217	"Mental problems or infection: Is there a chronic borreliosis despite therapy?. [german] psychische probleme oder infektion: Gibt es eine chronische borreliose trotz therapie?".ti. (0)

218	"[after the tick bite: Update borreliosis 2012]".ti. (1)

219	"Chronic borreliosis. Do combinations with hydroxychloroquine help?. [german] chronische borreliose. Helfen kombinationen mit hydroxychioroquin?".ti. (0)

220	"[bilateral tonic pupil as the only remaining ophthalmic sign of lyme disease: Case report]".ti. (1)

221	"[cutaneous marginal zone lymphoma (salt) and infection with borrelia burgdorferi]".ti. (1)

222	"[lyme borreliosis--diagnostic difficulties in interpreting serological results]".ti. (1)

223	"[early recurrence of neuroborreliosis with a fatal outcome]".ti. (1)

224	"[the swollen knee - differential diagnosis and therapy concepts from the pediatric rheumatologic perspective]".ti. (1)

225	"Clinical forms of tick bite. [spanish] picadura de garrapata: Formas clinicas".ti. (0)

226	"Ecopathological evidence and diagnosis of zoonotic diseases in the elderly".ti. (0)

227	"[pseudotumour cerebri in a patient with lyme disease and hypothyroidism]".ti. (1)

228	"Epidemiology of newly emerging and re-emerging zoonoses in bulgaria in the last decade".ti. (1)

229	"[diagnosis and treatment of lyme arthritis. Recommendations of the pharmacotherapy commission of the deutsche gesellschaft fur rheumatologie (german society for rheumatology)]".ti. (1)

230	"[acute transverse myelitis and lyme borreliosis: A case report]".ti. (1)

231	"Epidemiological investigation on lyme disease in changbai and tonghua county, jilin province".ti. (0)

232	"Anti-borrelia burgdorferi antibody response in 827 patients with suspected lyme disease".ti. (0)

233	"Borrelia burgdorferi antibody levels in 105 patients with suspected lyme disease".ti. (0)

234	"The first case of lyme disease in shanxi province, china: A case study".ti. (0)

235	"Tick vector-borne diseases: Human risk".ti. (0)

236	"[diseases and pathogenic agents transmitted by ticks in switzerland]".ti. (1)

237	"Neuroborreliose bij kinderen, herkenning en diagnostiek. [dutch] neuroborreliosis in children: Recognizing and diagnosing".ti. (0)

238	"[optic neuritis]".ti. and Wermund.au. (0)

239	"[alcohol and epilepsy: A case report between alcohol withdrawal seizures and neuroborreliosis]".ti. (1)

240	"Cooperation against work-related zoonoses".ti. (0)

241	"[a short journey through spirochetal infections]".ti. (1)

242	"[facial nerve paresis]".ti. and Glocker.au. (0)

243	"[ocular manifestations of lyme borreliosis in northwest croatia]".ti. (1)

244	"[diaphragmatic paralysis and arthromyalgia caused by lyme disease]".ti. (0)

245	"[interferon inductors in treatment of associative forms of tick-borne infection in children]".ti. (1)

246	"[seroepidemiologic study of lyme's borreliosis in mexico city and the northeast of the mexican republic]".ti. (0)

247	"Cranial polyneuritis in a case of neuroborreliosis with asymptomatic enhancement of cranial nerves. [german] polyneuritis cranialis: Neuroborreliose mit asymptomatischem hirnnervenenhancement".ti. (0)

248	"Facial palsies and neuroborreliosis in childhood. [german] fazialisparesen bei neuroborreliosen im kindesalter".ti. (0)

249	"Chronic lyme-disease as a chameleon of medicine: Burden of a special patient group".ti. (0)

250	"[idiopathic facial palsy]".ti. and Grosheva.au. (0)

251	"[re-infection with borrelia burgdorferi s.L in a patient with a history of neuroborreliosis--case report]".ti. (1)

252	"[failures of antibiotic treatment in lyme arthritis]".ti. (1)

253	"[delusional disorders in the course of tick-born encephalitis and borreliosis in patients with hemophilia a and posttraumatic epilepsy--diagnostic and therapeutic difficulties]".ti. (0)

254	"Seroepidemiological investigation of lyme disease in a part of forest region of xiao hinggan mountains".ti. (0)

255	"[reversible complete heart block by re-infection with borrelia burgdorferi with negative igm-antibodies]".ti. not erratum.pt. (1)

256	"[lyme borreliosis: Census of adult patients hospitalized in indre-et-loire (france), from the hospital discharge data (1999-2006)]".ti. (1)

257	"[seroprevalence of tick-borne encephalitis virus (tbev) among the residents of rural areas in sinop, central black-sea region, turkey]".ti. (1)

258	"Clinical characteristics of patients with intermediate uveitis".ti. (1)

259	"Cervical radiculitis as initial clinical feature of neuroborreliosis - case reports. [slovak] cervikalna radikulitida ako inicialny klinicky prejav neuroboreliozy - kazuistiky".ti. (0)

260	"[chronic fatigue syndrome following tick-borne diseases]".ti. (1)

261	"[lyme disease: Basis for treatment strategy, primary preventive care and secondary preventive care]".ti. (1)

262	"Epidemiological situation as to selected zoonoses in the slovak republic in 2007".ti. (0)

263	("Practice parameter: Treatment of nervous system lyme disease" and "report of the quality standards subcommittee of the american academy of neurology").ti. and Halperin.au. (0)

264	"[juvenile idiopathic arthritis and lyme disease: A case report]".ti. (1)

265	"[suspected borreliosis - what's behind it?]".ti. (1)

266	"[diagnosing lyme disease: The role of clinical and complementary examination]".ti. (1)

267	"[borrelia burgdorferi]".ti. and Hashimoto.au. and "2010".yr. (0)

268	"Stage-directed therapy of lyme borreliosis".ti. (1)

269	"[a case of suspected borreliosis]".ti. (1)

270	"Treatment after tick bites - problematics of borrelia infection".ti. and "2006".yr. and Tagliche Praxis.jn. (1)

271	"[purplish-erythematous plaque of the lower limb]".ti. (1)

272	"[polymorphic mental disorders in the course of lyme borreliosis--case study]".ti. (1)

273	"Lyme borreliosis during pregnancy: Consequences for the newborn?. [dutch] lyme-borreliose in de zwangerschap: Gevolgen voor de pasgeborene?".ti. (0)

274	"[borrelia burgdorferi myocarditis]".ti. (1)

275	"A case with an edge. [german] ein fall mit biss".ti. (0)

276	"Borrelia infections: Lyme disease and recurrent fever. [spanish] infecciones producidas por borrelias: Enfermedad de lyme y fiebre recurrente".ti. (0)

277	"[lyme borreliosis]".ti. and Herzer.au. and "2005".yr. (0)

278	"[45-year-old woman with joint pain and suspected lyme arthritis]".ti. (1)

279	"[lyme borreliosis]".ti. and Herzer.au. and "2014".yr. (0)

280	"Report from usa. [german] bericht aus usa".ti. (0)

281	"Clinical and therapeutic aspects of primary cutaneous marginal zone b-cell lymphoma. [dutch] klinische en therapeutische aspecten van het primair cutane marginale zone b-cel lymfoom".ti. (0)

282	"[early diagnosis of lyme borreliosis. Do not look only for erythema migrans]".ti. (0)

283	("[early diagnosis of lyme borreliosis]".ti. and Hofmann.au.) not "Do not look only for erythema migrans".ti. (0)

284	"[the variable spectrum of cutaneous lyme borreliosis. Diagnosis and therapy]".ti. (1)

285	"[early summer meningoencephalitis and lyme borreliosis. Relevant health risks in north bavaria]".ti. (0)

286	"[comatose state--think different]".ti. (1)

287	"Neuro-ehrlichiosis: A new tick-borne disease with nervous system involvement?".ti. (1)

288	"Analysis of imported lyme disease during beijing olympics".ti. (0)

289	"Human granulocytic anaplasmosis in bulgaria - confirmed cases and review of the literature".ti. (0)

290	"Prevalence of borrelia burgdorferi sensu lato species among patients in the czech republic; direct sequencing analysis and real-time polymerase chain reaction. [czech] prevalence druhu borrelia burgdorferi sensu lato u pacientu v ceske republice; prima sekvenc ni analyza a polymerazova rete zova reakce v realnem case".ti. (0)

291	"Erratum: Haufigste spate manifestation der lyme-borreliose (monatsschrift fur kinderheilkunde b 151:11 (1156-1162)). [german]".ti. (0)

292	"Lyme borreliosis (borrelia burgdorferi). [german] lyme-borreliose (borrelia burgdorferi)".ti. (0)

293	"One sided reddish-livid swelling of the earlobe - atheroma?".ti. (1)

294	"Rational diagnostic strategies for lyme borreliosis. [german] rationale diagnostik der lyme-borreliose".ti. (0)

295	("[lyme borreliosis in children and adolescents]".ti. and Huppertz.au.) not practice guideline.pt. (0)

296	"[lyme borreliosis]".ti. and Huppertz.au. and "2003".yr. (0)

297	"[clinical analysis of patients with peripheral facial palsy]".ti. (1)

298	"[a case of lyme disease requiring over 1 year to diagnose at an infectious-disease clinic]".ti. (1)

299	"Pediyatrik morfea (lokalize skleroderma) 14 olguya ait epidemiyolojik klinik ve laboratuvar bulguları".ti. (0)

300	"[inf-gamma and gm-csf concentrations in serum of patients with lyme disease]".ti. (0)

301	"[reddish-blue color lower leg with distinct vessel design. Venous insufficiency was lyme borreliosis]".ti. (0)

302	"[small risk of developing lyme borreliosis following a tick bite on ameland: Research in a general practice]".ti. (1)

303	"Nurse activities in accompanying children with lyme borreliosis".ti. (0)

304	"[synovitis score: Value of histopathological diagnostics in unclear arthritis. Case reports from rheumatological pathological practice]".ti. (1)

305	"[the role of pcr for the diagnosis of lyme arthritis]".ti. (0)

306	"Problems of false positive laboratory tests during qualification to the program of severe plaque psoriasis treatment on the basis of two cases. [polish] problemy falszywie dodatnich testow laboratoryjnych podczas kwalifikacji do programu leczenie ciezkiej postaci luszczycy plackowatej na podstawie dwoch przypadkow".ti. (0)

307	"[multilocular large erythemas and fever]".ti. (0)

308	"Unusual cutaneous borreliosis in alsace region. [french] la borreliose cutanee en alsace, dans tous ses etats".ti. (0)

309	"Bell palsy. Diagnostic and therapy. [german] idiopathische fazialisparese. Diagnostik und therapie".ti. (0)

310	"[cardiac arrest due to torsades de pointes ventricular tachycardia in a patient with lyme carditis.]".ti. (1)

311	"A multiplex fluorescent quantitative pcr method for detection of three rodent-carrying pathogens".ti. (0)

312	"Lyme disease in alsace: Incidence of a re-emerging disease. [french] incidence d'une maladie reemergente: La borreliose de lyme en alsace".ti. (0)

313	"The course of lyme disease in children - own observation. [polish] przebieg boreliozy u dzieci - obserwacje wlasne".ti. (0)

314	"[mental disorders in the course of lyme borreliosis and tick borne encephalitis]".ti. (1)

315	"[screening for antibodies against zoonotic agents among employees of the zoological garden of vienna, schonbrunn, austria]".ti. (0)

316	"Antibiotic therapy of lyme disease is criticized: Long-term therapy has no benefits, only side effects arzneimittel und therapie: Antibiose bei borreliose in der kritik: Langzeittherapie hat keinen nutzen, dafur nebenwirkungen".ti. (0)

317	"[a man with acutely starting pain in the right hip and thigh]".ti. (0)

318	"[complete atrio-ventricular block as a first symptom of borreliosis: Antibiotic treatment instead of pacemaker implantation]".ti. (1)

319	("Case reports" and "other association").ti. (1)

320	"[lyme disease from an ophthalmological point of view]".ti. (1)

321	"[psoriasis as koebner phenomenon in erythma migrans]".ti. (1)

322	"[clinical courses of acute and chronic neuroborreliosis following treatment with ceftriaxone]".ti. (1)

323	"[neuroborreliosis and diphasic meningoencephalitis -- common features and differences]".ti. (0)

324	"Cardiac arrest as atypical manifestation of early borreliosis - case report. [polish] nagle zatrzymanie krazenia jako nietypowy objaw wczesnej boreliozy - opis przypadku".ti. (0)

325	"Lyme borreliosis - not only endemic area disease - case report. [polish] borelioza - choroba nie tylko obszarow endemicznych - opis przypadku".ti. (0)

326	"Epidemiological aspects of the incidence of lyme borreliosis in košice region".ti. (0)

327	"Observation of the incidence of anti-anaplasma phagocytophilum antibodies in the risky groups of patients".ti. (0)

328	"Occurrence of zoonoses in the norwegian population".ti. (0)

329	"[usage of real time polymerase chain reaction for diagnostics of different tick-borne infections]".ti. (1)

330	"[isolation of the recombinant proteins ospc and the fragment flaa (f-flaa) from the western-siberian borrelia garinii nt29 isolates and the study of their immunochemical properties]".ti. (1)

331	"[lyme disease]".ti. and Kawabata.au. and "2003".yr. (0)

332	"A case of lyme disease with erythema migrans. [japanese]".ti. (0)

333	"[case of borrelia brainstem encephalitis presenting with severe dysphagia]".ti. (1)

334	"[evaluation of the usefulness cerebrospinal fluid myelin basic protein (mbp) concentration examination in patients with lyme neuroborreliosis--preliminary study]".ti. (1)

335	"[evaluation of cerebrospinal fluid serotonin (5-ht) concentration in patients with post-lyme disease syndrome--preliminary study]".ti. (1)

336	"[cranial neuritis with enhancement on post-contrast mri due to lyme disease]".ti. (1)

337	"[exposure to ticks and erythema chronicum migrans among borreliosis patients in lower silesia]".ti. (1)

338	"[eosinophilic fasciitis associated with borrelia afzelii infection]".ti. (1)

339	"Peripheral retinal vasculitis associated with lyme disease".ti. (1)

340	"A case of lyme disease with unusual cutaneous manifestations. [korean]".ti. (0)

341	"[acute urinary retention in a young woman with lyme borreliosis.]".ti. (1)

342	"A case of lyme disease. [japanese]".ti. (0)

343	"[preliminary studies on presence of antineuronal antibodies in serum of patients with lyme borreliosis]".ti. (1)

344	"[ambiguous swelling of the earlobe]".ti. (1)

345	"[prevalence of lyme disease among forestry workers]".ti. (1)

346	"[picture of the month: Erythema]".ti. and Kofoed.au. (0)

347	"Occupational infectious diseases in the west bohemian region in 1996-2000. [czech] profesionalni infekcni nemoci v zapadoceskem regionu v letech 1996-2000".ti. (0)

348	"[neuroimmune aspects of the pathogenesis and nitric oxide negative effects modifying the pathogenetic treatment of tick-borne infections]".ti. and "Kon'kova-Reidman".au. (0)

349	"[the polymerase chain reaction evaluation of borrelia burgdorferi dna presence in peripheral blood of patients with lyme disease]".ti. (1)

350	"[ehrlichia chaffeensis antibodies prevalence among patients bitten by ticks]".ti. (1)

351	"[effect of antibiotic therapy on levels of proinflammatory cytokines: Interleukin il-1, il-6 and tumor necrosis factor tnf-alpha in serum of patients with lyme borreliosis]".ti. (1)

352	"Fibrous nodules in lyme borreliosis. [czech] fibrozni uzly u lymeske boreliozy".ti. (0)

353	"[unclassified cardiomyopathy or lyme carditis? A three year follow-up]".ti. (1)

354	"[severe diarrhoea with invasive intestinal spirochaetosis]".ti. (1)

355	"Lyme disease in the ivory coast: A serological survey in abidjan. [french] borreliose de lyme en cote-d'ivoire: Mythe ou realite? Une enquete transversale de seropre valence a abidjan".ti. (0)

356	"[a japanese case of neuroborreliosis with papillitis]".ti. (1)

357	"Inflammatory dysfunctions of the facial nerve. [slovak] zapalove dysfunkcie tvaroveho nervu".ti. (0)

358	"Bullous localized scleroderma - presentation of two cases, therapeutic difficulties. [polish] twardzina ograniczona pecherzowa - przedstawienie dwoch przypadkow, trudnosci terapeutyczne".ti. (0)

359	"[neuroborreliosis in north jutland]".ti. (1)

360	"[early diagnosis of lyme arthritis]".ti. (1)

361	"[comparison of eia borrelia recombinant igm of the 3rd generation with eia test based on whole cell antigen in the diagnosis of lyme borreliosis]".ti. (0)

362	"[eye involvement of borrelia aetiology]".ti. (2)

363	"[multilocular erythema migrans in borreliosis]".ti. (1)

364	"[laboratory diagnosis of lyme borreliosis]".ti. and Kristiansen.au. (0)

365	"[the rate of detection and diagnostic significance of antibodies to borrelia burgdorferri in patients with eyes diseases of inflammatory nature]".ti. (1)

366	"[acrodermatitis chronica atrophicans: Late manifestation of lyme borreliosis]".ti. (0)

367	"[a man with a swollen nipple]".ti. (1)

368	"[risk for infection by lyme disease cause in green surfaces maintenance workers in belgrade]".ti. (1)

369	"Multiple foci of recurrent benign lymphoplasia in lyme disease".ti. (0)

370	"Early disseminated lyme borreliosis".ti. and Kryazheva.au. (0)

371	"[coexistence of two occupational infectious diseases in one patient--borreliosis and tick-borne encephalitis--a case report]".ti. (1)

372	"Comparison of results of measurements of visual evoked potentials in patients with multiple sclerosis and neuroborreliosis".ti. (1)

373	"Comprehensive biopsy-based diagnosis of inflammatory cardiomyopathy as a cause of new-onset heart failure of unclear etiology: Initial experience. [czech] komplexni biopticka diagnostika zane tlive kardiomyopatie jako priciny nove vznikleho srdecniho selhani nejasne etiologie: Prvni zkusenosti".ti. (0)

374	"Inflammatory cardiomyopathy secondary to borrelia infection, without a systemic serologic response".ti. (1)

375	"[clinical spectrum and incidence of neuro-borreliosis in the netherlands]".ti. (0)

376	"Early recurrence of a late lyme carditis: Successful treatment with roxithromycin and co-trimoxazole".ti. (1)

377	"Lyme disease: Current diagnosis and therapy".ti. (0)

378	"[comparative analysis of methods for laboratory diagnosis of lyme's disease in early stages of infection]".ti. (1)

379	"Role of borrelia burgdorferi in the development of morphea".ti. (0)

380	"Nipple disorders in children. [dutch] tepelafwijkingen bij kinderen".ti. (0)

381	"[the recurrence of lyme arthritis]".ti. (0)

382	"Dynamics of the polymerase chain reaction (pcr) and its value in lyme borreliosis. [czech] dynamika polymerazove retezove reakce (pcr) a jeji vyznam v diagnostice lymeske neuroboreliozy".ti. (0)

383	"[neuro-ophthamologic manifestations as unusual presentation of lyme disease]".ti. (1)

384	"Unusual finding in peripheral blood. [spanish] hallazgo infrecuente en sangre periferica".ti. (0)

385	"[lyme borreliosis--the most frequent vector-borne infection in denmark]".ti. (1)

386	"[lyme carditis--a bitter lesson or a delayed diagnostic success--a case report]".ti. (1)

387	"[borreliosis--simultaneous lyme carditis and psychiatric disorders--case report]".ti. (1)

388	"[lyme arthritis in children: A diagnostic trap]".ti. (1)

389	"[facial diplegia as the presenting feature of lyme disease]".ti. (1)

390	"[electromagnetic fields of extremely high frequency and radon baths in the treatment of lyme disease with nervous system lesions]".ti. (1)

391	"A case of selective paresis of the deep stabilization system as a result of borreliosis. [czech] pripad selektivni parezy hlubokeho stabilizacniho systemu jako nasledek boreliozy".ti. (0)

392	"Research advancement in action of cytokines in lyme disease. [chinese]".ti. (0)

393	"Study on tick-borne disease in northwest of china".ti. (0)

394	"Serological survey of lyme disease in some forest areas in shannxi province, china".ti. (0)

395	"Investigation of lyme disease natural epidemic focus in shaanxi province".ti. (0)

396	"Frequent incidence of lyme neuroborreliosis in children in the czech republic. [czech] casty vyskyt lymeske neuroboreliozy u deti v ceske republice".ti. (0)

397	"[the level of knowledge of general practitioners regarding the early phase of lyme borreliosis. Survey conducted among 106 general practitioners]".ti. (1)

398	"[ixodes tick-borne borreliosis and tick-borne encephalitis in the udmurt republic: Retrospective analysis of their spread]".ti. (1)

399	"[long-term morbidity and spatial distribution of tick-borne encephalitis and ixodes tick-borne borreliosis in udmurtia]".ti. (1)

400	"Serum epidemiological investigation on anaplasma phagocytophilum in population of the north forest area in fujian province".ti. (0)

401	"Surveillance and study of tick-borne diseases in zhejiang province, china".ti. (0)

402	"[invasion by trichinae in the patient hospitalized with suspicion of occupational borreliosis: A case report]".ti. (1)

403	"Anetoderma - a predictor of the occurrence of systemic autoimmunologic diseases. [polish] anetodermia - zwiastun wystapienia ukladowych shorzen autoimmunologicznych".ti. (0)

404	"Preliminary epidemiological investigation of lyme disease in hunan province".ti. (0)

405	"A seroepidemiological investigation of lyme disease in the shawan forest region of xinjiang, china".ti. (0)

406	"[study on the relation of transmission of lyme disease and ecological protection from western regions in china]".ti. (2)

407	"[ticks bite in foresters]".ti. (1)

408	"[neuroborreliosis in vest-agder]".ti. (1)

409	"Rare cutaneous signes of lyme disease in lorraine region".ti. (1)

410	"[differential diagnostics of neuroinfections: Lyme disease]".ti. (1)

411	"Cold, erythematous-violaceous macules on both breasts of a 66-year-old woman. [spanish] maculas eritemato-violaceas frias en ambas mamas de una paciente de 66 anos".ti. (0)

412	"Htlv-1 infection and alzheimer-type dementia. [spanish] infeccion por htlv-1 y demencia tipo alzheimer".ti. (0)

413	"[lyme disease. A tick bite, no panic! Emerging zoonoses in quebec]".ti. (1)

414	"[once again lyme disease]".ti. (1)

415	"Development of an indicator-based early warning system for lyme disease outbreaks".ti. (0)

416	"Neuroborreliosis. [croatian] neuroborelioza".ti. (0)

417	"[ usefulness of laboratory investigations for the diagnosis of acrodermatitis chronica atrophicans. ]".ti. (0)

418	"[onset of leber's hereditary optic neuropathy in association with borreliosis]".ti. (1)

419	"[atypical form of lyme disease]".ti. (1)

420	"[epidemiologic situation on dangerous infectious diseases on the territory of republic of abkhazia]".ti. (1)

421	"[lyme disease disclosed by pseudoneoplastic weight loss]".ti. (0)

422	"[preparations for serodiagnosis of diseases due to causative agents of ixode tick-borne borreliosis (lyme disease). Communication 1. Comparative study of the 1st generation enzyme immunoassay test-systems for detection of antibodies to borrelia burkdorferi sensu lato]".ti. (1)

423	"[preparations for serodiagnosis of diseases due to causative agents of ixodes tick-borne borreliosis (lyme disease). Communication 3. Comparative study of enzyme immunoassay test-systems for detection of igm to borrelia burgdorferi sensu lato]".ti. (1)

424	"[preparations for serodiagnosis of diseases due to causative agents of ixode tick-borne borreliosis (lyme disease). Communication 2. Comparative study of recombinant enzyme immunoassay test-systems (relisa) for serological diagnosis of ixode tick-borne borreliosis]".ti. (1)

425	"Acrodermatitis chronica atrophicans".ti. and Bottlaender.au. (0)

426	"First isolation of borrelia sp. (borrelia afzelii) from cerebrospinal fluid in a patient with neuroborreliosis in croatia. [croatian] prva izolacija borelije (borrelia afzelii) iz likvora bolesnika s neuroboreliozom u hrvatskoj".ti. (0)

427	"First isolation of borreliae garinii in croatia. [croatian] prva izolacija borreliae garinii u hrvatskoj".ti. (0)

428	"Azithromycin in the treatment of erythema migrans. [croatian] azitromycin u lijecenju erythema migrans".ti. (0)

429	"Fast onset cognitive impairment: Neuroborreliosis and positive 14-3-3 protein deterioro cognitivo de rapida instauracion: Neuroborreliosis y proteina 14-3-3 positiva".ti. (0)

430	"[red lower limb]".ti. (1)

431	"[psoriasiform exanthem in a patient with men-i]".ti. (1)

432	"[may lyme borreliosis lead to heart transplantation? - a case report]".ti. (1)

433	"[recurrent nerve palsy due to lyme disease: Report of two cases]".ti. (1)

434	"Risks of biotic agents".ti. (0)

435	"Characteristics of borrelia associated with lyme disease diagnosed in okinawa where lyme disease was not expected".ti. (0)

436	"[lyme uveitis: 2 case reports]".ti. (1)

437	"[red circles over the whole body]".ti. (0)

438	"[erythema chronicum migrans]".ti. and Maurin.au. (0)

439	"A case of a 14-year-old-girl with atrophoderma pasini-pierini. [polish] przypadek atrophodermii pasini-pierini u 14-letniej dziewczynki".ti. (0)

440	"[infectious diseases in poland in 2000]".ti. (1)

441	"[the onset of chronic lyme-borreliosis after the cure of tick-borne rickettsia infection in simultaneous infection]".ti. (1)

442	"[sensorineural loss of hearing in lower registers as the main symptom of lyme disease]".ti. (1)

443	"[spinal cord stimulation in teenager with complex regional pain syndrome for lymes disease. Case report and review of the literature]".ti. (0)

444	"[diagnosis, treatment and prevention of ixodes tick-borne borreliosis in the ul'ianovsk region]".ti. (1)

445	"[natural focal tick-borne transmissive infections in the children of perm territory: Differential diagnosis algorithm]".ti. (1)

446	"66-year-old patient with necrolytic migratory erythema, weight loss, anemia and diabetes mellitus. [german] 66-jahriger patient mit erythema necrolyticum migrans, gewichtsverlust, anamie und diabetes mellitus".ti. (0)

447	"Neuroborreliosis versus sclerosis multiplex. [czech]".ti. (0)

448	"[clinical and laboratory predictors for forecasting the outcomes of ixodes tick-borne borreliosis]".ti. (1)

449	"[seroprevalence of lyme borreliosis in workers from cordoba, colombia]".ti. (1)

450	"Prognosis of severity of tick-borne encephalitis based on selected parameters. [croatian] predvicrossed d signanje tezine krpeljnog meningoencefalitisa na temelju izabranih pokazatelja".ti. (0)

451	"[knowledge and prevention of tick-bite borreliosis: Survey of the population in alsace, an endemic area]".ti. (2)

452	"On an epidemiological study of lyme disease in hokkaido, japan".ti. (0)

453	"[risk of lyme disease development after a tick bite]".ti. (1)

454	"[borrelia infection with shoulder pain as the primary symptom]".ti. (1)

455	"[concentration of soluble forms of selectins in serum and in cerebrospinal fluid in group of patients with neuroborreliosis--a preliminary study]".ti. (1)

456	"Correlation between environmental and/or occupational risk and seropositivity for lyme disease. Analysis of data from liguria region. [italian] correlazione tra esposizione ambientale e/o professionale a rischio e sierologia nella borreliosi di lyme. Analisi del data base ligure".ti. (0)

457	"[detection of borrelia dna from patients with neuroborreliosis and erythema migrans]".ti. (0)

458	"Single test-tube nested polymerase chain reaction in the detection of borrelia burgdorferi in body fluids. [czech] dvoustupnova polymerazova retezova reakce (nested pcr) v jedne zkumavce na prukaz borrelia burgdorferi sensu lato v biologickych tekutinach".ti. (0)

459	"[prevalence of antiborrelia antibodies in two populations: Various interpretations of the same data]".ti. (0)

460	"[lyme disease with hepatitis and corticosteroids: A case report]".ti. (1)

461	"[long-term antibiotics are senseless]".ti. (1)

462	"[serological survey of the population in the territory endemic for ixodes tick-borne borreliosis]".ti. (1)

463	"[ecological and parasitological prerequisites for the spread of ixodes tick-borne borrelioses in the foci with various landscapes]".ti. (1)

464	"[causes affecting the incidence of ixodes tick-borne borreliosis]".ti. (1)

465	"[a late revealing of a case of tick-born borreliosis]".ti. (1)

466	"[clinical manifestations of neuroborreliosis in the volga region]".ti. (1)

467	"[cutaneous manifestations of the late stage of lyme's disease]".ti. (1)

468	"[a case of lyme neuroborreliosis coexistent with t-cell lymphoma]".ti. (1)

469	"Detection of anti-borrelia and anti-babesia antibodies in the serum of children with clinical manifestation and compatible epidemiology with lyme-like disease in the state of mato grosso do sul. [portuguese] resquisa de anticorpos anti-borrelia e anti-babesia em soro de criancas com manifestac-tioes cliinicas e epidemiologia compativeis com a doenc a de lyme-simile no estado de mato grosso do sul".ti. (0)

470	"[lyme borreliosis hepatitis]".ti. (0)

471	"[bilateral anterior uveiopapillitis, suspicious of lyme disease--case report]".ti. (1)

472	"[lyme disease--case report]".ti. and Nicula.au. and "2008".yr. (0)

473	"Multiple erythema migrans-like lesions".ti. (1)

474	("[tick-bite in penile skin" and "A case report]").ti. (1)

475	"[all physicians must be capable to diagnose borrelia infection. A case report, or how to be a patient]".ti. (1)

476	"[ixodes ticks of the town of cherkessk and its environs]".ti. (1)

477	"[lyme disease]".ti. and Weis.au. and Zeitschrift fur Gastroenterologie.jn. (0)

478	"Emergent zoonotic diseases in bosnia and herzegovina during 2007 to 2011 year".ti. (0)

479	"[two cases of rare unilateral sensorineural hearing loss]".ti. (1)

480	"[ticks and the pediatrician]".ti. (1)

481	"[coexisting of borreliosis, depression and psoriasis--case report]".ti. (1)

482	"[clinical manifestation of lyme borreliosis in children with positive and negatiwe western blot results]".ti. (1)

483	"[the significance of immunoblot tests in diagnosis of lyme borreliosis in children]".ti. (1)

484	"[unusual clinical manifestation of lyme disease--report of 2 cases]".ti. (1)

485	"[contribution of electronic health record in surveillance of notifiable diseases]".ti. (1)

486	"[improvement of federal epidemiologic surveillance, provision of biologic security of population of russian federation]".ti. (1)

487	"[no work disability in supposed post-borreliosis syndrome. On the decision of the olg saarbrucken of 19 may 2010]".ti. (1)

488	"Lyme borreliosis in children. [polish] borelioza z lyme u dzieci".ti. (0)

489	"[seroprevalence of lyme disease in navarra, spain]".ti. (1)

490	"[increase in childhood neuroborreliosis]".ti. (1)

491	"Hatay yöresinde kene kaynaklı lyme borelyozu seroprevalansı".ti. (0)

492	"[borrelia-associated lymphocytoma cutis]".ti. and Palmen.au. (0)

493	"[concentrations of pro-inflammatory cytokines ifn-gamma, il-6, il-12 and il-15 in serum and cerebrospinal fluid in patients with neuroborreliosis undergoing antibiotic treatment]".ti. (1)

494	"[anti yersinia enterocolitica antybodies titer in serum in patients with chronic lyme arthritis]".ti. (1)

495	"Borrelia and rickettsia in skin bioptates of erythema migrans patients borelije i rikecije u bioptatima koze bolesnika s erythema migrans".ti. (0)

496	"[tick bites, lyme disease and early summer meningoencephalitis. Current indications concerning infection risks, prevention and therapy]".ti. (1)

497	"[lyme borreliosis in poland in 2009]".ti. (1)

498	"[lyme borreliosis in poland in 2010]".ti. (1)

499	"Van bölgesindeki riskli gruplarda borrelia burgdorferi seropozitifliği".ti. (0)

500	"[seropositivity of borrelia burgdorferi in risky groups in van region, turkey]".ti. (0)

501	"A woman in her 50s with manic psychosis".ti. (1)

502	"[atrioventricular heart block in lyme disease]".ti. (1)

503	"[epidemiological characteristics of lyme-like disease in children]".ti. (1)

504	"[recurrent tick-borne fever: A possible diagnosis in patients returning from senegal]".ti. (1)

505	"borrelia infection diagnostic challenge. Manage when suspected also without serologic findings".ti. (0)

506	"Is diagnosis of lyme borreliosis still difficult?".ti. (1)

507	"Pediatric facial palsy. [spanish] paralisis facial infantil".ti. (0)

508	"[epidemiologic analysis of infectious diseases identified as occupational diseases in poland in the years 1998-2002]".ti. (0)

509	"[slowly enlarging erythematous macule]".ti. and Canas.au. (0)

510	"[facial palsy in a 7-year-old german tourist]".ti. (1)

511	"[results of etiologic diagnosis in clinical syndrome consistent with acute and chronic borreliosis]".ti. (1)

512	"[lyme borreliosis: From the erythema migrans to lyme arthritis]".ti. (1)

513	"Possibilities for prevention neuroinfections. [czech] moznosti prevence neuroinfekci".ti. (0)

514	"[the use of electron microscopy and virus isolation in the diagnosis of aseptic neuroinfections]".ti. (1)

515	"Diagnosis and treatment of neuroborreliosis: What is relevant and what is unnecessary".ti. (0)

516	"[infectious neurological diseases. Diagnostic confirmation from the cerebrospinal fluid]".ti. (1)

517	"[bell's palsy: Idiopathic or infectious?]".ti. (1)

518	"A contribution to the current in post-borrelia syndrome. [czech] prispevek k problematice postboreliove ho syndromu".ti. (0)

519	"[role of interleukin-18, interleukin-1beta and its soluble receptor (sil-1rii) in early and late lyme borreliosis]".ti. (1)

520	"[concentration of interleukin-18, interleukin-1beta, soluble receptor for interleukin-1 (sil-1rii) and c-reactive protein in patients with neuroborreliosis]".ti. (1)

521	"[the application of decision trees for constructing an algorithm for the differential diagnosis of zoonotic infections]".ti. (1)

522	"[the blood coagulation system and microcirculatory disorders in ixodid tick-borne borreliosis caused by borrelia miyamotoi]".ti. (1)

523	"[inflammatory borrelia - associated dilated cardiomyopathy]".ti. (1)

524	"[first report of three culture confirmed human lyme cases in turkey]".ti. (1)

525	"[choosing the threshold level in the detecting of antibodies to the causative agents of ixodes tick-borne borreliosis by lanthanide immonofluorescence assay]".ti. (1)

526	"[the clinical features of the acute period of mixed infection of tick-borne encephalitis and ixodes tick-borne borreliosis in children]".ti. (1)

527	"[epidemiologic situation by natural-foci infections in the crimea federal district in 2014-2015]".ti. (1)

528	"[isolated intracranial hypertension as the presenting sign of lyme disease]".ti. and article.pt. (1)

529	"[epidemiology and therapy of lyme arthritis and other manifestations of lyme borreliosis in germany: Results of a nation-wide survey]".ti. (1)

530	"Annual meeting and 1st symposium of the borreliosis society: Deplorable state of affairs in advanced lyme borreliosis. [german]".ti. (0)

531	"[clinical manifestations of 30 cases with lyme disease]".ti. (1)

532	"Establishment of a technique for rapid detection of the ospa outer membrane protein of borrelia burgdorferi via a gold-immuno-chromatographic assay".ti. (0)

533	"A serological study on tick-borne infectious diseases among residents in changbai mountainous area".ti. (0)

534	"[erythema migrans. Duration of antibiotic therapy?]".ti. (0)

535	"Recognition of the threats caused by infectious diseases in the netherlands in 2002 and 2003 by the weekly meetings of the early warning committee. [dutch]".ti. (0)

536	"[diagnosis of lyme borreliosis]".ti. and Rampini.au. (0)

537	"[red swollen helix, facial erythema a week later. Borrelia lymphocytoma]".ti. (2)

538	"[humoral response to selected antigens of yersinia enterocolitica and yersinia pseudotuberculosis in the course of yersiniosis in humans. Iv. Specificity of antigens]".ti. (1)

539	"[winged scapula in lyme borreliosis]".ti. (1)

540	"Anti-bodies against lyme's disease (borreliosis) in persons occupationally exposed to ticks in bavaria - what are the risk factors?. [german]".ti. (0)

541	"[borreliosis as the cause of disability pensions in norway]".ti. (1)

542	"[lyme borreliosis and pregnancy]".ti. and Relic.au. (0)

543	"Early clinical presentation following ticks' bite: Etiological diagnosis?. [french]".ti. (0)

544	"[acute hemiparesis revealing a neuroborreliosis in a child]".ti. (1)

545	"[physical factors in rehabilitation treatment of patients with ixodes tick-borne borreliosis with primary lesions of the joints]".ti. (1)

546	"[multiple ischemic strokes due to borrelia garinii meningovasculitis]".ti. (1)

547	"Lyme disease revealed by an atrio-ventricular block. [french, english]".ti. (0)

548	"[results of a prospective standardized study of 30 patients with chronic neurological and cognitive disorders after tick bites]".ti. (1)

549	"[lyme disease with cardiac involvement]".ti. and Rodionova.au. (0)

550	"[lyme disease in cuba? Report of possible cases]".ti. (1)

551	"The chronic atrophic acrodermatitis".ti. and Romashkina.au. (0)

552	"Lyme disease in the area of bjelovarsko-bilogorska county - negligible or neglected disease lymeska bolest na podrucju bjelovarsko-bilogorske zupanije - zanemariva ili zanemarena bolest".ti. (0)

553	"[growing boreliosis anxiety and its reasons]".ti. (1)

554	"[acrodermatitis chronica atrophicans can be difficult to diagnose]".ti. (1)

555	"[lyme carditis presenting as acute coronary syndrome: A case report]".ti. (1)

556	"[the organic disorders in the course of lyme disease]".ti. (1)

557	"[mental problems in lyme disease]".ti. (1)

558	"[clinical reasoning and decision-making in practice. An older man with prostate carcinoma and a painless paraparesis of the legs]".ti. (0)

559	"[peripheral facial palsy caused by lyme disease]".ti. (1)

560	"[cytokine cxcl13--a possible early csf marker for neuroborreliosis]".ti. (1)

561	"[why is my foot swollen and red?]".ti. (1)

562	"Lyme disease and juvenile idiopathic arthritis - a pediatric case report".ti. and Moreira.au. and case report.pt. (0)

563	"About the central control of breathing: Becuase of a woman with obstructive sleep apnea, lyme disease and chronic consumption of opiods. [spanish] sobre el control central de la respiracion: A proposito de una mujer con apnea obstructiva del sueno, enfermedad de lyme y consumo cronico de opioides".ti. (0)

564	"Lyme disease in childhood: Data from an endemic area in sicily. [italian] la borreliosi di lyme in eta pediatrica: Individuata un'area endemica in sicilia".ti. (0)

565	"[acquired nystagmus in a 12-year-old boy as initial presentation of lyme disease]".ti. (1)

566	"[relapsing (recurrent) disease caused by borrelia miyamotoi]".ti. (1)

567	"[ocular lyme disease occurring during childhood: Five case reports]".ti. (1)

568	"Lyme disease".ti. and Nyman.au. (0)

569	"Borrelial lymphocytoma. [dutch] borrelia-lymfocytoom".ti. (0)

570	"[tick-borne encephalitis in the north part of the canton de vaud]".ti. (1)

571	"[en route in switzerland - tick-borne and hantavirus infections]".ti. (1)

572	"Epidemiological data on lyme disease in alsace, limousin and rhône-alpes".ti. (0)

573	"Lyme borreliosis during pregnancy. [german] lymeborreliose wahrend der schwangerschaft".ti. (0)

574	"Differential diagnosis of hyperpigmentation. What's behind the blemishes on the skin".ti. (1)

575	"[evidence of borrelia burgdorferi spirochetes in patients with early disseminated lyme borreliosis]".ti. (0)

576	"[a curious case of livedo]".ti. (1)

577	"Red knee - inflammation without trauma - lyme disease".ti. (1)

578	"[isolated left abducens nerve palsy secondary to lyme disease in an 11-year-old boy]".ti. (1)

579	"Reversible dementias and treatable dementias".ti. (1)

580	"Children lyme borreliosis: Retrospective study of 16 cases. [french] borreliose de lyme chez l'enfant: Etude retrospective de 16 cas".ti. (0)

581	"[children lyme borreliosis: Retrospective study of 16 cases]".ti. (1)

582	"[serological verification of tick-borne encephalitis cases in the altai republic]".ti. (1)

583	"[epidemiological characterization of ixodes tick-borne borreliosis in the krasnoyarsk territory]".ti. (1)

584	"Analysis of 92 patients with tick-borne diseases in mudanjiang forestry central hospital".ti. (0)

585	"Tick-borne infection: Doubtful borreliosis test".ti. (1)

586	"[case of the month: A boy with a notable skin rash]".ti. (1)

587	"[arthritis and clinical history]".ti. (1)

588	"[ixodes tick-borne borreliosis with erythema nodosum]".ti. (1)

589	"[cardiovascular manifestations of lyme disease]".ti. and Sinski.au. (0)

590	"Borrelia infection in swedish children - effective treatment and low risk of being affected by residual problems".ti. (0)

591	"[pcr sensitivity in detection of dna of borrelia burgdorferi sensu lato in different isolates]".ti. (1)

592	"[neuroboreliosis with motoric disturbations in the developmental age]".ti. (1)

593	"Signs and significance of a tick-bite: Psychiatric disorders associated with lyme disease".ti. (1)

594	"Cutaneous pseudolymphoma. An example of the relevance of minor surgery in pc. [spanish] seudolinfoma cutaneo. Un ejemplo de la pertinencia de la cirugia menor en atencion primaria".ti. (0)

595	"Epidemic of lyme disease research in jixian county of tianjin".ti. (0)

596	"Lyme disease: Which tests to prescribe to children?. [french] la maladie de lyme: Que prescrire comme examens complementaires chez l'enfant?".ti. (0)

597	"[recurrent ischemic strokes revealing lyme meningovascularitis]".ti. (1)

598	"[differential diagnosis of late-stage neuroborreliosis with affection of the central nervous system]".ti. (2)

599	"[differential aspects of multiple sclerosis and chronic borrelial encephalomyelitis]".ti. (1)

600	"[papilledema associated with visual field defects and motility disorders]".ti. (0)

601	"Nested-pcr real time as alternative molecular tool for detection of borrelia burgdorferi compared to the classical serological diagnosis of the blood".ti. (1)

602	("[psoriasis migrans" and "Erythema migrans as koebner phenomenon in psoriasis]").ti. (1)

603	"[lymphadenopathy and absences]".ti. (0)

604	"Lyme panniculitis. [dutch] lyme panniculitis".ti. (0)

605	"[epidemiology of tick-borne diseases in poland]".ti. (1)

606	"[lymphocytoma cutis benigna. Case report of a 9 year old boy]".ti. (1)

607	"[epidemiology of co-existing opisthorchiasis with ixodes tick-borne infections on tiumen region]".ti. (1)

608	"[even in fever, joint pain and erythema: Borrelia antibodies do not prove lyme disease]".ti. (0)

609	"[a non-itching lesion on the stomach]".ti. (1)

610	"Epidemiologic reports. [croatian] epidemioloska izvjesca".ti. (0)

611	"[the influence of child's behaviour on the course of neurological diagnosis]".ti. (1)

612	"[borrelia lymphocytoma in the breast]".ti. (1)

613	"[anetoderma with positive borrelia serology]".ti. (1)

614	"Erythema migrans and tetanus recall vaccination".ti. (1)

615	"Advances in the etiology of lichen sclerosus et atrophicus".ti. (1)

616	"[lyme disease in pediatrics: Different faces, the same disease. Cases series]".ti. (1)

617	"[the study of adaptation syndrome in mixed-infection of tick-borne encephalitis and borreliosis in children]".ti. (1)

618	"An investigation on lyme disease in xinjiang prospecting bureau of henan oilfield".ti. (0)

619	"[woman in her 70s with rapidly increasing cognitive impairment]".ti. (1)

620	"[anterior optic neuropathy in lyme disease, monosymptomatic form]".ti. (1)

621	"[occurrence of occupational diseases in poland, 2014]".ti. (1)

622	"Meningoradiculitis: A well-known but often missed expression of neuroborreliosis. [french]".ti. (0)

623	"A case of acute hepatitis due to lyme disease. [japanese]".ti. (0)

624	"[successful treatment of neuroborreliosis with combined administration of antibiotics and steroids: A case report]".ti. (1)

625	"[diagnostic difficulties in neuroborreliosis in children]".ti. (1)

626	"Clinical features of neurological manifestation of lyme disease in xinjing".ti. (0)

627	"Surveillance and analysis on the natural foci of lyme disease in southern mountainous area of urumqi".ti. (0)

628	"Molecular epidemiological study of borrelia burgdorferi infection among population in xinjiang".ti. (0)

629	"Seroprevalence of ehrlichiosis and lyme disease".ti. (0)

630	"Ocular involvement during rheumatic diseases in children".ti. (1)

631	"Neuroretinitis associated with interstitial keratitis: The first case report of lyme disease in the state of parana. [portuguese]".ti. (0)

632	"[suspected borreliosis: Therapy is incompatible with the french sun]".ti. (0)

633	"[seroprevalence of lyme borreliosis and tick-borne encephalitis in workers at risk, in eastern france]".ti. (1)

634	"[ataxic sensory neuropathy and lyme disease]".ti. (1)

635	"Cutaneous and systemic lyme disease: Pathogenesis - diagnosis - treatment. [german]".ti. (0)

636	"Serologic survey of lyme disease in mentougou district of beijing".ti. (0)

637	"An outbreak of lyme disease combined with scrub typhus".ti. (0)

638	"[significance of circulating immune complexes (cic) in the diagnosis of infections with borrelia burgdorferi]".ti. (1)

639	("monoinfections caused by borrelia burgdorferi and borrelia burgdorferi" and "anaplasma phagocytophilum co-infections in forestry workers and farmers").ti. (1)

640	"[risk of occupational infections caused by borrelia burgdorferi among forestry workers and farmers]".ti. (1)

641	"Seroprevalence of lyme borreliosis in forestry workers from lazio".ti. (1)

642	"Borreliosis. A proposito de un caso de enfermedad de lyme".ti. (1)

643	"Anosmia. [dutch] anosmie".ti. (0)

644	"[evaluation and prognosis of epidemiologic situation for lyme borreliosis in vladimir region]".ti. (1)

645	"[retrospective study of lyme borreliosis positive serologies in 2003]".ti. (1)

646	"Meningitis".ti. and Tsokos.au. and "2001".yr. (0)

647	"[cellular borrelia tests]".ti. (0)

648	"Tick-borne zoonoses occurring in poland".ti. (1)

649	"[the lyme borreliosis controversy]".ti. (2)

650	"[false positive serological test in suspected borreliosis]".ti. (1)

651	"[diagnosis at a glance]".ti. and Ushiki.au. (0)

652	"[neuroimaging in lyme disease]".ti. (1)

653	"[epidemiology of ixodes tick-borne borreliosis in the north of the volga-viatka region]".ti. (1)

654	"Seropositivity of lyme disease in erzurum province, turkey".ti. (0)

655	"[torticollis and seizures due to neuroborreliosis in a child]".ti. and "2012".yr. (1)

656	"Nueroborreliosis in belgian children. [dutch] neuroborreliose in een populatie belgische kinderen".ti. (0)

657	"Persistent symptoms after treatment of lyme disease: The lyme prospect study persisterende klachten na behandeling van lyme-borreliose: De lymeprospect-studie".ti. (0)

658	"[working together on occupational zoonoses]".ti. (1)

659	"Borrelial lymphocytome: About three cases. [french]".ti. (0)

660	"[central nervous system infection--tick-borne encephalitis, neuroborreliosis or both?]".ti. (1)

661	"Formation of multiplier educators for surveillance and zoonoses prevention in a medium city in brazil: From the analysis of health needs to the practical application of results".ti. (0)

662	"[the impact of climatic changes on ixodes (ixodidae) ticks and their related natural and focal infections]".ti. (1)

663	"[epidemiology of lyme disease in a healthcare area in north-west spain]".ti. (1)

664	"[lyme disease in paediatrics]".ti. and Diaz.au. (0)

665	"[clinical polymorphism of neuroborreliosis at a late stage of the disease]".ti. (1)

666	"[total atrioventricular block following a tick bite]".ti. and Verbunt.au. (0)

667	"Benign cutan lymphoid hyperplasia - atypical localisation: Case report".ti. (0)

668	"Ticks of public health importance in republic of croatia and protection measures".ti. (0)

669	("Tick bites and malaria" and "tekenbeten en malaria").ti. (0)

670	"[lyme-arthritis--a case report]".ti. and "von Ameln-Mayerhofer".au. (0)

671	"Evaluation of the diagnostic significance of the lymphocyte proliferation test in patients with lyme borreliosis".ti. (1)

672	"New lyme disease pathogen discovered neuer borreliose-erreger entdeckt".ti. (0)

673	"[clinical manifestations in acute lyme diseases]".ti. (1)

674	"[peripheral facial palsy in children: Test for lyme borreliosis only in the presence of other clinical signs]".ti. (1)

675	"Epidemiology of occupational infectious diseases in health care workers. [polish] epidemiologia chorob zawodowych o etiologii zakaznej u pracownikow sluzby zdrowia regionu wroclawskiego".ti. (0)

676	"[acute atrioventricular block in chronic lyme disease]".ti. (1)

677	("prevalence of antibodies against anaplasma phagocytophilum" and "babesia microti i borrelia burgdorferi in adults in north-eastern poland").ti. (1)

678	"lyme disease--a reason for sudden sensorineural hearing loss and vestibular neuronitis".ti. (1)

679	"[bilateral facial nerve paralysis-diagnosis and treatment]".ti. (1)

680	"Seroepidemiological study of lyme disease in shandong province".ti. (0)

681	"[lyme disease complicated with human granulocytic anaplasmosis in a case]".ti. (1)

682	"Lyme borreliosis - the difficult search for an appropiate prophylaxis".ti. (1)

683	"[subacute proximal entrapment neuropathy of the radial nerve at the hiatus radialis]".ti. (1)

684	"[lyme disease could mimic dermatomyositis]".ti. (1)

685	"[lyme disease]".ti. and Weiss.au. and Zeitschrift fur Gastroenterologie.jn. (0)

686	"[lyme disease]. [german] die lyme-borreliose".ti. (0)

687	"[tick bites and borreliosis in children attending forest kindergarten: A challenge for prevention]".ti. (1)

688	"[activity of lysosomal exoglycosidases in serum of patients with chronic borrelia arthritis]".ti. (1)

689	"Homeopathy in borreliosis: Plea for an unconventional therapeutic option. [german] homoopathie bei borreliose: Pladoyer fur eine unkonventionelle therapeutische option".ti. (0)

690	"[occupational diseases in poland]".ti. and Wilczynska.au. and "2005".ti. (0)

691	"[what is your diagnosis? Erythema migrans (bull eye ). Initial stage of lyme borreliosis]".ti. (1)

692	"[exacerbation of chronic lower back pain with an unexpected twist]".ti. (1)

693	"[extrapulmonary complications of mycoplasma pneumoniae infections]".ti. and Sobanska.au. (0)

694	"[guillain-barre syndrome in a course of early cutaneous type of lyme borreliosis: Diagnostic and therapeutic difficulties]".ti. (1)

695	"Borreliosis in humans in the region of warmia and mazury in 1999-2008".ti. (0)

696	"[acrodermatitis chronica atrophicans]".ti. and Wittmann.au. (0)

697	"Lesions in tappers' hands in a hevea brasiliensis variety farm in the southwest of ivory-coast: Epidemiologic and forensic aspects".ti. (2)

698	"[presence of the antinuclear antibodies and antibodies to borrelia burgdorferi among patients with morphea en plaque, deep linear scleroderma and atrophoderma pasini-pierini]".ti. (1)

699	"Neuroborreliosis or schizophrenia coexisting with a neuroinfectious disease - case report".ti. (1)

700	"Progress in epidemiological research of lyme disease".ti. (0)

701	"Study of the natural foci of lyme disease in the city of meizhou".ti. (0)

702	"Investigation of lyme disease in changping district, beijing".ti. (0)

703	("Forty cases of human infestations with hard ticks" and "japan").ti. (0)

704	"Signal regulation on interspecies spread of tick-borne borrelia burgdorferii sensu lato".ti. (0)

705	"[spirochetal pericarditis]".ti. and Yano.au. and "2007".yr. (0)

706	"Interpretation of the microbiologic tests used in the diagnosis of lyme borreliosis".ti. (0)

707	"Epidemiological investigation of lyme disease in parts of forest areas in qinghai province".ti. (0)

708	"[mesangioproliferative iga-nephritis in a patient with lyme borreliosis]".ti. (1)

709	"[clinical forms of neuroborreliosis--the analysis of patients diagnosed in department of infectious diseases and neuroinfection medical academy in bialystok between 2000-2005]".ti. (1)

710	"[comparison of two types of diagnostic test detecting antibodies against borrelia burgdorferi: Eia based on one genospecies antigens and elisa based on recombinant antigens]".ti. (1)

711	"[western-blot with vlse protein and in vivo antigens in lyme borreliosis diagnosis]".ti. (1)

712	"[encephalomeningitis caused by listeria monocytogenes in patient infected by tbe virus--case report]".ti. (1)

713	"[examination of intrathecal immunological response to borrelia burgdorferi antigens in patients with suspected neuroborreliosis--a preliminary communication]".ti. (1)

714	"Cases of cancer diagnosed at the department of infectious diseases and neuroinfections at the medical university of bialystok".ti. (1)

715	"[comparison of test with antigen vlse (c6) with tests with recombinant antigens in patients with lyme borreliosis]".ti. (0)

716	"[peripheral neuropathies in lyme borreliosis]".ti. (1)

717	"[presence of antibodies against gangliosides among patients with lyme borreliosis--preliminary study]".ti. (1)

718	"[effect of exposure to tick-bites on the course of lyme borreliosis in bialowieza residents]".ti. (1)

719	"Frequency of lyme arthritis in patients with unknown subacute arthritis".ti. (0)

720	"Neuroborreliosis at the universitiy hospital for infectious diseases".ti. (1)

721	"A piquant diagnosis. [french] un diagnostic piquant".ti. (0)

722	"Clinical analysis and diagnosis of 28 cases with neurological lyme disease".ti. (0)

723	"Nested pcr for testing serum samples of suspected lyme disease patients".ti. (0)

724	"[prediction of potential geographic distribution of lyme disease in qinghai province with maximum entropy model]".ti. (1)

725	"[infectious diseases in poland in 2001]".ti. (1)

726	"[infectious diseases in poland in 2002]".ti. (1)

727	"[enlarged lymph nodes, fever, abdominal cramps, numbness in arm and leg. Deciding evidence in the gluteal fold]".ti. (0)

728	"[prevalence of antibodies to anaplasma phagocytophilum and borrelia burgdorferi in forestry workers from the lublin region]".ti. (1)

729	"[lyme disease and general medicine, a survey of a sample of 133 instilled voluntary general practitioners in haute-savoie: A state of the art and a comparison with the recommendations of the 2006 consensus conference]".ti. (0)

730	"Advocate from vejen now leads denmark's first case of recognition of borrelia diagnoses from abroad. A woman was denied sickness benefits because her municipality would not recognize foreign medical records if she had borrelia".ti. (0)

731	"Usikkerhed om danske borrelia-diagnoser".ti. (0)

732	or/1-731 (411)

733	remove duplicates from 732 (401)

734	733 (401)

735	limit 734 to english language (16)

736	734 not 735 (385)

737	limit 734 to (english language or no language specified) (16)

738	734 not 737 (385)

739	limit 734 to (english language or no language specified or polyglot) (16)

740	734 not 739 (385)

741	((afrikaans or albanian or arabic or armenian or azerbaidzhani or azerbaijani or basque or belorussian or bengali or bulgarian or burmese or bosnian or catalan or catalan; valencian or chinese or croatian or czech or danish or dutch or dutch; flemish or esperanto or estonian or finnish or french or gallegan or georgian or german or greek or "greek, modern (1453-)" or hebrew or hindi or hungarian or icelandic or indonesian or italian or japanese or korean or latvian or lithuanian or macedonian or malay or maori or mongolian or norwegian or persian or polish or portuguese or pushto or romanian or russian or serbian or sinhalese or slovak or slovene or slovenian or spanish or spanish; castilian or swedish or tagalog or thai or turkish or ukrainian or urdu or uzbek or vietnamese) not english).lg. (4689622)

742	734 not 741 (16)

743	734 and 741 (385)

744	((afrikaans or albanian or arabic or armenian or azerbaidzhani or azerbaijani or basque or belorussian or bengali or bulgarian or burmese or bosnian or catalan or catalan; valencian or chinese or croatian or czech or danish or dutch or dutch; flemish or esperanto or estonian or finnish or french or gallegan or georgian or german or greek or "greek, modern (1453-)" or hebrew or hindi or hungarian or icelandic or indonesian or italian or japanese or korean or latvian or lithuanian or macedonian or malay or maori or mongolian or norwegian or persian or polish or portuguese or pushto or romanian or russian or serbian or sinhalese or slovak or slovene or slovenian or spanish or spanish; castilian or swedish or tagalog or thai or turkish or ukrainian or urdu or uzbek or vietnamese) not (english or polyglot)).lg. (4689622)

745	734 not 744 (16)

### **Dataset 4:**

**Includes: MEDLINE® ALL** via Ovid http://ovidsp.ovid.com/

Date range searched: <1946 to March 20, 2024>

Date searched: 21/03/2024

1	"Randomized controlled trial of moderate cardiovascular exercise for patients with drug-resistant epilepsy".ti. (1)

2	"Telephonic review for outpatients with epilepsy - a prospective randomized, parallel group study".ti. (1)

3	"Effects of a remotely delivered group-format epilepsy self-management program on adverse health outcomes in vulnerable people with epilepsy: A causal mediation analysis".ti. (1)

4	"Effects of a broad-spectrum behavioral medicine treatment program on children with refractory epileptic seizures: An 8-year follow-up".ti. (1)

5	"Effects of a broad-spectrum behavior modification treatment program on children with refractory epileptic seizures".ti. (1)

6	"Effects of a contingent relaxation treatment program on adults with refractory epileptic seizures".ti. (1)

7	"Meditation improves clinicoelectroencephalographic measures in drug-resistant epileptics".ti. (1)

8	"Improving antiepileptic drug tolerability among patients living with epilepsy: The impact of pharmaceutical care services".ti. (1)

9	"Impact of educational actions on the quality of life of patients with epilepsy: A randomised controlled trial".ti. (1)

10	"Effects of an exercise program on health of people with epilepsy: A randomized clinical trial".ti. (1)

11	("Behavioral interventions as a treatment for epilepsy: A multicenter randomized controlled trial".ti. and Haut.ax.) not comment.pt. (1)

12	"Exercise training results in positive outcomes in persons with epilepsy".ti. (0)

13	"A structured, nurse-led intervention program improves quality of life in patients with epilepsy: A randomized, controlled trial".ti. (1)

14	"Effect of intensive self-management education on seizure frequency and quality of life in epilepsy patients with prodromes or precipitating factors".ti. (1)

15	"Wechat-based remote follow-up management reduces the burden of home care and anxiety on parents of children with refractory epilepsy: A randomized controlled study".ti. (1)

16	"The role of clinical pharmacist in pediatrics' adherence to antiepileptic drugs".ti. (1)

17	"A comparison of the quality of life of people with epilepsy receiving home-based and clinic-based epilepsy care using the european quality of life five-dimension three-level (eq-5d-3l) scale".ti. (1)

18	"Yoga in children with epilepsy: A randomized controlled trial".ti. (1)

19	"Impact of exercise as a complementary management strategy in people with epilepsy: A randomized controlled trial".ti. (1)

20	"Ethnicity and health outcomes among people with epilepsy participating in an epilepsy self-management rct".ti. (1)

21	"Enhancing medical compliance of patients with convulsive epilepsy in rural community: A randomized intervention trial".ti. (1)

22	"Systemic family therapy of comorbidity of anxiety and depression with epilepsy in adolescents".ti. (1)

23	"A randomized trial evaluating the effects of exercise on psychological outcomes in adult patients with epilepsy".ti. (0)

24	"Evaluation of acceptance and commitment therapy for drug refractory epilepsy: A randomized controlled trial in south africa--a pilot study".ti. (1)

25	"Acceptance and commitment therapy and yoga for drug-refractory epilepsy: A randomized controlled trial".ti. (1)

26	"Preventing depression in adolescents with epilepsy".ti. (1)

27	"The efficacy of an educational treatment program for patients with epilepsy moses: Results of a controlled, randomized study".ti. (1)

28	"A prospective evaluation of the effects of a 12-week outpatient exercise program on clinical and behavioral outcomes in patients with epilepsy".ti. (1)

29	"A prospective evaluation of the effects of an outpatient exercise program on seizure activity and antiepileptic drug concentrations in patients with epilepsy".ti. (0)

30	"A randomized trial of a group based cognitive behavior therapy program for older adults with epilepsy: The impact on seizure frequency, depression and psychosocial well-being".ti. (1)

31	"The effectiveness of mindfulness intervention based on stress reduction on quality of life and the number of seizure attacks in patients with epilepsy".ti. (0)

32	"Wordless intervention for people with epilepsy and learning disabilities wield: A randomised controlled feasibility trial".ti. (1)

33	"Effects of an epilepsy-specific internet intervention emyna on depression: Results of the encode randomized controlled trial".ti. (1)

34	"Effects of an epilepsy-specific digital treatment emyna on depression: Results of a randomized controlled trial".ti. (0)

35	"Effectiveness of specialist epilepsy nurses: Evaluation of a pilot project".ti. (0)

36	"Effectiveness of epilepsy specialist nurse - results of a randomized, controlled study in adult outpatients with epilepsy".ti. (0)

37	"Efficacy of the epilepsy nurse: Results of a randomized controlled study".ti. (1)

38	"Stress management intervention for living with epilepsy: Results of a rct".ti. (0)

39	"Controlled examination of effects of progressive relaxation training on seizure reduction".ti. (1)

40	"Self-management education for adults with poorly controlled epilepsy: The smile trial".ti. (0)

41	"Newly diagnosed epilepsy: Can nurse specialists help? A randomized controlled trial".ti. (1)

42	"The effect of a special nurse on patients' knowledge of epilepsy and their emotional state".ti. (1)

43	"Self-management education for adults with poorly controlled epilepsy smile uk: A randomised controlled trial".ti. and "2018".yr. (1)

44	"Feasibility and effects of nurse run clinics for patients with epilepsy in general practice: Randomised controlled trial".ti. (1)

45	"Self-management education for adults with poorly controlled epilepsy (smile-uk)".ti. and Epilepsia.jn. (0)

46	"one-year follow-up of a remotely delivered epilepsy self-management program in high-risk people with epilepsy".ti. (1)

47	"A 6-month prospective randomized controlled trial of remotely delivered group format epilepsy self-management versus waitlist control for high-risk people with epilepsy".ti. (1)

48	"Modulation of cardiac autonomic balance with adjuvant yoga therapy in patients with refractory epilepsy".ti. (1)

49	"Does an additional virtual interactive session increase the impact of digital educational material given to epilepsy patients? A randomized controlled trial".ti. and "2021".yr. (1)

50	"Optimising epilepsy management with a smartphone application: A randomised controlled trial".ti. (1)

51	"Effects of the medical resonance therapy music in the complex treatment of epileptic patients".ti. (1)

52	"A cluster-randomized trial comparing home-based primary health care and usual clinic care for epilepsy in a resource-limited country".ti. (1)

53	"Health literacy and education level correlates of participation and outcome in a remotely delivered epilepsy self-management program".ti. (1)

54	"Cognitive-behavior therapy with adult patients with epilepsy: A controlled outcome study".ti. (1)

55	"Mindfulness-based psychological treatment and social support intervention for drug-resistant epilepsy: A randomized, assessor-blinded, controlled trial".ti. (0)

56	"Mindfulness-based therapy for drug-resistant epilepsy: An assessor-blinded randomized trial".ti. (1)

57	"A pragmatic randomised controlled trial of a prompt and reminder card in the care of people with epilepsy".ti. (1)

58	"The impact of a depression self-management intervention on seizure activity".ti. (1)

59	"Multidisciplinary management improves anxiety, depression, medication adherence, and quality of life among patients with epilepsy in eastern china: A prospective study".ti. (1)

60	or/1-59 (48)

61	limit 60 to english language (48)

62	60 not 61 (0)

63	limit 60 to (english language or no language specified) (48)

64	60 not 63 (0)

65	limit 60 to (english language or no language specified or multilingual or undetermined) (48)

66	60 not 65 (0)

67	((afrikaans or albanian or arabic or armenian or azerbaijani or belorussian or bengali or bosnian or bulgarian or burmese or catalan or chinese or croatian or czech or danish or dutch or esperanto or estonian or finnish or flemish or french or gaelic, scots or georgian or german or greek or hausa or hebrew or hindi or hungarian or icelandic or indonesian or italian or japanese or kinyarwanda or kirghiz or korean or latin or latvian or lithuanian or macedonian or malay or malayalam or marathi or maori or masai or norwegian or persian or polish or portuguese or pushto or romanian or rumanian or russian or serbian or slovak or slovene or spanish or swahili or swedish or tagalog or tamil or telugu or thai or turkish or ukrainian or urdu or vietnamese or welsh) not english).lg. (4607430)

68	60 not 67 (48)

69	60 and 67 (0)

70	((afrikaans or albanian or arabic or armenian or azerbaijani or belorussian or bengali or bosnian or bulgarian or burmese or catalan or chinese or croatian or czech or danish or dutch or esperanto or estonian or finnish or flemish or french or gaelic, scots or georgian or german or greek or hausa or hebrew or hindi or hungarian or icelandic or indonesian or italian or japanese or kinyarwanda or kirghiz or korean or latin or latvian or lithuanian or macedonian or malay or malayalam or marathi or maori or masai or norwegian or persian or polish or portuguese or pushto or romanian or rumanian or russian or serbian or slovak or slovene or spanish or swahili or swedish or tagalog or tamil or telugu or thai or turkish or ukrainian or urdu or vietnamese or welsh) not (english or undetermined or multilingual)).lg. (4607388)

71	60 not 70 (48)

72	60 and 70 (0)

**Includes: Embase** via Ovid http://ovidsp.ovid.com/

Date range searched: <1974 to 2024 March 20>

Date searched: 21/03/2024

1	"Randomized controlled trial of moderate cardiovascular exercise for patients with drug-resistant epilepsy".ti. (1)

2	"Telephonic review for outpatients with epilepsy - a prospective randomized, parallel group study".ti. (1)

3	"Effects of a remotely delivered group-format epilepsy self-management program on adverse health outcomes in vulnerable people with epilepsy: A causal mediation analysis".ti. (1)

4	"Effects of a broad-spectrum behavioral medicine treatment program on children with refractory epileptic seizures: An 8-year follow-up".ti. (1)

5	"Effects of a broad-spectrum behavior modification treatment program on children with refractory epileptic seizures".ti. (1)

6	"Effects of a contingent relaxation treatment program on adults with refractory epileptic seizures".ti. (1)

7	"Meditation improves clinicoelectroencephalographic measures in drug-resistant epileptics".ti. (1)

8	"Improving antiepileptic drug tolerability among patients living with epilepsy: The impact of pharmaceutical care services".ti. (1)

9	"Impact of educational actions on the quality of life of patients with epilepsy: A randomised controlled trial".ti. (1)

10	"Effects of an exercise program on health of people with epilepsy: A randomized clinical trial".ti. (1)

11	"Behavioral interventions as a treatment for epilepsy: A multicenter randomized controlled trial".ti. and "journal article".pt. (0)

12	"Exercise training results in positive outcomes in persons with epilepsy".ti. (0)

13	"A structured, nurse-led intervention program improves quality of life in patients with epilepsy: A randomized, controlled trial".ti. (1)

14	"Effect of intensive self-management education on seizure frequency and quality of life in epilepsy patients with prodromes or precipitating factors".ti. (1)

15	"Wechat-based remote follow-up management reduces the burden of home care and anxiety on parents of children with refractory epilepsy: A randomized controlled study".ti. (1)

16	"The role of clinical pharmacist in pediatrics' adherence to antiepileptic drugs".ti. (1)

17	"A comparison of the quality of life of people with epilepsy receiving home-based and clinic-based epilepsy care using the european quality of life five-dimension three-level (eq-5d-3l) scale".ti. (0)

18	"Yoga in children with epilepsy: A randomized controlled trial".ti. (1)

19	"Impact of exercise as a complementary management strategy in people with epilepsy: A randomized controlled trial".ti. (1)

20	"Ethnicity and health outcomes among people with epilepsy participating in an epilepsy self-management rct".ti. (1)

21	"Enhancing medical compliance of patients with convulsive epilepsy in rural community: A randomized intervention trial".ti. (1)

22	"Systemic family therapy of comorbidity of anxiety and depression with epilepsy in adolescents".ti. (1)

23	"A randomized trial evaluating the effects of exercise on psychological outcomes in adult patients with epilepsy".ti. (0)

24	"Evaluation of acceptance and commitment therapy for drug refractory epilepsy: A randomized controlled trial in south africa--a pilot study".ti. (1)

25	"Acceptance and commitment therapy and yoga for drug-refractory epilepsy: A randomized controlled trial".ti. (1)

26	"Preventing depression in adolescents with epilepsy".ti. (1)

27	"The efficacy of an educational treatment program for patients with epilepsy moses: Results of a controlled, randomized study".ti. (1)

28	"A prospective evaluation of the effects of a 12-week outpatient exercise program on clinical and behavioral outcomes in patients with epilepsy".ti. (0)

29	"A prospective evaluation of the effects of an outpatient exercise program on seizure activity and antiepileptic drug concentrations in patients with epilepsy".ti. (0)

30	"A randomized trial of a group based cognitive behavior therapy program for older adults with epilepsy: The impact on seizure frequency, depression and psychosocial well-being".ti. (1)

31	"The effectiveness of mindfulness intervention based on stress reduction on quality of life and the number of seizure attacks in patients with epilepsy".ti. (0)

32	"Wordless intervention for people with epilepsy and learning disabilities wield: A randomised controlled feasibility trial".ti. (1)

33	"Effects of an epilepsy-specific internet intervention emyna on depression: Results of the encode randomized controlled trial".ti. (1)

34	"Effects of an epilepsy-specific digital treatment emyna on depression: Results of a randomized controlled trial".ti. (1)

35	"Effectiveness of specialist epilepsy nurses: Evaluation of a pilot project".ti. (1)

36	"Effectiveness of epilepsy specialist nurse - results of a randomized, controlled study in adult outpatients with epilepsy".ti. (0)

37	"Efficacy of the epilepsy nurse: Results of a randomized controlled study".ti. not "article in press".pt. (1)

38	"Stress management intervention for living with epilepsy: Results of a rct".ti. (1)

39	"Controlled examination of effects of progressive relaxation training on seizure reduction".ti. (1)

40	"Self-management education for adults with poorly controlled epilepsy: The smile trial".ti. (1)

41	"Newly diagnosed epilepsy: Can nurse specialists help? A randomized controlled trial".ti. (1)

42	"The effect of a special nurse on patients' knowledge of epilepsy and their emotional state".ti. (1)

43	"Self-management education for adults with poorly controlled epilepsy smile uk: A randomised controlled trial".ti. and "Health Technology Assessment".jn. (1)

44	"Feasibility and effects of nurse run clinics for patients with epilepsy in general practice: Randomised controlled trial".ti. (1)

45	"Self-management education for adults with poorly controlled epilepsy (smile-uk)".ti. and Epilepsia.jn. (1)

46	"one-year follow-up of a remotely delivered epilepsy self-management program in high-risk people with epilepsy".ti. (1)

47	"A 6-month prospective randomized controlled trial of remotely delivered group format epilepsy self-management versus waitlist control for high-risk people with epilepsy".ti. (1)

48	"Modulation of cardiac autonomic balance with adjuvant yoga therapy in patients with refractory epilepsy".ti. (1)

49	"Does an additional virtual interactive session increase the impact of digital educational material given to epilepsy patients? A randomized controlled trial".ti. not letter.pt. (1)

50	"Optimising epilepsy management with a smartphone application: A randomised controlled trial".ti. (1)

51	"Effects of the medical resonance therapy music in the complex treatment of epileptic patients".ti. (1)

52	"A cluster-randomized trial comparing home-based primary health care and usual clinic care for epilepsy in a resource-limited country".ti. (1)

53	"Health literacy and education level correlates of participation and outcome in a remotely delivered epilepsy self-management program".ti. (1)

54	"Cognitive-behavior therapy with adult patients with epilepsy: A controlled outcome study".ti. (1)

55	"Mindfulness-based psychological treatment and social support intervention for drug-resistant epilepsy: A randomized, assessor-blinded, controlled trial".ti. (1)

56	"Mindfulness-based therapy for drug-resistant epilepsy: An assessor-blinded randomized trial".ti. (0)

57	"A pragmatic randomised controlled trial of a prompt and reminder card in the care of people with epilepsy".ti. (1)

58	"The impact of a depression self-management intervention on seizure activity".ti. (1)

59	"Multidisciplinary management improves anxiety, depression, medication adherence, and quality of life among patients with epilepsy in eastern china: A prospective study".ti. (1)

60	or/1-59 (50)

61	limit 60 to english language (50)

62	60 not 61 (0)

63	limit 60 to (english language or no language specified) (50)

64	60 not 63 (0)

65	limit 60 to (english language or no language specified or polyglot) (50)

66	60 not 65 (0)

67	((afrikaans or albanian or arabic or armenian or azerbaidzhani or azerbaijani or basque or belorussian or bengali or bulgarian or burmese or bosnian or catalan or catalan; valencian or chinese or croatian or czech or danish or dutch or dutch; flemish or esperanto or estonian or finnish or french or gallegan or georgian or german or greek or "greek, modern (1453-)" or hebrew or hindi or hungarian or icelandic or indonesian or italian or japanese or korean or latvian or lithuanian or macedonian or malay or maori or mongolian or norwegian or persian or polish or portuguese or pushto or romanian or russian or serbian or sinhalese or slovak or slovene or slovenian or spanish or spanish; castilian or swedish or tagalog or thai or turkish or ukrainian or urdu or uzbek or vietnamese) not english).lg. (4697819)

68	60 not 67 (50)

69	60 and 67 (0)

70	((afrikaans or albanian or arabic or armenian or azerbaidzhani or azerbaijani or basque or belorussian or bengali or bulgarian or burmese or bosnian or catalan or catalan; valencian or chinese or croatian or czech or danish or dutch or dutch; flemish or esperanto or estonian or finnish or french or gallegan or georgian or german or greek or "greek, modern (1453-)" or hebrew or hindi or hungarian or icelandic or indonesian or italian or japanese or korean or latvian or lithuanian or macedonian or malay or maori or mongolian or norwegian or persian or polish or portuguese or pushto or romanian or russian or serbian or sinhalese or slovak or slovene or slovenian or spanish or spanish; castilian or swedish or tagalog or thai or turkish or ukrainian or urdu or uzbek or vietnamese) not (english or polyglot)).lg. (4697819)

71	60 not 70 (50)

72	60 and 70 (0)

**Excludes: MEDLINE® ALL** via Ovid http://ovidsp.ovid.com/

Date range searched: <1946 to March 20, 2024>

Date searched 21/03/2024

1	"[effectiveness of acceptance and commitment therapy on psychological well-being, quality of life and depression in patients with epilepsy]".ti. (0)

2	"[effectiveness of acceptance and commitment therapy on psychological well-being, quality of life and depression in patients with epilepsy]".ti. (0)

3	"[development and evaluation of the empowering a self-efficacy (ease) program for children with epilepsy]".ti. (1)

4	"Effectiveness of cognitive-behavioral group therapy on depression in intractable epileptic patients (with focusing on dysfunctional attitudes in epilepsy)".ti. (0)

5	"Effectiveness of cognitive-behavioral group therapy on depression in intractable epileptic patients (with focusing on dysfunctional attitudes in epilepsy)".ti. (0)

6	"Effectiveness of cognitive-behavioral therapy on dysfunctional attitudes in epileptic patients".ti. (0)

7	"Effectiveness of cognitive-- behavioral therapy on dysfunctional attitudes in epileptic patients".ti. (0)

8	or/1-7 (1)

9	limit 8 to english language (0)

10	8 not 9 (1)

11	limit 8 to (english language or no language specified) (0)

12	8 not 11 (1)

13	limit 8 to (english language or no language specified or multilingual or undetermined) (0)

14	8 not 13 (1)

15	((afrikaans or albanian or arabic or armenian or azerbaijani or belorussian or bengali or bosnian or bulgarian or burmese or catalan or chinese or croatian or czech or danish or dutch or esperanto or estonian or finnish or flemish or french or gaelic, scots or georgian or german or greek or hausa or hebrew or hindi or hungarian or icelandic or indonesian or italian or japanese or kinyarwanda or kirghiz or korean or latin or latvian or lithuanian or macedonian or malay or malayalam or marathi or maori or masai or norwegian or persian or polish or portuguese or pushto or romanian or rumanian or russian or serbian or slovak or slovene or spanish or swahili or swedish or tagalog or tamil or telugu or thai or turkish or ukrainian or urdu or vietnamese or welsh) not english).lg. (4607430)

16	8 not 15 (0)

17	8 and 15 (1)

18	((afrikaans or albanian or arabic or armenian or azerbaijani or belorussian or bengali or bosnian or bulgarian or burmese or catalan or chinese or croatian or czech or danish or dutch or esperanto or estonian or finnish or flemish or french or gaelic, scots or georgian or german or greek or hausa or hebrew or hindi or hungarian or icelandic or indonesian or italian or japanese or kinyarwanda or kirghiz or korean or latin or latvian or lithuanian or macedonian or malay or malayalam or marathi or maori or masai or norwegian or persian or polish or portuguese or pushto or romanian or rumanian or russian or serbian or slovak or slovene or spanish or swahili or swedish or tagalog or tamil or telugu or thai or turkish or ukrainian or urdu or vietnamese or welsh) not (english or undetermined or multilingual)).lg. (4607388)

19	8 not 18 (0)

20	8 and 18 (1)

**Excludes: Embase** via Ovid http://ovidsp.ovid.com/

Date range searched: <1974 to 2024 March 20>

Date searched: 21/03/2024

1	"[effectiveness of acceptance and commitment therapy on psychological well-being, quality of life and depression in patients with epilepsy]".ti. (0)

2	"[effectiveness of acceptance and commitment therapy on psychological well-being, quality of life and depression in patients with epilepsy]".ti. (0)

3	"[development and evaluation of the empowering a self-efficacy (ease) program for children with epilepsy]".ti. (1)

4	"Effectiveness of cognitive-behavioral group therapy on depression in intractable epileptic patients (with focusing on dysfunctional attitudes in epilepsy)".ti. (0)

5	"Effectiveness of cognitive-behavioral group therapy on depression in intractable epileptic patients (with focusing on dysfunctional attitudes in epilepsy)".ti. (0)

6	"Effectiveness of cognitive-behavioral therapy on dysfunctional attitudes in epileptic patients".ti. (0)

7	"Effectiveness of cognitive-- behavioral therapy on dysfunctional attitudes in epileptic patients".ti. (0)

8	or/1-7 (1)

9	limit 8 to english language (0)

10	8 not 9 (1)

11	limit 8 to (english language or no language specified) (0)

12	8 not 11 (1)

13	limit 8 to (english language or no language specified or polyglot) (0)

14	8 not 13 (1)

15	((afrikaans or albanian or arabic or armenian or azerbaidzhani or azerbaijani or basque or belorussian or bengali or bulgarian or burmese or bosnian or catalan or catalan; valencian or chinese or croatian or czech or danish or dutch or dutch; flemish or esperanto or estonian or finnish or french or gallegan or georgian or german or greek or "greek, modern (1453-)" or hebrew or hindi or hungarian or icelandic or indonesian or italian or japanese or korean or latvian or lithuanian or macedonian or malay or maori or mongolian or norwegian or persian or polish or portuguese or pushto or romanian or russian or serbian or sinhalese or slovak or slovene or slovenian or spanish or spanish; castilian or swedish or tagalog or thai or turkish or ukrainian or urdu or uzbek or vietnamese) not english).lg. (4697819)

16	8 not 15 (0)

17	8 and 15 (1)

18	((afrikaans or albanian or arabic or armenian or azerbaidzhani or azerbaijani or basque or belorussian or bengali or bulgarian or burmese or bosnian or catalan or catalan; valencian or chinese or croatian or czech or danish or dutch or dutch; flemish or esperanto or estonian or finnish or french or gallegan or georgian or german or greek or "greek, modern (1453-)" or hebrew or hindi or hungarian or icelandic or indonesian or italian or japanese or korean or latvian or lithuanian or macedonian or malay or maori or mongolian or norwegian or persian or polish or portuguese or pushto or romanian or russian or serbian or sinhalese or slovak or slovene or slovenian or spanish or spanish; castilian or swedish or tagalog or thai or turkish or ukrainian or urdu or uzbek or vietnamese) not (english or polyglot)).lg. (4697819)

19	8 not 18 (0)

20	8 and 18 (1)

### **Dataset 5:**

**Includes: MEDLINE® ALL** via Ovid http://ovidsp.ovid.com/

Date range searched: <1946 to March 21, 2024>

Date searched: 22/03/2024

1	"Effect of two physical exercise protocols on cognition and depressive symptoms in oldest‐old people: A randomized controlled trial".ti. (0)

2	"Effects of physical activity program on cognitive function and sleep quality in elderly with mild cognitive impairment: A randomized controlled trial".ti. (1)

3	"A multi-ingredient nutritional supplement in combination with resistance exercise and high-intensity interval training improves cognitive function and increases n-3 index in healthy older men: A randomized controlled trial".ti. and "2019".yr. (1)

4	"Effects of high-intensity combined training on women over 65".ti. (1)

5	"A multi-modal exercise intervention that improves cognitive function and physical performance, elderly with mobility-related disability: A randomized controlled trial".ti. (1)

6	"Moderate exercise improves cognitive function in healthy elderly people: Results of a randomized controlled trial".ti. (1)

7	"Effects of resistance exercise with instability on cognitive function rei study: A proof-of-concept randomized controlled trial in older adults with cognitive complaints".ti. (1)

8	"Improvements in cognition and associations with measures of aerobic fitness and muscular power following structured exercise".ti. (1)

9	"The effects of virtual kayak paddling exercise on postural balance, muscle performance, and cognitive function in older adults with mild cognitive impairment: A randomized controlled trial".ti. and "Randomized Controlled Trial".pt. (1)

10	"The effect of structured strength and balance training on cognitive function in frail, cognitive impaired elderly long-term care residents".ti. (1)

11	"Effects of physical activity intervention on physical and cognitive function in sedentary adults with and without diabetes".ti. (1)

12	"Impact of different aquatic exercise programs on body composition, functional fitness and cognitive function of non-institutionalized elderly adults: A randomized controlled trial".ti. (1)

13	"Effects of a 12-week exercise training program on physical function in institutionalized frail elderly".ti. (1)

14	"Long-term effects of individually tailored physical training and activity on physical function, well-being and cognition in scandinavian nursing home residents: A randomized controlled trial".ti. (1)

15	"Progressive resistance and balance training for falls prevention in long-term residential aged care: A cluster randomized trial of the sunbeam program".ti. (1)

16	"Effects of 12-week resistance exercise on electroencephalogram patterns and cognitive function in the elderly with mild cognitive impairment: A randomized controlled trial".ti. (1)

17	"Effects of physical, virtual reality-based, and brain exercise on physical, cognition, and preference in older persons: A randomized controlled trial".ti. (1)

18	"Effects of yoga therapy on postural stability in patients with schizophrenia-spectrum disorders: A single-blind randomized controlled trial".ti. (1)

19	"Effect of combined bifidobacteria supplementation and resistance training on cognitive function, body composition and bowel habits of healthy elderly subjects".ti. (1)

20	"Traditional thai exercise (ruesi dadton) for improving motor and cognitive functions in mild cognitive impairment: A randomized controlled trial".ti. (1)

21	"Effect of a dual-task net-step exercise on cognitive and gait function in older adults".ti. (1)

22	"Would older adults with mild cognitive impairment adhere to and benefit from a structured lifestyle activity intervention to enhance cognition?: A cluster randomized controlled trial".ti. (1)

23	"Benefits of physical exercise training on cognition and quality of life in frail older adults".ti. (1)

24	"Effect of exercise on cognition, conditioning, muscle endurance, and balance in older adults with mild cognitive impairment: A randomized controlled trial".ti. (1)

25	"Effect of physical activity on cognitive function in older adults at risk for alzheimer disease".ti. (1)

26	"Effects of functional tasks exercise on cognitive functions of older adults with mild cognitive impairment: A randomized controlled pilot trial".ti. (1)

27	"Effects of momentum-based dumbbell training on cognitive function in older adults with mild cognitive impairment: A pilot randomized controlled trial".ti. (1)

28	"The effects of different types of physical exercise on physical and cognitive function in frail institutionalized older adults with mild to moderate cognitive impairment. A randomized controlled trial".ti. (1)

29	"Physical activity and cognitive performance in the older population".ti. (0)

30	"Chronic endurance exercise training prevents aging‐related cognitive decline in healthy older adults: A randomized controlled trial".ti. (0)

31	"The effect of aerobic dance intervention on brain spontaneous activity in older adults with mild cognitive impairment: A resting-state functional mri study".ti. (1)

32	"Resistance training does not have an effect on cognition or related serum biomarkers in nonagenarians: A randomized controlled trial".ti. (1)

33	"Effects of multicomponent exercise on cognitive function in older adults with amnestic mild cognitive impairment: A randomized controlled trial".ti. (1)

34	"Mind-body exercise improves cognitive function and modulates the function and structure of the hippocampus and anterior cingulate cortex in patients with mild cognitive impairment".ti. (1)

35	"A multicomponent exercise intervention that reverses frailty and improves cognition, emotion, and social networking in the community-dwelling frail elderly: A randomized clinical trial".ti. (1)

36	"Comparison of time‐matched aerobic, resistance, or concurrent exercise training in older adults".ti. (0)

37	"Effects of two different intensities of aerobic exercise on elderly people with mild cognitive impairment: A randomized pilot study".ti. (1)

38	"Positive effects of physical training in activity of daily living–dependent older adults".ti. (0)

39	"Effects and mediating mechanisms of a structured limbs-exercise program on general cognitive function in older adults with mild cognitive impairment: A randomized controlled trial".ti. (1)

40	"Effect of handball training on cognitive ability in elderly with mild cognitive impairment".ti. (1)

41	"Changes in cognitive function in a randomized trial of physical activity: Results of the lifestyle interventions and independence for elders pilot study".ti. (1)

42	"Effect of elastic band‐based high‐speed power training on cognitive function, physical performance and muscle strength in older women with mild cognitive impairment".ti. (0)

43	"Effects of a specially designed aerobic dance routine on mild cognitive impairment".ti. (1)

44	"Effects of 5 years aerobic exercise on cognition in older adults: The generation 100 study: A randomized controlled trial".ti. (1)

45	or/1-44 (38)

46	limit 45 to english language (38)

47	45 not 46 (0)

48	limit 45 to (english language or no language specified) (38)

49	45 not 48 (0)

50	limit 45 to (english language or no language specified or multilingual or undetermined) (38)

51	45 not 50 (0)

52	((afrikaans or albanian or arabic or armenian or azerbaijani or belorussian or bengali or bosnian or bulgarian or burmese or catalan or chinese or croatian or czech or danish or dutch or esperanto or estonian or finnish or flemish or french or gaelic, scots or georgian or german or greek or hausa or hebrew or hindi or hungarian or icelandic or indonesian or italian or japanese or kinyarwanda or kirghiz or korean or latin or latvian or lithuanian or macedonian or malay or malayalam or marathi or maori or masai or norwegian or persian or polish or portuguese or pushto or romanian or rumanian or russian or serbian or slovak or slovene or spanish or swahili or swedish or tagalog or tamil or telugu or thai or turkish or ukrainian or urdu or vietnamese or welsh) not english).lg. (4607642)

53	45 not 52 (38)

54	45 and 52 (0)

55	((afrikaans or albanian or arabic or armenian or azerbaijani or belorussian or bengali or bosnian or bulgarian or burmese or catalan or chinese or croatian or czech or danish or dutch or esperanto or estonian or finnish or flemish or french or gaelic, scots or georgian or german or greek or hausa or hebrew or hindi or hungarian or icelandic or indonesian or italian or japanese or kinyarwanda or kirghiz or korean or latin or latvian or lithuanian or macedonian or malay or malayalam or marathi or maori or masai or norwegian or persian or polish or portuguese or pushto or romanian or rumanian or russian or serbian or slovak or slovene or spanish or swahili or swedish or tagalog or tamil or telugu or thai or turkish or ukrainian or urdu or vietnamese or welsh) not (english or undetermined or multilingual)).lg. (4607600)

56	45 not 55 (38)

57	45 and 55 (0)

**Includes: Embase** via Ovid http://ovidsp.ovid.com/

Date range searched: <1974 to 2024 March 21>

Date searched: 22/03/2024

1	"Effect of two physical exercise protocols on cognition and depressive symptoms in oldest‐old people: A randomized controlled trial".ti. (0)

2	"Effects of physical activity program on cognitive function and sleep quality in elderly with mild cognitive impairment: A randomized controlled trial".ti. (1)

3	"A multi-ingredient nutritional supplement in combination with resistance exercise and high-intensity interval training improves cognitive function and increases n-3 index in healthy older men: A randomized controlled trial".ti. and "2019".yr. (1)

4	"Effects of high-intensity combined training on women over 65".ti. (1)

5	"A multi-modal exercise intervention that improves cognitive function and physical performance, elderly with mobility-related disability: A randomized controlled trial".ti. (1)

6	"Moderate exercise improves cognitive function in healthy elderly people: Results of a randomized controlled trial".ti. (1)

7	"Effects of resistance exercise with instability on cognitive function rei study: A proof-of-concept randomized controlled trial in older adults with cognitive complaints".ti. (1)

8	"Improvements in cognition and associations with measures of aerobic fitness and muscular power following structured exercise".ti. (1)

9	"The effects of virtual kayak paddling exercise on postural balance, muscle performance, and cognitive function in older adults with mild cognitive impairment: A randomized controlled trial".ti. (1)

10	"The effect of structured strength and balance training on cognitive function in frail, cognitive impaired elderly long-term care residents".ti. (1)

11	"Effects of physical activity intervention on physical and cognitive function in sedentary adults with and without diabetes".ti. (1)

12	"Impact of different aquatic exercise programs on body composition, functional fitness and cognitive function of non-institutionalized elderly adults: A randomized controlled trial".ti. (1)

13	"Effects of a 12-week exercise training program on physical function in institutionalized frail elderly".ti. (1)

14	"Long-term effects of individually tailored physical training and activity on physical function, well-being and cognition in scandinavian nursing home residents: A randomized controlled trial".ti. (1)

15	"Progressive resistance and balance training for falls prevention in long-term residential aged care: A cluster randomized trial of the sunbeam program".ti. (1)

16	"Effects of 12-week resistance exercise on electroencephalogram patterns and cognitive function in the elderly with mild cognitive impairment: A randomized controlled trial".ti. (1)

17	"Effects of physical, virtual reality-based, and brain exercise on physical, cognition, and preference in older persons: A randomized controlled trial".ti. (0)

18	"Effects of yoga therapy on postural stability in patients with schizophrenia-spectrum disorders: A single-blind randomized controlled trial".ti. (1)

19	"Effect of combined bifidobacteria supplementation and resistance training on cognitive function, body composition and bowel habits of healthy elderly subjects".ti. (1)

20	"Traditional thai exercise (ruesi dadton) for improving motor and cognitive functions in mild cognitive impairment: A randomized controlled trial".ti. (0)

21	"Effect of a dual-task net-step exercise on cognitive and gait function in older adults".ti. (1)

22	"Would older adults with mild cognitive impairment adhere to and benefit from a structured lifestyle activity intervention to enhance cognition?: A cluster randomized controlled trial".ti. (1)

23	"Benefits of physical exercise training on cognition and quality of life in frail older adults".ti. (1)

24	"Effect of exercise on cognition, conditioning, muscle endurance, and balance in older adults with mild cognitive impairment: A randomized controlled trial".ti. (1)

25	"Effect of physical activity on cognitive function in older adults at risk for alzheimer disease".ti. and "2008".yr. (1)

26	"Effects of functional tasks exercise on cognitive functions of older adults with mild cognitive impairment: A randomized controlled pilot trial".ti. (1)

27	"Effects of momentum-based dumbbell training on cognitive function in older adults with mild cognitive impairment: A pilot randomized controlled trial".ti. (1)

28	"The effects of different types of physical exercise on physical and cognitive function in frail institutionalized older adults with mild to moderate cognitive impairment. A randomized controlled trial".ti. (1)

29	"Physical activity and cognitive performance in the older population".ti. (0)

30	"Chronic endurance exercise training prevents aging‐related cognitive decline in healthy older adults: A randomized controlled trial".ti. (0)

31	"The effect of aerobic dance intervention on brain spontaneous activity in older adults with mild cognitive impairment: A resting-state functional mri study".ti. (1)

32	"Resistance training does not have an effect on cognition or related serum biomarkers in nonagenarians: A randomized controlled trial".ti. (1)

33	"Effects of multicomponent exercise on cognitive function in older adults with amnestic mild cognitive impairment: A randomized controlled trial".ti. (1)

34	"Mind-body exercise improves cognitive function and modulates the function and structure of the hippocampus and anterior cingulate cortex in patients with mild cognitive impairment".ti. (1)

35	"A multicomponent exercise intervention that reverses frailty and improves cognition, emotion, and social networking in the community-dwelling frail elderly: A randomized clinical trial".ti. (1)

36	"Comparison of time‐matched aerobic, resistance, or concurrent exercise training in older adults".ti. (0)

37	"Effects of two different intensities of aerobic exercise on elderly people with mild cognitive impairment: A randomized pilot study".ti. (1)

38	"Positive effects of physical training in activity of daily living–dependent older adults".ti. (0)

39	"Effects and mediating mechanisms of a structured limbs-exercise program on general cognitive function in older adults with mild cognitive impairment: A randomized controlled trial".ti. (1)

40	"Effect of handball training on cognitive ability in elderly with mild cognitive impairment".ti. (1)

41	"Changes in cognitive function in a randomized trial of physical activity: Results of the lifestyle interventions and independence for elders pilot study".ti. (1)

42	"Effect of elastic band‐based high‐speed power training on cognitive function, physical performance and muscle strength in older women with mild cognitive impairment".ti. (0)

43	"Effects of a specially designed aerobic dance routine on mild cognitive impairment".ti. (1)

44	"Effects of 5 years aerobic exercise on cognition in older adults: The generation 100 study: A randomized controlled trial".ti. (1)

45	or/1-44 (36)

46	limit 45 to english language (36)

47	45 not 46 (0)

48	limit 45 to (english language or no language specified) (36)

49	45 not 48 (0)

50	limit 45 to (english language or no language specified or polyglot) (36)

51	45 not 50 (0)

52	((afrikaans or albanian or arabic or armenian or azerbaidzhani or azerbaijani or basque or belorussian or bengali or bulgarian or burmese or bosnian or catalan or catalan; valencian or chinese or croatian or czech or danish or dutch or dutch; flemish or esperanto or estonian or finnish or french or gallegan or georgian or german or greek or "greek, modern (1453-)" or hebrew or hindi or hungarian or icelandic or indonesian or italian or japanese or korean or latvian or lithuanian or macedonian or malay or maori or mongolian or norwegian or persian or polish or portuguese or pushto or romanian or russian or serbian or sinhalese or slovak or slovene or slovenian or spanish or spanish; castilian or swedish or tagalog or thai or turkish or ukrainian or urdu or uzbek or vietnamese) not english).lg. (4698032)

53	45 not 52 (36)

54	45 and 52 (0)

55	((afrikaans or albanian or arabic or armenian or azerbaidzhani or azerbaijani or basque or belorussian or bengali or bulgarian or burmese or bosnian or catalan or catalan; valencian or chinese or croatian or czech or danish or dutch or dutch; flemish or esperanto or estonian or finnish or french or gallegan or georgian or german or greek or "greek, modern (1453-)" or hebrew or hindi or hungarian or icelandic or indonesian or italian or japanese or korean or latvian or lithuanian or macedonian or malay or maori or mongolian or norwegian or persian or polish or portuguese or pushto or romanian or russian or serbian or sinhalese or slovak or slovene or slovenian or spanish or spanish; castilian or swedish or tagalog or thai or turkish or ukrainian or urdu or uzbek or vietnamese) not (english or polyglot)).lg. (4698032)

56	45 not 55 (36)

57	45 and 55 (0)

**Excludes: MEDLINE® ALL** via Ovid http://ovidsp.ovid.com/

Date range searched: <1946 to March 21, 2024>

Date searched: 22/03/2024

1	"Effects of exercise on the cognition of older women treated with lovastatin".ti. (1)

2	"Estudio piloto de metilfenidato y entrenamiento a padres en el tratamiento de niños con trastorno por déficit de atención-hiperactividad".ti. (0)

3	"The effective of facial exercises on the mental health in elderly adults".ti. (1)

4	"Effect of balance exercise on functional reach test and body sway for the frail elderly".ti. (1)

5	or/1-4 (3)

6	limit 5 to english language (1)

7	5 not 6 (2)

8	limit 5 to (english language or no language specified) (1)

9	5 not 8 (2)

10	limit 5 to (english language or no language specified or multilingual or undetermined) (1)

11	5 not 10 (2)

12	((afrikaans or albanian or arabic or armenian or azerbaijani or belorussian or bengali or bosnian or bulgarian or burmese or catalan or chinese or croatian or czech or danish or dutch or esperanto or estonian or finnish or flemish or french or gaelic, scots or georgian or german or greek or hausa or hebrew or hindi or hungarian or icelandic or indonesian or italian or japanese or kinyarwanda or kirghiz or korean or latin or latvian or lithuanian or macedonian or malay or malayalam or marathi or maori or masai or norwegian or persian or polish or portuguese or pushto or romanian or rumanian or russian or serbian or slovak or slovene or spanish or swahili or swedish or tagalog or tamil or telugu or thai or turkish or ukrainian or urdu or vietnamese or welsh) not english).lg. (4607642)

13	5 not 12 (1)

14	5 and 12 (2)

15	((afrikaans or albanian or arabic or armenian or azerbaijani or belorussian or bengali or bosnian or bulgarian or burmese or catalan or chinese or croatian or czech or danish or dutch or esperanto or estonian or finnish or flemish or french or gaelic, scots or georgian or german or greek or hausa or hebrew or hindi or hungarian or icelandic or indonesian or italian or japanese or kinyarwanda or kirghiz or korean or latin or latvian or lithuanian or macedonian or malay or malayalam or marathi or maori or masai or norwegian or persian or polish or portuguese or pushto or romanian or rumanian or russian or serbian or slovak or slovene or spanish or swahili or swedish or tagalog or tamil or telugu or thai or turkish or ukrainian or urdu or vietnamese or welsh) not (english or undetermined or multilingual)).lg. (4607600)

16	5 not 15 (1)

17	5 and 15 (2)

**Excludes: Embase** via Ovid http://ovidsp.ovid.com/

Date range searched: <1974 to 2024 March 21>

Date searched: 22/03/2024

1	"Effects of exercise on the cognition of older women treated with lovastatin".ti. (1)

2	"Estudio piloto de metilfenidato y entrenamiento a padres en el tratamiento de niños con trastorno por déficit de atención-hiperactividad".ti. (0)

3	"The effective of facial exercises on the mental health in elderly adults".ti. (1)

4	"Effect of balance exercise on functional reach test and body sway for the frail elderly".ti. (1)

5	or/1-4 (3)

6	limit 5 to english language (1)

7	5 not 6 (2)

8	limit 5 to (english language or no language specified) (1)

9	5 not 8 (2)

10	limit 5 to (english language or no language specified or polyglot) (1)

11	5 not 10 (2)

12	((afrikaans or albanian or arabic or armenian or azerbaidzhani or azerbaijani or basque or belorussian or bengali or bulgarian or burmese or bosnian or catalan or catalan; valencian or chinese or croatian or czech or danish or dutch or dutch; flemish or esperanto or estonian or finnish or french or gallegan or georgian or german or greek or "greek, modern (1453-)" or hebrew or hindi or hungarian or icelandic or indonesian or italian or japanese or korean or latvian or lithuanian or macedonian or malay or maori or mongolian or norwegian or persian or polish or portuguese or pushto or romanian or russian or serbian or sinhalese or slovak or slovene or slovenian or spanish or spanish; castilian or swedish or tagalog or thai or turkish or ukrainian or urdu or uzbek or vietnamese) not english).lg. (4698032)

13	5 not 12 (1)

14	5 and 12 (2)

15	((afrikaans or albanian or arabic or armenian or azerbaidzhani or azerbaijani or basque or belorussian or bengali or bulgarian or burmese or bosnian or catalan or catalan; valencian or chinese or croatian or czech or danish or dutch or dutch; flemish or esperanto or estonian or finnish or french or gallegan or georgian or german or greek or "greek, modern (1453-)" or hebrew or hindi or hungarian or icelandic or indonesian or italian or japanese or korean or latvian or lithuanian or macedonian or malay or maori or mongolian or norwegian or persian or polish or portuguese or pushto or romanian or russian or serbian or sinhalese or slovak or slovene or slovenian or spanish or spanish; castilian or swedish or tagalog or thai or turkish or ukrainian or urdu or uzbek or vietnamese) not (english or polyglot)).lg. (4698032)

16	5 not 15 (1)

17	5 and 15 (2)

### **Dataset 6:**

**Includes: MEDLINE® ALL** via Ovid http://ovidsp.ovid.com/

Date range searched: <1946 to February 28, 2024>

Date searched: 29th February 2024

1	"Effect of high-intensity interval training on cardiorespiratory fitness, physical activity and body composition in people with schizophrenia: A randomized controlled trial".ti. and "2020".yr. (1)

2	"A 20-week program of resistance or concurrent exercise improves symptoms of schizophrenia: Results of a blind, randomized controlled trial".ti. and "2015".yr. (1)

3	"The impact of aerobic exercise training on cardiopulmonary functioning in individuals with schizophrenia".ti. and "2016".yr. (1)

4	"Soccer practice as an add-on treatment in the management of individuals with a diagnosis of schizophrenia".ti. and "2013".yr. (1)

5	"Effects of exercise on mental and physical health parameters of persons with schizophrenia".ti. and "2005".yr. (1)

6	"A randomised controlled trial of adjunctive yoga and adjunctive physical exercise training for cognitive dysfunction in schizophrenia".ti. and "2017".yr. (1)

7	"The health benefits and challenges of exercise training in persons living with schizophrenia: A pilot study".ti. and "2013".yr. (1)

8	"Psychological wellness, yoga and quality of life in patients affected by schizophrenia spectrum disorders: A pilot study".ti. and "2019".yr. (1)

9	"Positive impact of prescribed physical activity on symptoms of schizophrenia: Randomized clinical trial".ti. and "2017".yr. (1)

10	"Therapeutic effects of maximal strength training on walking efficiency in patients with schizophrenia - a pilot study".ti. and "2012".yr. (1)

11	"Effects of high aerobic intensity training in patients with schizophrenia: A controlled trial".ti. and "2011".yr. (1)

12	"The effect of 12 weeks of aerobic, resistance or combination exercise training on cardiovascular risk factors in the overweight and obese in a randomized trial".ti. and "2012".yr. (1)

13	"Incongruent changes in heart rate variability and body weight after discontinuing aerobic exercise in patients with schizophrenia".ti. and "2016".yr. (1)

14	"Effects of yoga therapy on postural stability in patients with schizophrenia-spectrum disorders: A single-blind randomized controlled trial".ti. and "2013".yr. (1)

15	"Functional and psychosocial effects of either a traditional dancing or a formal exercising training program in patients with chronic heart failure: A comparative randomized controlled study".ti. and "2013".yr. (0)

16	"Increase of circulating bdnf levels and its relation to improvement of physical fitness following 12 weeks of combined exercise in chronic patients with schizophrenia: A pilot study".ti. and "2014".yr. (1)

17	"The impact of aerobic exercise on brain-derived neurotrophic factor and neurocognition in individuals with schizophrenia: A single-blind, randomized clinical trial".ti. and "2015".yr. (1)

18	"Structured walking and chronic institutionalized schizophrenia inmates: A pilot rct study on quality of life".ti. and "2015".yr. (1)

19	"Feasibility and effects of a group-based resistance and aerobic exercise program for individuals with severe schizophrenia: A multidisciplinary approach".ti. and "2009".yr. (0)

20	"Hippocampal plasticity in response to exercise in schizophrenia".ti. and "2010".yr. (1)

21	"Effect of body-oriented psychological therapy on negative symptoms in schizophrenia: A randomized controlled trial".ti. and "2006".yr. (1)

22	"Outdoor cycling improves clinical symptoms, cognition and objectively measured physical activity in patients with schizophrenia: A randomized controlled trial".ti. and "2020".yr. (1)

23	"Effects of exercise therapy on cardiovascular fitness and the metabolic syndrome in schizophrenia: A randomized clinical trial".ti. and "2012".yr. (0)

24	"Exercise therapy, cardiorespiratory fitness and their effect on brain volumes: A randomised controlled trial in patients with schizophrenia and healthy controls".ti. and "2013".yr. (1)

25	"Aerobic exercise and cognitive functioning in schizophrenia: A pilot randomized controlled trial".ti. and "2019".yr. (1)

26	"An interactive sports video game as an intervention for rehabilitation of community-living patients with schizophrenia: A controlled, single-blind, crossover study".ti. and "2017".yr. (1)

27	"Physical exercise keeps the brain connected: Biking increases white matter integrity in patients with schizophrenia and healthy controls".ti. and "2015".yr. (1)

28	"Therapeutic efficacy of add-on yogasana intervention in stabilized outpatient schizophrenia: Randomized controlled comparison with exercise and waitlist".ti. and "2012".yr. (1)

29	or/1-28 (25)

30	29 (25)

31	limit 30 to english language (25)

32	30 not 31 (0)

33	limit 30 to (english language or no language specified) (25)

34	30 not 33 (0)

35	limit 30 to (english language or no language specified or multilingual or undetermined) (25)

36	30 not 35 (0)

37	((afrikaans or albanian or arabic or armenian or azerbaijani or belorussian or bengali or bosnian or bulgarian or burmese or catalan or chinese or croatian or czech or danish or dutch or esperanto or estonian or finnish or flemish or french or gaelic, scots or georgian or german or greek or hausa or hebrew or hindi or hungarian or icelandic or indonesian or italian or japanese or kinyarwanda or kirghiz or korean or latin or latvian or lithuanian or macedonian or malay or malayalam or marathi or maori or masai or norwegian or persian or polish or portuguese or pushto or romanian or rumanian or russian or serbian or slovak or slovene or spanish or swahili or swedish or tagalog or tamil or telugu or thai or turkish or ukrainian or urdu or vietnamese or welsh) not english).lg. (4604123)

38	30 not 37 (25)

39	30 and 37 (0)

40	((afrikaans or albanian or arabic or armenian or azerbaijani or belorussian or bengali or bosnian or bulgarian or burmese or catalan or chinese or croatian or czech or danish or dutch or esperanto or estonian or finnish or flemish or french or gaelic, scots or georgian or german or greek or hausa or hebrew or hindi or hungarian or icelandic or indonesian or italian or japanese or kinyarwanda or kirghiz or korean or latin or latvian or lithuanian or macedonian or malay or malayalam or marathi or maori or masai or norwegian or persian or polish or portuguese or pushto or romanian or rumanian or russian or serbian or slovak or slovene or spanish or swahili or swedish or tagalog or tamil or telugu or thai or turkish or ukrainian or urdu or vietnamese or welsh) not (english or undetermined or multilingual)).lg. (4604081)

41	30 not 40 (25)

42	30 and 40 (0)

**Includes: Embase** via Ovid http://ovidsp.ovid.com/

Date range searched: <1974 to 2024 February 28>

Date searched: 29th February 2024

1	"Effect of high-intensity interval training on cardiorespiratory fitness, physical activity and body composition in people with schizophrenia: A randomized controlled trial".ti. and "2020".yr. (1)

2	"A 20-week program of resistance or concurrent exercise improves symptoms of schizophrenia: Results of a blind, randomized controlled trial".ti. and "2015".yr. (1)

3	"The impact of aerobic exercise training on cardiopulmonary functioning in individuals with schizophrenia".ti. and "2016".yr. (1)

4	"Soccer practice as an add-on treatment in the management of individuals with a diagnosis of schizophrenia".ti. and "2013".yr. (1)

5	"Effects of exercise on mental and physical health parameters of persons with schizophrenia".ti. and "2005".yr. (1)

6	"A randomised controlled trial of adjunctive yoga and adjunctive physical exercise training for cognitive dysfunction in schizophrenia".ti. and "2017".yr. (1)

7	"The health benefits and challenges of exercise training in persons living with schizophrenia: A pilot study".ti. and "2013".yr. (1)

8	"Psychological wellness, yoga and quality of life in patients affected by schizophrenia spectrum disorders: A pilot study".ti. and "2019".yr. (1)

9	"Positive impact of prescribed physical activity on symptoms of schizophrenia: Randomized clinical trial".ti. and "2017".yr. (1)

10	"Therapeutic effects of maximal strength training on walking efficiency in patients with schizophrenia - a pilot study".ti. and "2012".yr. (1)

11	"Effects of high aerobic intensity training in patients with schizophrenia: A controlled trial".ti. and "2011".yr. (1)

12	"The effect of 12 weeks of aerobic, resistance or combination exercise training on cardiovascular risk factors in the overweight and obese in a randomized trial".ti. and "2012".yr. (1)

13	"Incongruent changes in heart rate variability and body weight after discontinuing aerobic exercise in patients with schizophrenia".ti. and "2016".yr. (1)

14	"Effects of yoga therapy on postural stability in patients with schizophrenia-spectrum disorders: A single-blind randomized controlled trial".ti. and "2013".yr. (1)

15	"Functional and psychosocial effects of either a traditional dancing or a formal exercising training program in patients with chronic heart failure: A comparative randomized controlled study".ti. and "2013".yr. (0)

16	"Increase of circulating bdnf levels and its relation to improvement of physical fitness following 12 weeks of combined exercise in chronic patients with schizophrenia: A pilot study".ti. and "2014".yr. (1)

17	"The impact of aerobic exercise on brain-derived neurotrophic factor and neurocognition in individuals with schizophrenia: A single-blind, randomized clinical trial".ti. and "2015".yr. and Schizophrenia Bulletin.jn. (1)

18	"Structured walking and chronic institutionalized schizophrenia inmates: A pilot rct study on quality of life".ti. and "2015".yr. (0)

19	"Feasibility and effects of a group-based resistance and aerobic exercise program for individuals with severe schizophrenia: A multidisciplinary approach".ti. and "2009".yr. (1)

20	"Hippocampal plasticity in response to exercise in schizophrenia".ti. and "2010".yr. (1)

21	"Effect of body-oriented psychological therapy on negative symptoms in schizophrenia: A randomized controlled trial".ti. and "2006".yr. (1)

22	"Outdoor cycling improves clinical symptoms, cognition and objectively measured physical activity in patients with schizophrenia: A randomized controlled trial".ti. and "2020".yr. (1)

23	"Effects of exercise therapy on cardiovascular fitness and the metabolic syndrome in schizophrenia: A randomized clinical trial".ti. and "2012".yr. (1)

24	"Exercise therapy, cardiorespiratory fitness and their effect on brain volumes: A randomised controlled trial in patients with schizophrenia and healthy controls".ti. and "2013".yr. and Scheewe.au. (0)

25	"Aerobic exercise and cognitive functioning in schizophrenia: A pilot randomized controlled trial".ti. and "2019".yr. (1)

26	"An interactive sports video game as an intervention for rehabilitation of community-living patients with schizophrenia: A controlled, single-blind, crossover study".ti. and "2017".yr. (1)

27	"Physical exercise keeps the brain connected: Biking increases white matter integrity in patients with schizophrenia and healthy controls".ti. and "2015".yr. (1)

28	"Therapeutic efficacy of add-on yogasana intervention in stabilized outpatient schizophrenia: Randomized controlled comparison with exercise and waitlist".ti. and "2012".yr. (1)

29	or/1-28 (25)

30	29 (25)

31	limit 30 to english language (25)

32	30 not 31 (0)

33	limit 30 to (english language or no language specified) (25)

34	30 not 33 (0)

35	limit 30 to (english language or no language specified or polyglot) (25)

36	30 not 35 (0)

37	((afrikaans or albanian or arabic or armenian or azerbaidzhani or azerbaijani or basque or belorussian or bengali or bulgarian or burmese or bosnian or catalan or catalan; valencian or chinese or croatian or czech or danish or dutch or dutch; flemish or esperanto or estonian or finnish or french or gallegan or georgian or german or greek or "greek, modern (1453-)" or hebrew or hindi or hungarian or icelandic or indonesian or italian or japanese or korean or latvian or lithuanian or macedonian or malay or maori or mongolian or norwegian or persian or polish or portuguese or pushto or romanian or russian or serbian or sinhalese or slovak or slovene or slovenian or spanish or spanish; castilian or swedish or tagalog or thai or turkish or ukrainian or urdu or uzbek or vietnamese) not english).lg. (4689552)

38	30 not 37 (25)

39	30 and 37 (0)

40	((afrikaans or albanian or arabic or armenian or azerbaidzhani or azerbaijani or basque or belorussian or bengali or bulgarian or burmese or bosnian or catalan or catalan; valencian or chinese or croatian or czech or danish or dutch or dutch; flemish or esperanto or estonian or finnish or french or gallegan or georgian or german or greek or "greek, modern (1453-)" or hebrew or hindi or hungarian or icelandic or indonesian or italian or japanese or korean or latvian or lithuanian or macedonian or malay or maori or mongolian or norwegian or persian or polish or portuguese or pushto or romanian or russian or serbian or sinhalese or slovak or slovene or slovenian or spanish or spanish; castilian or swedish or tagalog or thai or turkish or ukrainian or urdu or uzbek or vietnamese) not (english or polyglot)).lg. (4689552)

41	30 not 40 (25)

42	30 and 40 (0)

**Excludes: MEDLINE® ALL** via Ovid http://ovidsp.ovid.com/

Date range searched: <1946 to February 28, 2024>

Date searched: 29th February 2024

1	"The effect of life skills training on functioning in schizophrenia: A randomized controlled trial".ti. and "2020".yr. (1)

2	"[erythrocyte ketone reductase activity, total plasma haloperidol and acute psychoses]".ti. and "1995".yr. (1)

3	"Psychoedukativ-psycho- therapeutische behandlung von schizophrenen patienten und ihren bezugspersonen".ti. and "1999".yr. (0)

4	"[randomized controlled trial on effectiveness of the community re-entry program to inpatients with schizophrenia spectrum disorder, centering around acquisition of illness self-management knowledge]".ti. and "2003".yr. (1)

5	"Neuropsychologie der psychoedukation bei schizophrenie".ti. and "2011".yr. (0)

6	"Effekte neuer kognitiv-behavioraler therapieprogramme zur verbesserung spezifischer sozialer fertigkeiten bei schizophren erkrankten eine kontrollierte studie".ti. and "2001".yr. (0)

7	"Vado approach efficacy in psychiatric rehabilitation: A controlled study".ti. and "2003".yr. (0)

8	or/1-7 (3)

9	8 (3)

10	limit 9 to english language (1)

11	9 not 10 (2)

12	limit 9 to (english language or no language specified) (1)

13	9 not 12 (2)

14	limit 9 to (english language or no language specified or multilingual or undetermined) (1)

15	9 not 14 (2)

16	((afrikaans or albanian or arabic or armenian or azerbaijani or belorussian or bengali or bosnian or bulgarian or burmese or catalan or chinese or croatian or czech or danish or dutch or esperanto or estonian or finnish or flemish or french or gaelic, scots or georgian or german or greek or hausa or hebrew or hindi or hungarian or icelandic or indonesian or italian or japanese or kinyarwanda or kirghiz or korean or latin or latvian or lithuanian or macedonian or malay or malayalam or marathi or maori or masai or norwegian or persian or polish or portuguese or pushto or romanian or rumanian or russian or serbian or slovak or slovene or spanish or swahili or swedish or tagalog or tamil or telugu or thai or turkish or ukrainian or urdu or vietnamese or welsh) not english).lg. (4604123)

17	9 not 16 (1)

18	9 and 16 (2)

19	((afrikaans or albanian or arabic or armenian or azerbaijani or belorussian or bengali or bosnian or bulgarian or burmese or catalan or chinese or croatian or czech or danish or dutch or esperanto or estonian or finnish or flemish or french or gaelic, scots or georgian or german or greek or hausa or hebrew or hindi or hungarian or icelandic or indonesian or italian or japanese or kinyarwanda or kirghiz or korean or latin or latvian or lithuanian or macedonian or malay or malayalam or marathi or maori or masai or norwegian or persian or polish or portuguese or pushto or romanian or rumanian or russian or serbian or slovak or slovene or spanish or swahili or swedish or tagalog or tamil or telugu or thai or turkish or ukrainian or urdu or vietnamese or welsh) not (english or undetermined or multilingual)).lg. (4604081)

20	9 not 19 (1)

21	9 and 19 (2)

22	10 or 12 or 14 or 17 or 20 (1)

**Excludes: Embase** via Ovid http://ovidsp.ovid.com/

Date range searched: <1974 to 2024 February 29>

Date searched: 1st March 2024

1	"The effect of life skills training on functioning in schizophrenia: A randomized controlled trial".ti. and "2020".yr. and Article.pt. (1)

2	"[erythrocyte ketone reductase activity, total plasma haloperidol and acute psychoses]".ti. and "1995".yr. (0)

3	"Psychoedukativ-psycho- therapeutische behandlung von schizophrenen patienten und ihren bezugspersonen".ti. and "1999".yr. (0)

4	"[randomized controlled trial on effectiveness of the community re-entry program to inpatients with schizophrenia spectrum disorder, centering around acquisition of illness self-management knowledge]".ti. and "2003".yr. (1)

5	"Neuropsychologie der psychoedukation bei schizophrenie".ti. and "2011".yr. (0)

6	"Effekte neuer kognitiv-behavioraler therapieprogramme zur verbesserung spezifischer sozialer fertigkeiten bei schizophren erkrankten eine kontrollierte studie".ti. and "2001".yr. (0)

7	"Vado approach efficacy in psychiatric rehabilitation: A controlled study".ti. and "2003".yr. (1)

8	or/1-7 (3)

9	8 (3)

10	limit 9 to english language (1)

11	9 not 10 (2)

12	limit 9 to (english language or no language specified) (1)

13	9 not 12 (2)

14	limit 9 to (english language or no language specified or polyglot) (1)

15	9 not 14 (2)

16	((afrikaans or albanian or arabic or armenian or azerbaidzhani or azerbaijani or basque or belorussian or bengali or bulgarian or burmese or bosnian or catalan or catalan; valencian or chinese or croatian or czech or danish or dutch or dutch; flemish or esperanto or estonian or finnish or french or gallegan or georgian or german or greek or "greek, modern (1453-)" or hebrew or hindi or hungarian or icelandic or indonesian or italian or japanese or korean or latvian or lithuanian or macedonian or malay or maori or mongolian or norwegian or persian or polish or portuguese or pushto or romanian or russian or serbian or sinhalese or slovak or slovene or slovenian or spanish or spanish; castilian or swedish or tagalog or thai or turkish or ukrainian or urdu or uzbek or vietnamese) not english).lg. (4690169)

17	9 not 16 (1)

18	9 and 16 (2)

19	((afrikaans or albanian or arabic or armenian or azerbaidzhani or azerbaijani or basque or belorussian or bengali or bulgarian or burmese or bosnian or catalan or catalan; valencian or chinese or croatian or czech or danish or dutch or dutch; flemish or esperanto or estonian or finnish or french or gallegan or georgian or german or greek or "greek, modern (1453-)" or hebrew or hindi or hungarian or icelandic or indonesian or italian or japanese or korean or latvian or lithuanian or macedonian or malay or maori or mongolian or norwegian or persian or polish or portuguese or pushto or romanian or russian or serbian or sinhalese or slovak or slovene or slovenian or spanish or spanish; castilian or swedish or tagalog or thai or turkish or ukrainian or urdu or uzbek or vietnamese) not (english or polyglot)).lg. (4690169)

20	9 not 19 (1)

21	9 and 19 (2)

### **Dataset 7:**

**Includes: MEDLINE® ALL** via Ovid http://ovidsp.ovid.com/

Date range searched: <1946 to March 21, 2024>

Date searched: 22/03/2024

1	"Nurses' perceptions of futile care: A qualitative study".ti. and "2016".yr. (1)

2	"Reasons behind providing futile medical treatments in iran".ti. and "2017".yr. (1)

3	"Perceptions and lived experiences of nurses who provide care to extremely immature newborns".ti. and "2013".yr. (0)

4	"The experience of the nurse in providing futile care".ti. and "1995".yr. (0)

5	"Paradox of prescribing late chemotherapy: Oncologists explain".ti. and "2016".yr. (1)

6	"Exploring nurses' attitudes about the value of a hospital based medical futility policy".ti. and "2016".yr. (0)

7	"Surgeons' perspectives on avoiding nonbeneficial treatments in seriously ill older patients with surgical emergencies: A qualitative study".ti. and "2016".yr. (1)

8	"Understanding early decisions to withdraw life-sustaining therapy in cardiac arrest survivors. A qualitative investigation".ti. and "2016".yr. (1)

9	"Therapeutic futility as an ethical issue: Intensive care unit nurses".ti. and "2009".yr. (1)

10	"Dysthanasia: Nursing professionals' perception".ti. and "2009".yr. (1)

11	"Moral distress amongst american physician trainees regarding futile treatments at the end of life: A qualitative study".ti. and "2016".yr. (1)

12	"Homing in on the social: System-level influences on overly aggressive treatments at the end of life".ti. and "2018".yr. (1)

13	"Understanding the moral distress of nurses witnessing medically futile care".ti. and "2006".yr. (1)

14	"Understanding psycho-social processes underpinning engagement with services in motor neurone disease: A qualitative study".ti. and "2014".yr. (1)

15	"Futile treatment in hospital: Doctors' intergroup language".ti. and "2015".yr. (0)

16	"Being on the frontline: Nurses' experiences providing end-of-life care to adults with hematologic malignancies".ti. and "2018".yr. (1)

17	"System-level barriers and facilitators for foregoing or withdrawing dialysis: A qualitative study of nephrologists in the united states and england".ti. and "2017".yr. (1)

18	"Giving moral distress a voice: Ethical concerns among neonatal intensive care unit personnel".ti. and "1999".yr. (1)

19	"Fruitful or futile: Intensive care nurses' experiences and perceptions of medical futility".ti. and "2006".yr. (1)

20	"Pain management in persons who are terminally ill in rural acute care: Barriers and facilitators".ti. and "2012".yr. (0)

21	"Medical futility at the end of life: The perspectives of intensive care and palliative care clinicians".ti. and "2012".yr. (1)

22	"The window of opportunity for death after severe brain injury: Family experiences".ti. and "2013".yr. (1)

23	"Discussing conservative management with older patients with ckd: An interview study of nephrologists".ti. and "2018".yr. (1)

24	"Rather one more chemo than one less...: Oncologists and oncology nurses' reasons for aggressive treatment of young adults with advanced cancer".ti. and "2018".yr. (1)

25	"Working in the grey zone: The moral suffering of critical care nurses".ti. and "1994".yr. (0)

26	"U.S. Physicians' perceptions of malpractice liability factors in aggressive treatment of dying patients".ti. and "1987".yr. (1)

27	"Non-treatment decisions on grounds of medical futility and quality of life: Interviews with fourteen dutch neonatologists".ti. and "2010".yr. (1)

28	"Shared decision-making in acute surgical illness: The surgeon's perspective".ti. and "2018".yr. (1)

29	"Ethical problems in intensive care unit admission and discharge decisions: A qualitative study among physicians and nurses in the netherlands".ti. and "2015".yr. (1)

30	"Narratives of aggressive care: Knowledge, time, and responsibility".ti. and "2014".yr. (1)

31	"Orthothanasia and dysthanasia: Perception of health professionals of an intensive care".ti. and "2016".yr. (0)

32	"Perceptions of patients on the utility or futility of end-of-life treatment".ti. and "2006".yr. (1)

33	"Current attitudes of anesthesiologists towards medically futile care".ti. and "2013".yr. (0)

34	"The ethical decisions uk doctors make regarding advanced cancer patients at the end of life--the perceived (in) appropriateness of anticoagulation for venous thromboembolism: A qualitative study".ti. and "2012".yr. (1)

35	"Perceptions of futile care among caregivers in intensive care units".ti. and "2007".yr. (1)

36	"Reflections on life and death in a technological society: Experiences of doctors and nurses with dying patients in intensive care".ti. and "1996".yr. (0)

37	"Women physicians' narratives about being in ethically difficult care situations in paediatrics".ti. and "2000".yr. (1)

38	"Conflict over treatment withdrawal in the intensive care unit: A description of the health care providers' experience".ti. and "1998".yr. (0)

39	"Nurses' experiences of futile care at intensive care units: A phenomenological study".ti. and "2015".yr. (1)

40	"Surrogate decision makers' responses to physicians' predictions of medical futility".ti. and "2009".yr. (1)

41	"Exploring the moral distress of registered nurses".ti. and "2007".yr. (1)

42	or/1-41 (31)

43	limit 42 to english language (31)

44	42 not 43 (0)

45	limit 42 to (english language or no language specified) (31)

46	42 not 45 (0)

47	limit 42 to (english language or no language specified or multilingual or undetermined) (31)

48	42 not 47 (0)

49	((afrikaans or albanian or arabic or armenian or azerbaijani or belorussian or bengali or bosnian or bulgarian or burmese or catalan or chinese or croatian or czech or danish or dutch or esperanto or estonian or finnish or flemish or french or gaelic, scots or georgian or german or greek or hausa or hebrew or hindi or hungarian or icelandic or indonesian or italian or japanese or kinyarwanda or kirghiz or korean or latin or latvian or lithuanian or macedonian or malay or malayalam or marathi or maori or masai or norwegian or persian or polish or portuguese or pushto or romanian or rumanian or russian or serbian or slovak or slovene or spanish or swahili or swedish or tagalog or tamil or telugu or thai or turkish or ukrainian or urdu or vietnamese or welsh) not english).lg. (4607642)

50	42 not 49 (31)

51	42 and 49 (0)

52	((afrikaans or albanian or arabic or armenian or azerbaijani or belorussian or bengali or bosnian or bulgarian or burmese or catalan or chinese or croatian or czech or danish or dutch or esperanto or estonian or finnish or flemish or french or gaelic, scots or georgian or german or greek or hausa or hebrew or hindi or hungarian or icelandic or indonesian or italian or japanese or kinyarwanda or kirghiz or korean or latin or latvian or lithuanian or macedonian or malay or malayalam or marathi or maori or masai or norwegian or persian or polish or portuguese or pushto or romanian or rumanian or russian or serbian or slovak or slovene or spanish or swahili or swedish or tagalog or tamil or telugu or thai or turkish or ukrainian or urdu or vietnamese or welsh) not (english or undetermined or multilingual)).lg. (4607600)

53	42 not 52 (31)

54	42 and 52 (0)

**Includes: Embase** via Ovid http://ovidsp.ovid.com/

Date range searched: <1974 to 2024 March 21>

Date searched: 22/03/2024

1	"Nurses' perceptions of futile care: A qualitative study".ti. and "2016".yr. (1)

2	"Reasons behind providing futile medical treatments in iran".ti. and "2017".yr. (1)

3	"Perceptions and lived experiences of nurses who provide care to extremely immature newborns".ti. and "2013".yr. (0)

4	"The experience of the nurse in providing futile care".ti. and "1995".yr. (0)

5	"Paradox of prescribing late chemotherapy: Oncologists explain".ti. and "2016".yr. (1)

6	"Exploring nurses' attitudes about the value of a hospital based medical futility policy".ti. and "2016".yr. (0)

7	"Surgeons' perspectives on avoiding nonbeneficial treatments in seriously ill older patients with surgical emergencies: A qualitative study".ti. and "2016".yr. (1)

8	"Understanding early decisions to withdraw life-sustaining therapy in cardiac arrest survivors. A qualitative investigation".ti. and "2016".yr. (1)

9	"Therapeutic futility as an ethical issue: Intensive care unit nurses".ti. and "2009".yr. (1)

10	"Dysthanasia: Nursing professionals' perception".ti. and "2009".yr. (1)

11	"Moral distress amongst american physician trainees regarding futile treatments at the end of life: A qualitative study".ti. and "2016".yr. (1)

12	"Homing in on the social: System-level influences on overly aggressive treatments at the end of life".ti. and "2018".yr. (1)

13	"Understanding the moral distress of nurses witnessing medically futile care".ti. and "2006".yr. (1)

14	"Understanding psycho-social processes underpinning engagement with services in motor neurone disease: A qualitative study".ti. and "2014".yr. (1)

15	"Futile treatment in hospital: Doctors' intergroup language".ti. and "2015".yr. (0)

16	"Being on the frontline: Nurses' experiences providing end-of-life care to adults with hematologic malignancies".ti. and "2018".yr. (1)

17	"System-level barriers and facilitators for foregoing or withdrawing dialysis: A qualitative study of nephrologists in the united states and england".ti. and "2017".yr. (1)

18	"Giving moral distress a voice: Ethical concerns among neonatal intensive care unit personnel".ti. and "1999".yr. (1)

19	"Fruitful or futile: Intensive care nurses' experiences and perceptions of medical futility".ti. and "2006".yr. (1)

20	"Pain management in persons who are terminally ill in rural acute care: Barriers and facilitators".ti. and "2012".yr. (0)

21	"Medical futility at the end of life: The perspectives of intensive care and palliative care clinicians".ti. and "2012".yr. (1)

22	"The window of opportunity for death after severe brain injury: Family experiences".ti. and "2013".yr. (1)

23	"Discussing conservative management with older patients with ckd: An interview study of nephrologists".ti. and "2018".yr. (1)

24	"Rather one more chemo than one less...: Oncologists and oncology nurses' reasons for aggressive treatment of young adults with advanced cancer".ti. and "2018".yr. (1)

25	"Working in the grey zone: The moral suffering of critical care nurses".ti. and "1994".yr. (0)

26	"U.S. Physicians' perceptions of malpractice liability factors in aggressive treatment of dying patients".ti. and "1987".yr. (1)

27	"Non-treatment decisions on grounds of medical futility and quality of life: Interviews with fourteen dutch neonatologists".ti. and "2010".yr. and "01 Jun 2010".dp. (1)

28	"Shared decision-making in acute surgical illness: The surgeon's perspective".ti. and "2018".yr. (1)

29	"Ethical problems in intensive care unit admission and discharge decisions: A qualitative study among physicians and nurses in the netherlands".ti. and "2015".yr. (1)

30	"Narratives of aggressive care: Knowledge, time, and responsibility".ti. and "2014".yr. (1)

31	"Orthothanasia and dysthanasia: Perception of health professionals of an intensive care".ti. and "2016".yr. (0)

32	"Perceptions of patients on the utility or futility of end-of-life treatment".ti. and "2006".yr. (0)

33	"Current attitudes of anesthesiologists towards medically futile care".ti. and "2013".yr. (0)

34	"The ethical decisions uk doctors make regarding advanced cancer patients at the end of life--the perceived (in) appropriateness of anticoagulation for venous thromboembolism: A qualitative study".ti. and "2012".yr. (1)

35	"Perceptions of futile care among caregivers in intensive care units".ti. and "2007".yr. (1)

36	"Reflections on life and death in a technological society: Experiences of doctors and nurses with dying patients in intensive care".ti. and "1996".yr. (0)

37	"Women physicians' narratives about being in ethically difficult care situations in paediatrics".ti. and "2000".yr. (1)

38	"Conflict over treatment withdrawal in the intensive care unit: A description of the health care providers' experience".ti. and "1998".yr. (0)

39	"Nurses' experiences of futile care at intensive care units: A phenomenological study".ti. and "2015".yr. (1)

40	"Surrogate decision makers' responses to physicians' predictions of medical futility".ti. and "2009".yr. (1)

41	"Exploring the moral distress of registered nurses".ti. and "2007".yr. (1)

42	or/1-41 (30)

43	limit 42 to english language (28)

44	42 not 43 (2)

45	limit 42 to (english language or no language specified) (28)

46	42 not 45 (2)

47	((afrikaans or albanian or arabic or armenian or azerbaidzhani or azerbaijani or basque or belorussian or bengali or bulgarian or burmese or bosnian or catalan or catalan; valencian or chinese or croatian or czech or danish or dutch or dutch; flemish or esperanto or estonian or finnish or french or gallegan or georgian or german or greek or "greek, modern (1453-)" or hebrew or hindi or hungarian or icelandic or indonesian or italian or japanese or korean or latvian or lithuanian or macedonian or malay or maori or mongolian or norwegian or persian or polish or portuguese or pushto or romanian or russian or serbian or sinhalese or slovak or slovene or slovenian or spanish or spanish; castilian or swedish or tagalog or thai or turkish or ukrainian or urdu or uzbek or vietnamese) not english).lg. (4698032)

48	42 not 47 (28)

49	42 and 47 (2)

50	((afrikaans or albanian or arabic or armenian or azerbaidzhani or azerbaijani or basque or belorussian or bengali or bulgarian or burmese or bosnian or catalan or catalan; valencian or chinese or croatian or czech or danish or dutch or dutch; flemish or esperanto or estonian or finnish or french or gallegan or georgian or german or greek or "greek, modern (1453-)" or hebrew or hindi or hungarian or icelandic or indonesian or italian or japanese or korean or latvian or lithuanian or macedonian or malay or maori or mongolian or norwegian or persian or polish or portuguese or pushto or romanian or russian or serbian or sinhalese or slovak or slovene or slovenian or spanish or spanish; castilian or swedish or tagalog or thai or turkish or ukrainian or urdu or uzbek or vietnamese) not (english or polyglot)).lg. (4698032)

51	42 not 50 (28)

52	42 and 50 (2)

**Excludes: MEDLINE® ALL** via Ovid http://ovidsp.ovid.com/

Date range searched: <1946 to March 21, 2024>

Date searched: 22/03/2024

1	"Definition of (in)appropriate care seen from the patient's perspective".ti. and "2018".yr. (0)

2	"Concepções de enfermeiras sobre o prolongamento artificial da vida".ti. and "2014".yr. (0)

3	"Age and costs - factors in treatment decisions at the end-of-life? An analysis of informal knowledge structures of doctors and nurses".ti. and "2007".yr. (0)

4	"Therapeutic obstinacy at an intensive care unit: Perspective from doctors and nurses".ti. and "2012".yr. (0)

5	or/1-4 (0)

6	limit 5 to english language (0)

7	5 not 6 (0)

8	limit 5 to (english language or no language specified) (0)

9	5 not 8 (0)

10	limit 5 to (english language or no language specified or multilingual or undetermined) (0)

11	5 not 10 (0)

12	((afrikaans or albanian or arabic or armenian or azerbaijani or belorussian or bengali or bosnian or bulgarian or burmese or catalan or chinese or croatian or czech or danish or dutch or esperanto or estonian or finnish or flemish or french or gaelic, scots or georgian or german or greek or hausa or hebrew or hindi or hungarian or icelandic or indonesian or italian or japanese or kinyarwanda or kirghiz or korean or latin or latvian or lithuanian or macedonian or malay or malayalam or marathi or maori or masai or norwegian or persian or polish or portuguese or pushto or romanian or rumanian or russian or serbian or slovak or slovene or spanish or swahili or swedish or tagalog or tamil or telugu or thai or turkish or ukrainian or urdu or vietnamese or welsh) not english).lg. (4607642)

13	5 not 12 (0)

14	5 and 12 (0)

15	((afrikaans or albanian or arabic or armenian or azerbaijani or belorussian or bengali or bosnian or bulgarian or burmese or catalan or chinese or croatian or czech or danish or dutch or esperanto or estonian or finnish or flemish or french or gaelic, scots or georgian or german or greek or hausa or hebrew or hindi or hungarian or icelandic or indonesian or italian or japanese or kinyarwanda or kirghiz or korean or latin or latvian or lithuanian or macedonian or malay or malayalam or marathi or maori or masai or norwegian or persian or polish or portuguese or pushto or romanian or rumanian or russian or serbian or slovak or slovene or spanish or swahili or swedish or tagalog or tamil or telugu or thai or turkish or ukrainian or urdu or vietnamese or welsh) not (english or undetermined or multilingual)).lg. (4607600)

16	5 not 15 (0)

17	5 and 15 (0)

**Excludes: Embase** via Ovid http://ovidsp.ovid.com/

Date range searched: <1974 to 2024 March 21>

Date searched: 22/03/2024

1	"Definition of (in)appropriate care seen from the patient's perspective".ti. and "2018".yr. (1)

2	"Concepções de enfermeiras sobre o prolongamento artificial da vida".ti. and "2014".yr. (0)

3	"Age and costs - factors in treatment decisions at the end-of-life? An analysis of informal knowledge structures of doctors and nurses".ti. and "2007".yr. (0)

4	"Therapeutic obstinacy at an intensive care unit: Perspective from doctors and nurses".ti. and "2012".yr. (0)

5	or/1-4 (1)

6	limit 5 to english language (1)

7	5 not 6 (0)

8	limit 5 to (english language or no language specified) (1)

9	5 not 8 (0)

10	limit 5 to (english language or no language specified or polyglot) (1)

11	5 not 10 (0)

12	((afrikaans or albanian or arabic or armenian or azerbaidzhani or azerbaijani or basque or belorussian or bengali or bulgarian or burmese or bosnian or catalan or catalan; valencian or chinese or croatian or czech or danish or dutch or dutch; flemish or esperanto or estonian or finnish or french or gallegan or georgian or german or greek or "greek, modern (1453-)" or hebrew or hindi or hungarian or icelandic or indonesian or italian or japanese or korean or latvian or lithuanian or macedonian or malay or maori or mongolian or norwegian or persian or polish or portuguese or pushto or romanian or russian or serbian or sinhalese or slovak or slovene or slovenian or spanish or spanish; castilian or swedish or tagalog or thai or turkish or ukrainian or urdu or uzbek or vietnamese) not english).lg. (4698032)

13	5 not 12 (1)

14	5 and 12 (0)

15	((afrikaans or albanian or arabic or armenian or azerbaidzhani or azerbaijani or basque or belorussian or bengali or bulgarian or burmese or bosnian or catalan or catalan; valencian or chinese or croatian or czech or danish or dutch or dutch; flemish or esperanto or estonian or finnish or french or gallegan or georgian or german or greek or "greek, modern (1453-)" or hebrew or hindi or hungarian or icelandic or indonesian or italian or japanese or korean or latvian or lithuanian or macedonian or malay or maori or mongolian or norwegian or persian or polish or portuguese or pushto or romanian or russian or serbian or sinhalese or slovak or slovene or slovenian or spanish or spanish; castilian or swedish or tagalog or thai or turkish or ukrainian or urdu or uzbek or vietnamese) not (english or polyglot)).lg. (4698032)

16	5 not 15 (1)

17	5 and 15 (0)
